# Cancer incidence and competing mortality risk following 15 presenting symptoms in primary care: a population-based cohort study using electronic healthcare records

**DOI:** 10.1101/2024.05.21.24307662

**Authors:** Matthew Barclay, Cristina Renzi, Hannah Harrison, Ana Torralbo, Becky White, Samantha Ip, Juliet Usher-Smith, Jane Lange, Nora Pashayan, Spiros Denaxas, Angela Wood, Antonis C Antoniou, Georgios Lyratzopoulos

**Affiliations:** Department of Behavioural Science and Health, Institute of Epidemiology and Healthcare, University College London, London, United Kingdom; Faculty of Medicine, University Vita-Salute San Raffaele, Milan, Italy; Department of Public Health and Primary Care, School of Clinical Medicine, University of Cambridge, Cambridge, United Kingdom; Institute of Health Informatics, University College London, London, United Kingdom; Victor Phillip Dahdaleh Heart and Lung Research Institute, University of Cambridge, Cambridge, United Kingdom; Cancer Early Detection Advanced Research Center, Oregon Health & Science University, Portland, Oregon, United States of America; Department of Applied Health Research, Institute of Epidemiology and Healthcare, University College London, London, United Kingdom; British Heart Foundation Centre of Research Excellence, University of Cambridge, Cambridge, United Kingdom; National Institute for Health and Care Research Blood and Transplant Research Unit in Donor Health and Behaviour, University of Cambridge, Cambridge, United Kingdom; Health Data Research UK Cambridge, Wellcome Genome Campus and University of Cambridge, Cambridge, United Kingdom; Cambridge Centre for Artificial Intelligence in Medicine, University of Cambridge, Cambridge, United Kingdom

**Author notes:** The Corresponding Author has the right to grant on behalf of all authors and does grant on behalf of all authors, an exclusive licence (or non exclusive for government employees) on a worldwide basis to the BMJ Publishing Group Ltd to permit this article (if accepted) to be published in BMJ editions and any other BMJPGL products and sublicences such use and exploit all subsidiary rights, as set out in our licence.

## Abstract

**Objectives:** Assessment of age, sex and smoking-specific risk of cancer diagnosis and non-cancer mortality following primary care consultation for 15 new-onset symptoms.

**Methods and analysis:** Data on patients aged 18-99 in 2007 – 2017 were extracted from a UK primary care database (CPRD Gold), comprising a randomly-selected reference group and a symptomatic cohort of patients presenting with one of 15 new onset symptoms (abdominal pain, abdominal bloating, rectal bleed, change in bowel habit, dyspepsia, dysphagia, dyspnoea, haemoptysis, haematuria, fatigue, night sweats, weight loss, jaundice, breast lump, post-menopausal bleed).

Time-to-event models were used to estimate outcome-specific hazards for site-specific cancer diagnosis and non-cancer mortality, and used to estimate cumulative incidence up to 12 months following index consultation.

**Results:** Data included 1,622,419 patients, of whom 36,802 had a cancer diagnosis and 28,857 died without a cancer diagnosis within 12 months of index.

Risk of specific cancers exceeded the UK urgent referral risk threshold of 3% from a relatively young age for patients with red flag symptoms. For non-organ-specific symptoms, the risk of individual cancer sites either did not reach the threshold at any age, or reached it only in older patients.

**Conclusion:** Patients with new-onset symptoms in primary care often have comparable risk of cancer diagnosis and of non-cancer mortality. A holistic approach to risk assessment that includes the risk of different cancer types alongside mortality risk, especially among older patients, is needed to inform management of symptomatic patients in primary care, particularly for patients with non-organ-specific symptoms.

**Summary box:** *What is already known on this topic:*

- Evidence describing the diagnostic value of symptoms for cancer can help to assess which patients who present to primary care need urgent specialist assessment
- Current evidence is limited as age is often handled categorically, smoking status is not taken into account and study periods are historical.
- Further, evidence is concentrated on assessing the risk of specific cancer sites, although the same symptom can be related to cancer of different organs.

*What this study adds:*

- We present evidence on age-, sex-, and smoking status-specific estimates of risk of cancer of different organs and overall, alongside estimates of non-cancer death.
- Estimates relate to patients who present with one of 15 possible cancer symptoms, from a relatively recent time period.
- Certain symptoms such as jaundice and dysphagia are associated with high risk of non-cancer death in older patients.
- Other symptoms, such as unintended weight loss, fatigue and abdominal pain, are associated with excess risk of a range of different cancers, and such evidence can guide the choice of diagnostic strategies and the design of multi-cancer diagnostic services.

## Introduction

Most patients with cancer are diagnosed after symptomatic presentation [1], and, given the paucity of effective tests to enable population-based cancer screening, this is likely to be the case for the coming decade. Appropriately suspecting the diagnosis of cancer in symptomatic patients is difficult, as symptoms may be caused by many other diseases. Even so-termed ‘alarm’ or ‘red-flag’ symptoms typically have positive predictive values for cancer that do not exceed 5% in women of any age or in men younger than 70 [2]. In the UK, many patients with cancer experience diagnostic delays in the form of multiple pre-referral consultations and prolonged intervals to diagnosis, despite practice guidelines issued by the National Institute for Health and Social Care Excellence (NICE) that aimed to enable prompt diagnosis of cancer in primary care [7,8]. Such delays are associated with adverse patient experience and worse clinical outcomes [3–6],

Currently, most evidence supporting practice guidelines comes from case-control studies, examining symptom-related risk of specific cancer sites. This study design ignores that presenting symptoms are often shared between different cancers and diseases other than cancer; there has been no comprehensive examination of the risk of the full spectrum of possible cancer types for most relevant presenting symptoms. Further, guideline recommendations handle major cancer risk factors sub-optimally, as smoking status is typically ignored as a risk stratifier, and age typically not considered as a continuous variable, leading to information loss. Competing risk of death is also ignored, meaning that management decisions centred on cancer risk ignore risks related to other diseases.

This study is motivated by the need for evidence to support the updating of clinical practice guidelines for the primary care management of patients who present with symptoms of possible underlying cancer. Such evidence is needed both in terms of quantifying the absolute risk of different cancer types and also the probability of patients dying without a cancer diagnosis. We also aim to aid the development of and complement the use of risk prediction tools by describing in detail the associations between symptoms and cancer risk [9,10]. We therefore provide a comprehensive assessment of risk of cancer diagnosis and non-cancer mortality following consultation for 15 new-onset symptoms.

## Methods

### Study population

We used medical records from English National Health Service general practices that contributed anonymized primary-care electronic health records to the Clinical Practice Research Datalink Gold (CPRD), covering approximately 6.9% of the UK population [11]. Patients in CPRD are broadly representative of the UK general population with respect to age, sex, and ethnicity [11]. CPRD was linked to cancer diagnosis information from the English national cancer registry [12].

We first extracted a random sample of patients from CPRD for use as a reference group, choosing index dates randomly from ‘valid’ follow-up during 2007-01-01 to 2017-12-31. We then created a symptomatic cohort of all patients in CPRD Gold who had consulted for any of 15 presenting symptoms and who were not in the reference group, choosing the index date as the date of their first ‘valid’ consultation for a symptom during 2007-01-01 to 2017-12-31.

For an individual patient, follow-up was judged to be ‘valid’ if: they had been registered at their practice for at least one year; their practice was judged by CPRD to be providing data of a suitable standard for use in research (i.e., after the practice’s “up-to-standard” date); it was before the last data transfer to CPRD (i.e., the “last collection” date); the patient was registered at a CPRD practice (i.e., before the patient’s “transfer out” date, and before their death); the patient was aged 30-99; and the patient had not yet had a recorded cancer diagnosis in the cancer registry (excluding non-melanoma skin cancer).

A study flowchart is given in Appendix 1 Table 1.

### Outcomes

Both mortality and cancer diagnoses were considered. Mortality was identified from the primary care record; such information is highly concordant with the ‘gold standard’ official death registration records and is correct within one month 98% of the time [13]. Cancers were split into seven groups for men and eight groups for women, summarised below and with a full ICD10 codelist in Appendix 1 Table 2, guided by underlying body systems and corresponding major clinical specialities receiving urgent referrals for suspected cancer in England [14]. Cancer diagnoses were sourced from linkages with the national cancer registry and only the first cancer diagnosis was considered; available cancer data covered diagnoses up until 2018-12-31. As non-melanoma skin cancer is imperfectly registered and primarily managed in primary care, diagnoses of non-melanoma skin cancer were not considered in this study.

The cancer groups considered were:

- Breast cancer (women only), including invasive breast and in-situ breast cancers
- Gynaecological cancer (women only), including invasive cervical, in-situ cervical, ovarian, uterine, and vulvar cancers
- Lung, including lung cancer and mesothelioma
- Upper gastrointestinal (GI), including liver, oesophageal, pancreatic and stomach cancers
- Lower GI, including colon and rectal cancers
- Urological, including bladder, in-situ bladder, kidney and other urinary tract cancers
- Prostate cancer (men only)
- Haematological, including Hodgkin lymphoma, non-Hodgkin lymphoma, acute myeloid leukaemia, chronic lymphocytic leukaemia, other leukaemias, myeloma, and other haematological cancers
- Other, including all other sites, specifically including melanoma, unknown primary, thyroid, and meningeal cancers, also including testicular cancer and male breast cancer

The first outcome (of cancer diagnosis or non-cancer death) experienced by each patient was considered in the analysis. This means, for example, that in the analyses of cumulative incidence a patient who died shortly following a cancer diagnosis would only be considered to have had a cancer diagnosis, and their death would not contribute to the estimation of mortality risk irrespective of cause of death. Patients with a cancer diagnosis on the same day as their death (including, for example, death certificate only registrations of cancer) were treated as having had a cancer diagnosis rather than having died, noting that death certificate only registrations remained <0.4% through the study period [15].

### Symptoms

We considered a subset of symptoms known to have an association with risk of specific types of cancer and that are already included in referral guidelines for symptomatic cancer [7,16]. The included symptoms form part of the presentation in 40% of all patients with cancer England [1]. We identified symptoms from coded primary care data using existing Read v2 phenotyping algorithms [16]. The symptoms we considered were:

- Abdominal symptoms

- Abdominal pain
- Abdominal bloating
- Rectal bleeding
- Change in bowel habit
- Dyspepsia
- Dysphagia
- Jaundice
- Respiratory symptoms

- Dyspnoea
- Haemoptysis
- Urological symptoms

- Haematuria
- Non-specific symptoms

- Fatigue
- Night sweats
- Weight loss
- Breast and reproductive organ symptoms

- Breast lump (including in men)
- Post-menopausal bleeding

Only the first presenting symptom for each patient was included, and each patient was included at most once in the analysis. For example, if a patient had a consultation for breast lump in 2007 that did not result in a cancer diagnosis and a consultation for abdominal pain in 2010 that did result in a cancer diagnosis, only the risk after the 2007 consultation for breast lump would be included in analysis. If two or more of the examined symptoms presented on the same day, all were included as index symptoms (such occurrences were rare, see end of Results).

### Smoking status, sex, and age

Patients were categorised as ever-smokers or never-smokers. Ever-smokers included all patients with a record of being a current or ex-smokers in their entire primary care record, including periods after cancer diagnosis or before their record became eligible for use in this study; never-smokers included all other patients. Patients were classed as male or female based on the recorded gender in their primary care record. Patients’ age was estimated as the number of years between the mid-point of their year of birth and their index date.

### Statistical methods

Initial analysis described the distribution of patients in the sample and counts of cancer diagnoses and deaths within 12 months of any index symptom.

Hazards for specific cancers and non-cancer mortality were estimated using semi-parametric (Royston-Parmar) time-to-event models [17]. Follow-up for these analyses was censored at the earliest of 18 months after the index symptom, at first event (i.e., cancer diagnosis or death), or at the end of the available cancer registry follow-up on 2018-12-31. Models were stratified by sex and included the following covariates:

- Age (restricted cubic spline with six knots)
- Smoking status (binary, ever record of smoking in primary care data vs never)
- Index symptom (15 binary variables indicating the symptom(s) each patient had on their index date (all zero for patients in the reference group))
- An interaction with follow-up time in months for each index symptom, allowing the association between symptom and cause-specific risk to decay over time. This was motivated by the fact that following many possible symptoms of cancer, excess risk is highest in the first months following presentation (e.g., [18])

Cumulative incidence of cancer group and non-cancer mortality was estimated by combining each of the cause-specific models using the latent failure time approach [19]. We report cumulative incidence for combinations of age-sex-smoking-symptom up to 12 months follow-up, with results focusing on estimated cumulative incidence at 12 months and age considered in five-year intervals. To sense-check these model-based estimates, we additionally examined the crude cumulative incidence for each cancer group and non-cancer mortality within 12 months of each symptom by sex and smoking status using Aalen-Johansen non-parametric cumulative incidence curves [20,21].

Concordant with the methods and evidence that informed the development of NICE guidelines, we have considered the modelled cumulative incidence at 12 months to represent the positive predictive value for the outcome for the symptom [7]. Further, we calculated the (sex/smoking/symptom-specific) age at which cancer risk exceeded the 3% risk threshold for referrals used in the UK. We additionally present similar estimates for each individual cancer group.

Statistical modelling used Stata 17 MP. Simulation of failure times was performed on a high-performance cluster using Stata 16 MP. Survival models were fit using the *merlin* package [22], and multistate modelling was facilitated by the *multistate* package [23]. Data extraction and analysis code are available at https://github.com/MattEBarclay/cprd_symptom_cancer_1.

### Patient and public involvement

The study forms part of a programme of work examining the predictive value of symptoms for cancer diagnosis using electronic health records data. To support this programme, we ran three focus groups in August and September 2023 including a total of 15 patient and public involvement volunteers. Study reporting was informed by PPI input, but no specific changes were made.

## Results

The analysis cohort included 1,622,419 patients, 835,995 with an eligible first symptom recorded between 2007 and 2017 (Table 1). More than half of the cohort (64%, 1,040,762) were aged under 60 at index, with 24,731 (1.5%) patients aged 90 or older. The distribution of symptoms was uneven, with 14.4% of the cohort having abdominal pain as index symptom, followed by dyspnoea (8.7%), fatigue (8.1%), dyspepsia (6.7%), rectal bleeding (3.0%), breast lump (2.4%), haematuria (1.6%), abdominal bloating (1.4%), weight loss (1.2%), change in bowel habit (1.1%), dysphagia (0.9%), post-menopausal bleeding (0.5%), night sweats (0.5%), haemoptysis (0.4%), and jaundice (0.1%). The majority of patients (64%) had at least one smoking-related Read code in their records and were identified as ever-smokers. Within 12 months of their first recorded symptom, 36,802 patients had a cancer diagnosis and 28,867 patients died without a cancer diagnosis (a further 9,288 died following a cancer diagnosis); both cancer and mortality risk were higher in older patients. Ever-smokers had slightly higher cancer risk than patients without any smoking-related codes.

**Table 1.** Cohort summary.

|  | Cohort |  | Cancers within 12 months |  | Deaths within 12 months, no preceding cancer diagnosis |  | Deaths within 12 months, following a cancer diagnosis |  |
| --- | --- | --- | --- | --- | --- | --- | --- | --- |
|  | N | (col %) | N | (row %) | N | (row %) | N | (row %) |
| **Total** | 1,622,419 |  | 36,802 | (2.3%) | 28,867 | (1.8%) | 9,288 | (0.6%) |
| **Age at index (grouped)** |  |  |  |  |  |  |  |  |
| 30 to 39 | 395,313 | (24.4%) | 1,571 | (0.4%) | 426 | (0.1%) | 62 | (0.0%) |
| 40 to 49 | 350,133 | (21.6%) | 3,063 | (0.9%) | 792 | (0.2%) | 235 | (0.1%) |
| 50 to 59 | 295,316 | (18.2%) | 5,080 | (1.7%) | 1,343 | (0.5%) | 762 | (0.3%) |
| 60 to 69 | 259,039 | (16.0%) | 9,014 | (3.5%) | 2,829 | (1.1%) | 1,970 | (0.8%) |
| 70 to 79 | 185,854 | (11.5%) | 10,142 | (5.5%) | 6,007 | (3.2%) | 2,960 | (1.6%) |
| 80 to 89 | 111,933 | (6.9%) | 6,818 | (6.1%) | 11,453 | (10.2%) | 2,720 | (2.4%) |
| 90 to 99 | 24,731 | (1.5%) | 1,114 | (4.5%) | 6,017 | (24.3%) | 579 | (2.3%) |
| **Sex** |  |  |  |  |  |  |  |  |
| Women | 880,888 | (54.3%) | 19,808 | (2.2%) | 15,671 | (1.8%) | 4,259 | (0.5%) |
| Men | 741,531 | (45.7%) | 16,994 | (2.3%) | 13,196 | (1.8%) | 5,029 | (0.7%) |
| **IMD group** |  |  |  |  |  |  |  |  |
| Least deprived | 377,575 | (23.3%) | 8,934 | (2.4%) | 5,661 | (1.5%) | 2,001 | (0.5%) |
| 2 | 356,859 | (22.0%) | 8,347 | (2.3%) | 6,177 | (1.7%) | 2,031 | (0.6%) |
| 3 | 342,184 | (21.1%) | 7,755 | (2.3%) | 6,355 | (1.9%) | 1,889 | (0.6%) |
| 4 | 294,638 | (18.2%) | 6,483 | (2.2%) | 5,559 | (1.9%) | 1,805 | (0.6%) |
| Most deprived | 251,163 | (15.5%) | 5,283 | (2.1%) | 5,115 | (2.0%) | 1,562 | (0.6%) |
| **Any record of smoking** |  |  |  |  |  |  |  |  |
| Never smoker | 586,639 | (36.2%) | 10,390 | (1.8%) | 10,043 | (1.7%) | 2,259 | (0.4%) |
| Ever smoker | 1,035,780 | (63.8%) | 26,412 | (2.5%) | 18,824 | (1.8%) | 7,029 | (0.7%) |
| **Index symptom** |  |  |  |  |  |  |  |  |
| Reference group | 786,424 | (48.5%) | 7,536 | (1.0%) | 12,520 | (1.6%) | 2,034 | (0.3%) |
| Abdominal pain | 233,933 | (14.4%) | 5,605 | (2.4%) | 2,163 | (0.9%) | 1,640 | (0.7%) |
| Abdominal bloating | 22,629 | (1.4%) | 628 | (2.8%) | 261 | (1.2%) | 169 | (0.7%) |
| Rectal bleeding | 48,515 | (3.0%) | 1,868 | (3.9%) | 860 | (1.8%) | 220 | (0.5%) |
| Change in bowel habit | 17,212 | (1.1%) | 1,067 | (6.2%) | 163 | (0.9%) | 197 | (1.1%) |
| Dyspepsia | 108,488 | (6.7%) | 2,120 | (2.0%) | 959 | (0.9%) | 609 | (0.6%) |
| Dysphagia | 14,992 | (0.9%) | 1,036 | (6.9%) | 1,167 | (7.8%) | 451 | (3.0%) |
| Jaundice | 1,817 | (0.1%) | 456 | (25.1%) | 217 | (11.9%) | 280 | (15.4%) |
| Dyspnoea | 141,094 | (8.7%) | 3,945 | (2.8%) | 6,268 | (4.4%) | 1,490 | (1.1%) |
| Haemoptysis | 5,859 | (0.4%) | 412 | (7.0%) | 146 | (2.5%) | 183 | (3.1%) |
| Haematuria | 25,753 | (1.6%) | 2,770 | (10.8%) | 591 | (2.3%) | 378 | (1.5%) |
| Fatigue | 141,932 | (8.7%) | 2,405 | (1.7%) | 2,212 | (1.6%) | 739 | (0.5%) |
| Night sweats | 7,675 | (0.5%) | 133 | (1.7%) | 30 | (0.4%) | 35 | (0.5%) |
| Weight loss | 19,617 | (1.2%) | 1,238 | (6.3%) | 1,173 | (6.0%) | 623 | (3.2%) |
| Breast lump | 38,307 | (2.4%) | 4,789 | (12.5%) | 88 | (0.2%) | 185 | (0.5%) |
| Post-menopausal bleed | 8,172 | (0.5%) | 794 | (9.7%) | 49 | (0.6%) | 55 | (0.7%) |

### Age-adjusted cancer-specific hazard ratios for smoking and each index symptom

Both male and female ever-smokers had far higher cancer-specific hazard of lung cancer than non-smokers (Figure 1 and Appendix 4, HR 4.8, 95%CI 4.2-5.6, for women and HR 4.0, 95%CI 3.5-4.6, for men), and elevated hazards of urological (e.g., for men: HR 1.4, 95%CI 1.2-1.5, Appendix 4 Table 4) and upper GI cancers (e.g., for men: HR 1.4, 95%CI 1.2-1.5, Appendix 4 Table 1).

**Figure 1.**
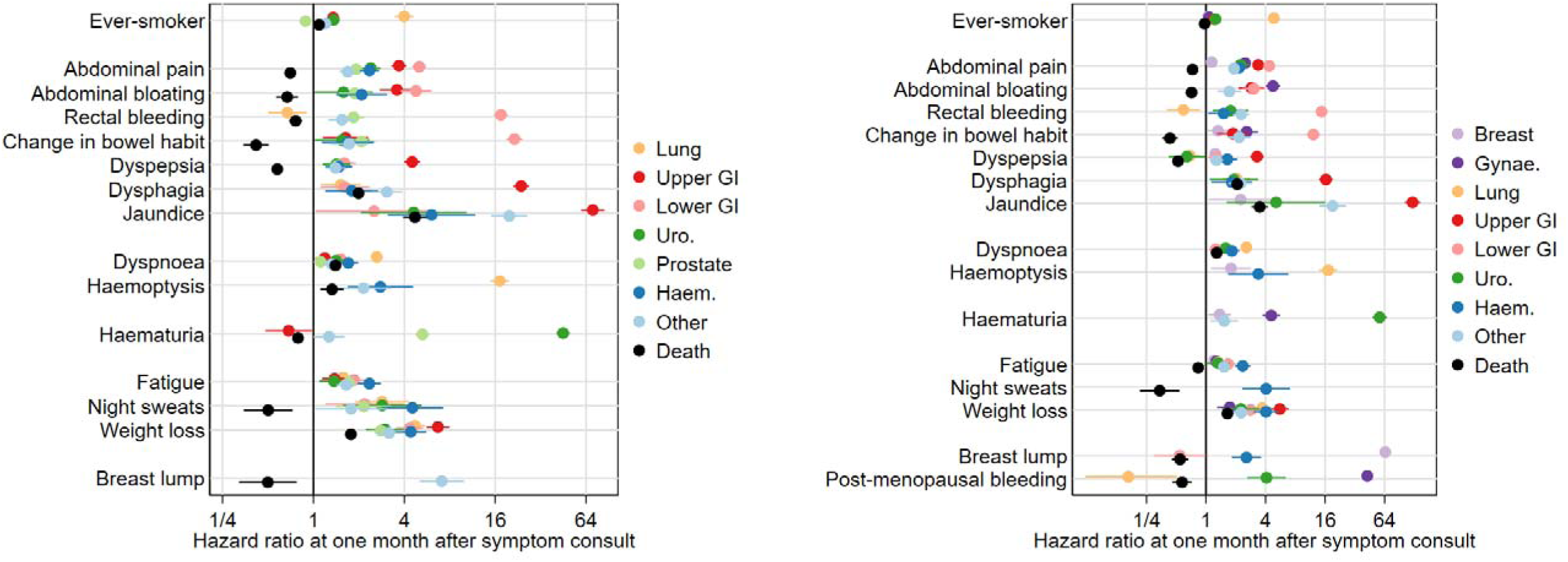
Hazard ratios for each cancer site and for non-cancer death at one month after index, for men (left) and women (right). Ever-smoker is compared to never-smoker; each symptom is compared to the control group. Models are stratified by sex, and adjusted for age, smoking status, and the presence of symptoms at index date.

Patients consulting for symptoms of possible cancer had similar or greater cause-specific hazards for almost every cancer site than the reference population (Figure 1 and Appendix 4). Yet for ten of the fifteen studied symptoms, the symptom was associated with lower cause-specific hazards for death than the reference group (the exceptions being dysphagia, jaundice, dyspnoea, haemoptysis, and weight loss). Further, for many symptoms associated with very high initial hazard of a specific cancer, while the hazard typically remained elevated at least to 12 months after the index consultation, it tended to reduce over time (Figure 1).

### Abdominal symptoms (abdominal pain, abdominal bloating, rectal bleeding, change in bowel habit, dyspepsia, dysphagia, jaundice)

For both men and women presentations with abdominal symptoms were associated with increased hazard of multiple types of cancer. At the same time, abdominal symptoms were associated with decreased hazard of death without a cancer diagnosis when compared with the reference group, except for dysphagia and jaundice (Figure 1, and Appendix 4 Tables 2-3 and 12-13). Cause-specific hazard ratios at one month after presentation were highest regarding lower GI cancer for rectal bleeding and change in bowel habit (e.g., CIBH for men: HR 17.4, 95% CI 15.7-19.4) and highest regarding upper GI cancer for jaundice and dysphagia (e.g., dysphagia in women: HR 16.4, 95%CI 14.0-19.2); hazard ratios decreased substantially over follow-up for these symptoms. Abdominal pain and abdominal bloating were associated with hazard ratios at consultation of around 4 for both upper and lower GI cancers (e.g., abdominal bloating in women with HR for lower GI cancer of 3.0, 95%CI 2.3-4.0), with abdominal bloating having a similar association for gynaecological cancers in women (HR 4.8, 95%CI 4.0-5.6), while dyspepsia was associated with a hazard ratio of around 4 for upper GI cancer. Patients with abdominal symptoms also appeared at elevated risk for urological and haematological cancers, and for prostate and gynaecological cancers.

### Respiratory symptoms (dyspnoea, haemoptysis)

Respiratory symptoms were primarily associated with lung cancer, but the strength of the association varied (Figure 1 and Appendix 4 Tables 1 and 11). Patients with haemoptysis had a cause-specific hazard ratio of around 16 at consultation compared with the reference group (e.g., for men, HR 17.1, 95%CI 14.8-19.8), while the association with dyspnoea was weaker but still notable (e.g., for men, HR 2.6, 95%CI 2.4-2.9). Other types of cancer, notably haematological cancers, also had elevated cause-specific hazards; (e.g., for men, the HR for haematological cancer being 2.8, 95%CI 1.7-4.6, Appendix 4 Tables 6 and 15).

### Urological symptoms (Haematuria)

Haematuria in women was primarily associated with urological cancers (HR 57, 95%CI 48-67) and with gynaecological cancers (HR 4.6, 95%CI 3.7-5.6) (Figure 1 and Appendix 4 Tables 10 and 14). In men, it was associated with urological cancers (HR 45, 95%CI 40-50) and prostate cancer (HR 5.3, 95%CI 4.8-5.8) (Appendix 4, Tables 4 and 5).

### Non-specific symptoms (Fatigue, night sweats, weight loss)

Non-specific symptoms were typically associated with elevated cause-specific hazard ratios for all cancer groups considered (Figure 1 and Appendix 4), and generally HRs appeared relatively similar in strength for each of the three non-specific symptoms. Weight loss had the strongest associations overall (cancer-specific HRs general between 2 and 5), followed by night sweats (HRs generally between 1 and 4, though imprecisely estimated), followed by fatigue (HRs between 1 and 2). It often appeared that the strongest cause-specific associations were for haematological cancers, though confidence intervals tended to overlap with those of other cancer groups.

*Breast and reproductive organ symptoms (breast lump, post-menopausal bleeding)* Post-menopausal bleeding was associated with large cause-specific hazard ratios for gynaecological cancer (HR 43, 95%CI 39-47) and substantial cause-specific HRs for urological cancer (HR 4.1, 95%CI 2.6-6.4) (Figure 1 and Appendix 4 Tables 10 and 14).

Breast lump in women was associated principally with breast cancer (HR 65, 95%CI 61-69) and to a lesser extent with haematological cancer (HR 2.6, 95%CI 1.80-3.6) (Appendix 4 Tables 9 and 15). A small number of men present with breast lump, and these men had cause-specific hazard ratios for the ‘other cancers’ group, which included male breast cancer, of 7.1 (95%CI 5.0-10.0) (Appendix 4 Table 7).

### Risk of specific cancer sites by age, sex, and smoking status

After symptom presentation for patients with single index symptoms, and based on simulations combining the cause-specific models, we present simulated cumulative incidence of each cancer site and of death without cancer at 3 months (Appendix 2), 6 months (Appendix 3), and 12 months (Figures 2-5, Appendix 5). Hereafter in this section, we discuss cumulative incidence at 12 months after symptom consultation. Unlike the hazard ratios presented above, estimates of cumulative incidence varied substantially by sex, as women have lower baseline cancer risk.

**Figure 2.**
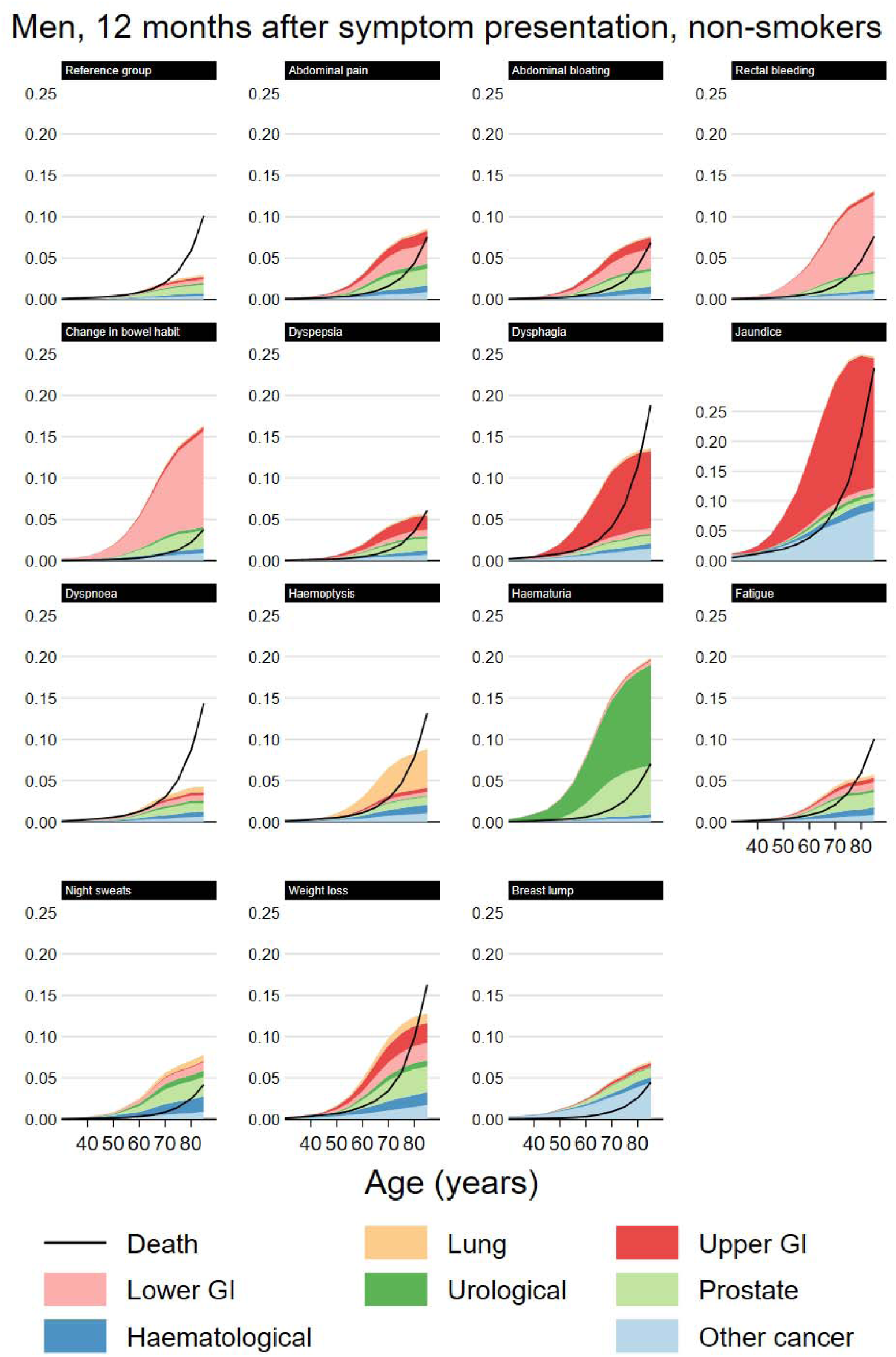
Modelled cancer and mortality risk at 12 months by index symptom, male non-smokers.

**Figure 3.**
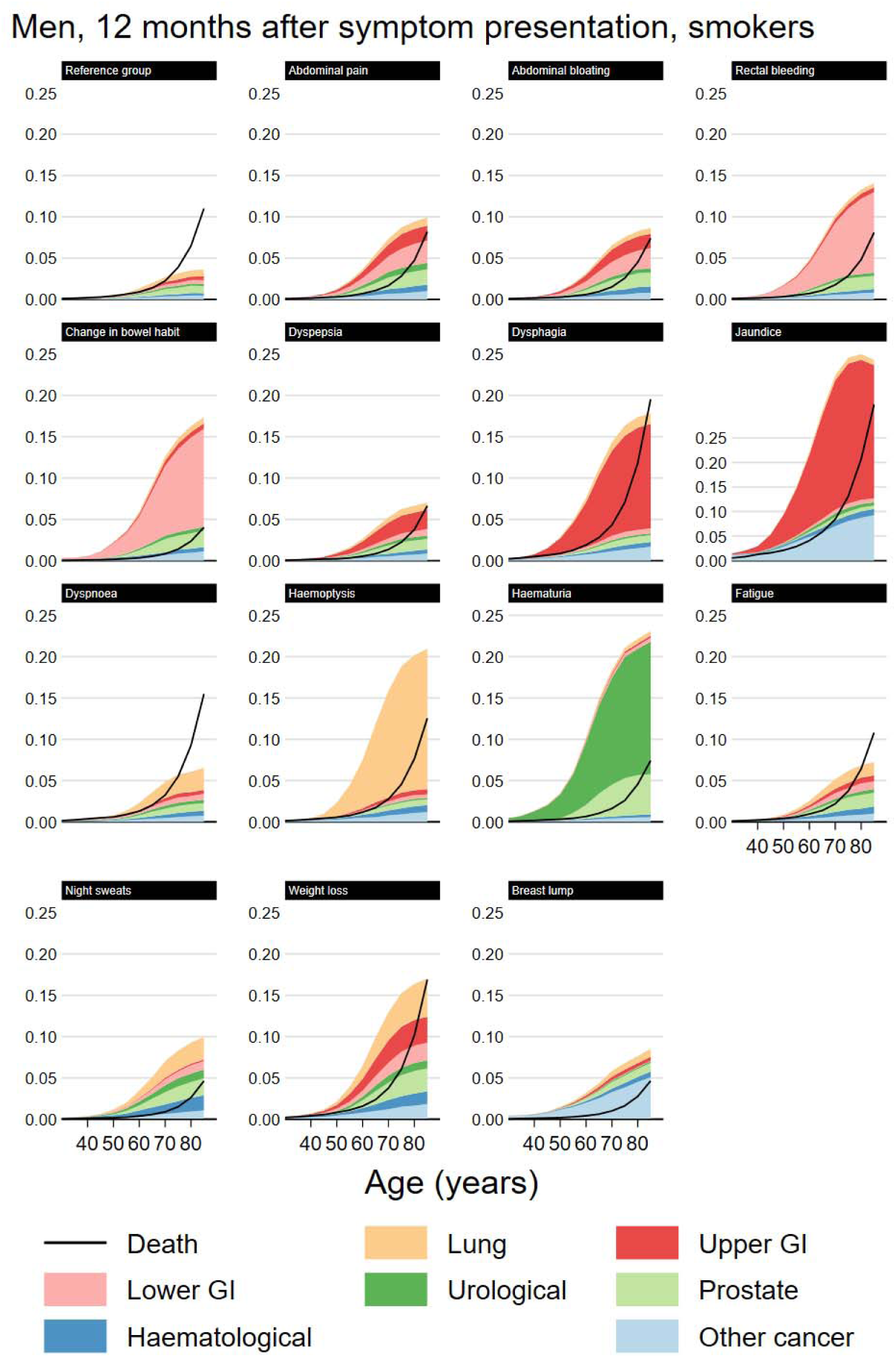
Modelled cancer and mortality risk at 12 months by index symptom, male smokers.

**Figure 4.**
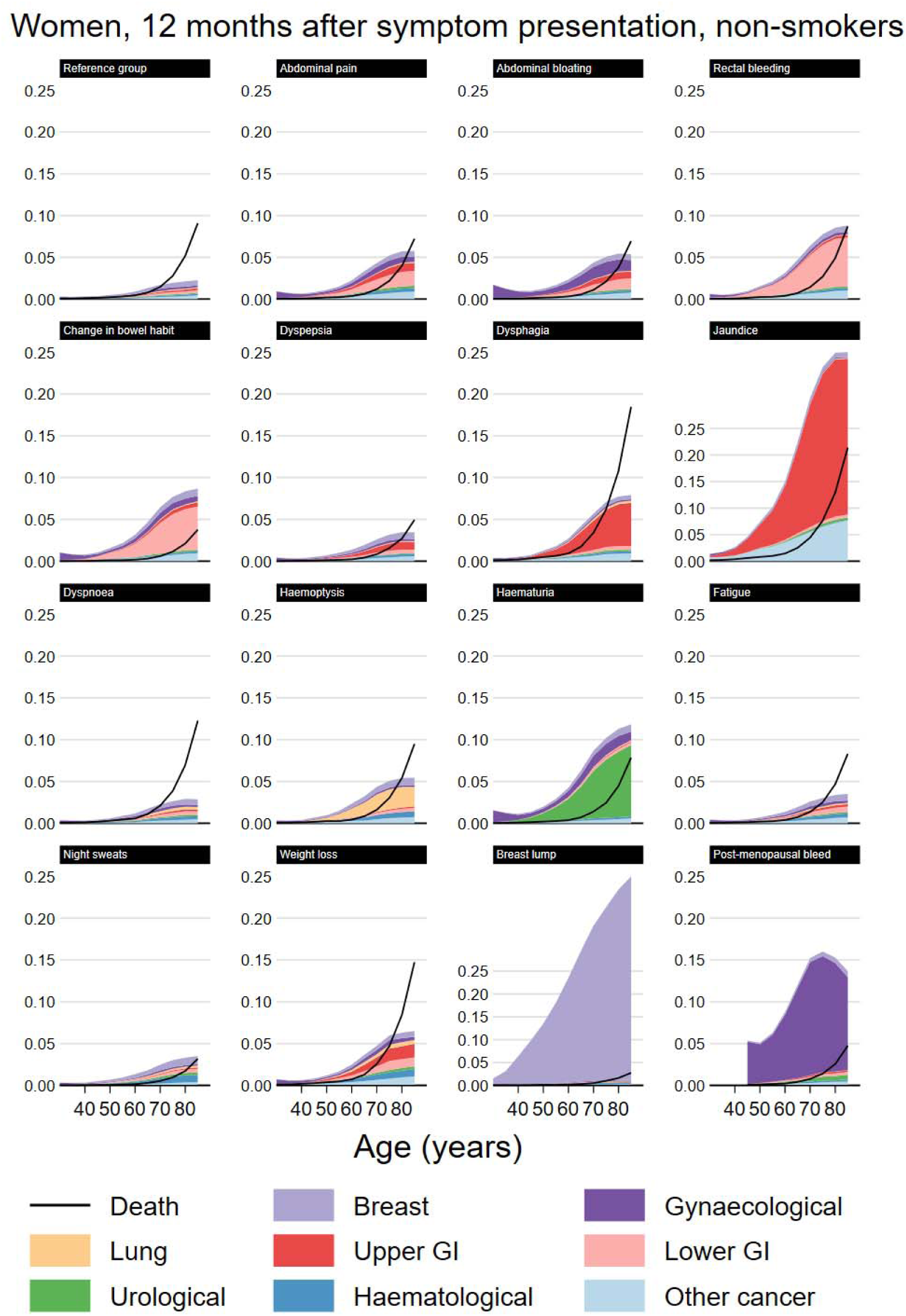
Modelled cancer and mortality risk at 12 months by index symptom, female non-smokers.

**Figure 5.**
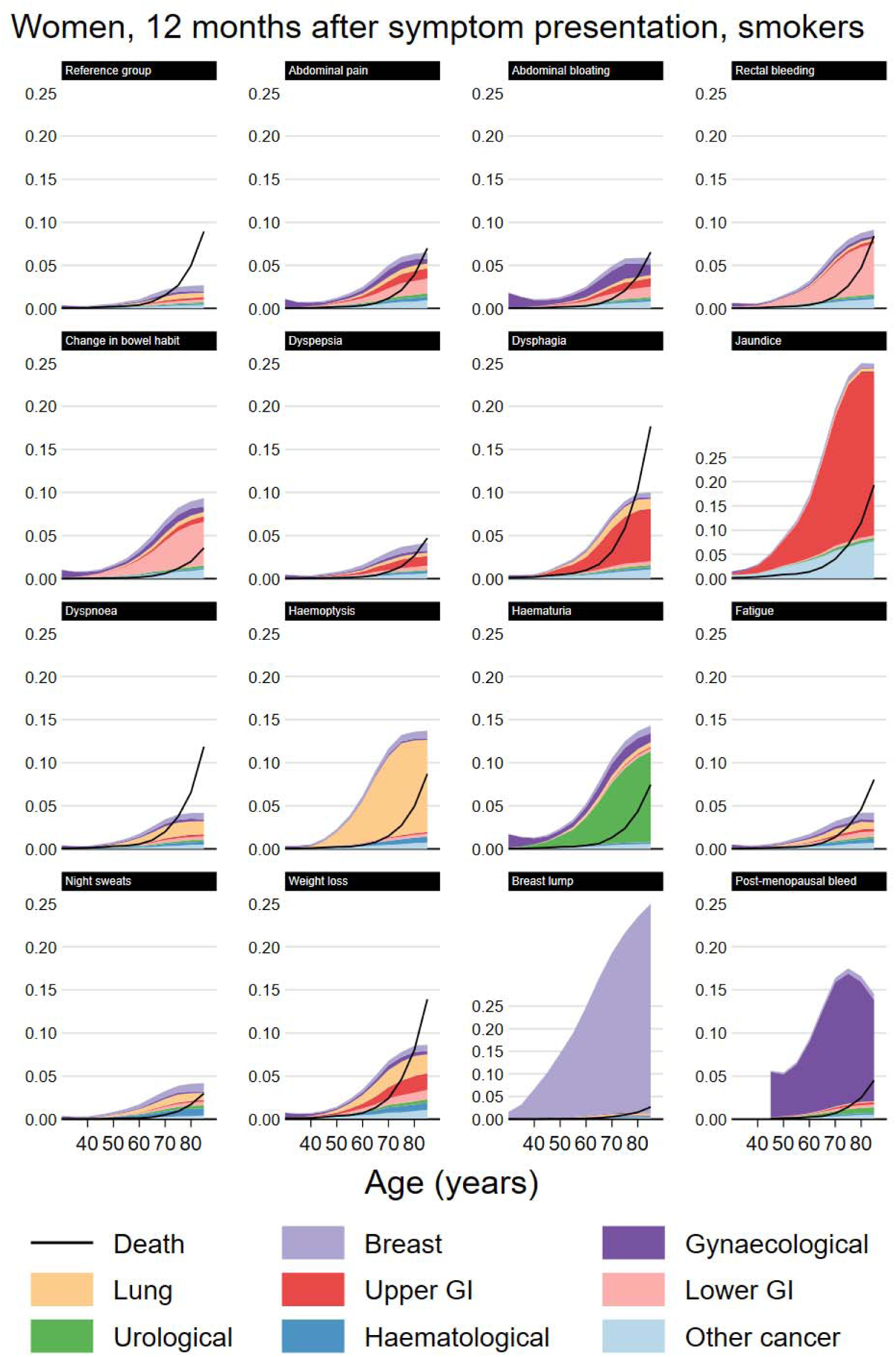
Modelled cancer and mortality risk at 12 months by index symptom, female smokers.

### 3% any cancer risk thresholds at 12 months

Patients reaching a 3% risk of any cancer may not reach such a risk level for any specific cancer group, especially for symptoms associated with multiple types of cancer. For example, female smokers presenting with weight loss had a 3% risk of cancer from age 60, but did not reach the 3% risk threshold at any age when any of the individual cancer groups were considered on their own (Table 2). For male non-smokers, risk of any cancer reached the 3% threshold from the following ages and onwards: 45 for jaundice; 55 for dysphagia, weight loss, haematuria, and change in bowel habit; 60 for haemoptysis and rectal bleeding; 65 for abdominal pain and bloating, night sweats and breast lump; and 70 for dyspepsia, dyspnoea, and fatigue (Table 2). For smokers, this threshold was often reached up to five years younger. Conversely, compared with male patients presenting with the same symptom, female patients reached the 3% threshold at an older age on average, with the main exception being breast lump for which the 3% threshold (in women) was reached from age 40.

**Table 2.** Modelled age at which patients presenting with each symptom had a 3%-risk (i.e., high enough to trigger urgent referral for suspected cancer in England) of all cancers combined and of specific cancer sites, by smoking status and sex.

| Men | Never-smokers | Ever-smokers |
| --- | --- | --- |
| Reference group | Any cancer (90) | Any cancer (75) |
| Abdominal pain | Any cancer (60) | Any cancer (60) |
| Abdominal bloating | Any cancer (65) | Any cancer (60) |
| Rectal bleeding | Any cancer (60); lower GI (65) | Any cancer (60); lower GI (60) |
| Change in bowel habit | Any cancer (55); lower GI (60) | Any cancer (55); lower GI (60) |
| Dyspepsia | Any cancer (65) | Any cancer (65) |
| Dysphagia | Any cancer (55); upper GI (60) | Any cancer (55); upper GI (55) |
| Jaundice | Any cancer (45); upper GI (50); other (55) | Any cancer (45); upper GI (50); other (55) |
| Dyspnoea | Any cancer (70) | Any cancer (65) |
| Haemoptysis | Any cancer (60); lung (70) | Any cancer (55); lung (55) |
| Haematuria | Any cancer (55); urological (55); prostate (65) | Any cancer (50); urological (55); prostate (70) |
| Fatigue | Any cancer (65) | Any cancer (65) |
| Night sweats | Any cancer (65) | Any cancer (60) |
| Weight loss | Any cancer (60); prostate (80) | Any cancer (55); lung (70); upper GI (75) |
| Breast lump | Any cancer (65); other (75) | Any cancer (60); other (70) |
| Women | Never-smokers | Ever-smokers |
| Reference group |  |  |
| Abdominal pain | Any cancer (65) | Any cancer (65) |
| Abdominal bloating | Any cancer (65) | Any cancer (65) |
| Rectal bleeding | Any cancer (60); lower GI (70) | Any cancer (60); lower GI (70) |
| Change in bowel habit | Any cancer (60); lower GI (70) | Any cancer (60); lower GI (70) |
| Dyspepsia | Any cancer (75) | Any cancer (70) |
| Dysphagia | Any cancer (65); upper GI (70) | Any cancer (60); upper GI (70) |
| Jaundice | Any cancer (45); upper GI (50); other (60) | Any cancer (40); upper GI (45); other (55) |
| Dyspnoea |  | Any cancer (70) |
| Haemoptysis | Any cancer (65) | Any cancer (55); lung (60) |
| Haematuria | Any cancer (60); urological (65) | Any cancer (55); urological (60) |
| Fatigue | Any cancer (75) | Any cancer (70) |
| Night sweats | Any cancer (75) | Any cancer (70) |
| Weight loss | Any cancer (65) | Any cancer (60) |
| Breast lump | Any cancer (35); breast (40) | Any cancer (35); breast (35) |
| Post-menopausal bleeding | Any cancer (30); gynaecological (30) | Any cancer (30); gynaecological (30) |

Notably, male smokers in the reference group had a 3% risk of any cancer from age 75, and male non-smokers from age 90; women in the reference group did not reach a 3% risk of cancer at any age.

A summary of risk of individual cancers is given in Appendix 6, plus additional graphical and tabular results in Appendices 3 and 5.

### Risk of non-cancer mortality

For most of the studied symptoms, symptomatic patients were less likely to die (without a cancer diagnosis) than similar patients in the reference group (Figures 2-5). The three principal exceptions were jaundice, dysphagia and weight loss, for which post-presentation mortality exceeded that in the reference group, and also older patients with less-specific symptoms for whom the risk of non-cancer mortality was often higher than the risk of any cancer. For example, for male smokers presenting with dyspnoea, around 6% who presented at age 80 would develop cancer within 12 months while 9% would die (Figure 3, Appendix 5 Table 1).

### Presentation with multiple symptoms

Among symptomatic patients, 1.2% (10,360 of 835,995) consulted for more than one of the fifteen studied symptoms on their index date, and a further 2.5% (21,167) consulted for an additional studied symptom within 30 days of an index symptom but before a cancer diagnosis (Table 3). The proportion of patients with multiple index symptoms subsequently diagnosed with cancer within 12 months of index (4.6%, 95% CI 4.2% to 5.1%) was higher than for patients with a single index symptom (3.5%, 95% CI 3.5% to 3.5%). This higher risk of cancer in patients with multiple index symptoms appeared applicable to many of the symptoms considered, but sample size limitations meant proportions developing cancer could often not be estimated precisely.

**Table 3.** Summary of cancer outcomes for patients with multiple different recorded symptoms at index presentation, and within 30 days of index symptom.

| Index symptom | Any other symptoms at index | Patients | Cancers within 12 months of index |  |  |
| --- | --- | --- | --- | --- | --- |
|  |  |  | N | % | (95% CI) |
| Any | No | 825,635 | 28,834 | 3.5% | (3.5%, 3.5%) |
|  | Yes | 10,360 | 480 | 4.6% | (4.2%, 5.1%) |
|  | Within 30 days* | 21,167 | 1429 | 6.8% | (6.4%, 7.1%) |
| Abdominal pain | No | 231,598 | 5,510 | 2.4% | (2.3%, 2.4%) |
|  | Yes | 2,335 | 101 | 4.3% | (3.6%, 5.2%) |
|  | Within 30 days* | 6,122 | 379 | 6.2% | (5.6%, 6.8%) |
| Abdominal bloating | No | 825,635 | 28,834 | 3.5% | (3.5%, 3.5%) |
|  | Yes | 10,360 | 480 | 4.6% | (4.2%, 5.1%) |
|  | Within 30 days* | 21,167 | 1429 | 6.8% | (6.4%, 7.1%) |
| Rectal bleeding | No | 47,774 | 1,831 | 3.8% | (3.7%, 4.0%) |
|  | Yes | 741 | 38 | 5.1% | (3.8%, 7.0%) |
|  | Within 30 days* | 1,116 | 61 | 5.5% | (4.3%, 7.0%) |
| Change in bowel habit | No | 16,857 | 1,042 | 6.2% | (5.8%, 6.6%) |
|  | Yes | 355 | 25 | 7.0% | (4.8%, 10.2%) |
|  | Within 30 days* | 520 | 77 | 14.8% | (12.0%, 18.1%) |
| Dyspepsia | No | 106,843 | 2,090 | 2.0% | (1.9%, 2.0%) |
|  | Yes | 1,645 | 35 | 2.1% | (1.5%, 2.9%) |
|  | Within 30 days* | 3,282 | 219 | 6.7% | (5.9%, 7.6%) |
| Dysphagia | No | 14,760 | 1,021 | 6.9% | (6.5%, 7.3%) |
|  | Yes | 232 | 17 | 7.3% | (4.6%, 11.4%) |
|  | Within 30 days* | 1,054 | 56 | 5.3% | (4.1%, 6.8%) |
| Jaundice | No | 1,759 | 450 | 25.6% | (23.6%, 27.7%) |
|  | Yes | 58 | 9 | 15.5% | (8.4%, 26.9%) |
|  | Within 30 days* | 81 | 17 | 21.0% | (13.5%, 31.1%) |
| Dyspnoea | No | 139,758 | 3,899 | 2.8% | (2.7%, 2.9%) |
|  | Yes | 1,336 | 61 | 4.6% | (3.6%, 5.8%) |
|  | Within 30 days* | 2,655 | 173 | 6.5% | (5.6%, 7.5%) |
| Haemoptysis | No | 5,750 | 406 | 7.1% | (6.4%, 7.8%) |
|  | Yes | 109 | 6 | 5.5% | (2.5%, 11.5%) |
|  | Within 30 days* | 198 | 20 | 10.1% | (6.6%, 15.1%) |
| Haematuria | No | 25,438 | 2,749 | 10.8% | (10.4%, 11.2%) |
|  | Yes | 315 | 22 | 7.0% | (4.7%, 10.3%) |
|  | Within 30 days* | 636 | 76 | 12.0% | (9.7%, 14.7%) |
| Fatigue | No | 140,212 | 2,353 | 1.7% | (1.6%, 1.7%) |
|  | Yes | 1,720 | 58 | 3.4% | (2.6%, 4.3%) |
|  | Within 30 days* | 3,132 | 157 | 5.0% | (4.3%, 5.8%) |
| Night sweats | No | 7,527 | 128 | 1.7% | (1.4%, 2.0%) |
|  | Yes | 148 | 5 | 3.4% | (1.5%, 7.7%) |
|  | Within 30 days* | 162 | 6 | 3.7% | (1.7%, 7.8%) |
| Weight loss | No | 19,168 | 1,193 | 6.2% | (5.9%, 6.6%) |
|  | Yes | 449 | 52 | 11.6% | (8.9%, 14.9%) |
|  | Within 30 days* | 725 | 90 | 12.4% | (10.2%, 15.0%) |
| Breast lump | No | 38,045 | 4,765 | 12.5% | (12.2%, 12.9%) |
|  | Yes | 262 | 25 | 9.5% | (6.5%, 13.7%) |
|  | Within 30 days* | 345 | 18 | 5.2% | (3.3%, 8.1%) |
| Post-menopausal bleed | No | 8,092 | 784 | 9.7% | (9.1%, 10.4%) |
|  | Yes | 80 | 10 | 12.5% | (6.9%, 21.5%) |
|  | Within 30 days* | 145 | 14 | 9.7% | (5.8%, 15.6%) |

\*subset of patients with no other symptoms at index

The cause-specific time-to-event models accommodated multiple index symptoms that were consulted for on the same day, so for example the cause-specific hazard ratio for upper GI cancer for abdominal pain is already adjusted for the presence of dysphagia, for the infrequent occasions (see above) where both were recorded – although possible interaction effects were not considered. Symptoms that were not consulted for on the same day as index were not considered. In principle, estimates of cancer risk for any combination of symptoms can be estimated from the cause-specific models, but these have not been produced due to computational limitations and the very large number of potential combinations.

## Discussion

Using a cohort design, we comprehensively estimated the risk of different cancer diagnoses and non-cancer mortality following presentation in primary care with one of 15 index symptoms, and in a reference group that was not selected based on symptom status and so should approximate the risk in the general population. There was considerable variation in risk by age and by sex. Smoking-status was highly informative for cancer risk for patients with respiratory or non-organ-specific symptoms. Smokers typically reached the 3% threshold warranting referral for cancer investigations up to five years younger than non-smokers. The findings highlight the importance of including smoking status in clinical guidelines and referral decisions in patients with a new onset symptom. Even symptoms with strong, well-established associations (e.g., dyspnoea and lung cancer) have notable associations with other types of cancer (e.g., haematological cancers). We also provide estimates of cancer risk while considering the potential for non-cancer mortality. For the oldest patients – and for those with symptoms such as dysphagia or jaundice – risk of death without a cancer diagnosis reached or exceeded the risk of cancer. Referral decisions based on a universally applied 3% cancer risk threshold, as currently set out in UK clinical guidelines, may not be appropriate for these patients.

### Strengths and weaknesses

Key strengths of the study are (a) the large representative dataset – allowing examination of a range of both common and rare symptoms and outcomes – (b) the joint estimation of the risks of the different outcomes, including of non-cancer mortality and risk of different types of cancer, and (c) the use of cancer registry data to ascertain presence of cancer, as cancer may be under- or over-recorded in non-registry sources [24]. While this study represents the most comprehensive and detailed description of risk of cancer in symptomatic patients to date, there are various areas where future work could make further improvements.

Considering limitations, the study only considers deaths in patients without cancer, but it may be important to understand if patients die quickly after a cancer diagnosis. Our measure of smoking status does not allow for a refined appreciation of smoking history and dose-response relationships. Additionally, our analytical approach only allowed each patient to be included once, not making full use of the longitudinal nature of EHR datasets [25]. We did not consider interactions between symptoms and simulated outcomes for patients with a single symptom only, in part due to only few patients having multiple symptoms. We did not have access to free-text data, despite evidence that coded data does not capture all symptoms [26,27]. Finally, we only examined 15 symptoms, ignoring the many other symptoms and important health conditions that may be associated with risk of cancer [1,16,28]. A more detailed examination of potential limitations is given in Appendix 7.

### Comparison with literature

A large and growing literature describes risk of cancer following symptom presentations in primary care; Moore and colleagues summarised the literature pre-2020 [16], and there are several recent papers [18,31–33]. Existing literature (a) rarely considers competing non-cancer mortality risk, (b) rarely considers smoking status, and (c) frequently provides no or only limited information on the age-dependent and sex-specific nature of the risk of different cancers. Much of the previous evidence additionally considers either the risk of all cancers combined or focuses on specific cancer sites judged to be of relevance to the specific examined symptoms *a priori*. We improve on previous descriptive studies by presenting a broad range of possible cancer diagnoses following presentation with wider spectrum of index symptoms. Further research is needed to extend analyses similar to those reported here to a wider collection of symptoms.

Some existing evidence on so-called red flag symptoms such as rectal bleeding and haemoptysis suggests the risk of cancer exceeds 3% for all ages, but did not examine the risk in different age groups [16]; our findings indicate that risk of cancer following these symptoms only exceeds 3% beyond certain age cut-offs. Furthermore, we show that for non-specific symptoms, the risk of any cancer exceeds 3% at a considerably earlier age than the risk of a specific cancer type, underscoring the need for studies that comprehensively examine all major cancer types. Weight loss provides a cardinal example, where risk of any cancer exceeded 3% in male non-smokers from age 55 but risk of any individual site only reached 3% at age 85.

Other studies have aimed to develop risk prediction tools for cancer intended for use in a primary care setting (see for example, [34–36]), and in particular the QCancer risk prediction tool [9,10] already considers a range of symptoms and risk of diagnosis of different types of cancer. For decisions about the management of an individual patient, a risk prediction tool including multiple potential predictors may be more suitable than the results presented in this paper. We view our results as complementary; by describing what is effectively the average risk in patients presenting with these symptoms (by age, sex, and smoking status), we can inform high-level policy decisions around symptomatic diagnosis of cancer such as clinical guideline recommendations, and help developers of more detailed risk prediction models by highlighting symptoms they may wish to consider. Further, our consideration of mortality risk provides relevant information that is frequently missing from current risk prediction tools (including QCancer) and that is especially important in frail and elderly populations.

### Implications

Symptoms recorded in primary care data can be highly informative about both cancer risk and short-term mortality risk. In some cases, for example lung cancer, smoking-status is very strongly associated with the risk of cancer following a certain symptom. Risk of cancer and non-cancer mortality varies considerably by age; describing “overall” risk of cancer following a symptom may be misleading if non-cancer mortality is not considered. Some (non-cancer) deaths will relate to as-yet undiagnosed disease which, like cancer diagnosis, necessitates specialist assessment in secondary care, though this should be the subject of future enquiries.

For researchers, our results underline the methodological importance of accounting for the fact that symptoms may be associated with multiple different disease outcomes. Advanced statistical modelling strategies are helpful in assessing diagnostic outcomes using EHR data, and current statistical packages allow for relatively straightforward handling of competing risks either by directly modelling cumulative incidence (e.g., the Fine-Gray model [37]) or, as here, by combining several cause-specific models [38]. Diagnostic research should adopt strategies that allow consideration of risk of several potentially related diseases (e.g., multiple types of cancer, as in this study), which can be done even with simple analytical approaches such as appropriate use of logistic regression [32].

For clinicians and policy makers, our systematic assessment of risk of cancer (and of non-cancer mortality) in symptomatic patients in primary care raises two key questions.

First, whether all age-sex-smoking status groups presenting with each of the studied symptoms and with an estimated any-cancer risk of above 3% should explicitly be added to NICE referral guidelines. This may indeed be justified, though given the high mortality rates in the oldest patients, there might also be a risk of over-testing in older men in particular.

However, the degree to which risk of over-testing is a concern relates to the exact causes of non-cancer mortality and the extent to which it relates to pre-diagnosed or new non-neoplastic diseases which could benefit from specialist diagnostic assessment and earlier diagnosis. As the components of non-cancer mortality due to pre-existing or new conditions is unclear, this should be addressed by future research. The current approach to cancer referral uses a normative threshold applicable to patients of any age and with any symptoms, and the results highlight the importance of considering whether patients are likely to benefit from prompt diagnosis.

Second, whether current referral pathways are necessarily ideal. For example, many abdominal symptoms were strongly associated with lower GI, upper GI and gynaecological cancers, and some form of referral pathway offering combined multi-specialty assessment may be justified for patients with these symptoms. Further, symptoms were often strongly associated with less common cancers such as haematological neoplasms but, due to the low incidence of these conditions, absolute risk rarely or never reached 3%; optimal diagnostic management of these patients is clearly challenging. Our findings may be helpful in clarifying referral criteria for new non-specific cancer pathways.

### Conclusions

The risk of cancer diagnosis and non-cancer mortality after symptomatic presentation can be comparable and both should be considered in referral and investigation decisions – alongside age, sex, and smoking status. A holistic and stratified assessment of risk in symptomatic patients, which considers the risk of a cancer diagnosis, the risk of a diagnosis of individual types of cancer, and the risk of non-cancer mortality is needed particularly for patients presenting with which are vague or non-specific symptoms associated with multiple cancer types and appreciable non-cancer mortality risk. Our results can support the updating of referral and management guidelines for symptomatic patients presenting in primary care.

## Data Availability

Potential concerns around patient confidentiality prevent open sharing of the underlying data for this study. CPRD Gold data can be obtained from CPRD, subject to protocol approval via CPRD's Research Data Governance Process. Further details can be found at https://cprd.com/data-access. Data extraction and analysis code are available at https://github.com/MattEBarclay/cprd_symptom_cancer_1.

## Ethics statements

### Ethical approval

This study was approved by the UK Medicines and Healthcare products Regulatory Agency Independent Scientific Advisory Committee (ISAC Protocol number 18_299), under Section 251 (NHS Social Care Act 2006). This study is based on data from the Clinical Practice Research Datalink obtained under license from the UK Medicines and Healthcare products Regulatory Agency. The data is provided by patients and collected by the UK National Health Service (NHS) as part of their care and support.

## Acknowledgements

The work was supported by the International Alliance for Cancer Early Detection, a partnership between Cancer Research UK (C18081/A31373), Canary Center at Stanford University, the University of Cambridge, OHSU Knight Cancer Institute, University College London, and the University of Manchester. SI is additionally supported by Cancer Research UK (EDDPMA-May22\100062) and HH and MB by CRUK International Alliance for Cancer Early Detection (ACED) Pathway Awards (EDDAPA-2022/100001 and EDDAPA-2022/100002, respectively). GL was supported by a Cancer Research UK (C18081/A18180) Advanced Clinician Scientist Fellowship. CR acknowledges funding from Cancer Research UK Early Detection and Diagnosis Committee (grant number EDDCPJT\100018). JUS is supported by a National Institute of Health Research Advanced Fellowship (NIHR300861). ACA is support by Cancer Research UK grant: PPRPGM-Nov20\100002. SI, AW and ACA are supported by the National Institute for Health and Care Research (NIHR) Cambridge Biomedical Research Centre (NIHR203312) [*]. AW is part of the BigData@Heart Consortium, funded by the Innovative Medicines Initiative-2 Joint Undertaking under grant agreement No 116074. AW and SI are supported by the British Heart Foundation (RG/18/13/33946: RG/F/23/110103) and by Health Data Research UK, which is funded by the UK Medical Research Council, Engineering and Physical Sciences Research Council, Economic and Social Research Council, Department of Health and Social Care (England), Chief Scientist Office of the Scottish Government Health and Social Care Directorates, Health and Social Care Research and Development Division (Welsh Government), Public Health Agency (Northern Ireland), British Heart Foundation and Wellcome.

*The views expressed are those of the authors and not necessarily those of the NIHR or the Department of Health and Social Care.

The funders had no role in study design, data collection and analysis, decision to publish, or preparation of the manuscript. All authors had access to statistical reports, tables, and analysis code. MB, CR, BW, SI and GL had full access to all of the data.

## Transparency declaration

The lead author affirms that this manuscript is an honest, accurate, and transparent account of the study being reported; that no important aspects of the study have been omitted; and that any discrepancies from the study as planned have been explained.

## Contributors

MB designed the statistical analysis, wrote analytical code, cleaned and analysed the data, and drafted and revised the paper. He is the guarantor. CR, HH and GL contributed to drafting the paper. CR, JU-S, NP and GL provided clinical interpretation. HH, AT, BW, SI and SD contributed to data management and phenotyping. JL, AW and ACA contributed to the design and interpretation of the analysis. All authors provided revisions to the paper and gave final approval to the submitted manuscript.

