## Supplementary material for "Cancer incidence and competing mortality risk following 15 presenting symptoms in primary care: a population-based cohort study using electronic healthcare records"

### Appendix 1. Supplementary methods.

Appendix 1 Figure 1A. Cohort flowchart for the reference group.

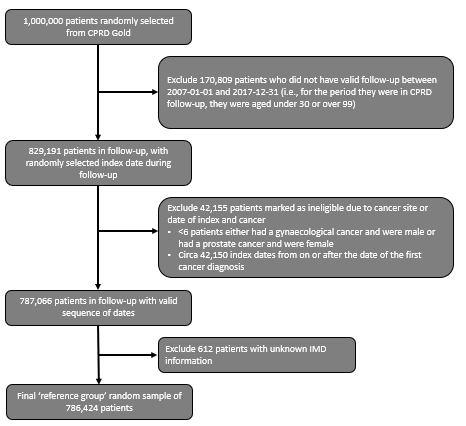

Appendix 1 Figure 1B. Cohort flowchart for the symptomatic cohort and the final analysis cohort.

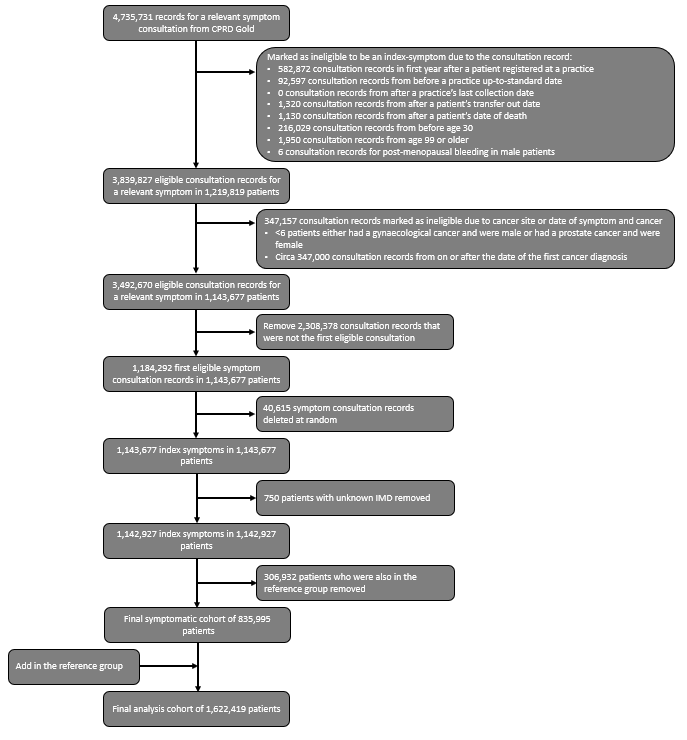

Appendix 1 Table 2. Cancer site codelist.

| Analysis grouping | Cancer site - specific | ICD10 code |
| --- | --- | --- |
| Breast | Breast | C50 |
| Breast | Breast (in-situ) | D05 |
| Gynaecological | Cervix | C53 |
| Gynaecological | Cervix (in-situ) | D06 |
| Gynaecological | Ovary | C56 |
| Gynaecological | Ovary | C57 |
| Gynaecological | Uterus | C54 |
| Gynaecological | Uterus | C55 |
| Gynaecological | Vulva | C51 |
| Lung | Lung | C33 |
| Lung | Lung | C34 |
| Lung | Mesothelioma | C45 |
| Upper GI | Liver | C22 |
| Upper GI | Oesophagus | C15 |
| Upper GI | Pancreas | C25 |
| Upper GI | Stomach | C16 |
| Lower GI | Colon | C18 |
| Lower GI | Colon | C19 |
| Lower GI | Rectum | C20 |
| Urological | Bladder | C67 |
| Urological | Bladder (in-situ) | D09 |
| Urological | Kidney | C64 |
| Urological | Other and unspecified urinary | C65 |
| Urological | Other and unspecified urinary | C66 |
| Urological | Other and unspecified urinary | C68 |
| Prostate | Prostate | C61 |
| Haematological | Acute myeloid leukaemia | C920 |
| Haematological | Acute myeloid leukaemia | C924 |
| Haematological | Acute myeloid leukaemia | C925 |
| Haematological | Acute myeloid leukaemia | C930 |
| Haematological | Acute myeloid leukaemia | C940 |
| Haematological | Acute myeloid leukaemia | C942 |
| Haematological | Chronic lymphocytic leukaemia | C911 |
| Haematological | Hodgkin lymphoma | C81 |
| Haematological | Multiple myeloma | C90 |
| Haematological | Non-Hodgkin lymphoma | C82 |
| Haematological | Non-Hodgkin lymphoma | C83 |
| Haematological | Non-Hodgkin lymphoma | C84 |
| Haematological | Non-Hodgkin lymphoma | C85 |
| Haematological | Other haematological | C88 |
| Haematological | Other haematological | C912 |
| Haematological | Other haematological | C913 |
| Haematological | Other haematological | C914 |
| Haematological | Other haematological | C915 |
| Haematological | Other haematological | C917 |
| Haematological | Other haematological | C919 |
| Haematological | Other haematological | C922 |
| Haematological | Other haematological | C923 |
| Haematological | Other haematological | C927 |
| Haematological | Other haematological | C929 |
| Haematological | Other haematological | C931 |
| Haematological | Other haematological | C932 |
| Haematological | Other haematological | C937 |
| Haematological | Other haematological | C939 |
| Haematological | Other haematological | C943 |
| Haematological | Other haematological | C944 |
| Haematological | Other haematological | C945 |
| Haematological | Other haematological | C947 |
| Haematological | Other haematological | C95 |
| Haematological | Other haematological | C96 |
| Haematological | Other leukaemia | C910 |
| Haematological | Other leukaemia | C921 |
| Other cancer | Bone sarcoma | C40 |
| Other cancer | Bone sarcoma | C41 |
| Other cancer | Brain | C71 |
| Other cancer | Brain | D330 |
| Other cancer | Brain | D331 |
| Other cancer | Brain | D332 |
| Other cancer | Brain | D430 |
| Other cancer | Brain | D431 |
| Other cancer | Brain | D432 |
| Other cancer | Connective and soft tissue sarcoma | C48 |
| Other cancer | Connective and soft tissue sarcoma | C49 |
| Other cancer | Larynx | C32 |
| Other cancer | Melanoma | C43 |
| Other cancer | Meninges | C70 |
| Other cancer | Meninges | D32 |
| Other cancer | Meninges | D42 |
| Other cancer | Non-specific head and neck | C00 |
| Other cancer | Non-specific head and neck | C14 |
| Other cancer | Non-specific head and neck | C31 |
| Other cancer | Oral cavity | C02 |
| Other cancer | Oral cavity | C03 |
| Other cancer | Oral cavity | C04 |
| Other cancer | Oral cavity | C06 |
| Other cancer | Oropharynx | C01 |
| Other cancer | Oropharynx | C09 |
| Other cancer | Oropharynx | C10 |
| Other cancer | Other CNS and intracranial | C720 |
| Other cancer | Other CNS and intracranial | C721 |
| Other cancer | Other CNS and intracranial | C722 |
| Other cancer | Other CNS and intracranial | C723 |
| Other cancer | Other CNS and intracranial | C724 |
| Other cancer | Other CNS and intracranial | C725 |
| Other cancer | Other CNS and intracranial | C751 |
| Other cancer | Other CNS and intracranial | C752 |
| Other cancer | Other CNS and intracranial | C753 |
| Other cancer | Other CNS and intracranial | D333 |
| Other cancer | Other CNS and intracranial | D334 |
| Other cancer | Other CNS and intracranial | D35 |
| Other cancer | Other CNS and intracranial | D433 |
| Other cancer | Other CNS and intracranial | D434 |
| Other cancer | Other CNS and intracranial | D44 |
| Other cancer | Other head and neck | C05 |
| Other cancer | Other head and neck | C07 |
| Other cancer | Other head and neck | C08 |
| Other cancer | Other head and neck | C11 |
| Other cancer | Other head and neck | C12 |
| Other cancer | Other head and neck | C13 |
| Other cancer | Other malignant neoplasms | C17 |
| Other cancer | Other malignant neoplasms | C21 |
| Other cancer | Other malignant neoplasms | C23 |
| Other cancer | Other malignant neoplasms | C24 |
| Other cancer | Other malignant neoplasms | C26 |
| Other cancer | Other malignant neoplasms | C30 |
| Other cancer | Other malignant neoplasms | C37 |
| Other cancer | Other malignant neoplasms | C38 |
| Other cancer | Other malignant neoplasms | C39 |
| Other cancer | Other malignant neoplasms | C46 |
| Other cancer | Other malignant neoplasms | C47 |
| Other cancer | Other malignant neoplasms | C52 |
| Other cancer | Other malignant neoplasms | C58 |
| Other cancer | Other malignant neoplasms | C60 |
| Other cancer | Other malignant neoplasms | C63 |
| Other cancer | Other malignant neoplasms | C69 |
| Other cancer | Other malignant neoplasms | C728 |
| Other cancer | Other malignant neoplasms | C729 |
| Other cancer | Other malignant neoplasms | C74 |
| Other cancer | Other malignant neoplasms | C750 |
| Other cancer | Other malignant neoplasms | C754 |
| Other cancer | Other malignant neoplasms | C755 |
| Other cancer | Other malignant neoplasms | C758 |
| Other cancer | Other malignant neoplasms | C759 |
| Other cancer | Other malignant neoplasms | C76 |
| Other cancer | Other malignant neoplasms | C97 |
| Other cancer | Testis | C62 |
| Other cancer | Thyroid | C73 |
| Other cancer | Unknown primary | C77 |
| Other cancer | Unknown primary | C78 |
| Other cancer | Unknown primary | C79 |
| Other cancer | Unknown primary | C80 |

### Appendix 2. Additional non-parametric and descriptive results.

Appendix 2 Table 1. Most common specific cancer sites in the 12 months following index by index symptom.

| **Index symptom** | **Incident cancers (first only)** | **Female never-smokers** | **Female ever-smokers** | **Male never-smokers** | **Male ever-smokers** |
| --- | --- | --- | --- | --- | --- |
| Reference cohort | No cancer | 164528 | 223702 | 145642 | 245016 |
| Reference cohort | Lung | 50 | 423 | 28 | 550 |
| Reference cohort | Breast | 359 | 540 |  | 7 |
| Reference cohort | Prostate |  |  | 216 | 613 |
| Reference cohort | Colon | 112 | 172 | 71 | 237 |
| Reference cohort | Cervix (in-situ) | 100 | 211 |  |  |
| Reference cohort | Unknown primary | 55 | 104 | 25 | 94 |
| Reference cohort | Non-Hodgkin lymphoma | 49 | 64 | 36 | 114 |
| Reference cohort | Pancreas | 51 | 89 | 18 | 91 |
| Reference cohort | Melanoma | 43 | 57 | 48 | 90 |
| Reference cohort | Rectum | 43 | 62 | 26 | 94 |
| Reference cohort | Bladder | 26 | 38 | 23 | 119 |
| Reference cohort | Oesophagus | 16 | 53 | 26 | 108 |
| Reference cohort | Stomach | 31 | 36 | 25 | 108 |
| Reference cohort | Ovary | 79 | 99 |  |  |
| Reference cohort | Kidney | 29 | 37 | 25 | 72 |
| Reference cohort | Other malignant neoplasms | 28 | 55 | 18 | 56 |
| Reference cohort | Brain | 25 | 39 | 27 | 55 |
| Reference cohort | Uterus | 65 | 74 |  |  |
| Reference cohort | Multiple myeloma | 14 | 27 | 19 | 55 |
| Reference cohort | Liver | 21 | 23 |  | 63 |
| Reference cohort | Chronic lymphocytic leukaemia | 12 | 19 | 10 | 41 |
| Reference cohort | Bladder (in-situ) |  | 15 | 13 | 52 |
| Reference cohort | Breast (in-situ) | 32 | 43 |  |  |
| Reference cohort | Mesothelioma | 6 | 5 | 14 | 42 |
| Reference cohort | Acute myeloid leukaemia | 12 | 15 | 8 | 31 |
| Reference cohort | Oral cavity | 8 | 15 | 5 | 27 |
| Reference cohort | Thyroid | 15 | 21 | 9 | 8 |
| Reference cohort | Cervix | 16 | 33 |  |  |
| Reference cohort | Connective and soft tissue sarcoma | 13 | 13 |  | 19 |
| Reference cohort | Meninges | 19 | 17 |  | 9 |
| Reference cohort | Oropharynx |  | 14 |  | 25 |
| Reference cohort | Other haematological | 5 | 6 | 8 | 19 |
| Reference cohort | Larynx |  | 6 |  | 32 |
| Reference cohort | Other CNS and intracranial | 13 | 7 | 5 | 9 |
| Reference cohort | Testis |  |  | 19 | 14 |
| Reference cohort | Other head and neck |  | 12 | 6 | 12 |
| Reference cohort | Vulva | 9 | 20 |  |  |
| Reference cohort | Hodgkin lymphoma | 8 | 5 |  | 15 |
| Reference cohort | Other leukaemia |  | 8 | 7 | 7 |
| Reference cohort | Other and unspecified urinary |  | 6 |  | 14 |
| Reference cohort | Non-specific head and neck |  |  |  | 5 |
| Reference cohort | Aggregated (sites with counts <5) | 21 | 3 | 19 | 4 |
| Abdominal pain | No cancer | 53508 | 86276 | 27390 | 61154 |
| Abdominal pain | Colon | 225 | 351 | 140 | 457 |
| Abdominal pain | Prostate |  |  | 135 | 327 |
| Abdominal pain | Pancreas | 72 | 148 | 49 | 169 |
| Abdominal pain | Ovary | 148 | 223 |  |  |
| Abdominal pain | Lung | 22 | 132 | 12 | 156 |
| Abdominal pain | Breast | 110 | 194 |  |  |
| Abdominal pain | Non-Hodgkin lymphoma | 60 | 74 | 33 | 100 |
| Abdominal pain | Unknown primary | 57 | 97 | 15 | 73 |
| Abdominal pain | Other malignant neoplasms | 40 | 70 | 14 | 77 |
| Abdominal pain | Stomach | 26 | 41 | 27 | 105 |
| Abdominal pain | Kidney | 26 | 56 | 26 | 90 |
| Abdominal pain | Cervix (in-situ) | 41 | 139 |  |  |
| Abdominal pain | Oesophagus | 19 | 39 | 24 | 90 |
| Abdominal pain | Rectum | 20 | 32 | 21 | 67 |
| Abdominal pain | Liver | 23 | 35 | 17 | 53 |
| Abdominal pain | Uterus | 55 | 51 |  |  |
| Abdominal pain | Bladder | 12 | 24 | 12 | 58 |
| Abdominal pain | Melanoma | 27 | 25 | 8 | 19 |
| Abdominal pain | Connective and soft tissue sarcoma | 15 | 27 | 6 | 14 |
| Abdominal pain | Multiple myeloma | 10 | 19 | 9 | 24 |
| Abdominal pain | Mesothelioma | 6 | 9 | 9 | 19 |
| Abdominal pain | Cervix | 15 | 24 |  |  |
| Abdominal pain | Breast (in-situ) | 7 | 25 |  |  |
| Abdominal pain | Bladder (in-situ) |  |  |  | 32 |
| Abdominal pain | Chronic lymphocytic leukaemia |  | 6 | 5 | 16 |
| Abdominal pain | Brain |  | 8 | 5 | 10 |
| Abdominal pain | Thyroid | 8 | 11 |  |  |
| Abdominal pain | Acute myeloid leukaemia |  |  | 8 | 11 |
| Abdominal pain | Testis |  |  | 9 | 7 |
| Abdominal pain | Other leukaemia |  |  | 5 | 6 |
| Abdominal pain | Larynx |  |  |  | 9 |
| Abdominal pain | Other and unspecified urinary |  |  |  | 8 |
| Abdominal pain | Other haematological |  |  |  | 8 |
| Abdominal pain | Oral cavity |  |  |  | 7 |
| Abdominal pain | Meninges |  | 7 |  |  |
| Abdominal pain | Other CNS and intracranial |  |  |  | 6 |
| Abdominal pain | Other head and neck |  |  |  | 5 |
| Abdominal pain | Aggregated (sites with counts <5) | 30 | 24 | 14 | 14 |
| Abdominal bloating | No cancer | 5495 | 9048 | 2245 | 5213 |
| Abdominal bloating | Ovary | 55 | 64 |  |  |
| Abdominal bloating | Colon | 14 | 25 | 12 | 32 |
| Abdominal bloating | Prostate |  |  | 17 | 28 |
| Abdominal bloating | Pancreas | 8 | 19 |  | 16 |
| Abdominal bloating | Rectum | 8 | 8 |  | 22 |
| Abdominal bloating | Breast | 11 | 24 |  |  |
| Abdominal bloating | Lung |  | 9 |  | 14 |
| Abdominal bloating | Cervix (in-situ) |  | 20 |  |  |
| Abdominal bloating | Connective and soft tissue sarcoma | 5 | 8 |  |  |
| Abdominal bloating | Other malignant neoplasms | 5 |  |  | 8 |
| Abdominal bloating | Oesophagus |  |  |  | 12 |
| Abdominal bloating | Liver |  |  |  | 11 |
| Abdominal bloating | Non-Hodgkin lymphoma |  |  |  | 11 |
| Abdominal bloating | Stomach |  | 5 |  | 5 |
| Abdominal bloating | Breast (in-situ) | 7 |  |  |  |
| Abdominal bloating | Melanoma | 6 |  |  |  |
| Abdominal bloating | Kidney | 6 |  |  |  |
| Abdominal bloating | Uterus |  | 6 |  |  |
| Abdominal bloating | Unknown primary | 5 |  |  |  |
| Abdominal bloating | Aggregated (sites with counts <5) | 24 | 36 | 32 | 30 |
| Rectal bleeding | No cancer | 8108 | 12371 | 8402 | 17766 |
| Rectal bleeding | Rectum | 81 | 137 | 101 | 317 |
| Rectal bleeding | Colon | 99 | 161 | 92 | 233 |
| Rectal bleeding | Prostate |  |  | 38 | 92 |
| Rectal bleeding | Other malignant neoplasms | 14 | 37 | 6 | 18 |
| Rectal bleeding | Breast | 16 | 44 |  |  |
| Rectal bleeding | Lung |  | 13 |  | 27 |
| Rectal bleeding | Stomach | 8 | 7 |  | 19 |
| Rectal bleeding | Non-Hodgkin lymphoma | 6 | 6 | 8 | 13 |
| Rectal bleeding | Melanoma |  | 14 |  | 11 |
| Rectal bleeding | Unknown primary | 6 | 10 |  | 7 |
| Rectal bleeding | Uterus | 8 | 11 |  |  |
| Rectal bleeding | Cervix (in-situ) | 5 | 12 |  |  |
| Rectal bleeding | Kidney |  |  |  | 15 |
| Rectal bleeding | Ovary | 7 | 6 |  |  |
| Rectal bleeding | Bladder |  | 6 |  | 7 |
| Rectal bleeding | Oesophagus |  |  |  | 11 |
| Rectal bleeding | Bladder (in-situ) |  |  |  | 8 |
| Rectal bleeding | Multiple myeloma |  |  |  | 8 |
| Rectal bleeding | Liver |  |  |  | 6 |
| Rectal bleeding | Pancreas |  |  |  | 5 |
| Rectal bleeding | Other head and neck |  |  |  | 5 |
| Rectal bleeding | Brain |  |  | 5 |  |
| Rectal bleeding | Aggregated (sites with counts <5) | 29 | 36 | 21 | 16 |
| Change in bowel habit | No cancer | 3188 | 5357 | 2331 | 5269 |
| Change in bowel habit | Colon | 53 | 76 | 51 | 161 |
| Change in bowel habit | Rectum | 42 | 51 | 51 | 162 |
| Change in bowel habit | Prostate |  |  | 12 | 60 |
| Change in bowel habit | Breast | 13 | 29 |  |  |
| Change in bowel habit | Ovary | 15 | 24 |  |  |
| Change in bowel habit | Lung |  | 16 |  | 21 |
| Change in bowel habit | Pancreas | 6 | 12 |  | 8 |
| Change in bowel habit | Other malignant neoplasms | 7 | 7 | 6 | 5 |
| Change in bowel habit | Unknown primary |  | 7 |  | 13 |
| Change in bowel habit | Non-Hodgkin lymphoma |  | 5 |  | 9 |
| Change in bowel habit | Uterus | 6 | 6 |  |  |
| Change in bowel habit | Oesophagus |  |  |  | 7 |
| Change in bowel habit | Liver |  |  |  | 7 |
| Change in bowel habit | Bladder |  |  |  | 6 |
| Change in bowel habit | Kidney |  |  |  | 5 |
| Change in bowel habit | Breast (in-situ) |  | 5 |  |  |
| Change in bowel habit | Stomach |  |  |  | 5 |
| Change in bowel habit | Aggregated (sites with counts <5) | 29 | 25 | 25 | 19 |
| Dyspepsia | No cancer | 20569 | 34203 | 14478 | 37118 |
| Dyspepsia | Oesophagus | 30 | 55 | 38 | 163 |
| Dyspepsia | Stomach | 20 | 44 | 34 | 114 |
| Dyspepsia | Prostate |  |  | 40 | 162 |
| Dyspepsia | Breast | 70 | 120 |  |  |
| Dyspepsia | Colon | 34 | 50 | 24 | 70 |
| Dyspepsia | Lung | 7 | 55 | 5 | 103 |
| Dyspepsia | Pancreas | 32 | 33 | 8 | 48 |
| Dyspepsia | Non-Hodgkin lymphoma | 13 | 25 | 12 | 35 |
| Dyspepsia | Unknown primary | 14 | 27 | 7 | 30 |
| Dyspepsia | Other malignant neoplasms | 11 | 20 | 8 | 20 |
| Dyspepsia | Kidney | 9 | 5 | 9 | 28 |
| Dyspepsia | Ovary | 19 | 27 |  |  |
| Dyspepsia | Uterus | 16 | 22 |  |  |
| Dyspepsia | Melanoma | 13 | 13 |  | 12 |
| Dyspepsia | Cervix (in-situ) | 9 | 26 |  |  |
| Dyspepsia | Liver | 5 | 5 |  | 23 |
| Dyspepsia | Bladder |  |  | 5 | 27 |
| Dyspepsia | Multiple myeloma | 7 | 7 | 6 | 10 |
| Dyspepsia | Rectum |  | 11 |  | 17 |
| Dyspepsia | Brain | 6 | 6 |  | 9 |
| Dyspepsia | Breast (in-situ) | 7 | 13 |  |  |
| Dyspepsia | Chronic lymphocytic leukaemia |  | 8 |  | 9 |
| Dyspepsia | Oropharynx |  |  | 5 | 9 |
| Dyspepsia | Other CNS and intracranial |  |  |  | 8 |
| Dyspepsia | Bladder (in-situ) |  |  |  | 8 |
| Dyspepsia | Acute myeloid leukaemia |  |  |  | 7 |
| Dyspepsia | Larynx |  |  |  | 7 |
| Dyspepsia | Other leukaemia |  | 6 |  |  |
| Dyspepsia | Meninges |  | 6 |  |  |
| Dyspepsia | Thyroid |  | 6 |  |  |
| Dyspepsia | Connective and soft tissue sarcoma |  |  |  | 5 |
| Dyspepsia | Mesothelioma |  |  |  | 5 |
| Dyspepsia | Aggregated (sites with counts <5) | 12 | 27 | 19 | 20 |
| Dysphagia | No cancer | 3112 | 4085 | 2142 | 4617 |
| Dysphagia | Oesophagus | 67 | 134 | 59 | 298 |
| Dysphagia | Stomach | 7 | 16 | 20 | 72 |
| Dysphagia | Lung |  | 35 |  | 34 |
| Dysphagia | Prostate |  |  | 6 | 25 |
| Dysphagia | Colon | 12 |  | 5 | 9 |
| Dysphagia | Breast | 8 | 14 |  |  |
| Dysphagia | Non-Hodgkin lymphoma |  | 5 |  | 14 |
| Dysphagia | Unknown primary |  | 5 |  | 9 |
| Dysphagia | Larynx |  |  |  | 11 |
| Dysphagia | Other head and neck |  |  |  | 11 |
| Dysphagia | Oropharynx |  |  |  | 10 |
| Dysphagia | Thyroid |  | 5 |  |  |
| Dysphagia | Ovary |  | 5 |  |  |
| Dysphagia | Non-specific head and neck |  |  |  | 5 |
| Dysphagia | Aggregated (sites with counts <5) | 33 | 49 | 23 | 30 |
| Jaundice | No cancer | 261 | 352 | 203 | 545 |
| Jaundice | Pancreas | 43 | 81 | 41 | 72 |
| Jaundice | Liver | 12 | 25 | 10 | 26 |
| Jaundice | Other malignant neoplasms | 7 | 20 | 10 | 33 |
| Jaundice | Unknown primary | 7 | 8 | 5 | 7 |
| Jaundice | Lung |  |  |  | 5 |
| Jaundice | Aggregated (sites with counts <5) | 6 | 13 | 7 | 18 |
| Dyspnoea | No cancer | 19397 | 51179 | 11162 | 55411 |
| Dyspnoea | Lung | 36 | 483 | 20 | 670 |
| Dyspnoea | Colon | 42 | 104 | 40 | 164 |
| Dyspnoea | Prostate |  |  | 64 | 283 |
| Dyspnoea | Breast | 73 | 216 |  | 5 |
| Dyspnoea | Mesothelioma | 7 | 15 | 27 | 115 |
| Dyspnoea | Non-Hodgkin lymphoma | 21 | 39 | 27 | 75 |
| Dyspnoea | Unknown primary | 16 | 53 | 10 | 82 |
| Dyspnoea | Bladder |  | 27 | 5 | 68 |
| Dyspnoea | Kidney | 11 | 29 | 10 | 39 |
| Dyspnoea | Oesophagus | 11 | 23 | 5 | 48 |
| Dyspnoea | Stomach | 9 | 11 | 11 | 54 |
| Dyspnoea | Pancreas | 10 | 26 |  | 35 |
| Dyspnoea | Ovary | 18 | 47 |  |  |
| Dyspnoea | Other malignant neoplasms | 7 | 22 | 6 | 28 |
| Dyspnoea | Acute myeloid leukaemia | 6 | 22 | 6 | 26 |
| Dyspnoea | Multiple myeloma | 10 | 14 | 10 | 25 |
| Dyspnoea | Melanoma | 7 | 17 | 6 | 20 |
| Dyspnoea | Liver | 5 | 6 | 5 | 33 |
| Dyspnoea | Rectum | 5 | 12 |  | 31 |
| Dyspnoea | Uterus | 12 | 28 |  |  |
| Dyspnoea | Larynx |  | 8 |  | 30 |
| Dyspnoea | Bladder (in-situ) |  | 11 |  | 25 |
| Dyspnoea | Cervix (in-situ) | 8 | 27 |  |  |
| Dyspnoea | Chronic lymphocytic leukaemia |  | 12 | 5 | 11 |
| Dyspnoea | Breast (in-situ) | 8 | 14 |  |  |
| Dyspnoea | Oropharynx |  | 7 |  | 13 |
| Dyspnoea | Brain | 5 |  |  | 14 |
| Dyspnoea | Oral cavity |  | 7 |  | 11 |
| Dyspnoea | Connective and soft tissue sarcoma |  | 8 |  | 8 |
| Dyspnoea | Other haematological |  | 8 |  | 7 |
| Dyspnoea | Thyroid | 5 | 6 |  |  |
| Dyspnoea | Cervix |  | 9 |  |  |
| Dyspnoea | Other leukaemia |  |  |  | 7 |
| Dyspnoea | Meninges |  | 6 |  |  |
| Dyspnoea | Non-specific head and neck |  |  |  | 5 |
| Dyspnoea | Hodgkin lymphoma |  |  |  | 5 |
| Dyspnoea | Other head and neck |  |  |  | 5 |
| Dyspnoea | Aggregated (sites with counts <5) | 31 | 18 | 33 | 15 |
| Haemoptysis | No cancer | 616 | 1362 | 780 | 2689 |
| Haemoptysis | Lung | 8 | 88 | 10 | 204 |
| Haemoptysis | Prostate |  |  |  | 10 |
| Haemoptysis | Breast |  | 9 |  |  |
| Haemoptysis | Larynx |  |  |  | 6 |
| Haemoptysis | Colon |  |  |  | 5 |
| Haemoptysis | Stomach |  |  |  | 5 |
| Haemoptysis | Aggregated (sites with counts <5) | 10 | 17 | 7 | 33 |
| Haematuria | No cancer | 3095 | 5324 | 4097 | 10467 |
| Haematuria | Bladder | 64 | 162 | 139 | 708 |
| Haematuria | Prostate |  |  | 119 | 351 |
| Haematuria | Bladder (in-situ) | 22 | 63 | 75 | 288 |
| Haematuria | Kidney | 18 | 30 | 56 | 161 |
| Haematuria | Other and unspecified urinary | 14 | 17 | 20 | 50 |
| Haematuria | Uterus | 30 | 37 |  |  |
| Haematuria | Lung |  | 19 |  | 42 |
| Haematuria | Colon | 6 | 5 | 6 | 27 |
| Haematuria | Breast | 18 | 16 |  |  |
| Haematuria | Unknown primary |  | 7 |  | 15 |
| Haematuria | Non-Hodgkin lymphoma |  | 5 |  | 15 |
| Haematuria | Other malignant neoplasms |  |  |  | 10 |
| Haematuria | Ovary |  | 9 |  |  |
| Haematuria | Stomach |  |  |  | 6 |
| Haematuria | Cervix |  | 6 |  |  |
| Haematuria | Melanoma |  |  |  | 6 |
| Haematuria | Multiple myeloma |  |  |  | 6 |
| Haematuria | Brain |  |  |  | 5 |
| Haematuria | Rectum |  |  |  | 5 |
| Haematuria | Aggregated (sites with counts <5) | 28 | 27 | 29 | 28 |
| Fatigue | No cancer | 37144 | 55527 | 14691 | 32165 |
| Fatigue | Lung | 21 | 144 | 12 | 170 |
| Fatigue | Prostate |  |  | 63 | 191 |
| Fatigue | Breast | 101 | 149 |  |  |
| Fatigue | Colon | 60 | 79 | 28 | 83 |
| Fatigue | Non-Hodgkin lymphoma | 24 | 40 | 17 | 45 |
| Fatigue | Cervix (in-situ) | 28 | 74 |  |  |
| Fatigue | Brain | 20 | 25 | 12 | 25 |
| Fatigue | Unknown primary | 21 | 25 | 7 | 26 |
| Fatigue | Pancreas | 16 | 21 | 7 | 24 |
| Fatigue | Melanoma | 19 | 18 | 6 | 20 |
| Fatigue | Kidney | 6 | 21 | 9 | 26 |
| Fatigue | Other malignant neoplasms | 9 | 20 | 6 | 16 |
| Fatigue | Ovary | 28 | 22 |  |  |
| Fatigue | Rectum | 7 | 15 | 7 | 19 |
| Fatigue | Liver | 8 | 12 | 8 | 19 |
| Fatigue | Acute myeloid leukaemia | 7 | 17 | 6 | 16 |
| Fatigue | Uterus | 19 | 26 |  |  |
| Fatigue | Multiple myeloma | 10 | 10 | 5 | 17 |
| Fatigue | Bladder | 5 | 10 |  | 25 |
| Fatigue | Oesophagus |  | 8 |  | 27 |
| Fatigue | Stomach | 5 | 10 |  | 18 |
| Fatigue | Chronic lymphocytic leukaemia | 6 | 6 |  | 16 |
| Fatigue | Breast (in-situ) | 12 | 15 |  |  |
| Fatigue | Meninges | 7 | 13 |  |  |
| Fatigue | Mesothelioma |  |  | 5 | 15 |
| Fatigue | Hodgkin lymphoma | 7 | 6 |  | 6 |
| Fatigue | Connective and soft tissue sarcoma |  | 7 | 5 | 6 |
| Fatigue | Thyroid |  | 12 |  |  |
| Fatigue | Other haematological |  |  |  | 11 |
| Fatigue | Bladder (in-situ) |  |  |  | 7 |
| Fatigue | Cervix |  | 6 |  |  |
| Fatigue | Oral cavity |  | 6 |  |  |
| Fatigue | Other CNS and intracranial |  | 6 |  |  |
| Fatigue | Oropharynx |  |  |  | 5 |
| Fatigue | Aggregated (sites with counts <5) | 28 | 20 | 28 | 24 |
| Night sweats | No cancer | 1454 | 3101 | 839 | 2148 |
| Night sweats | Lung |  | 7 |  | 16 |
| Night sweats | Prostate |  |  |  | 14 |
| Night sweats | Breast | 6 | 5 |  |  |
| Night sweats | Hodgkin lymphoma |  |  |  | 7 |
| Night sweats | Colon |  |  |  | 6 |
| Night sweats | Kidney |  |  |  | 5 |
| Night sweats | Aggregated (sites with counts <5) | 9 | 23 | 14 | 21 |
| Weight loss | No cancer | 3477 | 6506 | 2036 | 6360 |
| Weight loss | Lung | 6 | 90 | 11 | 151 |
| Weight loss | Colon | 27 | 25 | 15 | 58 |
| Weight loss | Prostate |  |  | 31 | 93 |
| Weight loss | Pancreas | 12 | 26 | 17 | 37 |
| Weight loss | Unknown primary | 13 | 14 | 9 | 41 |
| Weight loss | Stomach | 10 | 23 | 5 | 37 |
| Weight loss | Oesophagus | 6 | 14 | 6 | 30 |
| Weight loss | Non-Hodgkin lymphoma | 6 | 9 | 14 | 24 |
| Weight loss | Breast | 17 | 25 |  |  |
| Weight loss | Kidney | 5 |  |  | 28 |
| Weight loss | Rectum | 8 |  | 5 | 17 |
| Weight loss | Other malignant neoplasms | 5 | 12 |  | 11 |
| Weight loss | Liver | 5 |  | 5 | 13 |
| Weight loss | Multiple myeloma |  | 6 | 6 | 8 |
| Weight loss | Ovary | 9 | 11 |  |  |
| Weight loss | Bladder |  | 7 |  | 8 |
| Weight loss | Mesothelioma |  |  |  | 8 |
| Weight loss | Brain |  |  |  | 8 |
| Weight loss | Chronic lymphocytic leukaemia |  |  |  | 7 |
| Weight loss | Hodgkin lymphoma |  |  |  | 6 |
| Weight loss | Cervix (in-situ) |  | 6 |  |  |
| Weight loss | Bladder (in-situ) |  |  |  | 6 |
| Weight loss | Oral cavity |  |  |  | 5 |
| Weight loss | Other haematological |  |  |  | 5 |
| Weight loss | Larynx |  |  |  | 5 |
| Weight loss | Aggregated (sites with counts <5) | 35 | 48 | 16 | 12 |
| Breast lump | No cancer | 12312 | 19045 | 615 | 1546 |
| Breast lump | Breast | 1741 | 2666 | 8 | 20 |
| Breast lump | Breast (in-situ) | 68 | 108 |  |  |
| Breast lump | Cervix (in-situ) | 12 | 26 |  |  |
| Breast lump | Non-Hodgkin lymphoma | 8 | 20 |  |  |
| Breast lump | Melanoma | 6 | 7 |  |  |
| Breast lump | Lung |  | 11 |  |  |
| Breast lump | Ovary |  | 5 |  |  |
| Breast lump | Aggregated (sites with counts <5) | 23 | 38 | 7 | 15 |
| Post-menopausal bleed | No cancer | 2932 | 4446 |  |  |
| Post-menopausal bleed | Uterus | 231 | 356 |  |  |
| Post-menopausal bleed | Ovary | 27 | 29 |  |  |
| Post-menopausal bleed | Cervix | 14 | 31 |  |  |
| Post-menopausal bleed | Breast | 5 | 19 |  |  |
| Post-menopausal bleed | Bladder |  | 11 |  |  |
| Post-menopausal bleed | Cervix (in-situ) |  | 5 |  |  |
| Post-menopausal bleed | Colon | 5 |  |  |  |
| Post-menopausal bleed | Aggregated (sites with counts <5) | 26 | 35 |  |  |

Appendix 2 Figure 1. Aalen-Johansen estimates of cumulative incidence up to 12 months, male non-smokers

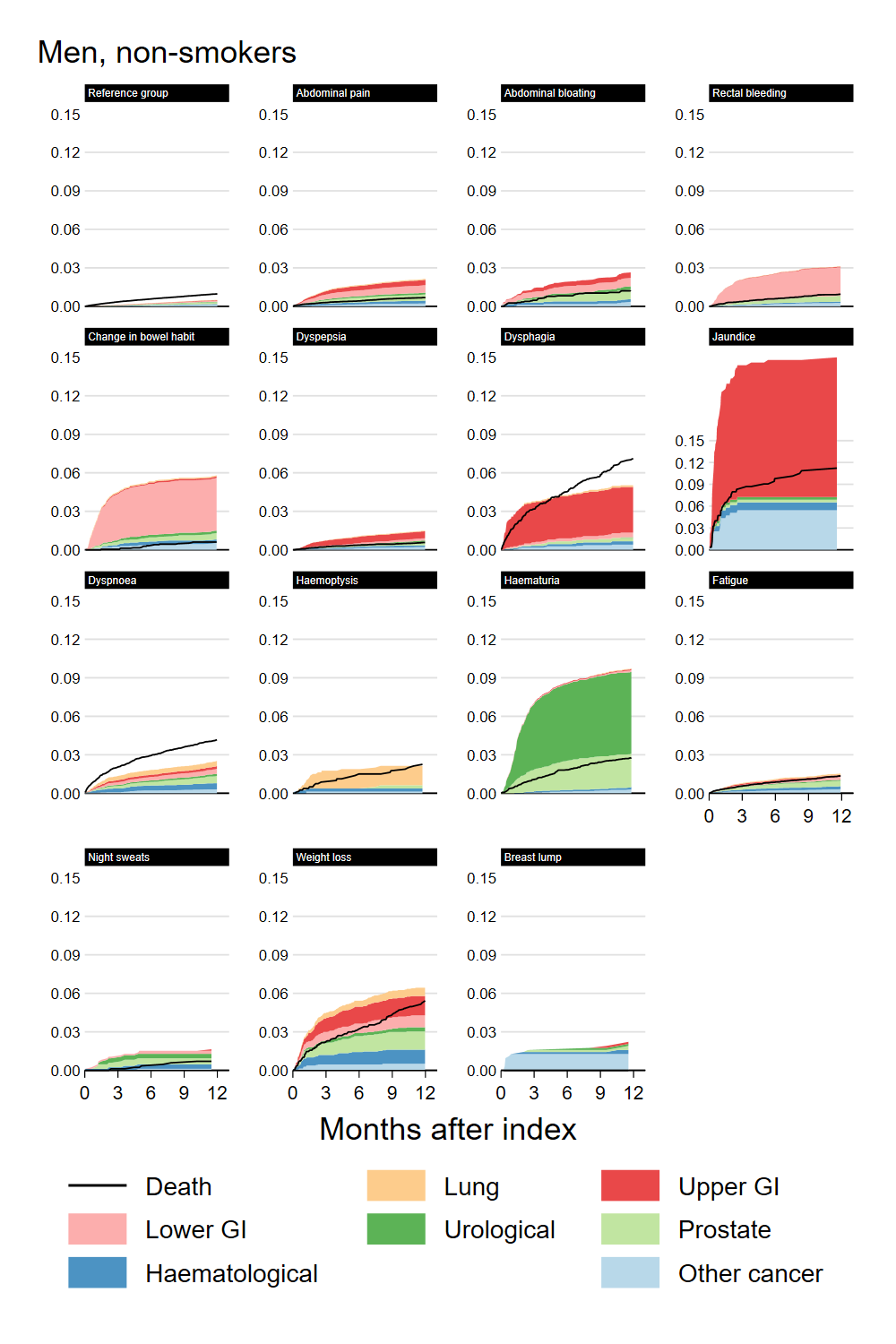

Appendix 2 Figure 2. Aalen-Johansen estimates of cumulative incidence up to 12 months, male smokers

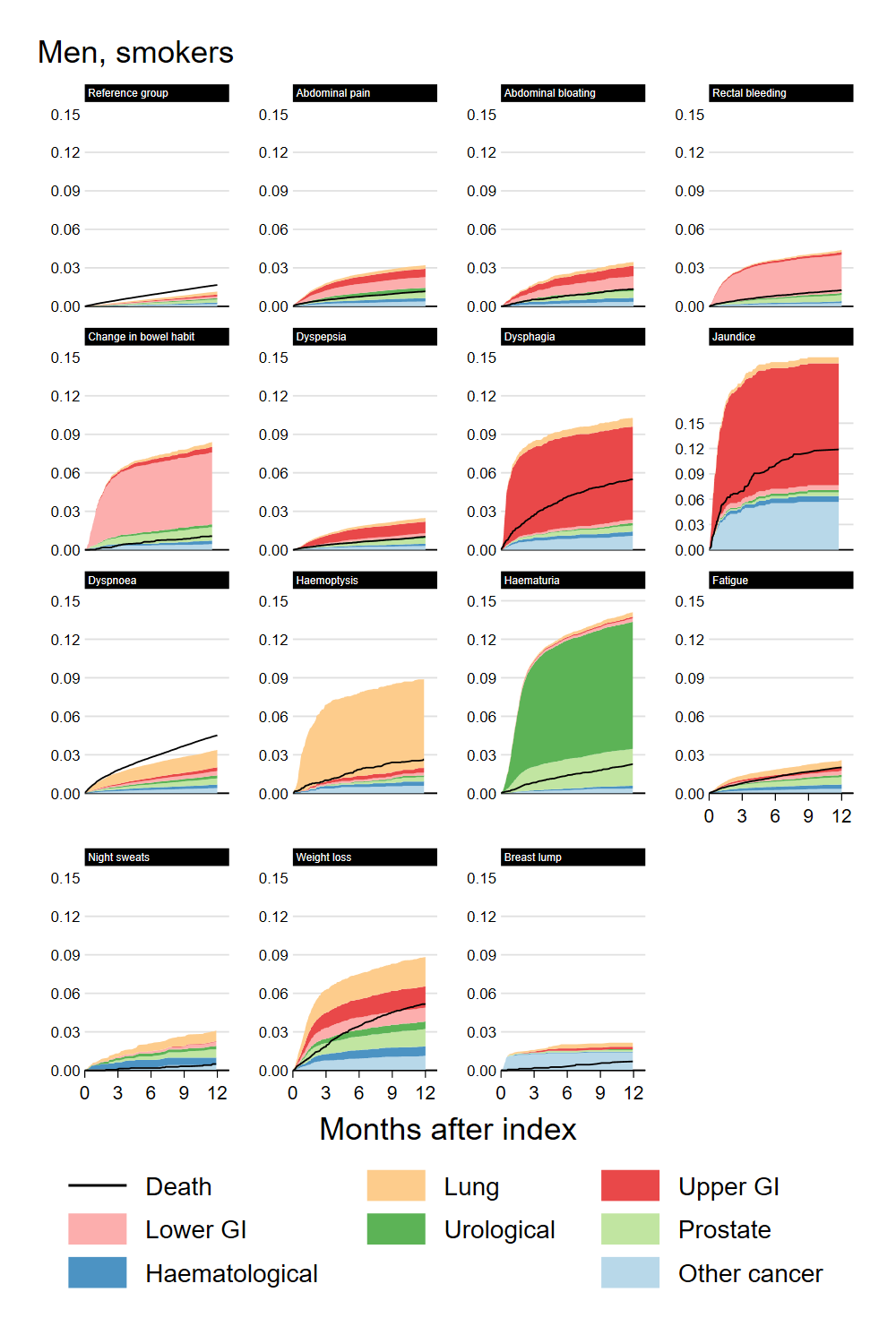

Appendix 2 Figure 3. Aalen-Johansen estimates of cumulative incidence up to 12 months, female non-smokers

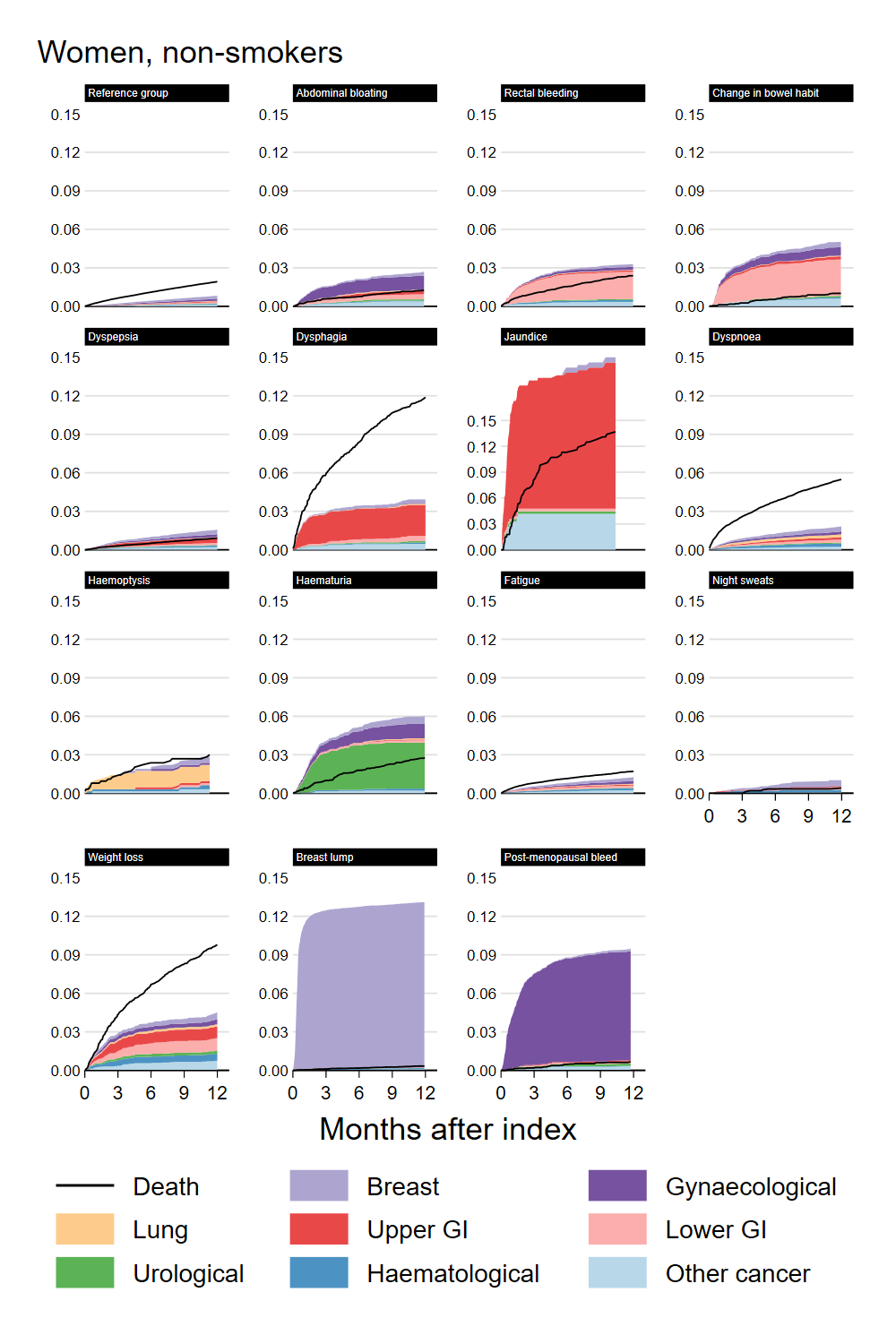

Appendix 2 Figure 4. Aalen-Johansen estimates of cumulative incidence up to 12 months, female smokers

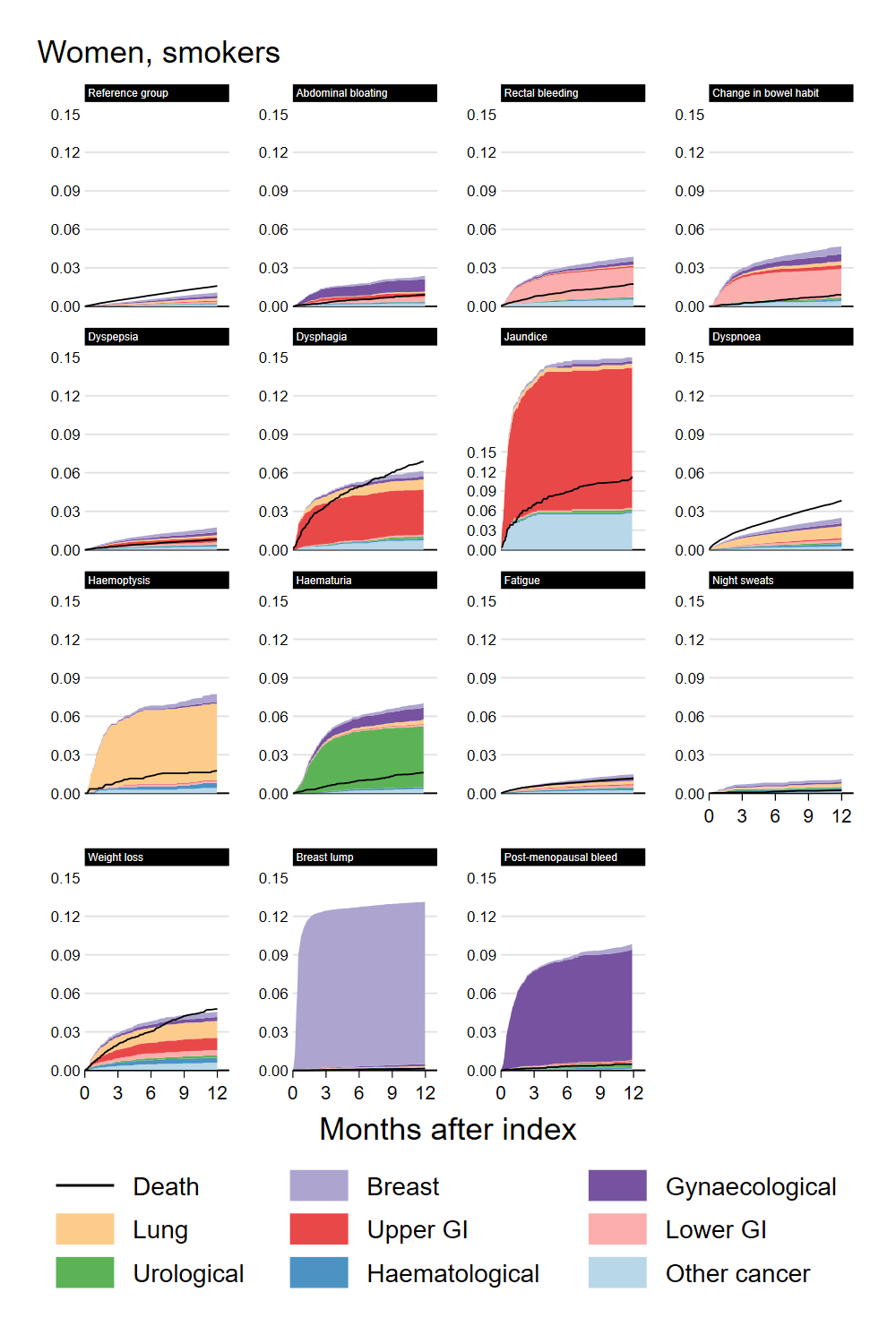

### Appendix 3. Figures showing simulated outcomes at 3 and 6 months after index symptom by age, sex and smoking status.

Appendix 3 Figure 1. Cumulative incidence at 3 months, male non-smokers

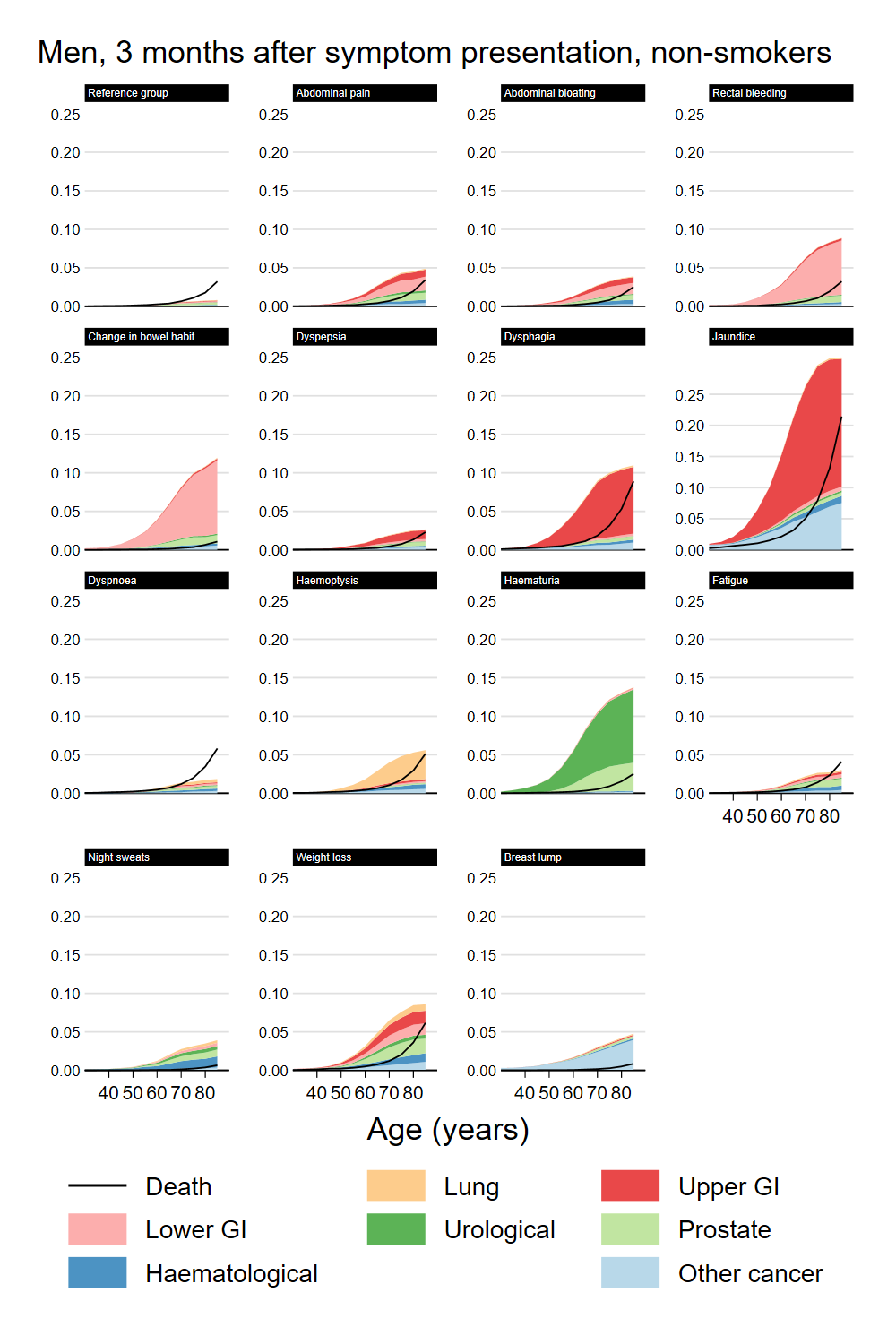

Appendix 3 Figure 2. Cumulative incidence at 3 months, male smokers

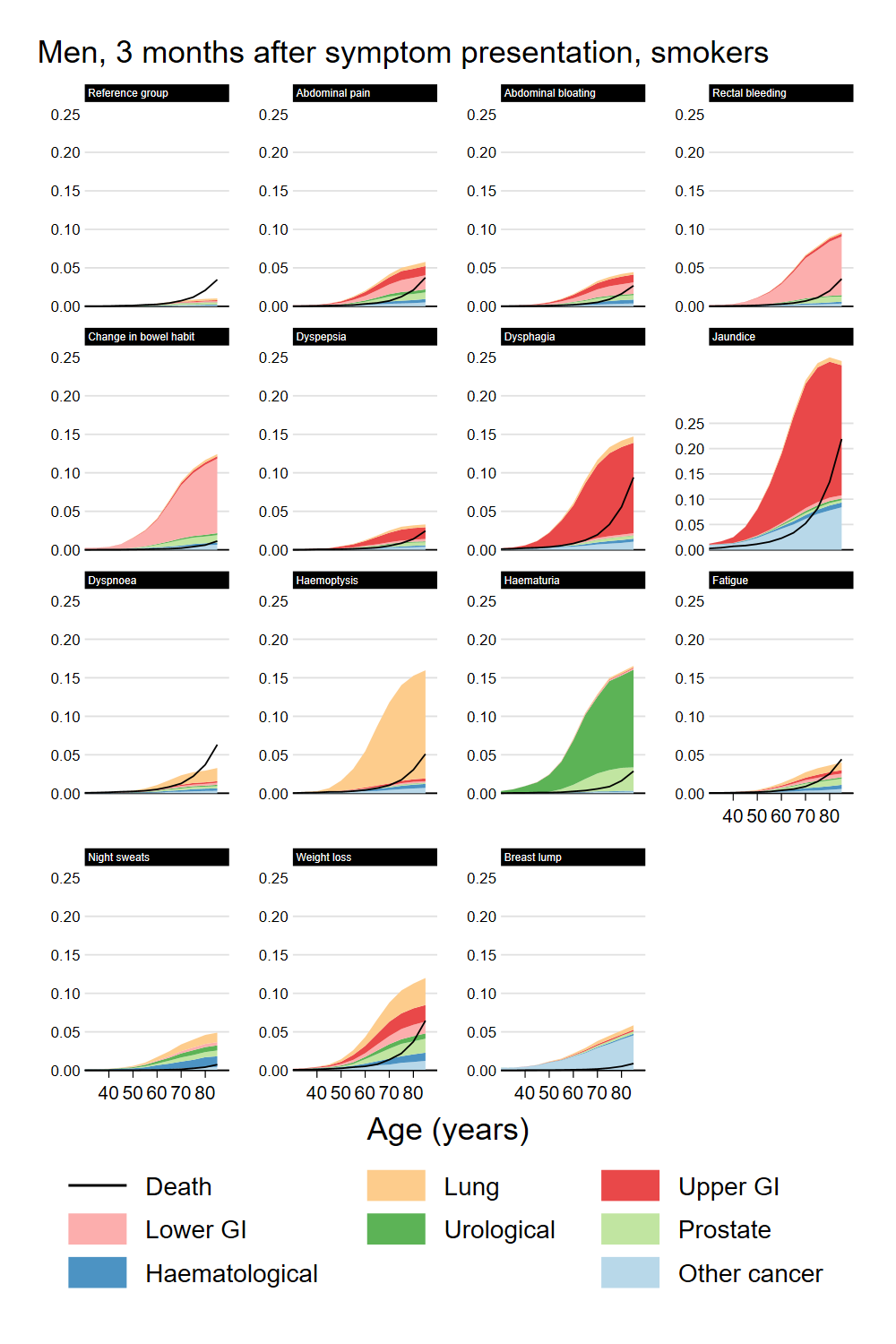

Appendix 3 Figure 3. Cumulative incidence at 3 months, female non-smokers

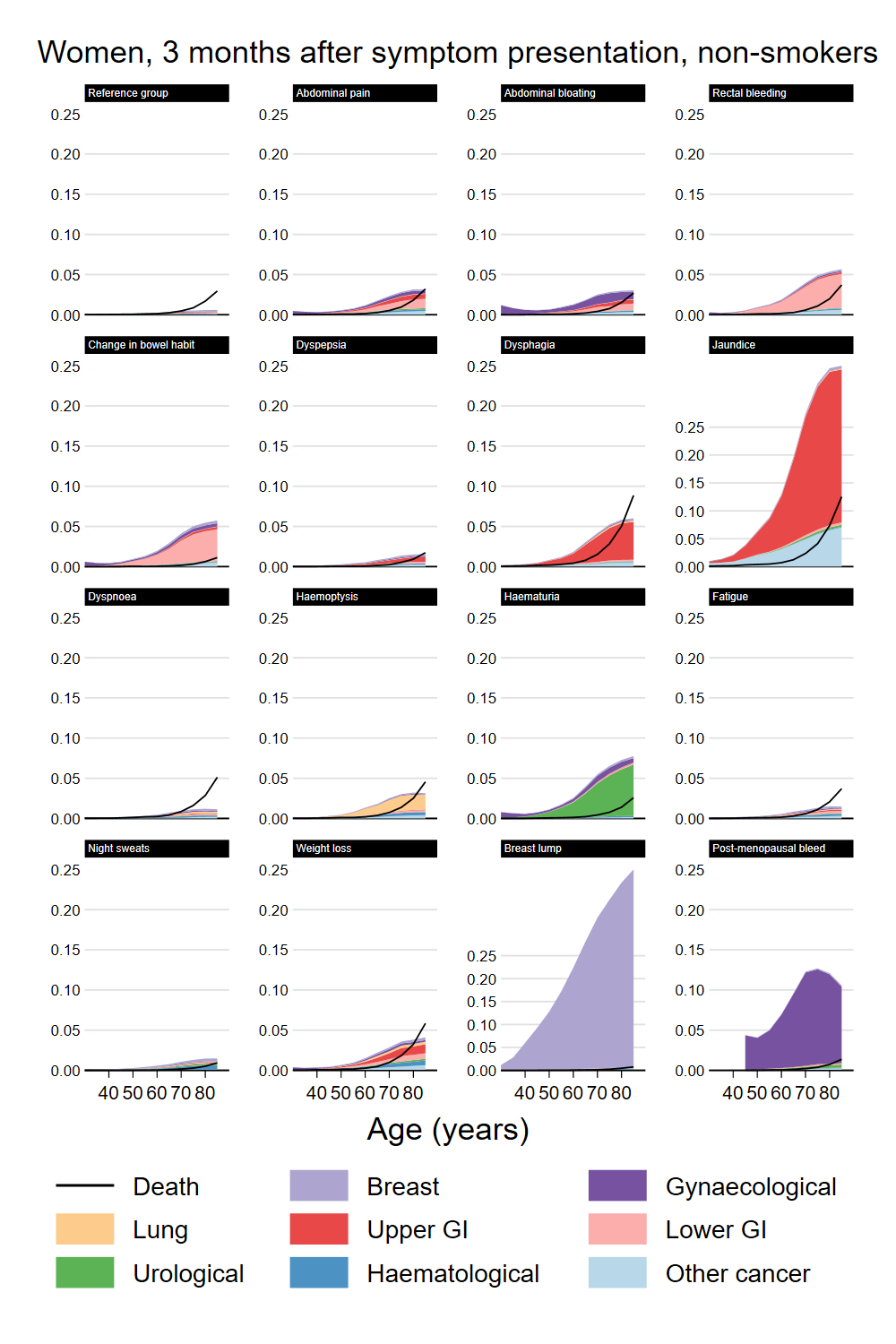

Appendix 3 Figure 4. Cumulative incidence at 3 months, female smokers

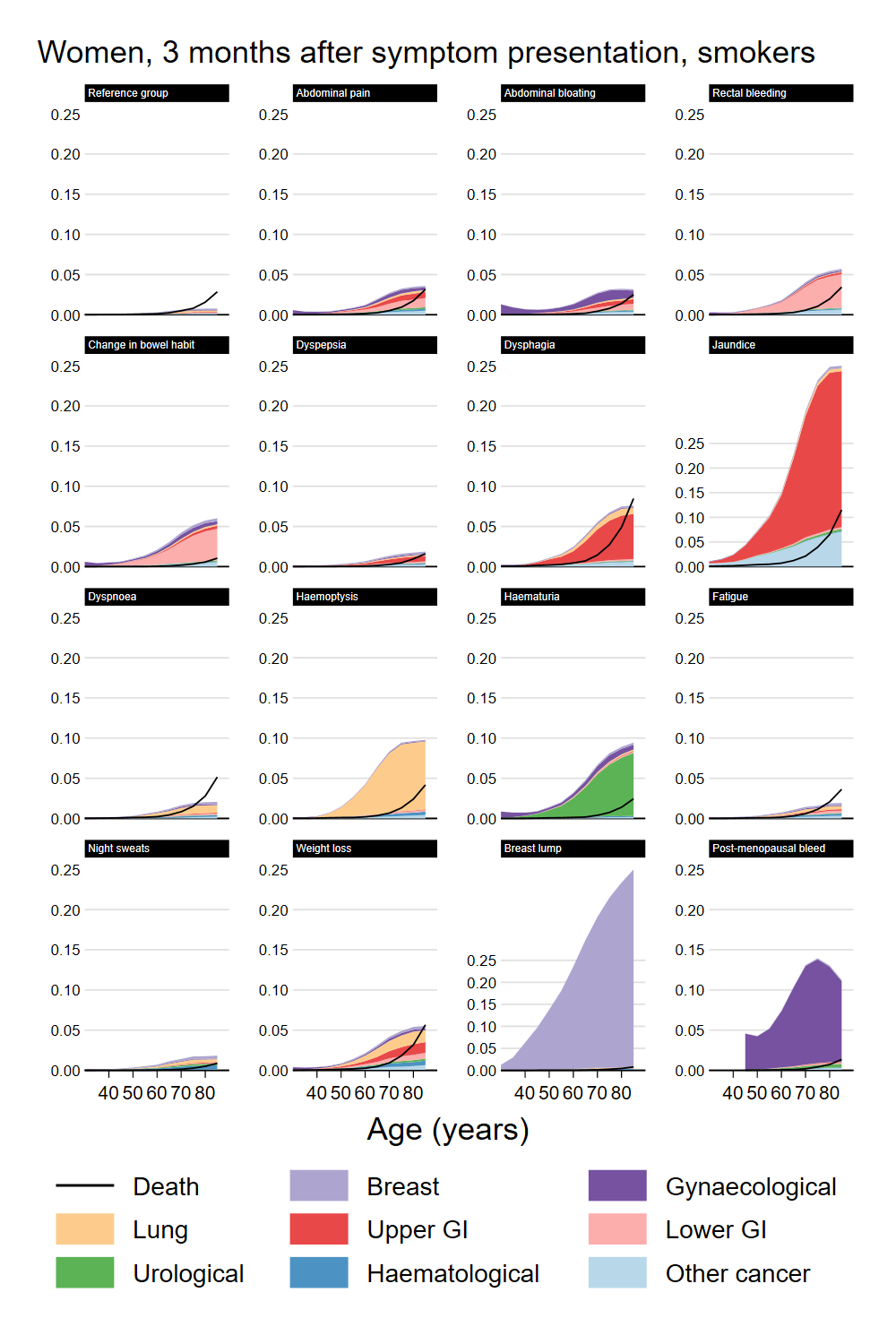

Appendix 3 Figure 5. Cumulative incidence at 6 months, male non-smokers

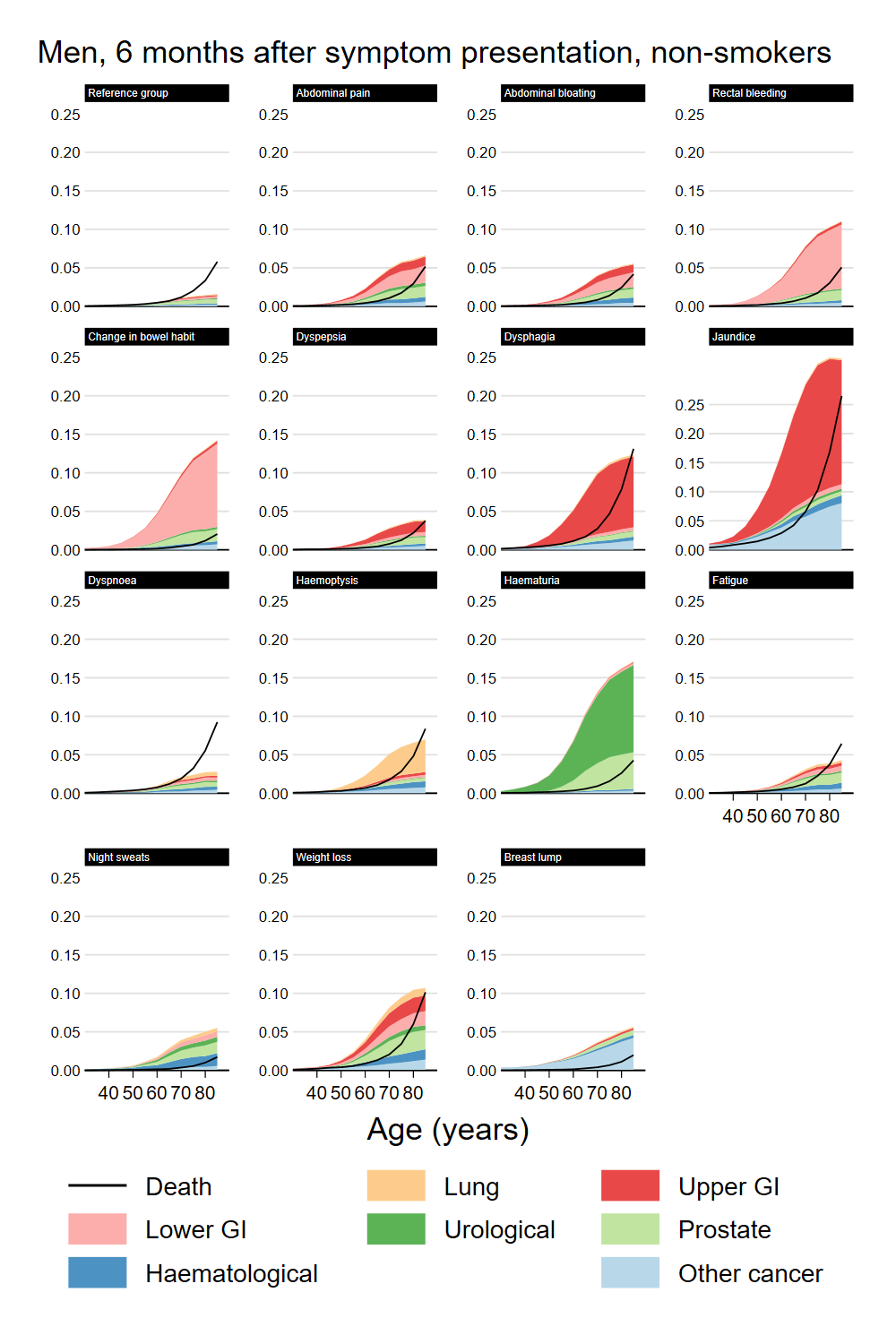

Appendix 3 Figure 6. Cumulative incidence at 6 months, male smokers

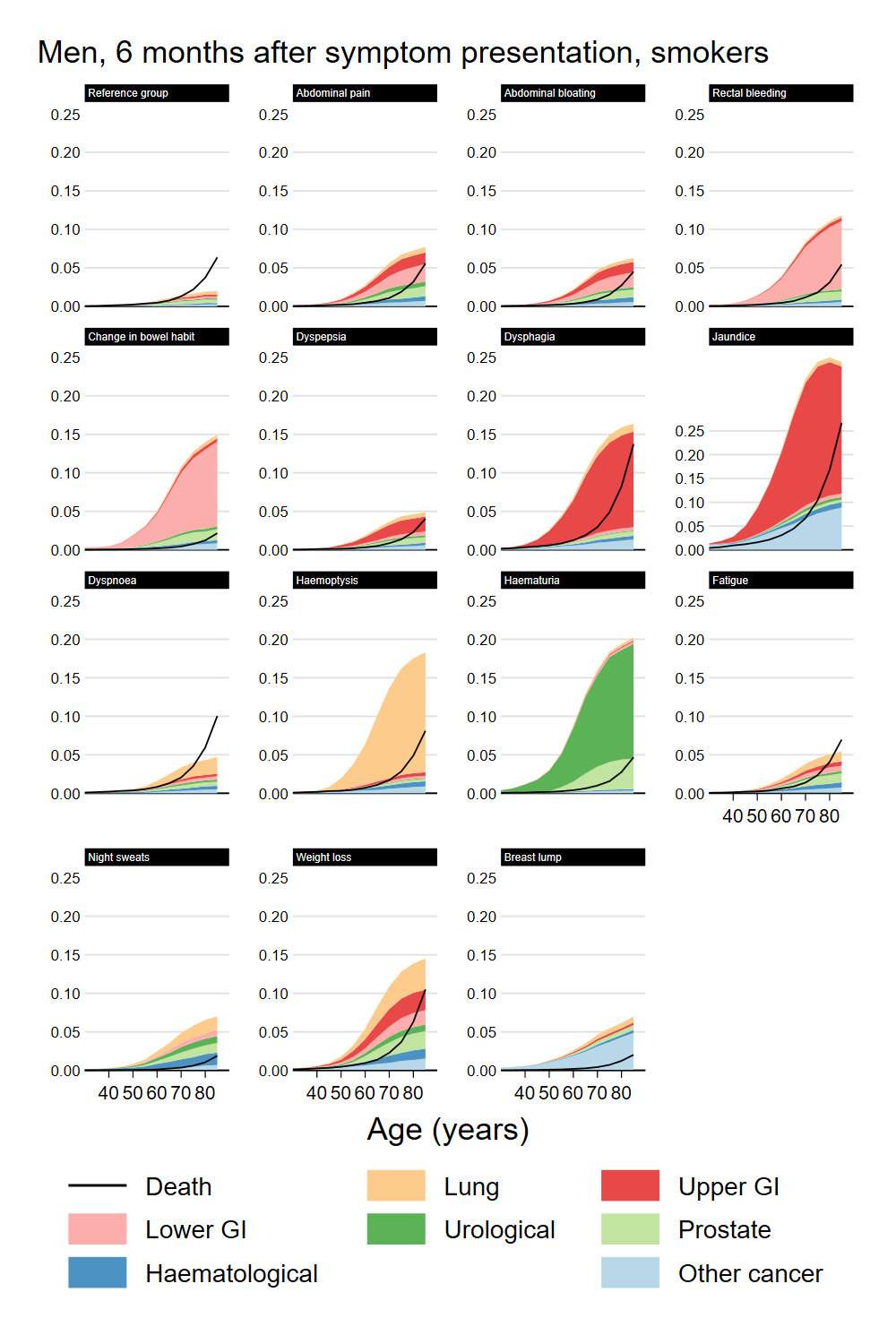

Appendix 3 Figure 7. Cumulative incidence at 6 months, female non-smokers

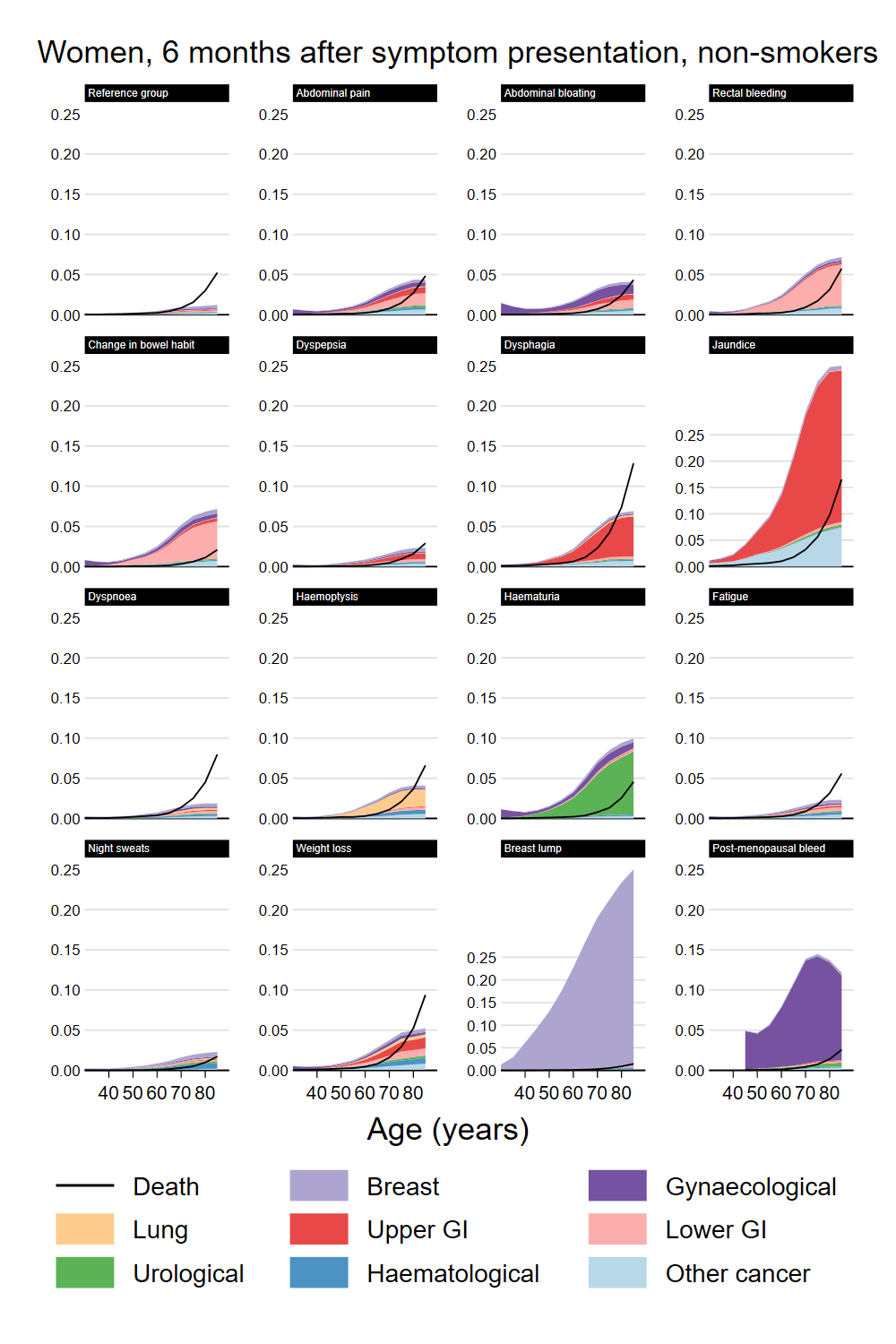

Appendix 3 Figure 8. Cumulative incidence at 6 months, female smokers

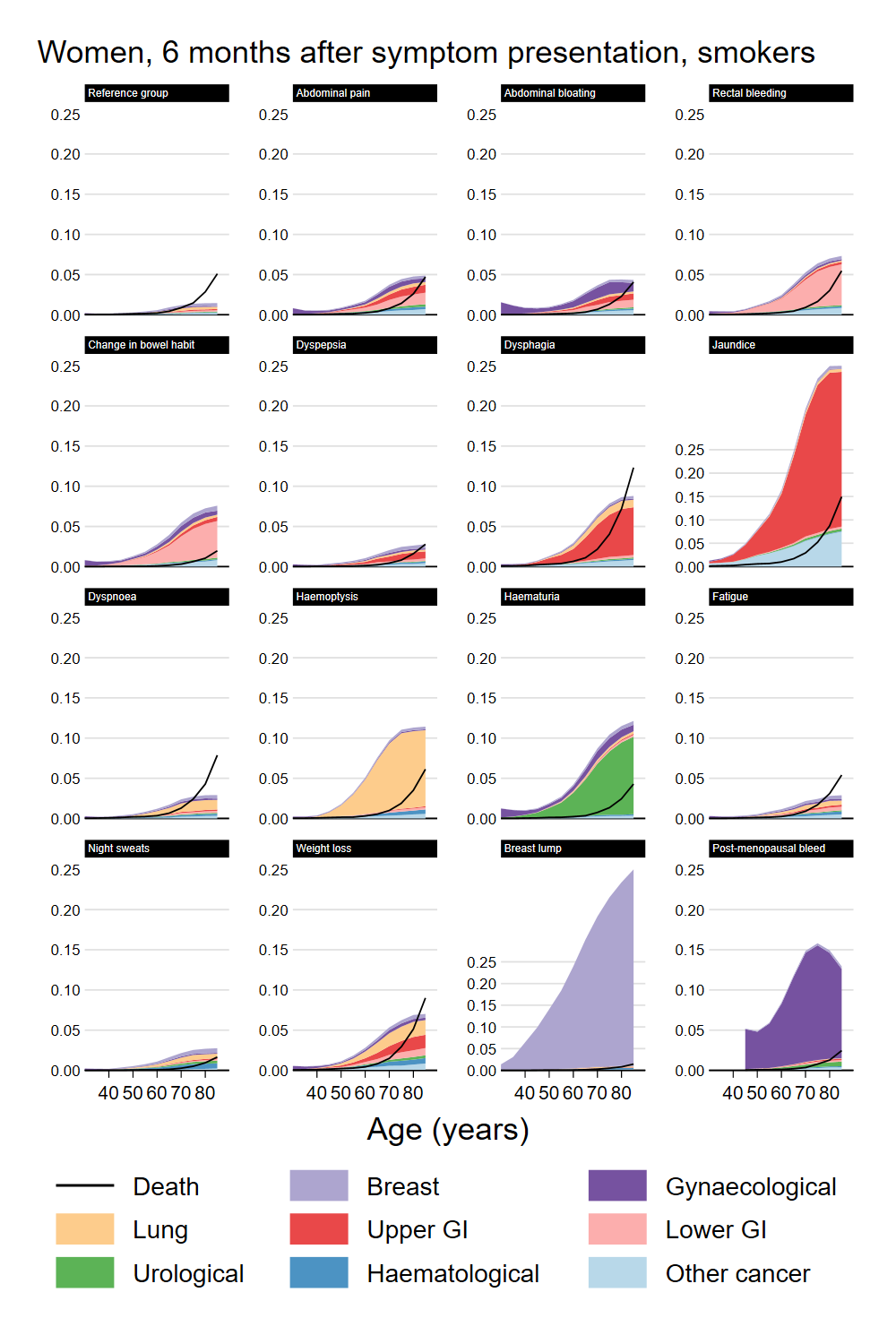

### Appendix 4. Details of the outcome-specific models.

Appendix 4 Table 1. Hazard ratios for lung cancer, men.

|  | **Hazard ratio** | |  |
| --- | --- | --- | --- |
| **Outcome: Lung** | **Estimate** | **(95% CI)** | **p-value** |
| **Smoking status** |  |  |  |
| Never smoker | (ref) |  |  |
| Ever-smoker | 3.99 | (3.45, 4.60) | <0.001 |
| **Symptoms** |  |  |  |
| Reference cohort | (ref) |  |  |
| Abdominal pain | 0.99 | (0.86, 1.14) | 0.860 |
| ln(time) * Abdominal pain | 0.81 | (0.78, 0.85) | <0.001 |
| Abdominal bloating | 0.73 | (0.47, 1.13) | 0.159 |
| ln(time) * Abdominal bloating | 0.84 | (0.74, 0.95) | 0.007 |
| Rectal bleeding | 0.67 | (0.50, 0.90) | 0.008 |
| ln(time) * Rectal bleeding | 0.95 | (0.85, 1.07) | 0.415 |
| Change in bowel habit | 0.88 | (0.61, 1.27) | 0.492 |
| ln(time) * Change in bowel habit | 0.91 | (0.80, 1.03) | 0.118 |
| Dyspepsia | 0.95 | (0.80, 1.13) | 0.585 |
| ln(time) * Dyspepsia | 0.88 | (0.83, 0.93) | <0.001 |
| Dysphagia | 1.52 | (1.12, 2.05) | 0.007 |
| ln(time) * Dysphagia | 0.79 | (0.73, 0.84) | <0.001 |
| Jaundice | 1.92 | (0.85, 4.31) | 0.116 |
| ln(time) * Jaundice | 0.77 | (0.65, 0.90) | 0.001 |
| Dyspnoea | 2.62 | (2.40, 2.87) | <0.001 |
| ln(time) * Dyspnoea | 0.78 | (0.75, 0.80) | <0.001 |
| Haemoptysis | 17.14 | (14.81, 19.84) | <0.001 |
| ln(time) * Haemoptysis | 0.71 | (0.69, 0.74) | <0.001 |
| Haematuria | 0.79 | (0.61, 1.02) | 0.072 |
| ln(time) * Haematuria | 0.94 | (0.86, 1.04) | 0.231 |
| Fatigue | 1.57 | (1.36, 1.82) | <0.001 |
| ln(time) * Fatigue | 0.79 | (0.76, 0.82) | <0.001 |
| Night sweats | 2.85 | (1.89, 4.31) | <0.001 |
| ln(time) * Night sweats | 0.86 | (0.76, 0.98) | 0.018 |
| Weight loss | 4.70 | (4.01, 5.50) | <0.001 |
| ln(time) * Weight loss | 0.74 | (0.71, 0.77) | <0.001 |
| Breast lump | 0.93 | (0.39, 2.25) | 0.876 |
| ln(time) * Breast lump | 0.79 | (0.66, 0.95) | 0.013 |

Appendix 4 Table 2. Hazard ratios for upper GI cancer, men.

|  | **Hazard ratio** | |  |
| --- | --- | --- | --- |
| **Outcome: Upper GI** | **Estimate** | **(95% CI)** | **p-value** |
| **Smoking status** |  |  |  |
| Never smoker | (ref) |  |  |
| Ever-smoker | 1.35 | (1.23, 1.48) | <0.001 |
| **Symptoms** |  |  |  |
| Reference cohort | (ref) |  |  |
| Abdominal pain | 3.68 | (3.29, 4.12) | <0.001 |
| ln(time) * Abdominal pain | 0.75 | (0.72, 0.78) | <0.001 |
| Abdominal bloating | 3.57 | (2.75, 4.64) | <0.001 |
| ln(time) * Abdominal bloating | 0.78 | (0.73, 0.84) | <0.001 |
| Rectal bleeding | 1.26 | (0.96, 1.65) | 0.094 |
| ln(time) * Rectal bleeding | 0.77 | (0.72, 0.81) | <0.001 |
| Change in bowel habit | 1.63 | (1.15, 2.30) | 0.006 |
| ln(time) * Change in bowel habit | 0.85 | (0.76, 0.94) | 0.002 |
| Dyspepsia | 4.50 | (3.99, 5.08) | <0.001 |
| ln(time) * Dyspepsia | 0.75 | (0.72, 0.78) | <0.001 |
| Dysphagia | 23.78 | (21.06, 26.86) | <0.001 |
| ln(time) * Dysphagia | 0.67 | (0.64, 0.69) | <0.001 |
| Jaundice | 70.89 | (59.44, 84.56) | <0.001 |
| ln(time) * Jaundice | 0.67 | (0.64, 0.69) | <0.001 |
| Dyspnoea | 1.19 | (1.03, 1.38) | 0.020 |
| ln(time) * Dyspnoea | 0.88 | (0.84, 0.93) | <0.001 |
| Haemoptysis | 1.42 | (0.78, 2.58) | 0.247 |
| ln(time) * Haemoptysis | 0.81 | (0.69, 0.94) | 0.006 |
| Haematuria | 0.69 | (0.48, 0.98) | 0.038 |
| ln(time) * Haematuria | 0.92 | (0.81, 1.05) | 0.202 |
| Fatigue | 1.38 | (1.14, 1.67) | <0.001 |
| ln(time) * Fatigue | 0.78 | (0.74, 0.82) | <0.001 |
| Night sweats | 0.84 | (0.34, 2.05) | 0.700 |
| ln(time) * Night sweats | 1.97 | (0.76, 5.10) | 0.162 |
| Weight loss | 6.67 | (5.60, 7.93) | <0.001 |
| ln(time) * Weight loss | 0.74 | (0.71, 0.78) | <0.001 |
| Breast lump | 1.12 | (0.42, 3.00) | 0.821 |
| ln(time) * Breast lump | 0.88 | (0.64, 1.22) | 0.445 |

Appendix 4 Table 3. Hazard ratios for lower GI cancer, men.

|  | **Hazard ratio** | |  |
| --- | --- | --- | --- |
| **Outcome: Lower GI** | **Estimate** | **(95% CI)** | **p-value** |
| **Smoking status** |  |  |  |
| Never smoker | (ref) |  |  |
| Ever-smoker | 1.04 | (0.96, 1.12) | 0.359 |
| **Symptoms** |  |  |  |
| Reference cohort | (ref) |  |  |
| Abdominal pain | 5.02 | (4.51, 5.59) | <0.001 |
| ln(time) * Abdominal pain | 0.76 | (0.73, 0.78) | <0.001 |
| Abdominal bloating | 4.77 | (3.78, 6.02) | <0.001 |
| ln(time) * Abdominal bloating | 0.79 | (0.75, 0.84) | <0.001 |
| Rectal bleeding | 17.45 | (15.69, 19.40) | <0.001 |
| ln(time) * Rectal bleeding | 0.73 | (0.70, 0.75) | <0.001 |
| Change in bowel habit | 21.47 | (18.98, 24.28) | <0.001 |
| ln(time) * Change in bowel habit | 0.71 | (0.69, 0.74) | <0.001 |
| Dyspepsia | 1.60 | (1.34, 1.91) | <0.001 |
| ln(time) * Dyspepsia | 0.91 | (0.85, 0.97) | 0.002 |
| Dysphagia | 1.62 | (1.11, 2.35) | 0.012 |
| ln(time) * Dysphagia | 0.89 | (0.79, 1.00) | 0.045 |
| Jaundice | 2.52 | (1.04, 6.13) | 0.041 |
| ln(time) * Jaundice | 0.74 | (0.64, 0.85) | <0.001 |
| Dyspnoea | 1.52 | (1.32, 1.74) | <0.001 |
| ln(time) * Dyspnoea | 0.95 | (0.90, 1.00) | 0.032 |
| Haemoptysis | 1.09 | (0.54, 2.19) | 0.812 |
| ln(time) * Haemoptysis | 0.90 | (0.72, 1.13) | 0.357 |
| Haematuria | 1.13 | (0.85, 1.50) | 0.408 |
| ln(time) * Haematuria | 0.93 | (0.85, 1.03) | 0.186 |
| Fatigue | 1.86 | (1.57, 2.20) | <0.001 |
| ln(time) * Fatigue | 0.84 | (0.80, 0.88) | <0.001 |
| Night sweats | 2.18 | (1.20, 3.95) | 0.011 |
| ln(time) * Night sweats | 0.90 | (0.74, 1.10) | 0.304 |
| Weight loss | 4.33 | (3.52, 5.34) | <0.001 |
| ln(time) * Weight loss | 0.75 | (0.72, 0.79) | <0.001 |
| Breast lump | 0.26 | (0.03, 1.89) | 0.182 |
| ln(time) * Breast lump | 2.09 | (0.24, 18.57) | 0.508 |

Appendix 4 Table 4. Hazard ratios for urological cancer, men.

|  | **Hazard ratio** | |  |
| --- | --- | --- | --- |
| **Outcome: Urological** | **Estimate** | **(95% CI)** | **p-value** |
| **Smoking status** |  |  |  |
| Never smoker | (ref) |  |  |
| Ever-smoker | 1.36 | (1.24, 1.49) | <0.001 |
| **Symptoms** |  |  |  |
| Reference cohort | (ref) |  |  |
| Abdominal pain | 2.39 | (2.06, 2.78) | <0.001 |
| ln(time) * Abdominal pain | 0.81 | (0.77, 0.85) | <0.001 |
| Abdominal bloating | 1.58 | (1.01, 2.47) | 0.046 |
| ln(time) * Abdominal bloating | 0.90 | (0.76, 1.06) | 0.195 |
| Rectal bleeding | 1.29 | (0.95, 1.76) | 0.108 |
| ln(time) * Rectal bleeding | 0.88 | (0.79, 0.98) | 0.019 |
| Change in bowel habit | 1.55 | (1.02, 2.35) | 0.041 |
| ln(time) * Change in bowel habit | 0.80 | (0.71, 0.90) | <0.001 |
| Dyspepsia | 1.42 | (1.15, 1.75) | 0.001 |
| ln(time) * Dyspepsia | 0.96 | (0.88, 1.05) | 0.412 |
| Dysphagia | 1.22 | (0.74, 2.01) | 0.433 |
| ln(time) * Dysphagia | 1.14 | (0.86, 1.50) | 0.362 |
| Jaundice | 4.61 | (2.06, 10.33) | <0.001 |
| ln(time) * Jaundice | 0.84 | (0.67, 1.05) | 0.121 |
| Dyspnoea | 1.41 | (1.20, 1.66) | <0.001 |
| ln(time) * Dyspnoea | 0.89 | (0.84, 0.95) | <0.001 |
| Haemoptysis | 0.89 | (0.37, 2.15) | 0.792 |
| ln(time) * Haemoptysis | 1.06 | (0.69, 1.64) | 0.787 |
| Haematuria | 45.08 | (40.56, 50.10) | <0.001 |
| ln(time) * Haematuria | 0.69 | (0.66, 0.72) | <0.001 |
| Fatigue | 1.37 | (1.10, 1.71) | 0.006 |
| ln(time) * Fatigue | 0.85 | (0.79, 0.91) | <0.001 |
| Night sweats | 2.85 | (1.57, 5.19) | <0.001 |
| ln(time) * Night sweats | 0.74 | (0.66, 0.83) | <0.001 |
| Weight loss | 2.97 | (2.23, 3.97) | <0.001 |
| ln(time) * Weight loss | 0.73 | (0.68, 0.77) | <0.001 |
| Breast lump | 1.03 | (0.33, 3.25) | 0.959 |
| ln(time) * Breast lump | 1.63 | (0.59, 4.52) | 0.351 |

Appendix 4 Table 5. Hazard ratios for prostate cancer, men.

|  | **Hazard ratio** | |  |
| --- | --- | --- | --- |
| **Outcome: Prostate** | **Estimate** | **(95% CI)** | **p-value** |
| **Smoking status** |  |  |  |
| Never smoker | (ref) |  |  |
| Ever-smoker | 0.89 | (0.82, 0.95) | <0.001 |
| **Symptoms** |  |  |  |
| Reference cohort | (ref) |  |  |
| Abdominal pain | 1.92 | (1.74, 2.12) | <0.001 |
| ln(time) * Abdominal pain | 0.84 | (0.82, 0.88) | <0.001 |
| Abdominal bloating | 1.88 | (1.47, 2.40) | <0.001 |
| ln(time) * Abdominal bloating | 0.93 | (0.85, 1.03) | 0.156 |
| Rectal bleeding | 1.84 | (1.56, 2.17) | <0.001 |
| ln(time) * Rectal bleeding | 0.86 | (0.82, 0.91) | <0.001 |
| Change in bowel habit | 2.08 | (1.68, 2.58) | <0.001 |
| ln(time) * Change in bowel habit | 0.83 | (0.78, 0.89) | <0.001 |
| Dyspepsia | 1.45 | (1.27, 1.65) | <0.001 |
| ln(time) * Dyspepsia | 0.94 | (0.89, 0.99) | 0.020 |
| Dysphagia | 1.04 | (0.76, 1.43) | 0.808 |
| ln(time) * Dysphagia | 0.85 | (0.77, 0.94) | 0.001 |
| Jaundice | 1.25 | (0.52, 3.03) | 0.622 |
| ln(time) * Jaundice | 0.79 | (0.65, 0.96) | 0.018 |
| Dyspnoea | 1.11 | (1.00, 1.24) | 0.043 |
| ln(time) * Dyspnoea | 0.97 | (0.93, 1.02) | 0.232 |
| Haemoptysis | 1.17 | (0.74, 1.87) | 0.499 |
| ln(time) * Haemoptysis | 1.16 | (0.89, 1.51) | 0.274 |
| Haematuria | 5.27 | (4.76, 5.84) | <0.001 |
| ln(time) * Haematuria | 0.77 | (0.75, 0.80) | <0.001 |
| Fatigue | 1.72 | (1.52, 1.94) | <0.001 |
| ln(time) * Fatigue | 0.83 | (0.80, 0.87) | <0.001 |
| Night sweats | 2.15 | (1.41, 3.28) | <0.001 |
| ln(time) * Night sweats | 0.91 | (0.78, 1.06) | 0.229 |
| Weight loss | 2.79 | (2.34, 3.32) | <0.001 |
| ln(time) * Weight loss | 0.76 | (0.73, 0.80) | <0.001 |
| Breast lump | 1.17 | (0.58, 2.34) | 0.665 |
| ln(time) * Breast lump | 0.99 | (0.72, 1.38) | 0.965 |

Append 4 Table 6. Hazard ratios for haematological cancer, men.

|  | **Hazard ratio** | |  |
| --- | --- | --- | --- |
| **Outcome: Haematological** | **Estimate** | **(95% CI)** | **p-value** |
| **Smoking status** |  |  |  |
| Never smoker | (ref) |  |  |
| Ever-smoker | 0.98 | (0.88, 1.10) | 0.742 |
| **Symptoms** |  |  |  |
| Reference cohort | (ref) |  |  |
| Abdominal pain | 2.35 | (2.03, 2.72) | <0.001 |
| ln(time) * Abdominal pain | 0.80 | (0.77, 0.85) | <0.001 |
| Abdominal bloating | 2.08 | (1.42, 3.07) | <0.001 |
| ln(time) * Abdominal bloating | 0.75 | (0.69, 0.82) | <0.001 |
| Rectal bleeding | 1.26 | (0.93, 1.71) | 0.142 |
| ln(time) * Rectal bleeding | 0.81 | (0.75, 0.88) | <0.001 |
| Change in bowel habit | 1.69 | (1.14, 2.50) | 0.009 |
| ln(time) * Change in bowel habit | 0.83 | (0.75, 0.93) | 0.001 |
| Dyspepsia | 1.47 | (1.20, 1.81) | <0.001 |
| ln(time) * Dyspepsia | 0.89 | (0.83, 0.95) | 0.001 |
| Dysphagia | 1.79 | (1.20, 2.68) | 0.005 |
| ln(time) * Dysphagia | 0.76 | (0.70, 0.83) | <0.001 |
| Jaundice | 6.06 | (3.11, 11.81) | <0.001 |
| ln(time) * Jaundice | 0.76 | (0.66, 0.87) | <0.001 |
| Dyspnoea | 1.70 | (1.46, 1.98) | <0.001 |
| ln(time) * Dyspnoea | 0.81 | (0.77, 0.85) | <0.001 |
| Haemoptysis | 2.77 | (1.69, 4.57) | <0.001 |
| ln(time) * Haemoptysis | 0.79 | (0.70, 0.88) | <0.001 |
| Haematuria | 1.33 | (0.98, 1.80) | 0.066 |
| ln(time) * Haematuria | 0.95 | (0.84, 1.07) | 0.372 |
| Fatigue | 2.34 | (1.97, 2.79) | <0.001 |
| ln(time) * Fatigue | 0.77 | (0.73, 0.81) | <0.001 |
| Night sweats | 4.52 | (2.82, 7.23) | <0.001 |
| ln(time) * Night sweats | 0.71 | (0.66, 0.78) | <0.001 |
| Weight loss | 4.41 | (3.47, 5.59) | <0.001 |
| ln(time) * Weight loss | 0.75 | (0.71, 0.79) | <0.001 |
| Breast lump | 2.14 | (0.95, 4.78) | 0.065 |
| ln(time) * Breast lump | 0.97 | (0.69, 1.36) | 0.848 |

Appendix 4 Table 7. Hazard ratios for other cancers, men.

|  | **Hazard ratio** | |  |
| --- | --- | --- | --- |
| **Outcome: Other cancer** | **Estimate** | **(95% CI)** | **p-value** |
| **Smoking status** |  |  |  |
| Never smoker | (ref) |  |  |
| Ever-smoker | 1.20 | (1.09, 1.32) | <0.001 |
| **Symptoms** |  |  |  |
| Reference cohort | (ref) |  |  |
| Abdominal pain | 1.69 | (1.49, 1.90) | <0.001 |
| ln(time) * Abdominal pain | 0.82 | (0.79, 0.85) | <0.001 |
| Abdominal bloating | 1.40 | (0.98, 1.99) | 0.063 |
| ln(time) * Abdominal bloating | 0.85 | (0.76, 0.95) | 0.004 |
| Rectal bleeding | 1.54 | (1.26, 1.90) | <0.001 |
| ln(time) * Rectal bleeding | 0.87 | (0.81, 0.93) | <0.001 |
| Change in bowel habit | 1.72 | (1.28, 2.32) | <0.001 |
| ln(time) * Change in bowel habit | 0.78 | (0.72, 0.83) | <0.001 |
| Dyspepsia | 1.40 | (1.20, 1.64) | <0.001 |
| ln(time) * Dyspepsia | 0.85 | (0.81, 0.89) | <0.001 |
| Dysphagia | 3.06 | (2.41, 3.89) | <0.001 |
| ln(time) * Dysphagia | 0.77 | (0.73, 0.82) | <0.001 |
| Jaundice | 19.84 | (15.05, 26.16) | <0.001 |
| ln(time) * Jaundice | 0.69 | (0.66, 0.72) | <0.001 |
| Dyspnoea | 1.32 | (1.17, 1.50) | <0.001 |
| ln(time) * Dyspnoea | 0.86 | (0.82, 0.90) | <0.001 |
| Haemoptysis | 2.15 | (1.41, 3.28) | <0.001 |
| ln(time) * Haemoptysis | 0.80 | (0.71, 0.89) | <0.001 |
| Haematuria | 1.27 | (1.00, 1.61) | 0.049 |
| ln(time) * Haematuria | 0.91 | (0.84, 0.99) | 0.032 |
| Fatigue | 1.65 | (1.42, 1.92) | <0.001 |
| ln(time) * Fatigue | 0.81 | (0.77, 0.85) | <0.001 |
| Night sweats | 1.77 | (1.03, 3.07) | 0.040 |
| ln(time) * Night sweats | 0.88 | (0.73, 1.04) | 0.141 |
| Weight loss | 3.17 | (2.57, 3.90) | <0.001 |
| ln(time) * Weight loss | 0.76 | (0.72, 0.79) | <0.001 |
| Breast lump | 7.07 | (5.04, 9.91) | <0.001 |
| ln(time) * Breast lump | 0.67 | (0.64, 0.70) | <0.001 |

Appendix 4 Table 8. Hazard ratios for death (no cancer), men.

|  | **Hazard ratio** | |  |
| --- | --- | --- | --- |
| **Outcome: Death** | **Estimate** | **(95% CI)** | **p-value** |
| **Smoking status** |  |  |  |
| Never smoker | (ref) |  |  |
| Ever-smoker | 1.09 | (1.05, 1.13) | <0.001 |
| **Symptoms** |  |  |  |
| Reference cohort | (ref) |  |  |
| Abdominal pain | 0.70 | (0.66, 0.75) | <0.001 |
| ln(time) * Abdominal pain | 0.90 | (0.88, 0.92) | <0.001 |
| Abdominal bloating | 0.67 | (0.57, 0.79) | <0.001 |
| ln(time) * Abdominal bloating | 0.97 | (0.91, 1.02) | 0.243 |
| Rectal bleeding | 0.76 | (0.69, 0.84) | <0.001 |
| ln(time) * Rectal bleeding | 0.92 | (0.90, 0.95) | <0.001 |
| Change in bowel habit | 0.42 | (0.34, 0.51) | <0.001 |
| ln(time) * Change in bowel habit | 1.06 | (0.97, 1.15) | 0.200 |
| Dyspepsia | 0.58 | (0.53, 0.63) | <0.001 |
| ln(time) * Dyspepsia | 0.96 | (0.93, 0.98) | 0.002 |
| Dysphagia | 1.99 | (1.82, 2.17) | <0.001 |
| ln(time) * Dysphagia | 0.90 | (0.88, 0.92) | <0.001 |
| Jaundice | 4.70 | (3.92, 5.63) | <0.001 |
| ln(time) * Jaundice | 0.84 | (0.81, 0.87) | <0.001 |
| Dyspnoea | 1.40 | (1.34, 1.45) | <0.001 |
| ln(time) * Dyspnoea | 0.93 | (0.92, 0.95) | <0.001 |
| Haemoptysis | 1.33 | (1.11, 1.59) | 0.002 |
| ln(time) * Haemoptysis | 0.95 | (0.90, 1.01) | 0.087 |
| Haematuria | 0.79 | (0.72, 0.86) | <0.001 |
| ln(time) * Haematuria | 0.98 | (0.95, 1.01) | 0.169 |
| Fatigue | 0.95 | (0.90, 1.02) | 0.147 |
| ln(time) * Fatigue | 0.93 | (0.92, 0.95) | <0.001 |
| Night sweats | 0.50 | (0.35, 0.73) | <0.001 |
| ln(time) * Night sweats | 1.27 | (1.01, 1.59) | 0.039 |
| Weight loss | 1.77 | (1.63, 1.92) | <0.001 |
| ln(time) * Weight loss | 0.97 | (0.94, 1.00) | 0.044 |
| Breast lump | 0.50 | (0.32, 0.77) | 0.002 |
| ln(time) * Breast lump | 1.19 | (0.93, 1.51) | 0.162 |

Appendix 4 Table 9. Hazard ratios for breast cancer, women.

|  | **Hazard ratio** | |  |
| --- | --- | --- | --- |
| **Outcome: Breast** | **Estimate** | **(95% CI)** | **p-value** |
| **Smoking status** |  |  |  |
| Never smoker | (ref) |  |  |
| Ever-smoker | 1.05 | (1.01, 1.10) | 0.020 |
| **Symptoms** |  |  |  |
| Reference cohort | (ref) |  |  |
| Abdominal pain | 1.14 | (1.03, 1.26) | 0.009 |
| ln(time) * Abdominal pain | 1.05 | (1.00, 1.11) | 0.052 |
| Abdominal bloating | 1.22 | (0.96, 1.56) | 0.106 |
| ln(time) * Abdominal bloating | 1.07 | (0.93, 1.22) | 0.345 |
| Rectal bleeding | 1.12 | (0.91, 1.37) | 0.275 |
| ln(time) * Rectal bleeding | 0.96 | (0.88, 1.05) | 0.352 |
| Change in bowel habit | 1.32 | (1.02, 1.71) | 0.033 |
| ln(time) * Change in bowel habit | 0.88 | (0.80, 0.97) | 0.012 |
| Dyspepsia | 1.24 | (1.10, 1.41) | <0.001 |
| ln(time) * Dyspepsia | 0.97 | (0.91, 1.03) | 0.270 |
| Dysphagia | 0.86 | (0.61, 1.20) | 0.372 |
| ln(time) * Dysphagia | 0.91 | (0.80, 1.03) | 0.149 |
| Jaundice | 2.24 | (1.06, 4.72) | 0.035 |
| ln(time) * Jaundice | 0.86 | (0.68, 1.08) | 0.196 |
| Dyspnoea | 1.06 | (0.95, 1.18) | 0.266 |
| ln(time) * Dyspnoea | 0.92 | (0.88, 0.97) | <0.001 |
| Haemoptysis | 1.79 | (1.12, 2.86) | 0.015 |
| ln(time) * Haemoptysis | 1.25 | (0.91, 1.72) | 0.162 |
| Haematuria | 1.37 | (1.06, 1.78) | 0.017 |
| ln(time) * Haematuria | 0.97 | (0.86, 1.09) | 0.585 |
| Fatigue | 1.21 | (1.08, 1.35) | <0.001 |
| ln(time) * Fatigue | 0.95 | (0.90, 1.00) | 0.034 |
| Night sweats | 1.35 | (0.85, 2.12) | 0.200 |
| ln(time) * Night sweats | 0.89 | (0.75, 1.05) | 0.165 |
| Weight loss | 1.07 | (0.82, 1.39) | 0.608 |
| ln(time) * Weight loss | 0.84 | (0.77, 0.92) | <0.001 |
| Breast lump | 64.98 | (61.10, 69.10) | <0.001 |
| ln(time) * Breast lump | 0.64 | (0.62, 0.65) | <0.001 |
| Post-menopausal bleed | 1.22 | (0.90, 1.65) | 0.191 |
| ln(time) * Post-menopausal bleed | 1.09 | (0.92, 1.29) | 0.300 |

Appendix 4 Table 10. Hazard ratios for gynaecological cancer, women.

|  | **Hazard ratio** | |  |
| --- | --- | --- | --- |
| **Outcome: Gynaecological** | **Estimate** | **(95% CI)** | **p-value** |
| **Smoking status** |  |  |  |
| Never smoker | (ref) |  |  |
| Ever-smoker | 1.07 | (1.01, 1.15) | 0.032 |
| **Symptoms** |  |  |  |
| Reference cohort | (ref) |  |  |
| Abdominal pain | 2.49 | (2.27, 2.73) | <0.001 |
| ln(time) * Abdominal pain | 0.80 | (0.77, 0.82) | <0.001 |
| Abdominal bloating | 4.75 | (4.03, 5.60) | <0.001 |
| ln(time) * Abdominal bloating | 0.72 | (0.70, 0.75) | <0.001 |
| Rectal bleeding | 1.25 | (0.97, 1.60) | 0.086 |
| ln(time) * Rectal bleeding | 0.86 | (0.79, 0.94) | <0.001 |
| Change in bowel habit | 2.58 | (1.98, 3.36) | <0.001 |
| ln(time) * Change in bowel habit | 0.78 | (0.73, 0.84) | <0.001 |
| Dyspepsia | 1.15 | (0.97, 1.35) | 0.109 |
| ln(time) * Dyspepsia | 0.93 | (0.87, 1.00) | 0.042 |
| Dysphagia | 0.76 | (0.45, 1.30) | 0.318 |
| ln(time) * Dysphagia | 0.85 | (0.71, 1.00) | 0.056 |
| Jaundice | 1.34 | (0.34, 5.38) | 0.676 |
| ln(time) * Jaundice | 1.00 | (0.57, 1.78) | 0.989 |
| Dyspnoea | 1.06 | (0.92, 1.24) | 0.414 |
| ln(time) * Dyspnoea | 0.87 | (0.82, 0.92) | <0.001 |
| Haemoptysis | 0.74 | (0.28, 1.98) | 0.552 |
| ln(time) * Haemoptysis | 0.89 | (0.63, 1.27) | 0.528 |
| Haematuria | 4.57 | (3.72, 5.61) | <0.001 |
| ln(time) * Haematuria | 0.81 | (0.76, 0.86) | <0.001 |
| Fatigue | 1.27 | (1.12, 1.45) | <0.001 |
| ln(time) * Fatigue | 0.95 | (0.90, 1.01) | 0.104 |
| Night sweats | 0.74 | (0.37, 1.49) | 0.402 |
| ln(time) * Night sweats | 0.91 | (0.70, 1.19) | 0.500 |
| Weight loss | 1.74 | (1.30, 2.33) | <0.001 |
| ln(time) * Weight loss | 0.82 | (0.75, 0.89) | <0.001 |
| Breast lump | 1.19 | (0.95, 1.49) | 0.131 |
| ln(time) * Breast lump | 1.08 | (0.96, 1.21) | 0.216 |
| Post-menopausal bleed | 42.73 | (38.56, 47.34) | <0.001 |
| ln(time) * Post-menopausal bleed | 0.68 | (0.66, 0.70) | <0.001 |

Appendix 4 Table 11. Hazard ratios for lung cancer, women.

|  | **Hazard ratio** | |  |
| --- | --- | --- | --- |
| **Outcome: Lung** | **Estimate** | **(95% CI)** | **p-value** |
| **Smoking status** |  |  |  |
| Never smoker | (ref) |  |  |
| Ever-smoker | 4.85 | (4.22, 5.56) | <0.001 |
| **Symptoms** |  |  |  |
| Reference cohort | (ref) |  |  |
| Abdominal pain | 1.00 | (0.86, 1.17) | 0.995 |
| ln(time) * Abdominal pain | 0.90 | (0.85, 0.95) | <0.001 |
| Abdominal bloating | 0.66 | (0.41, 1.05) | 0.081 |
| ln(time) * Abdominal bloating | 0.87 | (0.75, 1.01) | 0.062 |
| Rectal bleeding | 0.59 | (0.40, 0.87) | 0.008 |
| ln(time) * Rectal bleeding | 1.04 | (0.87, 1.26) | 0.644 |
| Change in bowel habit | 0.94 | (0.62, 1.41) | 0.762 |
| ln(time) * Change in bowel habit | 0.95 | (0.81, 1.11) | 0.516 |
| Dyspepsia | 0.69 | (0.55, 0.86) | <0.001 |
| ln(time) * Dyspepsia | 0.92 | (0.84, 0.99) | 0.035 |
| Dysphagia | 2.04 | (1.49, 2.79) | <0.001 |
| ln(time) * Dysphagia | 0.79 | (0.73, 0.85) | <0.001 |
| Jaundice | 0.81 | (0.21, 3.10) | 0.757 |
| ln(time) * Jaundice | 0.55 | (0.46, 0.66) | <0.001 |
| Dyspnoea | 2.57 | (2.30, 2.86) | <0.001 |
| ln(time) * Dyspnoea | 0.81 | (0.78, 0.84) | <0.001 |
| Haemoptysis | 17.20 | (13.94, 21.22) | <0.001 |
| ln(time) * Haemoptysis | 0.73 | (0.70, 0.77) | <0.001 |
| Haematuria | 1.17 | (0.80, 1.70) | 0.424 |
| ln(time) * Haematuria | 1.05 | (0.88, 1.24) | 0.615 |
| Fatigue | 1.33 | (1.14, 1.56) | <0.001 |
| ln(time) * Fatigue | 0.86 | (0.81, 0.90) | <0.001 |
| Night sweats | 1.57 | (0.86, 2.85) | 0.140 |
| ln(time) * Night sweats | 0.91 | (0.73, 1.12) | 0.358 |
| Weight loss | 3.70 | (3.03, 4.52) | <0.001 |
| ln(time) * Weight loss | 0.78 | (0.74, 0.82) | <0.001 |
| Breast lump | 0.82 | (0.52, 1.28) | 0.384 |
| ln(time) * Breast lump | 0.88 | (0.77, 1.02) | 0.086 |
| Post-menopausal bleed | 0.16 | (0.05, 0.51) | 0.002 |
| ln(time) * Post-menopausal bleed | 0.88 | (0.62, 1.27) | 0.506 |

Appendix 4 Table 12. Hazard ratios for upper GI cancer, women.

|  | **Hazard ratio** | |  |
| --- | --- | --- | --- |
| **Outcome: Upper GI** | **Estimate** | **(95% CI)** | **p-value** |
| **Smoking status** |  |  |  |
| Never smoker | (ref) |  |  |
| Ever-smoker | 1.20 | (1.10, 1.32) | <0.001 |
| **Symptoms** |  |  |  |
| Reference cohort | (ref) |  |  |
| Abdominal pain | 3.37 | (2.95, 3.85) | <0.001 |
| ln(time) * Abdominal pain | 0.75 | (0.72, 0.79) | <0.001 |
| Abdominal bloating | 2.88 | (2.14, 3.88) | <0.001 |
| ln(time) * Abdominal bloating | 0.78 | (0.72, 0.84) | <0.001 |
| Rectal bleeding | 1.10 | (0.76, 1.58) | 0.624 |
| ln(time) * Rectal bleeding | 0.76 | (0.69, 0.82) | <0.001 |
| Change in bowel habit | 1.87 | (1.29, 2.73) | 0.001 |
| ln(time) * Change in bowel habit | 0.79 | (0.71, 0.88) | <0.001 |
| Dyspepsia | 3.28 | (2.81, 3.84) | <0.001 |
| ln(time) * Dyspepsia | 0.77 | (0.73, 0.81) | <0.001 |
| Dysphagia | 16.36 | (13.95, 19.18) | <0.001 |
| ln(time) * Dysphagia | 0.66 | (0.63, 0.69) | <0.001 |
| Jaundice | 122.63 | (102.30, 147.01) | <0.001 |
| ln(time) * Jaundice | 0.66 | (0.63, 0.69) | <0.001 |
| Dyspnoea | 0.88 | (0.73, 1.08) | 0.218 |
| ln(time) * Dyspnoea | 0.90 | (0.84, 0.97) | 0.009 |
| Haemoptysis | 0.61 | (0.15, 2.45) | 0.487 |
| ln(time) * Haemoptysis | 0.88 | (0.56, 1.40) | 0.598 |
| Haematuria | 0.49 | (0.23, 1.03) | 0.058 |
| ln(time) * Haematuria | 0.78 | (0.65, 0.93) | 0.007 |
| Fatigue | 1.04 | (0.84, 1.29) | 0.689 |
| ln(time) * Fatigue | 0.81 | (0.76, 0.87) | <0.001 |
| Night sweats | 0.50 | (0.13, 2.01) | 0.331 |
| ln(time) * Night sweats | 0.77 | (0.56, 1.06) | 0.104 |
| Weight loss | 5.57 | (4.52, 6.87) | <0.001 |
| ln(time) * Weight loss | 0.74 | (0.70, 0.78) | <0.001 |
| Breast lump | 0.78 | (0.44, 1.38) | 0.389 |
| ln(time) * Breast lump | 0.98 | (0.76, 1.25) | 0.853 |
| Post-menopausal bleed | 1.06 | (0.58, 1.92) | 0.856 |
| ln(time) * Post-menopausal bleed | 1.01 | (0.77, 1.33) | 0.938 |

Appendix 4 Table 13. Hazard ratios for lower GI cancer, women.

|  | **Hazard ratio** | |  |
| --- | --- | --- | --- |
| **Outcome: Lower GI** | **Estimate** | **(95% CI)** | **p-value** |
| **Smoking status** |  |  |  |
| Never smoker | (ref) |  |  |
| Ever-smoker | 0.96 | (0.89, 1.04) | 0.339 |
| **Symptoms** |  |  |  |
| Reference cohort | (ref) |  |  |
| Abdominal pain | 4.35 | (3.89, 4.86) | <0.001 |
| ln(time) * Abdominal pain | 0.71 | (0.68, 0.74) | <0.001 |
| Abdominal bloating | 3.04 | (2.34, 3.95) | <0.001 |
| ln(time) * Abdominal bloating | 0.72 | (0.67, 0.77) | <0.001 |
| Rectal bleeding | 14.74 | (13.08, 16.62) | <0.001 |
| ln(time) * Rectal bleeding | 0.68 | (0.65, 0.72) | <0.001 |
| Change in bowel habit | 12.18 | (10.45, 14.21) | <0.001 |
| ln(time) * Change in bowel habit | 0.66 | (0.63, 0.69) | <0.001 |
| Dyspepsia | 1.28 | (1.05, 1.56) | 0.013 |
| ln(time) * Dyspepsia | 0.84 | (0.78, 0.90) | <0.001 |
| Dysphagia | 1.35 | (0.89, 2.05) | 0.157 |
| ln(time) * Dysphagia | 0.85 | (0.73, 0.98) | 0.029 |
| Jaundice | 2.05 | (0.65, 6.45) | 0.222 |
| ln(time) * Jaundice | 0.70 | (0.57, 0.86) | <0.001 |
| Dyspnoea | 1.26 | (1.07, 1.47) | 0.004 |
| ln(time) * Dyspnoea | 0.88 | (0.82, 0.94) | <0.001 |
| Haemoptysis | 1.01 | (0.38, 2.71) | 0.977 |
| ln(time) * Haemoptysis | 0.74 | (0.58, 0.94) | 0.015 |
| Haematuria | 0.97 | (0.60, 1.57) | 0.892 |
| ln(time) * Haematuria | 0.84 | (0.71, 0.99) | 0.041 |
| Fatigue | 1.67 | (1.43, 1.96) | <0.001 |
| ln(time) * Fatigue | 0.78 | (0.74, 0.83) | <0.001 |
| Night sweats | 1.48 | (0.73, 2.99) | 0.272 |
| ln(time) * Night sweats | 1.25 | (0.76, 2.08) | 0.380 |
| Weight loss | 2.82 | (2.19, 3.64) | <0.001 |
| ln(time) * Weight loss | 0.71 | (0.67, 0.76) | <0.001 |
| Breast lump | 0.54 | (0.30, 0.99) | 0.046 |
| ln(time) * Breast lump | 0.95 | (0.73, 1.25) | 0.729 |
| Post-menopausal bleed | 1.07 | (0.63, 1.81) | 0.814 |
| ln(time) * Post-menopausal bleed | 0.87 | (0.71, 1.08) | 0.204 |

Appendix 4 Table 14. Hazard ratios for urological cancer, women.

|  | **Hazard ratio** | |  |
| --- | --- | --- | --- |
| **Outcome: Urological** | **Estimate** | **(95% CI)** | **p-value** |
| **Smoking status** |  |  |  |
| Never smoker | (ref) |  |  |
| Ever-smoker | 1.25 | (1.10, 1.42) | <0.001 |
| **Symptoms** |  |  |  |
| Reference cohort | (ref) |  |  |
| Abdominal pain | 2.23 | (1.80, 2.75) | <0.001 |
| ln(time) * Abdominal pain | 0.82 | (0.77, 0.88) | <0.001 |
| Abdominal bloating | 1.56 | (0.89, 2.73) | 0.119 |
| ln(time) * Abdominal bloating | 0.95 | (0.76, 1.19) | 0.674 |
| Rectal bleeding | 1.77 | (1.17, 2.68) | 0.007 |
| ln(time) * Rectal bleeding | 0.97 | (0.82, 1.14) | 0.710 |
| Change in bowel habit | 1.71 | (0.98, 3.00) | 0.060 |
| ln(time) * Change in bowel habit | 0.89 | (0.74, 1.08) | 0.240 |
| Dyspepsia | 0.64 | (0.42, 0.98) | 0.040 |
| ln(time) * Dyspepsia | 0.86 | (0.76, 0.98) | 0.029 |
| Dysphagia | 1.92 | (1.09, 3.36) | 0.023 |
| ln(time) * Dysphagia | 0.96 | (0.77, 1.19) | 0.700 |
| Jaundice | 5.11 | (1.62, 16.14) | 0.005 |
| ln(time) * Jaundice | 0.73 | (0.60, 0.89) | 0.002 |
| Dyspnoea | 1.58 | (1.26, 1.99) | <0.001 |
| ln(time) * Dyspnoea | 1.03 | (0.93, 1.14) | 0.527 |
| Haemoptysis | 0.00 | (0.00, 0.00) | 0.999 |
| ln(time) * Haemoptysis | 2.54 | (0.00, 0.00) | 1.000 |
| Haematuria | 57.06 | (48.37, 67.29) | <0.001 |
| ln(time) * Haematuria | 0.72 | (0.68, 0.77) | <0.001 |
| Fatigue | 1.32 | (1.01, 1.74) | 0.046 |
| ln(time) * Fatigue | 0.95 | (0.86, 1.06) | 0.391 |
| Night sweats | 2.04 | (0.76, 5.49) | 0.158 |
| ln(time) * Night sweats | 0.76 | (0.64, 0.90) | 0.002 |
| Weight loss | 2.26 | (1.45, 3.50) | <0.001 |
| ln(time) * Weight loss | 0.77 | (0.69, 0.85) | <0.001 |
| Breast lump | 1.01 | (0.50, 2.06) | 0.970 |
| ln(time) * Breast lump | 1.15 | (0.79, 1.69) | 0.467 |
| Post-menopausal bleed | 4.09 | (2.61, 6.41) | <0.001 |
| ln(time) * Post-menopausal bleed | 0.76 | (0.68, 0.84) | <0.001 |

Appendix 4 Table 15. Hazard ratios for haematological cancer, women.

|  | **Hazard ratio** | |  |
| --- | --- | --- | --- |
| **Outcome: Haematological** | **Estimate** | **(95% CI)** | **p-value** |
| **Smoking status** |  |  |  |
| Never smoker | (ref) |  |  |
| Ever-smoker | 0.98 | (0.88, 1.11) | 0.794 |
| **Symptoms** |  |  |  |
| Reference cohort | (ref) |  |  |
| Abdominal pain | 2.16 | (1.82, 2.56) | <0.001 |
| ln(time) * Abdominal pain | 0.82 | (0.78, 0.88) | <0.001 |
| Abdominal bloating | 1.39 | (0.87, 2.24) | 0.169 |
| ln(time) * Abdominal bloating | 0.84 | (0.73, 0.98) | 0.026 |
| Rectal bleeding | 1.51 | (1.05, 2.17) | 0.026 |
| ln(time) * Rectal bleeding | 0.83 | (0.74, 0.93) | 0.001 |
| Change in bowel habit | 1.14 | (0.65, 1.98) | 0.648 |
| ln(time) * Change in bowel habit | 0.92 | (0.75, 1.14) | 0.460 |
| Dyspepsia | 1.64 | (1.31, 2.06) | <0.001 |
| ln(time) * Dyspepsia | 0.88 | (0.81, 0.95) | 0.002 |
| Dysphagia | 1.82 | (1.13, 2.93) | 0.013 |
| ln(time) * Dysphagia | 1.05 | (0.83, 1.32) | 0.708 |
| Jaundice | 0.45 | (0.05, 4.23) | 0.487 |
| ln(time) * Jaundice | 0.51 | (0.40, 0.65) | <0.001 |
| Dyspnoea | 1.83 | (1.52, 2.19) | <0.001 |
| ln(time) * Dyspnoea | 0.82 | (0.77, 0.87) | <0.001 |
| Haemoptysis | 3.38 | (1.68, 6.81) | <0.001 |
| ln(time) * Haemoptysis | 0.78 | (0.66, 0.93) | 0.005 |
| Haematuria | 1.34 | (0.78, 2.28) | 0.287 |
| ln(time) * Haematuria | 0.83 | (0.71, 0.96) | 0.013 |
| Fatigue | 2.35 | (1.96, 2.82) | <0.001 |
| ln(time) * Fatigue | 0.79 | (0.74, 0.84) | <0.001 |
| Night sweats | 4.05 | (2.33, 7.05) | <0.001 |
| ln(time) * Night sweats | 0.77 | (0.67, 0.89) | <0.001 |
| Weight loss | 4.04 | (3.04, 5.37) | <0.001 |
| ln(time) * Weight loss | 0.76 | (0.71, 0.82) | <0.001 |
| Breast lump | 2.56 | (1.83, 3.60) | <0.001 |
| ln(time) * Breast lump | 0.73 | (0.68, 0.79) | <0.001 |
| Post-menopausal bleed | 1.33 | (0.73, 2.43) | 0.351 |
| ln(time) * Post-menopausal bleed | 0.94 | (0.74, 1.19) | 0.593 |

Appendix 4 Table 16. Hazard ratios for other cancers, women.

|  | **Hazard ratio** | |  |
| --- | --- | --- | --- |
| **Outcome: Other cancer** | **Estimate** | **(95% CI)** | **p-value** |
| **Smoking status** |  |  |  |
| Never smoker | (ref) |  |  |
| Ever-smoker | 1.06 | (0.98, 1.15) | 0.149 |
| **Symptoms** |  |  |  |
| Reference cohort | (ref) |  |  |
| Abdominal pain | 1.92 | (1.72, 2.15) | <0.001 |
| ln(time) * Abdominal pain | 0.79 | (0.76, 0.83) | <0.001 |
| Abdominal bloating | 1.72 | (1.31, 2.27) | <0.001 |
| ln(time) * Abdominal bloating | 0.82 | (0.75, 0.90) | <0.001 |
| Rectal bleeding | 2.27 | (1.87, 2.76) | <0.001 |
| ln(time) * Rectal bleeding | 0.77 | (0.73, 0.82) | <0.001 |
| Change in bowel habit | 2.16 | (1.65, 2.83) | <0.001 |
| ln(time) * Change in bowel habit | 0.81 | (0.75, 0.88) | <0.001 |
| Dyspepsia | 1.27 | (1.08, 1.50) | 0.004 |
| ln(time) * Dyspepsia | 0.84 | (0.80, 0.90) | <0.001 |
| Dysphagia | 2.23 | (1.69, 2.94) | <0.001 |
| ln(time) * Dysphagia | 0.78 | (0.72, 0.84) | <0.001 |
| Jaundice | 19.08 | (13.98, 26.04) | <0.001 |
| ln(time) * Jaundice | 0.66 | (0.62, 0.70) | <0.001 |
| Dyspnoea | 1.10 | (0.96, 1.27) | 0.179 |
| ln(time) * Dyspnoea | 0.86 | (0.82, 0.91) | <0.001 |
| Haemoptysis | 1.62 | (0.84, 3.13) | 0.150 |
| ln(time) * Haemoptysis | 0.78 | (0.66, 0.94) | 0.008 |
| Haematuria | 1.53 | (1.10, 2.12) | 0.011 |
| ln(time) * Haematuria | 0.97 | (0.84, 1.12) | 0.650 |
| Fatigue | 1.52 | (1.33, 1.74) | <0.001 |
| ln(time) * Fatigue | 0.83 | (0.79, 0.87) | <0.001 |
| Night sweats | 0.96 | (0.48, 1.92) | 0.898 |
| ln(time) * Night sweats | 1.01 | (0.72, 1.42) | 0.943 |
| Weight loss | 2.27 | (1.79, 2.88) | <0.001 |
| ln(time) * Weight loss | 0.78 | (0.73, 0.83) | <0.001 |
| Breast lump | 1.24 | (0.93, 1.66) | 0.149 |
| ln(time) * Breast lump | 0.92 | (0.82, 1.03) | 0.139 |
| Post-menopausal bleed | 1.08 | (0.70, 1.66) | 0.733 |
| ln(time) * Post-menopausal bleed | 0.80 | (0.71, 0.91) | <0.001 |

Appendix 4 Table 17. Hazard ratios for death (no cancer), women.

|  | **Hazard ratio** | |  |
| --- | --- | --- | --- |
| **Outcome: Death** | **Estimate** | **(95% CI)** | **p-value** |
| **Smoking status** |  |  |  |
| Never smoker | (ref) |  |  |
| Ever-smoker | 0.97 | (0.94, 1.00) | 0.025 |
| **Symptoms** |  |  |  |
| Reference cohort | (ref) |  |  |
| Abdominal pain | 0.73 | (0.69, 0.77) | <0.001 |
| ln(time) * Abdominal pain | 0.89 | (0.88, 0.91) | <0.001 |
| Abdominal bloating | 0.72 | (0.62, 0.82) | <0.001 |
| ln(time) * Abdominal bloating | 0.94 | (0.89, 0.98) | 0.010 |
| Rectal bleeding | 0.93 | (0.85, 1.01) | 0.091 |
| ln(time) * Rectal bleeding | 0.92 | (0.89, 0.95) | <0.001 |
| Change in bowel habit | 0.43 | (0.36, 0.52) | <0.001 |
| ln(time) * Change in bowel habit | 1.04 | (0.96, 1.13) | 0.361 |
| Dyspepsia | 0.52 | (0.48, 0.57) | <0.001 |
| ln(time) * Dyspepsia | 0.97 | (0.94, 1.00) | 0.047 |
| Dysphagia | 2.07 | (1.92, 2.22) | <0.001 |
| ln(time) * Dysphagia | 0.88 | (0.86, 0.90) | <0.001 |
| Jaundice | 3.49 | (2.88, 4.23) | <0.001 |
| ln(time) * Jaundice | 0.86 | (0.82, 0.90) | <0.001 |
| Dyspnoea | 1.28 | (1.24, 1.33) | <0.001 |
| ln(time) * Dyspnoea | 0.91 | (0.90, 0.92) | <0.001 |
| Haemoptysis | 0.97 | (0.75, 1.27) | 0.852 |
| ln(time) * Haemoptysis | 0.87 | (0.81, 0.94) | <0.001 |
| Haematuria | 0.93 | (0.82, 1.05) | 0.263 |
| ln(time) * Haematuria | 1.01 | (0.96, 1.06) | 0.770 |
| Fatigue | 0.84 | (0.79, 0.88) | <0.001 |
| ln(time) * Fatigue | 0.89 | (0.87, 0.90) | <0.001 |
| Night sweats | 0.34 | (0.21, 0.54) | <0.001 |
| ln(time) * Night sweats | 1.03 | (0.83, 1.26) | 0.806 |
| Weight loss | 1.65 | (1.53, 1.77) | <0.001 |
| ln(time) * Weight loss | 0.94 | (0.91, 0.96) | <0.001 |
| Breast lump | 0.55 | (0.45, 0.66) | <0.001 |
| ln(time) * Breast lump | 1.07 | (0.99, 1.17) | 0.099 |
| Post-menopausal bleed | 0.57 | (0.46, 0.72) | <0.001 |
| ln(time) * Post-menopausal bleed | 1.04 | (0.94, 1.16) | 0.414 |

### Appendix 5. Table showing simulated outcomes at 12 months after index symptom by age, sex and smoking status.

*Appendix 5 Table 1. Estimated probability of cancer and mortality within 12 months for patients presenting aged 30, 40, 50, 60, 70, 80, 90 with each index symptom. Only probabilities greater than 0.001 are shown.*

| **Sex** | **Index symptom** | **Smoking status** | **Age (years)** | **Outcome event** | **Probability of outcome** | **(95% CI)** |
| --- | --- | --- | --- | --- | --- | --- |
| Women | Abdominal pain | Never smoker | 30 | Gyanecological | 0.008 | (0.003, 0.021) |
| Women | Abdominal pain | Never smoker | 40 | Gyanecological | 0.003 | (0.002, 0.007) |
| Women | Abdominal pain | Never smoker | 50 | Breast | 0.002 | (<0.001, 0.003) |
| Women | Abdominal pain | Never smoker | 50 | Gyanecological | 0.003 | (0.001, 0.005) |
| Women | Abdominal pain | Never smoker | 50 | Lower GI | 0.003 | (0.001, 0.006) |
| Women | Abdominal pain | Never smoker | 50 | Other cancers | 0.002 | (<0.001, 0.005) |
| Women | Abdominal pain | Never smoker | 50 | Death w/o cancer | 0.002 | (<0.001, 0.004) |
| Women | Abdominal pain | Never smoker | 60 | Breast | 0.003 | (0.001, 0.009) |
| Women | Abdominal pain | Never smoker | 60 | Gyanecological | 0.004 | (0.002, 0.007) |
| Women | Abdominal pain | Never smoker | 60 | Upper GI | 0.003 | (0.002, 0.006) |
| Women | Abdominal pain | Never smoker | 60 | Lower GI | 0.005 | (0.003, 0.011) |
| Women | Abdominal pain | Never smoker | 60 | Haematological | 0.002 | (<0.001, 0.004) |
| Women | Abdominal pain | Never smoker | 60 | Other cancers | 0.004 | (0.001, 0.010) |
| Women | Abdominal pain | Never smoker | 60 | Death w/o cancer | 0.004 | (0.001, 0.011) |
| Women | Abdominal pain | Never smoker | 70 | Breast | 0.005 | (0.002, 0.011) |
| Women | Abdominal pain | Never smoker | 70 | Gyanecological | 0.007 | (0.003, 0.017) |
| Women | Abdominal pain | Never smoker | 70 | Upper GI | 0.007 | (0.003, 0.015) |
| Women | Abdominal pain | Never smoker | 70 | Lower GI | 0.012 | (0.007, 0.022) |
| Women | Abdominal pain | Never smoker | 70 | Urological | 0.002 | (0.001, 0.004) |
| Women | Abdominal pain | Never smoker | 70 | Haematological | 0.003 | (0.002, 0.008) |
| Women | Abdominal pain | Never smoker | 70 | Other cancers | 0.006 | (0.002, 0.014) |
| Women | Abdominal pain | Never smoker | 70 | Death w/o cancer | 0.012 | (0.007, 0.019) |
| Women | Abdominal pain | Never smoker | 80 | Breast | 0.007 | (0.003, 0.015) |
| Women | Abdominal pain | Never smoker | 80 | Gyanecological | 0.007 | (0.004, 0.013) |
| Women | Abdominal pain | Never smoker | 80 | Lung | 0.001 | (<0.001, 0.003) |
| Women | Abdominal pain | Never smoker | 80 | Upper GI | 0.010 | (0.006, 0.017) |
| Women | Abdominal pain | Never smoker | 80 | Lower GI | 0.017 | (0.011, 0.026) |
| Women | Abdominal pain | Never smoker | 80 | Urological | 0.003 | (0.001, 0.007) |
| Women | Abdominal pain | Never smoker | 80 | Haematological | 0.004 | (0.002, 0.008) |
| Women | Abdominal pain | Never smoker | 80 | Other cancers | 0.008 | (0.004, 0.019) |
| Women | Abdominal pain | Never smoker | 80 | Death w/o cancer | 0.040 | (0.027, 0.059) |
| Women | Abdominal pain | Never smoker | 90 | Breast | 0.008 | (0.004, 0.016) |
| Women | Abdominal pain | Never smoker | 90 | Gyanecological | 0.005 | (0.002, 0.012) |
| Women | Abdominal pain | Never smoker | 90 | Upper GI | 0.010 | (0.004, 0.027) |
| Women | Abdominal pain | Never smoker | 90 | Lower GI | 0.018 | (0.009, 0.033) |
| Women | Abdominal pain | Never smoker | 90 | Urological | 0.003 | (<0.001, 0.000) |
| Women | Abdominal pain | Never smoker | 90 | Haematological | 0.004 | (0.002, 0.009) |
| Women | Abdominal pain | Never smoker | 90 | Other cancers | 0.010 | (0.005, 0.018) |
| Women | Abdominal pain | Never smoker | 90 | Death w/o cancer | 0.124 | (0.107, 0.143) |
| Women | Abdominal pain | Ever smoker | 30 | Gyanecological | 0.010 | (0.003, 0.034) |
| Women | Abdominal pain | Ever smoker | 40 | Gyanecological | 0.004 | (0.001, 0.011) |
| Women | Abdominal pain | Ever smoker | 40 | Other cancers | 0.001 | (<0.001, 0.002) |
| Women | Abdominal pain | Ever smoker | 50 | Breast | 0.002 | (<0.001, 0.004) |
| Women | Abdominal pain | Ever smoker | 50 | Gyanecological | 0.003 | (0.001, 0.005) |
| Women | Abdominal pain | Ever smoker | 50 | Upper GI | 0.001 | (<0.001, 0.003) |
| Women | Abdominal pain | Ever smoker | 50 | Lower GI | 0.002 | (0.001, 0.005) |
| Women | Abdominal pain | Ever smoker | 50 | Other cancers | 0.002 | (0.001, 0.005) |
| Women | Abdominal pain | Ever smoker | 50 | Death w/o cancer | 0.002 | (<0.001, 0.004) |
| Women | Abdominal pain | Ever smoker | 60 | Breast | 0.004 | (0.002, 0.008) |
| Women | Abdominal pain | Ever smoker | 60 | Gyanecological | 0.004 | (0.002, 0.010) |
| Women | Abdominal pain | Ever smoker | 60 | Lung | 0.002 | (<0.001, 0.006) |
| Women | Abdominal pain | Ever smoker | 60 | Upper GI | 0.003 | (0.001, 0.007) |
| Women | Abdominal pain | Ever smoker | 60 | Lower GI | 0.005 | (0.002, 0.018) |
| Women | Abdominal pain | Ever smoker | 60 | Urological | 0.001 | (<0.001, 0.002) |
| Women | Abdominal pain | Ever smoker | 60 | Haematological | 0.002 | (<0.001, 0.004) |
| Women | Abdominal pain | Ever smoker | 60 | Other cancers | 0.003 | (0.001, 0.010) |
| Women | Abdominal pain | Ever smoker | 60 | Death w/o cancer | 0.003 | (0.002, 0.007) |
| Women | Abdominal pain | Ever smoker | 70 | Breast | 0.005 | (0.002, 0.013) |
| Women | Abdominal pain | Ever smoker | 70 | Gyanecological | 0.008 | (0.004, 0.017) |
| Women | Abdominal pain | Ever smoker | 70 | Lung | 0.005 | (0.002, 0.011) |
| Women | Abdominal pain | Ever smoker | 70 | Upper GI | 0.008 | (0.003, 0.022) |
| Women | Abdominal pain | Ever smoker | 70 | Lower GI | 0.012 | (0.007, 0.019) |
| Women | Abdominal pain | Ever smoker | 70 | Urological | 0.002 | (<0.001, 0.007) |
| Women | Abdominal pain | Ever smoker | 70 | Haematological | 0.003 | (0.002, 0.005) |
| Women | Abdominal pain | Ever smoker | 70 | Other cancers | 0.006 | (0.002, 0.017) |
| Women | Abdominal pain | Ever smoker | 70 | Death w/o cancer | 0.012 | (0.006, 0.024) |
| Women | Abdominal pain | Ever smoker | 80 | Breast | 0.006 | (<0.001, 0.000) |
| Women | Abdominal pain | Ever smoker | 80 | Gyanecological | 0.007 | (0.004, 0.014) |
| Women | Abdominal pain | Ever smoker | 80 | Lung | 0.006 | (0.003, 0.015) |
| Women | Abdominal pain | Ever smoker | 80 | Upper GI | 0.012 | (0.006, 0.023) |
| Women | Abdominal pain | Ever smoker | 80 | Lower GI | 0.016 | (0.010, 0.027) |
| Women | Abdominal pain | Ever smoker | 80 | Urological | 0.004 | (0.001, 0.009) |
| Women | Abdominal pain | Ever smoker | 80 | Haematological | 0.004 | (0.002, 0.009) |
| Women | Abdominal pain | Ever smoker | 80 | Other cancers | 0.008 | (0.004, 0.016) |
| Women | Abdominal pain | Ever smoker | 80 | Death w/o cancer | 0.039 | (0.028, 0.053) |
| Women | Abdominal pain | Ever smoker | 90 | Breast | 0.008 | (0.003, 0.021) |
| Women | Abdominal pain | Ever smoker | 90 | Gyanecological | 0.005 | (0.002, 0.016) |
| Women | Abdominal pain | Ever smoker | 90 | Lung | 0.005 | (0.003, 0.008) |
| Women | Abdominal pain | Ever smoker | 90 | Upper GI | 0.012 | (0.006, 0.021) |
| Women | Abdominal pain | Ever smoker | 90 | Lower GI | 0.017 | (0.011, 0.027) |
| Women | Abdominal pain | Ever smoker | 90 | Urological | 0.004 | (0.001, 0.011) |
| Women | Abdominal pain | Ever smoker | 90 | Haematological | 0.004 | (0.002, 0.009) |
| Women | Abdominal pain | Ever smoker | 90 | Other cancers | 0.011 | (0.005, 0.021) |
| Women | Abdominal pain | Ever smoker | 90 | Death w/o cancer | 0.120 | (0.102, 0.140) |
| Women | Abdominal bloating | Never smoker | 30 | Gyanecological | 0.016 | (0.010, 0.027) |
| Women | Abdominal bloating | Never smoker | 40 | Gyanecological | 0.007 | (0.003, 0.018) |
| Women | Abdominal bloating | Never smoker | 50 | Breast | 0.002 | (0.001, 0.004) |
| Women | Abdominal bloating | Never smoker | 50 | Gyanecological | 0.005 | (0.002, 0.015) |
| Women | Abdominal bloating | Never smoker | 50 | Lower GI | 0.002 | (<0.001, 0.004) |
| Women | Abdominal bloating | Never smoker | 50 | Other cancers | 0.002 | (<0.001, 0.003) |
| Women | Abdominal bloating | Never smoker | 50 | Death w/o cancer | 0.002 | (<0.001, 0.004) |
| Women | Abdominal bloating | Never smoker | 60 | Breast | 0.003 | (0.001, 0.008) |
| Women | Abdominal bloating | Never smoker | 60 | Gyanecological | 0.009 | (0.004, 0.022) |
| Women | Abdominal bloating | Never smoker | 60 | Upper GI | 0.002 | (<0.001, 0.005) |
| Women | Abdominal bloating | Never smoker | 60 | Lower GI | 0.004 | (0.002, 0.007) |
| Women | Abdominal bloating | Never smoker | 60 | Other cancers | 0.003 | (0.001, 0.009) |
| Women | Abdominal bloating | Never smoker | 60 | Death w/o cancer | 0.003 | (0.001, 0.008) |
| Women | Abdominal bloating | Never smoker | 70 | Breast | 0.005 | (0.002, 0.014) |
| Women | Abdominal bloating | Never smoker | 70 | Gyanecological | 0.015 | (0.009, 0.024) |
| Women | Abdominal bloating | Never smoker | 70 | Upper GI | 0.006 | (0.003, 0.015) |
| Women | Abdominal bloating | Never smoker | 70 | Lower GI | 0.009 | (0.004, 0.018) |
| Women | Abdominal bloating | Never smoker | 70 | Urological | 0.002 | (<0.001, 0.003) |
| Women | Abdominal bloating | Never smoker | 70 | Haematological | 0.002 | (<0.001, 0.004) |
| Women | Abdominal bloating | Never smoker | 70 | Other cancers | 0.005 | (0.002, 0.014) |
| Women | Abdominal bloating | Never smoker | 70 | Death w/o cancer | 0.011 | (0.006, 0.020) |
| Women | Abdominal bloating | Never smoker | 80 | Breast | 0.007 | (0.003, 0.015) |
| Women | Abdominal bloating | Never smoker | 80 | Gyanecological | 0.014 | (0.010, 0.021) |
| Women | Abdominal bloating | Never smoker | 80 | Upper GI | 0.008 | (0.005, 0.014) |
| Women | Abdominal bloating | Never smoker | 80 | Lower GI | 0.012 | (0.007, 0.021) |
| Women | Abdominal bloating | Never smoker | 80 | Urological | 0.002 | (<0.001, 0.004) |
| Women | Abdominal bloating | Never smoker | 80 | Haematological | 0.003 | (0.001, 0.006) |
| Women | Abdominal bloating | Never smoker | 80 | Other cancers | 0.007 | (0.003, 0.016) |
| Women | Abdominal bloating | Never smoker | 80 | Death w/o cancer | 0.038 | (0.025, 0.057) |
| Women | Abdominal bloating | Never smoker | 90 | Breast | 0.008 | (0.004, 0.015) |
| Women | Abdominal bloating | Never smoker | 90 | Gyanecological | 0.010 | (0.006, 0.016) |
| Women | Abdominal bloating | Never smoker | 90 | Upper GI | 0.008 | (0.003, 0.025) |
| Women | Abdominal bloating | Never smoker | 90 | Lower GI | 0.013 | (0.006, 0.025) |
| Women | Abdominal bloating | Never smoker | 90 | Urological | 0.002 | (0.001, 0.005) |
| Women | Abdominal bloating | Never smoker | 90 | Haematological | 0.003 | (0.001, 0.007) |
| Women | Abdominal bloating | Never smoker | 90 | Other cancers | 0.009 | (0.004, 0.019) |
| Women | Abdominal bloating | Never smoker | 90 | Death w/o cancer | 0.119 | (0.098, 0.143) |
| Women | Abdominal bloating | Ever smoker | 30 | Gyanecological | 0.017 | (0.010, 0.029) |
| Women | Abdominal bloating | Ever smoker | 40 | Breast | 0.001 | (<0.001, 0.001) |
| Women | Abdominal bloating | Ever smoker | 40 | Gyanecological | 0.008 | (0.004, 0.014) |
| Women | Abdominal bloating | Ever smoker | 50 | Breast | 0.002 | (0.001, 0.004) |
| Women | Abdominal bloating | Ever smoker | 50 | Gyanecological | 0.005 | (0.003, 0.009) |
| Women | Abdominal bloating | Ever smoker | 50 | Upper GI | 0.001 | (<0.001, 0.002) |
| Women | Abdominal bloating | Ever smoker | 50 | Lower GI | 0.002 | (<0.001, 0.003) |
| Women | Abdominal bloating | Ever smoker | 50 | Other cancers | 0.002 | (<0.001, 0.004) |
| Women | Abdominal bloating | Ever smoker | 50 | Death w/o cancer | 0.002 | (<0.001, 0.004) |
| Women | Abdominal bloating | Ever smoker | 60 | Breast | 0.003 | (0.001, 0.007) |
| Women | Abdominal bloating | Ever smoker | 60 | Gyanecological | 0.009 | (0.005, 0.016) |
| Women | Abdominal bloating | Ever smoker | 60 | Lung | 0.002 | (0.001, 0.003) |
| Women | Abdominal bloating | Ever smoker | 60 | Upper GI | 0.003 | (0.002, 0.005) |
| Women | Abdominal bloating | Ever smoker | 60 | Lower GI | 0.003 | (0.001, 0.009) |
| Women | Abdominal bloating | Ever smoker | 60 | Haematological | 0.001 | (<0.001, 0.002) |
| Women | Abdominal bloating | Ever smoker | 60 | Other cancers | 0.003 | (0.001, 0.008) |
| Women | Abdominal bloating | Ever smoker | 60 | Death w/o cancer | 0.003 | (0.001, 0.006) |
| Women | Abdominal bloating | Ever smoker | 70 | Breast | 0.006 | (0.003, 0.012) |
| Women | Abdominal bloating | Ever smoker | 70 | Gyanecological | 0.016 | (0.010, 0.027) |
| Women | Abdominal bloating | Ever smoker | 70 | Lung | 0.003 | (0.001, 0.008) |
| Women | Abdominal bloating | Ever smoker | 70 | Upper GI | 0.008 | (0.003, 0.020) |
| Women | Abdominal bloating | Ever smoker | 70 | Lower GI | 0.008 | (0.004, 0.014) |
| Women | Abdominal bloating | Ever smoker | 70 | Urological | 0.002 | (<0.001, 0.004) |
| Women | Abdominal bloating | Ever smoker | 70 | Haematological | 0.002 | (0.001, 0.003) |
| Women | Abdominal bloating | Ever smoker | 70 | Other cancers | 0.006 | (0.002, 0.016) |
| Women | Abdominal bloating | Ever smoker | 70 | Death w/o cancer | 0.011 | (0.005, 0.023) |
| Women | Abdominal bloating | Ever smoker | 80 | Breast | 0.007 | (0.003, 0.017) |
| Women | Abdominal bloating | Ever smoker | 80 | Gyanecological | 0.015 | (0.010, 0.023) |
| Women | Abdominal bloating | Ever smoker | 80 | Lung | 0.004 | (0.001, 0.011) |
| Women | Abdominal bloating | Ever smoker | 80 | Upper GI | 0.009 | (0.004, 0.020) |
| Women | Abdominal bloating | Ever smoker | 80 | Lower GI | 0.011 | (0.006, 0.021) |
| Women | Abdominal bloating | Ever smoker | 80 | Urological | 0.002 | (<0.001, 0.006) |
| Women | Abdominal bloating | Ever smoker | 80 | Haematological | 0.003 | (0.001, 0.004) |
| Women | Abdominal bloating | Ever smoker | 80 | Other cancers | 0.007 | (0.004, 0.014) |
| Women | Abdominal bloating | Ever smoker | 80 | Death w/o cancer | 0.037 | (0.027, 0.052) |
| Women | Abdominal bloating | Ever smoker | 90 | Breast | 0.008 | (0.003, 0.021) |
| Women | Abdominal bloating | Ever smoker | 90 | Gyanecological | 0.011 | (0.006, 0.021) |
| Women | Abdominal bloating | Ever smoker | 90 | Lung | 0.003 | (0.001, 0.010) |
| Women | Abdominal bloating | Ever smoker | 90 | Upper GI | 0.010 | (0.005, 0.019) |
| Women | Abdominal bloating | Ever smoker | 90 | Lower GI | 0.012 | (0.007, 0.020) |
| Women | Abdominal bloating | Ever smoker | 90 | Urological | 0.003 | (0.001, 0.008) |
| Women | Abdominal bloating | Ever smoker | 90 | Haematological | 0.003 | (0.001, 0.006) |
| Women | Abdominal bloating | Ever smoker | 90 | Other cancers | 0.009 | (0.005, 0.018) |
| Women | Abdominal bloating | Ever smoker | 90 | Death w/o cancer | 0.115 | (0.091, 0.145) |
| Women | Breast lump | Never smoker | 30 | Breast | 0.011 | (0.006, 0.021) |
| Women | Breast lump | Never smoker | 30 | Gyanecological | 0.003 | (0.001, 0.007) |
| Women | Breast lump | Never smoker | 40 | Breast | 0.062 | (0.046, 0.083) |
| Women | Breast lump | Never smoker | 40 | Gyanecological | 0.001 | (<0.001, 0.003) |
| Women | Breast lump | Never smoker | 50 | Breast | 0.132 | (0.110, 0.156) |
| Women | Breast lump | Never smoker | 50 | Haematological | 0.001 | (<0.001, 0.002) |
| Women | Breast lump | Never smoker | 50 | Other cancers | 0.001 | (<0.001, 0.002) |
| Women | Breast lump | Never smoker | 50 | Death w/o cancer | 0.001 | (<0.001, 0.003) |
| Women | Breast lump | Never smoker | 60 | Breast | 0.230 | (0.191, 0.274) |
| Women | Breast lump | Never smoker | 60 | Gyanecological | 0.001 | (<0.001, 0.003) |
| Women | Breast lump | Never smoker | 60 | Haematological | 0.002 | (<0.001, 0.004) |
| Women | Breast lump | Never smoker | 60 | Other cancers | 0.001 | (<0.001, 0.003) |
| Women | Breast lump | Never smoker | 60 | Death w/o cancer | 0.002 | (<0.001, 0.005) |
| Women | Breast lump | Never smoker | 70 | Breast | 0.340 | (0.302, 0.380) |
| Women | Breast lump | Never smoker | 70 | Gyanecological | 0.002 | (<0.001, 0.004) |
| Women | Breast lump | Never smoker | 70 | Haematological | 0.003 | (0.001, 0.007) |
| Women | Breast lump | Never smoker | 70 | Other cancers | 0.002 | (<0.001, 0.006) |
| Women | Breast lump | Never smoker | 70 | Death w/o cancer | 0.005 | (0.002, 0.013) |
| Women | Breast lump | Never smoker | 80 | Breast | 0.417 | (0.380, 0.455) |
| Women | Breast lump | Never smoker | 80 | Gyanecological | 0.002 | (<0.001, 0.003) |
| Women | Breast lump | Never smoker | 80 | Upper GI | 0.001 | (<0.001, 0.002) |
| Women | Breast lump | Never smoker | 80 | Lower GI | 0.001 | (<0.001, 0.002) |
| Women | Breast lump | Never smoker | 80 | Haematological | 0.004 | (0.001, 0.011) |
| Women | Breast lump | Never smoker | 80 | Other cancers | 0.003 | (0.001, 0.007) |
| Women | Breast lump | Never smoker | 80 | Death w/o cancer | 0.016 | (0.009, 0.028) |
| Women | Breast lump | Never smoker | 90 | Breast | 0.474 | (0.427, 0.521) |
| Women | Breast lump | Never smoker | 90 | Gyanecological | 0.001 | (<0.001, 0.002) |
| Women | Breast lump | Never smoker | 90 | Upper GI | 0.001 | (<0.001, 0.002) |
| Women | Breast lump | Never smoker | 90 | Lower GI | 0.001 | (<0.001, 0.002) |
| Women | Breast lump | Never smoker | 90 | Haematological | 0.004 | (0.002, 0.009) |
| Women | Breast lump | Never smoker | 90 | Other cancers | 0.003 | (0.001, 0.009) |
| Women | Breast lump | Never smoker | 90 | Death w/o cancer | 0.047 | (0.033, 0.068) |
| Women | Breast lump | Ever smoker | 30 | Breast | 0.012 | (0.006, 0.026) |
| Women | Breast lump | Ever smoker | 30 | Gyanecological | 0.003 | (0.001, 0.008) |
| Women | Breast lump | Ever smoker | 40 | Breast | 0.065 | (0.051, 0.082) |
| Women | Breast lump | Ever smoker | 40 | Gyanecological | 0.002 | (<0.001, 0.003) |
| Women | Breast lump | Ever smoker | 50 | Breast | 0.142 | (0.114, 0.175) |
| Women | Breast lump | Ever smoker | 50 | Other cancers | 0.001 | (<0.001, 0.002) |
| Women | Breast lump | Ever smoker | 50 | Death w/o cancer | 0.001 | (<0.001, 0.002) |
| Women | Breast lump | Ever smoker | 60 | Breast | 0.241 | (0.208, 0.277) |
| Women | Breast lump | Ever smoker | 60 | Gyanecological | 0.001 | (<0.001, 0.003) |
| Women | Breast lump | Ever smoker | 60 | Lung | 0.002 | (<0.001, 0.003) |
| Women | Breast lump | Ever smoker | 60 | Haematological | 0.002 | (<0.001, 0.004) |
| Women | Breast lump | Ever smoker | 60 | Other cancers | 0.002 | (<0.001, 0.003) |
| Women | Breast lump | Ever smoker | 60 | Death w/o cancer | 0.002 | (<0.001, 0.003) |
| Women | Breast lump | Ever smoker | 70 | Breast | 0.355 | (0.323, 0.387) |
| Women | Breast lump | Ever smoker | 70 | Gyanecological | 0.002 | (<0.001, 0.004) |
| Women | Breast lump | Ever smoker | 70 | Lung | 0.003 | (0.001, 0.008) |
| Women | Breast lump | Ever smoker | 70 | Upper GI | 0.001 | (<0.001, 0.002) |
| Women | Breast lump | Ever smoker | 70 | Haematological | 0.003 | (0.001, 0.007) |
| Women | Breast lump | Ever smoker | 70 | Other cancers | 0.003 | (0.001, 0.005) |
| Women | Breast lump | Ever smoker | 70 | Death w/o cancer | 0.005 | (0.002, 0.012) |
| Women | Breast lump | Ever smoker | 80 | Breast | 0.433 | (0.385, 0.482) |
| Women | Breast lump | Ever smoker | 80 | Gyanecological | 0.002 | (<0.001, 0.004) |
| Women | Breast lump | Ever smoker | 80 | Lung | 0.003 | (0.001, 0.008) |
| Women | Breast lump | Ever smoker | 80 | Upper GI | 0.001 | (<0.001, 0.002) |
| Women | Breast lump | Ever smoker | 80 | Lower GI | 0.001 | (<0.001, 0.002) |
| Women | Breast lump | Ever smoker | 80 | Haematological | 0.004 | (0.001, 0.011) |
| Women | Breast lump | Ever smoker | 80 | Other cancers | 0.003 | (0.001, 0.008) |
| Women | Breast lump | Ever smoker | 80 | Death w/o cancer | 0.015 | (0.010, 0.024) |
| Women | Breast lump | Ever smoker | 90 | Breast | 0.496 | (0.460, 0.533) |
| Women | Breast lump | Ever smoker | 90 | Gyanecological | 0.001 | (<0.001, 0.002) |
| Women | Breast lump | Ever smoker | 90 | Lung | 0.002 | (<0.001, 0.007) |
| Women | Breast lump | Ever smoker | 90 | Upper GI | 0.001 | (<0.001, 0.002) |
| Women | Breast lump | Ever smoker | 90 | Haematological | 0.004 | (0.002, 0.006) |
| Women | Breast lump | Ever smoker | 90 | Other cancers | 0.003 | (0.001, 0.008) |
| Women | Breast lump | Ever smoker | 90 | Death w/o cancer | 0.044 | (0.032, 0.061) |
| Women | Change in bowel habit | Never smoker | 30 | Gyanecological | 0.009 | (0.003, 0.022) |
| Women | Change in bowel habit | Never smoker | 40 | Gyanecological | 0.004 | (0.001, 0.012) |
| Women | Change in bowel habit | Never smoker | 40 | Lower GI | 0.002 | (<0.001, 0.003) |
| Women | Change in bowel habit | Never smoker | 50 | Breast | 0.002 | (0.001, 0.005) |
| Women | Change in bowel habit | Never smoker | 50 | Gyanecological | 0.003 | (0.001, 0.005) |
| Women | Change in bowel habit | Never smoker | 50 | Lower GI | 0.007 | (0.003, 0.015) |
| Women | Change in bowel habit | Never smoker | 50 | Other cancers | 0.002 | (0.001, 0.006) |
| Women | Change in bowel habit | Never smoker | 50 | Death w/o cancer | 0.001 | (<0.001, 0.002) |
| Women | Change in bowel habit | Never smoker | 60 | Breast | 0.004 | (0.002, 0.010) |
| Women | Change in bowel habit | Never smoker | 60 | Gyanecological | 0.005 | (0.002, 0.013) |
| Women | Change in bowel habit | Never smoker | 60 | Upper GI | 0.001 | (<0.001, 0.003) |
| Women | Change in bowel habit | Never smoker | 60 | Lower GI | 0.016 | (0.010, 0.025) |
| Women | Change in bowel habit | Never smoker | 60 | Other cancers | 0.003 | (0.002, 0.006) |
| Women | Change in bowel habit | Never smoker | 60 | Death w/o cancer | 0.002 | (<0.001, 0.004) |
| Women | Change in bowel habit | Never smoker | 70 | Breast | 0.007 | (0.002, 0.018) |
| Women | Change in bowel habit | Never smoker | 70 | Gyanecological | 0.008 | (0.004, 0.014) |
| Women | Change in bowel habit | Never smoker | 70 | Lung | 0.001 | (<0.001, 0.002) |
| Women | Change in bowel habit | Never smoker | 70 | Upper GI | 0.003 | (0.001, 0.009) |
| Women | Change in bowel habit | Never smoker | 70 | Lower GI | 0.037 | (0.027, 0.051) |
| Women | Change in bowel habit | Never smoker | 70 | Urological | 0.002 | (<0.001, 0.003) |
| Women | Change in bowel habit | Never smoker | 70 | Haematological | 0.001 | (<0.001, 0.003) |
| Women | Change in bowel habit | Never smoker | 70 | Other cancers | 0.006 | (0.003, 0.014) |
| Women | Change in bowel habit | Never smoker | 70 | Death w/o cancer | 0.006 | (0.002, 0.016) |
| Women | Change in bowel habit | Never smoker | 80 | Breast | 0.009 | (0.004, 0.017) |
| Women | Change in bowel habit | Never smoker | 80 | Gyanecological | 0.007 | (0.004, 0.013) |
| Women | Change in bowel habit | Never smoker | 80 | Upper GI | 0.005 | (0.002, 0.010) |
| Women | Change in bowel habit | Never smoker | 80 | Lower GI | 0.049 | (0.036, 0.066) |
| Women | Change in bowel habit | Never smoker | 80 | Urological | 0.002 | (<0.001, 0.004) |
| Women | Change in bowel habit | Never smoker | 80 | Haematological | 0.002 | (<0.001, 0.006) |
| Women | Change in bowel habit | Never smoker | 80 | Other cancers | 0.009 | (0.004, 0.020) |
| Women | Change in bowel habit | Never smoker | 80 | Death w/o cancer | 0.021 | (0.015, 0.031) |
| Women | Change in bowel habit | Never smoker | 90 | Breast | 0.010 | (0.006, 0.016) |
| Women | Change in bowel habit | Never smoker | 90 | Gyanecological | 0.005 | (0.003, 0.009) |
| Women | Change in bowel habit | Never smoker | 90 | Lung | 0.001 | (<0.001, 0.002) |
| Women | Change in bowel habit | Never smoker | 90 | Upper GI | 0.005 | (0.002, 0.012) |
| Women | Change in bowel habit | Never smoker | 90 | Lower GI | 0.055 | (0.036, 0.082) |
| Women | Change in bowel habit | Never smoker | 90 | Urological | 0.002 | (0.001, 0.004) |
| Women | Change in bowel habit | Never smoker | 90 | Haematological | 0.002 | (<0.001, 0.004) |
| Women | Change in bowel habit | Never smoker | 90 | Other cancers | 0.011 | (0.006, 0.022) |
| Women | Change in bowel habit | Never smoker | 90 | Death w/o cancer | 0.065 | (0.047, 0.090) |
| Women | Change in bowel habit | Ever smoker | 30 | Gyanecological | 0.009 | (0.004, 0.021) |
| Women | Change in bowel habit | Ever smoker | 40 | Breast | 0.001 | (<0.001, 0.002) |
| Women | Change in bowel habit | Ever smoker | 40 | Gyanecological | 0.004 | (0.002, 0.011) |
| Women | Change in bowel habit | Ever smoker | 40 | Lower GI | 0.002 | (<0.001, 0.004) |
| Women | Change in bowel habit | Ever smoker | 40 | Other cancers | 0.001 | (<0.001, 0.002) |
| Women | Change in bowel habit | Ever smoker | 50 | Breast | 0.002 | (0.001, 0.004) |
| Women | Change in bowel habit | Ever smoker | 50 | Gyanecological | 0.003 | (0.001, 0.006) |
| Women | Change in bowel habit | Ever smoker | 50 | Lower GI | 0.007 | (0.003, 0.015) |
| Women | Change in bowel habit | Ever smoker | 50 | Other cancers | 0.002 | (0.001, 0.006) |
| Women | Change in bowel habit | Ever smoker | 60 | Breast | 0.005 | (0.003, 0.008) |
| Women | Change in bowel habit | Ever smoker | 60 | Gyanecological | 0.005 | (0.002, 0.013) |
| Women | Change in bowel habit | Ever smoker | 60 | Lung | 0.002 | (0.001, 0.006) |
| Women | Change in bowel habit | Ever smoker | 60 | Upper GI | 0.002 | (<0.001, 0.004) |
| Women | Change in bowel habit | Ever smoker | 60 | Lower GI | 0.016 | (0.010, 0.028) |
| Women | Change in bowel habit | Ever smoker | 60 | Other cancers | 0.004 | (0.002, 0.008) |
| Women | Change in bowel habit | Ever smoker | 60 | Death w/o cancer | 0.002 | (<0.001, 0.004) |
| Women | Change in bowel habit | Ever smoker | 70 | Breast | 0.007 | (<0.001, 0.000) |
| Women | Change in bowel habit | Ever smoker | 70 | Gyanecological | 0.009 | (0.005, 0.014) |
| Women | Change in bowel habit | Ever smoker | 70 | Lung | 0.005 | (0.002, 0.013) |
| Women | Change in bowel habit | Ever smoker | 70 | Upper GI | 0.004 | (0.002, 0.011) |
| Women | Change in bowel habit | Ever smoker | 70 | Lower GI | 0.034 | (0.024, 0.047) |
| Women | Change in bowel habit | Ever smoker | 70 | Urological | 0.002 | (<0.001, 0.004) |
| Women | Change in bowel habit | Ever smoker | 70 | Haematological | 0.002 | (<0.001, 0.004) |
| Women | Change in bowel habit | Ever smoker | 70 | Other cancers | 0.006 | (0.002, 0.021) |
| Women | Change in bowel habit | Ever smoker | 70 | Death w/o cancer | 0.006 | (0.003, 0.014) |
| Women | Change in bowel habit | Ever smoker | 80 | Breast | 0.008 | (0.004, 0.016) |
| Women | Change in bowel habit | Ever smoker | 80 | Gyanecological | 0.008 | (0.004, 0.016) |
| Women | Change in bowel habit | Ever smoker | 80 | Lung | 0.005 | (0.002, 0.011) |
| Women | Change in bowel habit | Ever smoker | 80 | Upper GI | 0.006 | (0.002, 0.015) |
| Women | Change in bowel habit | Ever smoker | 80 | Lower GI | 0.049 | (0.036, 0.065) |
| Women | Change in bowel habit | Ever smoker | 80 | Urological | 0.003 | (0.001, 0.006) |
| Women | Change in bowel habit | Ever smoker | 80 | Haematological | 0.002 | (<0.001, 0.004) |
| Women | Change in bowel habit | Ever smoker | 80 | Other cancers | 0.009 | (0.003, 0.025) |
| Women | Change in bowel habit | Ever smoker | 80 | Death w/o cancer | 0.020 | (0.013, 0.032) |
| Women | Change in bowel habit | Ever smoker | 90 | Breast | 0.011 | (0.005, 0.021) |
| Women | Change in bowel habit | Ever smoker | 90 | Gyanecological | 0.005 | (0.003, 0.011) |
| Women | Change in bowel habit | Ever smoker | 90 | Lung | 0.005 | (0.002, 0.014) |
| Women | Change in bowel habit | Ever smoker | 90 | Upper GI | 0.006 | (0.003, 0.012) |
| Women | Change in bowel habit | Ever smoker | 90 | Lower GI | 0.051 | (0.036, 0.070) |
| Women | Change in bowel habit | Ever smoker | 90 | Urological | 0.003 | (0.001, 0.008) |
| Women | Change in bowel habit | Ever smoker | 90 | Haematological | 0.002 | (<0.001, 0.005) |
| Women | Change in bowel habit | Ever smoker | 90 | Other cancers | 0.011 | (0.005, 0.024) |
| Women | Change in bowel habit | Ever smoker | 90 | Death w/o cancer | 0.064 | (0.045, 0.091) |
| Women | Dyspepsia | Never smoker | 30 | Gyanecological | 0.003 | (0.002, 0.005) |
| Women | Dyspepsia | Never smoker | 40 | Gyanecological | 0.001 | (<0.001, 0.002) |
| Women | Dyspepsia | Never smoker | 50 | Breast | 0.002 | (<0.001, 0.005) |
| Women | Dyspepsia | Never smoker | 50 | Upper GI | 0.001 | (<0.001, 0.002) |
| Women | Dyspepsia | Never smoker | 50 | Other cancers | 0.002 | (<0.001, 0.003) |
| Women | Dyspepsia | Never smoker | 50 | Death w/o cancer | 0.001 | (<0.001, 0.003) |
| Women | Dyspepsia | Never smoker | 60 | Breast | 0.004 | (0.002, 0.009) |
| Women | Dyspepsia | Never smoker | 60 | Gyanecological | 0.002 | (<0.001, 0.004) |
| Women | Dyspepsia | Never smoker | 60 | Upper GI | 0.003 | (0.001, 0.006) |
| Women | Dyspepsia | Never smoker | 60 | Lower GI | 0.001 | (<0.001, 0.003) |
| Women | Dyspepsia | Never smoker | 60 | Haematological | 0.001 | (<0.001, 0.002) |
| Women | Dyspepsia | Never smoker | 60 | Other cancers | 0.002 | (<0.001, 0.006) |
| Women | Dyspepsia | Never smoker | 60 | Death w/o cancer | 0.002 | (0.001, 0.004) |
| Women | Dyspepsia | Never smoker | 70 | Breast | 0.006 | (0.002, 0.015) |
| Women | Dyspepsia | Never smoker | 70 | Gyanecological | 0.003 | (0.001, 0.006) |
| Women | Dyspepsia | Never smoker | 70 | Upper GI | 0.006 | (0.003, 0.014) |
| Women | Dyspepsia | Never smoker | 70 | Lower GI | 0.004 | (0.002, 0.008) |
| Women | Dyspepsia | Never smoker | 70 | Haematological | 0.003 | (0.001, 0.005) |
| Women | Dyspepsia | Never smoker | 70 | Other cancers | 0.004 | (0.001, 0.010) |
| Women | Dyspepsia | Never smoker | 70 | Death w/o cancer | 0.008 | (0.004, 0.017) |
| Women | Dyspepsia | Never smoker | 80 | Breast | 0.008 | (0.004, 0.015) |
| Women | Dyspepsia | Never smoker | 80 | Gyanecological | 0.003 | (0.001, 0.007) |
| Women | Dyspepsia | Never smoker | 80 | Upper GI | 0.010 | (0.003, 0.027) |
| Women | Dyspepsia | Never smoker | 80 | Lower GI | 0.004 | (0.002, 0.009) |
| Women | Dyspepsia | Never smoker | 80 | Haematological | 0.003 | (0.001, 0.007) |
| Women | Dyspepsia | Never smoker | 80 | Other cancers | 0.006 | (0.003, 0.012) |
| Women | Dyspepsia | Never smoker | 80 | Death w/o cancer | 0.027 | (0.020, 0.037) |
| Women | Dyspepsia | Never smoker | 90 | Breast | 0.009 | (0.004, 0.019) |
| Women | Dyspepsia | Never smoker | 90 | Gyanecological | 0.002 | (<0.001, 0.006) |
| Women | Dyspepsia | Never smoker | 90 | Upper GI | 0.009 | (0.005, 0.019) |
| Women | Dyspepsia | Never smoker | 90 | Lower GI | 0.004 | (0.002, 0.009) |
| Women | Dyspepsia | Never smoker | 90 | Haematological | 0.003 | (0.001, 0.009) |
| Women | Dyspepsia | Never smoker | 90 | Other cancers | 0.006 | (0.003, 0.011) |
| Women | Dyspepsia | Never smoker | 90 | Death w/o cancer | 0.088 | (0.068, 0.114) |
| Women | Dyspepsia | Ever smoker | 30 | Gyanecological | 0.004 | (0.001, 0.009) |
| Women | Dyspepsia | Ever smoker | 40 | Breast | 0.001 | (<0.001, 0.002) |
| Women | Dyspepsia | Ever smoker | 40 | Gyanecological | 0.001 | (<0.001, 0.003) |
| Women | Dyspepsia | Ever smoker | 50 | Breast | 0.002 | (<0.001, 0.005) |
| Women | Dyspepsia | Ever smoker | 50 | Gyanecological | 0.001 | (<0.001, 0.002) |
| Women | Dyspepsia | Ever smoker | 50 | Upper GI | 0.001 | (<0.001, 0.003) |
| Women | Dyspepsia | Ever smoker | 50 | Other cancers | 0.002 | (<0.001, 0.003) |
| Women | Dyspepsia | Ever smoker | 50 | Death w/o cancer | 0.001 | (<0.001, 0.002) |
| Women | Dyspepsia | Ever smoker | 60 | Breast | 0.004 | (0.002, 0.008) |
| Women | Dyspepsia | Ever smoker | 60 | Gyanecological | 0.002 | (0.001, 0.003) |
| Women | Dyspepsia | Ever smoker | 60 | Lung | 0.002 | (<0.001, 0.004) |
| Women | Dyspepsia | Ever smoker | 60 | Upper GI | 0.003 | (0.001, 0.009) |
| Women | Dyspepsia | Ever smoker | 60 | Lower GI | 0.001 | (<0.001, 0.003) |
| Women | Dyspepsia | Ever smoker | 60 | Haematological | 0.001 | (<0.001, 0.002) |
| Women | Dyspepsia | Ever smoker | 60 | Other cancers | 0.002 | (<0.001, 0.005) |
| Women | Dyspepsia | Ever smoker | 60 | Death w/o cancer | 0.002 | (<0.001, 0.005) |
| Women | Dyspepsia | Ever smoker | 70 | Breast | 0.006 | (0.003, 0.015) |
| Women | Dyspepsia | Ever smoker | 70 | Gyanecological | 0.003 | (0.001, 0.008) |
| Women | Dyspepsia | Ever smoker | 70 | Lung | 0.004 | (0.001, 0.011) |
| Women | Dyspepsia | Ever smoker | 70 | Upper GI | 0.008 | (0.004, 0.016) |
| Women | Dyspepsia | Ever smoker | 70 | Lower GI | 0.003 | (0.001, 0.007) |
| Women | Dyspepsia | Ever smoker | 70 | Haematological | 0.002 | (0.001, 0.005) |
| Women | Dyspepsia | Ever smoker | 70 | Other cancers | 0.004 | (0.001, 0.011) |
| Women | Dyspepsia | Ever smoker | 70 | Death w/o cancer | 0.008 | (0.004, 0.016) |
| Women | Dyspepsia | Ever smoker | 80 | Breast | 0.008 | (0.003, 0.018) |
| Women | Dyspepsia | Ever smoker | 80 | Gyanecological | 0.003 | (0.001, 0.008) |
| Women | Dyspepsia | Ever smoker | 80 | Lung | 0.004 | (0.001, 0.012) |
| Women | Dyspepsia | Ever smoker | 80 | Upper GI | 0.011 | (0.006, 0.020) |
| Women | Dyspepsia | Ever smoker | 80 | Lower GI | 0.004 | (0.002, 0.009) |
| Women | Dyspepsia | Ever smoker | 80 | Urological | 0.001 | (<0.001, 0.002) |
| Women | Dyspepsia | Ever smoker | 80 | Haematological | 0.003 | (0.001, 0.006) |
| Women | Dyspepsia | Ever smoker | 80 | Other cancers | 0.005 | (0.002, 0.015) |
| Women | Dyspepsia | Ever smoker | 80 | Death w/o cancer | 0.027 | (0.018, 0.038) |
| Women | Dyspepsia | Ever smoker | 90 | Breast | 0.010 | (0.005, 0.018) |
| Women | Dyspepsia | Ever smoker | 90 | Gyanecological | 0.003 | (0.001, 0.005) |
| Women | Dyspepsia | Ever smoker | 90 | Lung | 0.004 | (0.001, 0.009) |
| Women | Dyspepsia | Ever smoker | 90 | Upper GI | 0.012 | (0.007, 0.020) |
| Women | Dyspepsia | Ever smoker | 90 | Lower GI | 0.004 | (0.002, 0.009) |
| Women | Dyspepsia | Ever smoker | 90 | Urological | 0.001 | (<0.001, 0.002) |
| Women | Dyspepsia | Ever smoker | 90 | Haematological | 0.003 | (0.001, 0.008) |
| Women | Dyspepsia | Ever smoker | 90 | Other cancers | 0.007 | (0.002, 0.020) |
| Women | Dyspepsia | Ever smoker | 90 | Death w/o cancer | 0.086 | (0.069, 0.106) |
| Women | Dysphagia | Never smoker | 30 | Gyanecological | 0.002 | (0.001, 0.005) |
| Women | Dysphagia | Never smoker | 30 | Death w/o cancer | 0.001 | (<0.001, 0.002) |
| Women | Dysphagia | Never smoker | 40 | Gyanecological | 0.001 | (<0.001, 0.002) |
| Women | Dysphagia | Never smoker | 40 | Upper GI | 0.002 | (0.001, 0.003) |
| Women | Dysphagia | Never smoker | 40 | Other cancers | 0.001 | (<0.001, 0.002) |
| Women | Dysphagia | Never smoker | 40 | Death w/o cancer | 0.002 | (<0.001, 0.005) |
| Women | Dysphagia | Never smoker | 50 | Breast | 0.001 | (<0.001, 0.002) |
| Women | Dysphagia | Never smoker | 50 | Upper GI | 0.006 | (0.003, 0.013) |
| Women | Dysphagia | Never smoker | 50 | Other cancers | 0.003 | (<0.001, 0.007) |
| Women | Dysphagia | Never smoker | 50 | Death w/o cancer | 0.005 | (0.002, 0.012) |
| Women | Dysphagia | Never smoker | 60 | Breast | 0.003 | (0.001, 0.005) |
| Women | Dysphagia | Never smoker | 60 | Gyanecological | 0.001 | (<0.001, 0.003) |
| Women | Dysphagia | Never smoker | 60 | Lung | 0.001 | (<0.001, 0.002) |
| Women | Dysphagia | Never smoker | 60 | Upper GI | 0.014 | (0.008, 0.025) |
| Women | Dysphagia | Never smoker | 60 | Lower GI | 0.002 | (<0.001, 0.003) |
| Women | Dysphagia | Never smoker | 60 | Haematological | 0.001 | (<0.001, 0.003) |
| Women | Dysphagia | Never smoker | 60 | Other cancers | 0.004 | (0.002, 0.008) |
| Women | Dysphagia | Never smoker | 60 | Death w/o cancer | 0.009 | (0.005, 0.018) |
| Women | Dysphagia | Never smoker | 70 | Breast | 0.004 | (0.002, 0.009) |
| Women | Dysphagia | Never smoker | 70 | Gyanecological | 0.002 | (0.001, 0.005) |
| Women | Dysphagia | Never smoker | 70 | Lung | 0.002 | (<0.001, 0.006) |
| Women | Dysphagia | Never smoker | 70 | Upper GI | 0.035 | (0.024, 0.051) |
| Women | Dysphagia | Never smoker | 70 | Lower GI | 0.003 | (0.001, 0.007) |
| Women | Dysphagia | Never smoker | 70 | Urological | 0.002 | (<0.001, 0.003) |
| Women | Dysphagia | Never smoker | 70 | Haematological | 0.002 | (<0.001, 0.005) |
| Women | Dysphagia | Never smoker | 70 | Other cancers | 0.007 | (0.003, 0.017) |
| Women | Dysphagia | Never smoker | 70 | Death w/o cancer | 0.034 | (0.025, 0.046) |
| Women | Dysphagia | Never smoker | 80 | Breast | 0.005 | (0.003, 0.008) |
| Women | Dysphagia | Never smoker | 80 | Gyanecological | 0.002 | (<0.001, 0.005) |
| Women | Dysphagia | Never smoker | 80 | Lung | 0.003 | (<0.001, 0.006) |
| Women | Dysphagia | Never smoker | 80 | Upper GI | 0.050 | (0.034, 0.073) |
| Women | Dysphagia | Never smoker | 80 | Lower GI | 0.005 | (0.002, 0.009) |
| Women | Dysphagia | Never smoker | 80 | Urological | 0.002 | (<0.001, 0.004) |
| Women | Dysphagia | Never smoker | 80 | Haematological | 0.003 | (0.001, 0.006) |
| Women | Dysphagia | Never smoker | 80 | Other cancers | 0.009 | (0.004, 0.019) |
| Women | Dysphagia | Never smoker | 80 | Death w/o cancer | 0.108 | (0.088, 0.132) |
| Women | Dysphagia | Never smoker | 90 | Breast | 0.005 | (0.002, 0.011) |
| Women | Dysphagia | Never smoker | 90 | Gyanecological | 0.001 | (<0.001, 0.003) |
| Women | Dysphagia | Never smoker | 90 | Lung | 0.002 | (<0.001, 0.006) |
| Women | Dysphagia | Never smoker | 90 | Upper GI | 0.049 | (0.037, 0.065) |
| Women | Dysphagia | Never smoker | 90 | Lower GI | 0.004 | (0.001, 0.011) |
| Women | Dysphagia | Never smoker | 90 | Urological | 0.002 | (0.001, 0.004) |
| Women | Dysphagia | Never smoker | 90 | Haematological | 0.003 | (0.001, 0.006) |
| Women | Dysphagia | Never smoker | 90 | Other cancers | 0.010 | (0.005, 0.021) |
| Women | Dysphagia | Never smoker | 90 | Death w/o cancer | 0.310 | (0.282, 0.339) |
| Women | Dysphagia | Ever smoker | 30 | Gyanecological | 0.003 | (<0.001, 0.007) |
| Women | Dysphagia | Ever smoker | 30 | Death w/o cancer | 0.001 | (<0.001, 0.002) |
| Women | Dysphagia | Ever smoker | 40 | Upper GI | 0.002 | (<0.001, 0.004) |
| Women | Dysphagia | Ever smoker | 40 | Other cancers | 0.001 | (<0.001, 0.002) |
| Women | Dysphagia | Ever smoker | 40 | Death w/o cancer | 0.002 | (<0.001, 0.005) |
| Women | Dysphagia | Ever smoker | 50 | Breast | 0.002 | (<0.001, 0.003) |
| Women | Dysphagia | Ever smoker | 50 | Lung | 0.002 | (<0.001, 0.004) |
| Women | Dysphagia | Ever smoker | 50 | Upper GI | 0.008 | (0.004, 0.017) |
| Women | Dysphagia | Ever smoker | 50 | Other cancers | 0.003 | (0.001, 0.007) |
| Women | Dysphagia | Ever smoker | 50 | Death w/o cancer | 0.005 | (0.003, 0.008) |
| Women | Dysphagia | Ever smoker | 60 | Breast | 0.003 | (<0.001, 0.007) |
| Women | Dysphagia | Ever smoker | 60 | Gyanecological | 0.001 | (<0.001, 0.002) |
| Women | Dysphagia | Ever smoker | 60 | Lung | 0.006 | (0.002, 0.016) |
| Women | Dysphagia | Ever smoker | 60 | Upper GI | 0.017 | (0.010, 0.028) |
| Women | Dysphagia | Ever smoker | 60 | Lower GI | 0.001 | (<0.001, 0.003) |
| Women | Dysphagia | Ever smoker | 60 | Haematological | 0.001 | (<0.001, 0.003) |
| Women | Dysphagia | Ever smoker | 60 | Other cancers | 0.004 | (0.002, 0.011) |
| Women | Dysphagia | Ever smoker | 60 | Death w/o cancer | 0.010 | (0.006, 0.016) |
| Women | Dysphagia | Ever smoker | 70 | Breast | 0.004 | (0.002, 0.010) |
| Women | Dysphagia | Ever smoker | 70 | Gyanecological | 0.002 | (<0.001, 0.005) |
| Women | Dysphagia | Ever smoker | 70 | Lung | 0.011 | (0.006, 0.019) |
| Women | Dysphagia | Ever smoker | 70 | Upper GI | 0.044 | (0.030, 0.063) |
| Women | Dysphagia | Ever smoker | 70 | Lower GI | 0.003 | (0.001, 0.008) |
| Women | Dysphagia | Ever smoker | 70 | Urological | 0.002 | (0.001, 0.004) |
| Women | Dysphagia | Ever smoker | 70 | Haematological | 0.002 | (0.001, 0.004) |
| Women | Dysphagia | Ever smoker | 70 | Other cancers | 0.007 | (0.003, 0.015) |
| Women | Dysphagia | Ever smoker | 70 | Death w/o cancer | 0.031 | (0.023, 0.042) |
| Women | Dysphagia | Ever smoker | 80 | Breast | 0.005 | (0.002, 0.012) |
| Women | Dysphagia | Ever smoker | 80 | Gyanecological | 0.002 | (0.001, 0.005) |
| Women | Dysphagia | Ever smoker | 80 | Lung | 0.013 | (0.007, 0.024) |
| Women | Dysphagia | Ever smoker | 80 | Upper GI | 0.060 | (0.045, 0.080) |
| Women | Dysphagia | Ever smoker | 80 | Lower GI | 0.004 | (0.002, 0.008) |
| Women | Dysphagia | Ever smoker | 80 | Urological | 0.003 | (0.001, 0.005) |
| Women | Dysphagia | Ever smoker | 80 | Haematological | 0.003 | (0.001, 0.006) |
| Women | Dysphagia | Ever smoker | 80 | Other cancers | 0.010 | (0.005, 0.019) |
| Women | Dysphagia | Ever smoker | 80 | Death w/o cancer | 0.104 | (0.086, 0.125) |
| Women | Dysphagia | Ever smoker | 90 | Breast | 0.005 | (0.002, 0.015) |
| Women | Dysphagia | Ever smoker | 90 | Gyanecological | 0.001 | (<0.001, 0.002) |
| Women | Dysphagia | Ever smoker | 90 | Lung | 0.010 | (0.005, 0.019) |
| Women | Dysphagia | Ever smoker | 90 | Upper GI | 0.060 | (0.045, 0.079) |
| Women | Dysphagia | Ever smoker | 90 | Lower GI | 0.004 | (0.002, 0.009) |
| Women | Dysphagia | Ever smoker | 90 | Urological | 0.003 | (0.001, 0.006) |
| Women | Dysphagia | Ever smoker | 90 | Haematological | 0.002 | (0.001, 0.005) |
| Women | Dysphagia | Ever smoker | 90 | Other cancers | 0.010 | (0.005, 0.020) |
| Women | Dysphagia | Ever smoker | 90 | Death w/o cancer | 0.297 | (0.262, 0.335) |
| Women | Dyspnoea | Never smoker | 30 | Gyanecological | 0.003 | (0.001, 0.008) |
| Women | Dyspnoea | Never smoker | 40 | Gyanecological | 0.001 | (<0.001, 0.002) |
| Women | Dyspnoea | Never smoker | 40 | Death w/o cancer | 0.001 | (<0.001, 0.002) |
| Women | Dyspnoea | Never smoker | 50 | Breast | 0.002 | (<0.001, 0.003) |
| Women | Dyspnoea | Never smoker | 50 | Other cancers | 0.001 | (<0.001, 0.002) |
| Women | Dyspnoea | Never smoker | 50 | Death w/o cancer | 0.003 | (0.002, 0.006) |
| Women | Dyspnoea | Never smoker | 60 | Breast | 0.003 | (0.001, 0.007) |
| Women | Dyspnoea | Never smoker | 60 | Gyanecological | 0.002 | (<0.001, 0.004) |
| Women | Dyspnoea | Never smoker | 60 | Lung | 0.001 | (<0.001, 0.004) |
| Women | Dyspnoea | Never smoker | 60 | Lower GI | 0.002 | (<0.001, 0.003) |
| Women | Dyspnoea | Never smoker | 60 | Haematological | 0.001 | (<0.001, 0.003) |
| Women | Dyspnoea | Never smoker | 60 | Other cancers | 0.002 | (<0.001, 0.004) |
| Women | Dyspnoea | Never smoker | 60 | Death w/o cancer | 0.006 | (0.003, 0.012) |
| Women | Dyspnoea | Never smoker | 70 | Breast | 0.005 | (0.002, 0.013) |
| Women | Dyspnoea | Never smoker | 70 | Gyanecological | 0.003 | (0.001, 0.009) |
| Women | Dyspnoea | Never smoker | 70 | Lung | 0.003 | (0.001, 0.005) |
| Women | Dyspnoea | Never smoker | 70 | Upper GI | 0.002 | (<0.001, 0.003) |
| Women | Dyspnoea | Never smoker | 70 | Lower GI | 0.003 | (0.001, 0.007) |
| Women | Dyspnoea | Never smoker | 70 | Urological | 0.001 | (<0.001, 0.003) |
| Women | Dyspnoea | Never smoker | 70 | Haematological | 0.003 | (0.001, 0.006) |
| Women | Dyspnoea | Never smoker | 70 | Other cancers | 0.003 | (0.002, 0.006) |
| Women | Dyspnoea | Never smoker | 70 | Death w/o cancer | 0.021 | (0.013, 0.033) |
| Women | Dyspnoea | Never smoker | 80 | Breast | 0.007 | (0.003, 0.017) |
| Women | Dyspnoea | Never smoker | 80 | Gyanecological | 0.003 | (<0.001, 0.006) |
| Women | Dyspnoea | Never smoker | 80 | Lung | 0.003 | (0.001, 0.009) |
| Women | Dyspnoea | Never smoker | 80 | Upper GI | 0.002 | (0.001, 0.004) |
| Women | Dyspnoea | Never smoker | 80 | Lower GI | 0.004 | (0.002, 0.012) |
| Women | Dyspnoea | Never smoker | 80 | Urological | 0.002 | (<0.001, 0.004) |
| Women | Dyspnoea | Never smoker | 80 | Haematological | 0.004 | (0.002, 0.007) |
| Women | Dyspnoea | Never smoker | 80 | Other cancers | 0.004 | (0.002, 0.008) |
| Women | Dyspnoea | Never smoker | 80 | Death w/o cancer | 0.070 | (0.054, 0.090) |
| Women | Dyspnoea | Never smoker | 90 | Breast | 0.007 | (0.004, 0.013) |
| Women | Dyspnoea | Never smoker | 90 | Gyanecological | 0.002 | (0.001, 0.004) |
| Women | Dyspnoea | Never smoker | 90 | Lung | 0.003 | (<0.001, 0.009) |
| Women | Dyspnoea | Never smoker | 90 | Upper GI | 0.002 | (<0.001, 0.005) |
| Women | Dyspnoea | Never smoker | 90 | Lower GI | 0.005 | (0.002, 0.011) |
| Women | Dyspnoea | Never smoker | 90 | Urological | 0.002 | (<0.001, 0.004) |
| Women | Dyspnoea | Never smoker | 90 | Haematological | 0.004 | (0.001, 0.009) |
| Women | Dyspnoea | Never smoker | 90 | Other cancers | 0.005 | (0.002, 0.017) |
| Women | Dyspnoea | Never smoker | 90 | Death w/o cancer | 0.213 | (0.195, 0.232) |
| Women | Dyspnoea | Ever smoker | 30 | Gyanecological | 0.004 | (0.002, 0.008) |
| Women | Dyspnoea | Ever smoker | 40 | Gyanecological | 0.001 | (<0.001, 0.003) |
| Women | Dyspnoea | Ever smoker | 40 | Death w/o cancer | 0.001 | (<0.001, 0.003) |
| Women | Dyspnoea | Ever smoker | 50 | Breast | 0.002 | (<0.001, 0.004) |
| Women | Dyspnoea | Ever smoker | 50 | Lung | 0.002 | (<0.001, 0.005) |
| Women | Dyspnoea | Ever smoker | 50 | Other cancers | 0.001 | (<0.001, 0.003) |
| Women | Dyspnoea | Ever smoker | 50 | Death w/o cancer | 0.003 | (0.001, 0.008) |
| Women | Dyspnoea | Ever smoker | 60 | Breast | 0.003 | (0.002, 0.008) |
| Women | Dyspnoea | Ever smoker | 60 | Gyanecological | 0.001 | (<0.001, 0.003) |
| Women | Dyspnoea | Ever smoker | 60 | Lung | 0.007 | (0.003, 0.017) |
| Women | Dyspnoea | Ever smoker | 60 | Lower GI | 0.001 | (<0.001, 0.003) |
| Women | Dyspnoea | Ever smoker | 60 | Haematological | 0.001 | (<0.001, 0.003) |
| Women | Dyspnoea | Ever smoker | 60 | Other cancers | 0.002 | (<0.001, 0.004) |
| Women | Dyspnoea | Ever smoker | 60 | Death w/o cancer | 0.006 | (0.003, 0.011) |
| Women | Dyspnoea | Ever smoker | 70 | Breast | 0.005 | (0.002, 0.013) |
| Women | Dyspnoea | Ever smoker | 70 | Gyanecological | 0.003 | (0.001, 0.008) |
| Women | Dyspnoea | Ever smoker | 70 | Lung | 0.014 | (0.008, 0.024) |
| Women | Dyspnoea | Ever smoker | 70 | Upper GI | 0.002 | (<0.001, 0.004) |
| Women | Dyspnoea | Ever smoker | 70 | Lower GI | 0.003 | (0.001, 0.008) |
| Women | Dyspnoea | Ever smoker | 70 | Urological | 0.002 | (<0.001, 0.003) |
| Women | Dyspnoea | Ever smoker | 70 | Haematological | 0.003 | (0.001, 0.006) |
| Women | Dyspnoea | Ever smoker | 70 | Other cancers | 0.003 | (0.001, 0.008) |
| Women | Dyspnoea | Ever smoker | 70 | Death w/o cancer | 0.020 | (0.013, 0.030) |
| Women | Dyspnoea | Ever smoker | 80 | Breast | 0.007 | (0.003, 0.015) |
| Women | Dyspnoea | Ever smoker | 80 | Gyanecological | 0.003 | (0.001, 0.007) |
| Women | Dyspnoea | Ever smoker | 80 | Lung | 0.015 | (0.009, 0.027) |
| Women | Dyspnoea | Ever smoker | 80 | Upper GI | 0.003 | (0.001, 0.005) |
| Women | Dyspnoea | Ever smoker | 80 | Lower GI | 0.004 | (0.002, 0.007) |
| Women | Dyspnoea | Ever smoker | 80 | Urological | 0.002 | (0.001, 0.005) |
| Women | Dyspnoea | Ever smoker | 80 | Haematological | 0.003 | (0.001, 0.007) |
| Women | Dyspnoea | Ever smoker | 80 | Other cancers | 0.005 | (0.002, 0.012) |
| Women | Dyspnoea | Ever smoker | 80 | Death w/o cancer | 0.066 | (0.051, 0.084) |
| Women | Dyspnoea | Ever smoker | 90 | Breast | 0.008 | (0.003, 0.022) |
| Women | Dyspnoea | Ever smoker | 90 | Gyanecological | 0.002 | (<0.001, 0.005) |
| Women | Dyspnoea | Ever smoker | 90 | Lung | 0.014 | (0.008, 0.024) |
| Women | Dyspnoea | Ever smoker | 90 | Upper GI | 0.003 | (0.001, 0.006) |
| Women | Dyspnoea | Ever smoker | 90 | Lower GI | 0.004 | (0.002, 0.009) |
| Women | Dyspnoea | Ever smoker | 90 | Urological | 0.002 | (0.001, 0.006) |
| Women | Dyspnoea | Ever smoker | 90 | Haematological | 0.004 | (0.002, 0.008) |
| Women | Dyspnoea | Ever smoker | 90 | Other cancers | 0.006 | (0.004, 0.008) |
| Women | Dyspnoea | Ever smoker | 90 | Death w/o cancer | 0.202 | (0.178, 0.228) |
| Women | Fatigue | Never smoker | 30 | Gyanecological | 0.003 | (0.002, 0.007) |
| Women | Fatigue | Never smoker | 40 | Gyanecological | 0.001 | (<0.001, 0.003) |
| Women | Fatigue | Never smoker | 50 | Breast | 0.002 | (<0.001, 0.004) |
| Women | Fatigue | Never smoker | 50 | Gyanecological | 0.001 | (<0.001, 0.002) |
| Women | Fatigue | Never smoker | 50 | Other cancers | 0.002 | (<0.001, 0.003) |
| Women | Fatigue | Never smoker | 50 | Death w/o cancer | 0.002 | (<0.001, 0.005) |
| Women | Fatigue | Never smoker | 60 | Breast | 0.004 | (0.002, 0.007) |
| Women | Fatigue | Never smoker | 60 | Gyanecological | 0.002 | (<0.001, 0.004) |
| Women | Fatigue | Never smoker | 60 | Lower GI | 0.002 | (<0.001, 0.004) |
| Women | Fatigue | Never smoker | 60 | Haematological | 0.002 | (<0.001, 0.004) |
| Women | Fatigue | Never smoker | 60 | Other cancers | 0.003 | (<0.001, 0.007) |
| Women | Fatigue | Never smoker | 60 | Death w/o cancer | 0.004 | (0.002, 0.009) |
| Women | Fatigue | Never smoker | 70 | Breast | 0.006 | (0.003, 0.011) |
| Women | Fatigue | Never smoker | 70 | Gyanecological | 0.003 | (0.001, 0.009) |
| Women | Fatigue | Never smoker | 70 | Lung | 0.002 | (<0.001, 0.003) |
| Women | Fatigue | Never smoker | 70 | Upper GI | 0.002 | (0.001, 0.006) |
| Women | Fatigue | Never smoker | 70 | Lower GI | 0.004 | (0.002, 0.011) |
| Women | Fatigue | Never smoker | 70 | Urological | 0.001 | (<0.001, 0.002) |
| Women | Fatigue | Never smoker | 70 | Haematological | 0.003 | (0.002, 0.006) |
| Women | Fatigue | Never smoker | 70 | Other cancers | 0.005 | (0.002, 0.010) |
| Women | Fatigue | Never smoker | 70 | Death w/o cancer | 0.014 | (0.008, 0.023) |
| Women | Fatigue | Never smoker | 80 | Breast | 0.007 | (0.003, 0.016) |
| Women | Fatigue | Never smoker | 80 | Gyanecological | 0.003 | (0.002, 0.006) |
| Women | Fatigue | Never smoker | 80 | Lung | 0.002 | (<0.001, 0.003) |
| Women | Fatigue | Never smoker | 80 | Upper GI | 0.003 | (0.002, 0.005) |
| Women | Fatigue | Never smoker | 80 | Lower GI | 0.006 | (0.002, 0.015) |
| Women | Fatigue | Never smoker | 80 | Urological | 0.002 | (<0.001, 0.003) |
| Women | Fatigue | Never smoker | 80 | Haematological | 0.004 | (0.001, 0.014) |
| Women | Fatigue | Never smoker | 80 | Other cancers | 0.007 | (0.003, 0.016) |
| Women | Fatigue | Never smoker | 80 | Death w/o cancer | 0.047 | (0.034, 0.064) |
| Women | Fatigue | Never smoker | 90 | Breast | 0.008 | (0.003, 0.021) |
| Women | Fatigue | Never smoker | 90 | Gyanecological | 0.002 | (<0.001, 0.006) |
| Women | Fatigue | Never smoker | 90 | Lung | 0.002 | (<0.001, 0.003) |
| Women | Fatigue | Never smoker | 90 | Upper GI | 0.003 | (0.001, 0.007) |
| Women | Fatigue | Never smoker | 90 | Lower GI | 0.006 | (0.003, 0.012) |
| Women | Fatigue | Never smoker | 90 | Urological | 0.002 | (<0.001, 0.003) |
| Women | Fatigue | Never smoker | 90 | Haematological | 0.005 | (0.002, 0.010) |
| Women | Fatigue | Never smoker | 90 | Other cancers | 0.007 | (0.004, 0.014) |
| Women | Fatigue | Never smoker | 90 | Death w/o cancer | 0.145 | (0.125, 0.167) |
| Women | Fatigue | Ever smoker | 30 | Gyanecological | 0.004 | (0.002, 0.011) |
| Women | Fatigue | Ever smoker | 40 | Gyanecological | 0.002 | (<0.001, 0.003) |
| Women | Fatigue | Ever smoker | 50 | Breast | 0.002 | (0.001, 0.005) |
| Women | Fatigue | Ever smoker | 50 | Gyanecological | 0.001 | (<0.001, 0.002) |
| Women | Fatigue | Ever smoker | 50 | Lung | 0.001 | (<0.001, 0.002) |
| Women | Fatigue | Ever smoker | 50 | Other cancers | 0.002 | (<0.001, 0.004) |
| Women | Fatigue | Ever smoker | 50 | Death w/o cancer | 0.002 | (<0.001, 0.005) |
| Women | Fatigue | Ever smoker | 60 | Breast | 0.004 | (0.002, 0.009) |
| Women | Fatigue | Ever smoker | 60 | Gyanecological | 0.002 | (0.001, 0.004) |
| Women | Fatigue | Ever smoker | 60 | Lung | 0.004 | (0.001, 0.010) |
| Women | Fatigue | Ever smoker | 60 | Upper GI | 0.001 | (<0.001, 0.002) |
| Women | Fatigue | Ever smoker | 60 | Lower GI | 0.002 | (<0.001, 0.005) |
| Women | Fatigue | Ever smoker | 60 | Haematological | 0.002 | (<0.001, 0.005) |
| Women | Fatigue | Ever smoker | 60 | Other cancers | 0.003 | (0.001, 0.006) |
| Women | Fatigue | Ever smoker | 60 | Death w/o cancer | 0.004 | (0.002, 0.008) |
| Women | Fatigue | Ever smoker | 70 | Breast | 0.006 | (0.002, 0.016) |
| Women | Fatigue | Ever smoker | 70 | Gyanecological | 0.003 | (0.001, 0.008) |
| Women | Fatigue | Ever smoker | 70 | Lung | 0.007 | (0.003, 0.015) |
| Women | Fatigue | Ever smoker | 70 | Upper GI | 0.003 | (0.002, 0.004) |
| Women | Fatigue | Ever smoker | 70 | Lower GI | 0.004 | (0.002, 0.010) |
| Women | Fatigue | Ever smoker | 70 | Urological | 0.001 | (<0.001, 0.003) |
| Women | Fatigue | Ever smoker | 70 | Haematological | 0.004 | (0.002, 0.008) |
| Women | Fatigue | Ever smoker | 70 | Other cancers | 0.004 | (0.002, 0.008) |
| Women | Fatigue | Ever smoker | 70 | Death w/o cancer | 0.014 | (0.008, 0.024) |
| Women | Fatigue | Ever smoker | 80 | Breast | 0.007 | (0.004, 0.015) |
| Women | Fatigue | Ever smoker | 80 | Gyanecological | 0.003 | (0.002, 0.007) |
| Women | Fatigue | Ever smoker | 80 | Lung | 0.009 | (0.004, 0.018) |
| Women | Fatigue | Ever smoker | 80 | Upper GI | 0.003 | (0.001, 0.009) |
| Women | Fatigue | Ever smoker | 80 | Lower GI | 0.006 | (0.004, 0.011) |
| Women | Fatigue | Ever smoker | 80 | Urological | 0.002 | (<0.001, 0.004) |
| Women | Fatigue | Ever smoker | 80 | Haematological | 0.005 | (0.002, 0.013) |
| Women | Fatigue | Ever smoker | 80 | Other cancers | 0.007 | (0.003, 0.015) |
| Women | Fatigue | Ever smoker | 80 | Death w/o cancer | 0.046 | (0.035, 0.060) |
| Women | Fatigue | Ever smoker | 90 | Breast | 0.009 | (0.004, 0.017) |
| Women | Fatigue | Ever smoker | 90 | Gyanecological | 0.002 | (0.001, 0.005) |
| Women | Fatigue | Ever smoker | 90 | Lung | 0.007 | (0.004, 0.014) |
| Women | Fatigue | Ever smoker | 90 | Upper GI | 0.003 | (0.002, 0.007) |
| Women | Fatigue | Ever smoker | 90 | Lower GI | 0.006 | (0.003, 0.012) |
| Women | Fatigue | Ever smoker | 90 | Urological | 0.002 | (<0.001, 0.005) |
| Women | Fatigue | Ever smoker | 90 | Haematological | 0.005 | (0.002, 0.012) |
| Women | Fatigue | Ever smoker | 90 | Other cancers | 0.008 | (0.004, 0.016) |
| Women | Fatigue | Ever smoker | 90 | Death w/o cancer | 0.141 | (0.122, 0.162) |
| Women | Haematuria | Never smoker | 30 | Gyanecological | 0.014 | (0.009, 0.023) |
| Women | Haematuria | Never smoker | 40 | Breast | 0.001 | (<0.001, 0.002) |
| Women | Haematuria | Never smoker | 40 | Gyanecological | 0.006 | (0.002, 0.016) |
| Women | Haematuria | Never smoker | 40 | Urological | 0.003 | (0.001, 0.008) |
| Women | Haematuria | Never smoker | 50 | Breast | 0.002 | (<0.001, 0.005) |
| Women | Haematuria | Never smoker | 50 | Gyanecological | 0.005 | (0.002, 0.009) |
| Women | Haematuria | Never smoker | 50 | Urological | 0.010 | (0.005, 0.020) |
| Women | Haematuria | Never smoker | 50 | Other cancers | 0.002 | (<0.001, 0.003) |
| Women | Haematuria | Never smoker | 50 | Death w/o cancer | 0.002 | (<0.001, 0.004) |
| Women | Haematuria | Never smoker | 60 | Breast | 0.004 | (0.002, 0.008) |
| Women | Haematuria | Never smoker | 60 | Gyanecological | 0.007 | (0.003, 0.016) |
| Women | Haematuria | Never smoker | 60 | Lower GI | 0.001 | (<0.001, 0.003) |
| Women | Haematuria | Never smoker | 60 | Urological | 0.026 | (0.018, 0.038) |
| Women | Haematuria | Never smoker | 60 | Other cancers | 0.002 | (0.001, 0.005) |
| Women | Haematuria | Never smoker | 60 | Death w/o cancer | 0.004 | (0.002, 0.007) |
| Women | Haematuria | Never smoker | 70 | Breast | 0.006 | (0.004, 0.009) |
| Women | Haematuria | Never smoker | 70 | Gyanecological | 0.014 | (0.008, 0.024) |
| Women | Haematuria | Never smoker | 70 | Lower GI | 0.003 | (0.001, 0.005) |
| Women | Haematuria | Never smoker | 70 | Urological | 0.057 | (0.043, 0.075) |
| Women | Haematuria | Never smoker | 70 | Haematological | 0.002 | (<0.001, 0.005) |
| Women | Haematuria | Never smoker | 70 | Other cancers | 0.004 | (0.002, 0.008) |
| Women | Haematuria | Never smoker | 70 | Death w/o cancer | 0.014 | (0.007, 0.027) |
| Women | Haematuria | Never smoker | 80 | Breast | 0.008 | (0.002, 0.028) |
| Women | Haematuria | Never smoker | 80 | Gyanecological | 0.013 | (0.008, 0.022) |
| Women | Haematuria | Never smoker | 80 | Lung | 0.001 | (<0.001, 0.003) |
| Women | Haematuria | Never smoker | 80 | Upper GI | 0.001 | (<0.001, 0.002) |
| Women | Haematuria | Never smoker | 80 | Lower GI | 0.003 | (0.001, 0.008) |
| Women | Haematuria | Never smoker | 80 | Urological | 0.078 | (0.058, 0.106) |
| Women | Haematuria | Never smoker | 80 | Haematological | 0.002 | (0.001, 0.005) |
| Women | Haematuria | Never smoker | 80 | Other cancers | 0.005 | (0.003, 0.008) |
| Women | Haematuria | Never smoker | 80 | Death w/o cancer | 0.045 | (0.032, 0.063) |
| Women | Haematuria | Never smoker | 90 | Breast | 0.009 | (0.004, 0.018) |
| Women | Haematuria | Never smoker | 90 | Gyanecological | 0.009 | (0.003, 0.023) |
| Women | Haematuria | Never smoker | 90 | Upper GI | 0.001 | (<0.001, 0.003) |
| Women | Haematuria | Never smoker | 90 | Lower GI | 0.003 | (0.002, 0.007) |
| Women | Haematuria | Never smoker | 90 | Urological | 0.091 | (0.069, 0.120) |
| Women | Haematuria | Never smoker | 90 | Haematological | 0.003 | (0.001, 0.005) |
| Women | Haematuria | Never smoker | 90 | Other cancers | 0.006 | (0.002, 0.017) |
| Women | Haematuria | Never smoker | 90 | Death w/o cancer | 0.137 | (0.111, 0.167) |
| Women | Haematuria | Ever smoker | 30 | Gyanecological | 0.016 | (0.009, 0.026) |
| Women | Haematuria | Ever smoker | 40 | Gyanecological | 0.007 | (0.002, 0.019) |
| Women | Haematuria | Ever smoker | 40 | Urological | 0.004 | (0.002, 0.011) |
| Women | Haematuria | Ever smoker | 50 | Breast | 0.002 | (0.001, 0.005) |
| Women | Haematuria | Ever smoker | 50 | Gyanecological | 0.005 | (0.002, 0.012) |
| Women | Haematuria | Ever smoker | 50 | Urological | 0.013 | (0.007, 0.025) |
| Women | Haematuria | Ever smoker | 50 | Other cancers | 0.001 | (<0.001, 0.003) |
| Women | Haematuria | Ever smoker | 50 | Death w/o cancer | 0.003 | (0.001, 0.006) |
| Women | Haematuria | Ever smoker | 60 | Breast | 0.004 | (0.002, 0.009) |
| Women | Haematuria | Ever smoker | 60 | Gyanecological | 0.008 | (0.003, 0.019) |
| Women | Haematuria | Ever smoker | 60 | Lung | 0.003 | (0.001, 0.007) |
| Women | Haematuria | Ever smoker | 60 | Lower GI | 0.001 | (<0.001, 0.003) |
| Women | Haematuria | Ever smoker | 60 | Urological | 0.033 | (0.025, 0.044) |
| Women | Haematuria | Ever smoker | 60 | Haematological | 0.001 | (<0.001, 0.002) |
| Women | Haematuria | Ever smoker | 60 | Other cancers | 0.002 | (<0.001, 0.006) |
| Women | Haematuria | Ever smoker | 60 | Death w/o cancer | 0.004 | (0.002, 0.010) |
| Women | Haematuria | Ever smoker | 70 | Breast | 0.006 | (0.002, 0.017) |
| Women | Haematuria | Ever smoker | 70 | Gyanecological | 0.014 | (0.008, 0.023) |
| Women | Haematuria | Ever smoker | 70 | Lung | 0.005 | (0.002, 0.013) |
| Women | Haematuria | Ever smoker | 70 | Upper GI | 0.001 | (<0.001, 0.002) |
| Women | Haematuria | Ever smoker | 70 | Lower GI | 0.002 | (0.001, 0.005) |
| Women | Haematuria | Ever smoker | 70 | Urological | 0.070 | (0.053, 0.092) |
| Women | Haematuria | Ever smoker | 70 | Haematological | 0.002 | (<0.001, 0.006) |
| Women | Haematuria | Ever smoker | 70 | Other cancers | 0.004 | (0.002, 0.010) |
| Women | Haematuria | Ever smoker | 70 | Death w/o cancer | 0.013 | (0.008, 0.023) |
| Women | Haematuria | Ever smoker | 80 | Breast | 0.008 | (0.003, 0.022) |
| Women | Haematuria | Ever smoker | 80 | Gyanecological | 0.013 | (0.007, 0.023) |
| Women | Haematuria | Ever smoker | 80 | Lung | 0.005 | (0.002, 0.013) |
| Women | Haematuria | Ever smoker | 80 | Upper GI | 0.002 | (<0.001, 0.003) |
| Women | Haematuria | Ever smoker | 80 | Lower GI | 0.003 | (0.001, 0.007) |
| Women | Haematuria | Ever smoker | 80 | Urological | 0.098 | (0.082, 0.117) |
| Women | Haematuria | Ever smoker | 80 | Haematological | 0.002 | (0.001, 0.004) |
| Women | Haematuria | Ever smoker | 80 | Other cancers | 0.005 | (0.002, 0.015) |
| Women | Haematuria | Ever smoker | 80 | Death w/o cancer | 0.043 | (0.032, 0.059) |
| Women | Haematuria | Ever smoker | 90 | Breast | 0.009 | (0.005, 0.018) |
| Women | Haematuria | Ever smoker | 90 | Gyanecological | 0.009 | (0.005, 0.015) |
| Women | Haematuria | Ever smoker | 90 | Lung | 0.005 | (0.002, 0.014) |
| Women | Haematuria | Ever smoker | 90 | Upper GI | 0.002 | (<0.001, 0.004) |
| Women | Haematuria | Ever smoker | 90 | Lower GI | 0.003 | (0.001, 0.007) |
| Women | Haematuria | Ever smoker | 90 | Urological | 0.112 | (0.087, 0.143) |
| Women | Haematuria | Ever smoker | 90 | Haematological | 0.003 | (<0.001, 0.008) |
| Women | Haematuria | Ever smoker | 90 | Other cancers | 0.007 | (0.004, 0.013) |
| Women | Haematuria | Ever smoker | 90 | Death w/o cancer | 0.130 | (0.105, 0.159) |
| Women | Haemoptysis | Never smoker | 30 | Gyanecological | 0.002 | (<0.001, 0.006) |
| Women | Haemoptysis | Never smoker | 40 | Breast | 0.001 | (<0.001, 0.002) |
| Women | Haemoptysis | Never smoker | 40 | Death w/o cancer | 0.001 | (<0.001, 0.002) |
| Women | Haemoptysis | Never smoker | 50 | Breast | 0.002 | (0.001, 0.005) |
| Women | Haemoptysis | Never smoker | 50 | Lung | 0.003 | (<0.001, 0.010) |
| Women | Haemoptysis | Never smoker | 50 | Haematological | 0.001 | (<0.001, 0.003) |
| Women | Haemoptysis | Never smoker | 50 | Other cancers | 0.002 | (<0.001, 0.004) |
| Women | Haemoptysis | Never smoker | 50 | Death w/o cancer | 0.003 | (0.001, 0.007) |
| Women | Haemoptysis | Never smoker | 60 | Breast | 0.005 | (0.002, 0.010) |
| Women | Haemoptysis | Never smoker | 60 | Gyanecological | 0.001 | (<0.001, 0.002) |
| Women | Haemoptysis | Never smoker | 60 | Lung | 0.011 | (0.005, 0.023) |
| Women | Haemoptysis | Never smoker | 60 | Lower GI | 0.001 | (<0.001, 0.003) |
| Women | Haemoptysis | Never smoker | 60 | Haematological | 0.003 | (0.001, 0.008) |
| Women | Haemoptysis | Never smoker | 60 | Other cancers | 0.003 | (0.001, 0.007) |
| Women | Haemoptysis | Never smoker | 60 | Death w/o cancer | 0.005 | (0.002, 0.012) |
| Women | Haemoptysis | Never smoker | 70 | Breast | 0.007 | (0.002, 0.019) |
| Women | Haemoptysis | Never smoker | 70 | Gyanecological | 0.002 | (<0.001, 0.006) |
| Women | Haemoptysis | Never smoker | 70 | Lung | 0.021 | (0.015, 0.030) |
| Women | Haemoptysis | Never smoker | 70 | Lower GI | 0.003 | (<0.001, 0.006) |
| Women | Haemoptysis | Never smoker | 70 | Haematological | 0.005 | (0.002, 0.015) |
| Women | Haemoptysis | Never smoker | 70 | Other cancers | 0.005 | (0.002, 0.012) |
| Women | Haemoptysis | Never smoker | 70 | Death w/o cancer | 0.016 | (0.010, 0.026) |
| Women | Haemoptysis | Never smoker | 80 | Breast | 0.009 | (0.004, 0.020) |
| Women | Haemoptysis | Never smoker | 80 | Gyanecological | 0.002 | (0.001, 0.004) |
| Women | Haemoptysis | Never smoker | 80 | Lung | 0.024 | (0.015, 0.041) |
| Women | Haemoptysis | Never smoker | 80 | Upper GI | 0.002 | (<0.001, 0.004) |
| Women | Haemoptysis | Never smoker | 80 | Lower GI | 0.004 | (0.001, 0.011) |
| Women | Haemoptysis | Never smoker | 80 | Haematological | 0.007 | (0.002, 0.022) |
| Women | Haemoptysis | Never smoker | 80 | Other cancers | 0.007 | (0.002, 0.024) |
| Women | Haemoptysis | Never smoker | 80 | Death w/o cancer | 0.054 | (0.036, 0.081) |
| Women | Haemoptysis | Never smoker | 90 | Breast | 0.009 | (0.005, 0.017) |
| Women | Haemoptysis | Never smoker | 90 | Gyanecological | 0.001 | (<0.001, 0.003) |
| Women | Haemoptysis | Never smoker | 90 | Lung | 0.021 | (0.013, 0.034) |
| Women | Haemoptysis | Never smoker | 90 | Upper GI | 0.001 | (<0.001, 0.003) |
| Women | Haemoptysis | Never smoker | 90 | Lower GI | 0.004 | (0.001, 0.012) |
| Women | Haemoptysis | Never smoker | 90 | Haematological | 0.006 | (0.003, 0.013) |
| Women | Haemoptysis | Never smoker | 90 | Other cancers | 0.008 | (0.004, 0.019) |
| Women | Haemoptysis | Never smoker | 90 | Death w/o cancer | 0.165 | (0.128, 0.211) |
| Women | Haemoptysis | Ever smoker | 30 | Gyanecological | 0.002 | (0.001, 0.004) |
| Women | Haemoptysis | Ever smoker | 40 | Breast | 0.001 | (<0.001, 0.002) |
| Women | Haemoptysis | Ever smoker | 40 | Gyanecological | 0.001 | (<0.001, 0.002) |
| Women | Haemoptysis | Ever smoker | 40 | Lung | 0.002 | (<0.001, 0.005) |
| Women | Haemoptysis | Ever smoker | 40 | Death w/o cancer | 0.001 | (<0.001, 0.002) |
| Women | Haemoptysis | Ever smoker | 50 | Breast | 0.003 | (0.001, 0.006) |
| Women | Haemoptysis | Ever smoker | 50 | Lung | 0.016 | (0.008, 0.029) |
| Women | Haemoptysis | Ever smoker | 50 | Haematological | 0.002 | (<0.001, 0.003) |
| Women | Haemoptysis | Ever smoker | 50 | Other cancers | 0.002 | (<0.001, 0.004) |
| Women | Haemoptysis | Ever smoker | 50 | Death w/o cancer | 0.003 | (<0.001, 0.007) |
| Women | Haemoptysis | Ever smoker | 60 | Breast | 0.004 | (0.001, 0.014) |
| Women | Haemoptysis | Ever smoker | 60 | Gyanecological | 0.001 | (<0.001, 0.002) |
| Women | Haemoptysis | Ever smoker | 60 | Lung | 0.049 | (0.037, 0.064) |
| Women | Haemoptysis | Ever smoker | 60 | Lower GI | 0.001 | (<0.001, 0.002) |
| Women | Haemoptysis | Ever smoker | 60 | Haematological | 0.003 | (0.001, 0.006) |
| Women | Haemoptysis | Ever smoker | 60 | Other cancers | 0.003 | (0.001, 0.009) |
| Women | Haemoptysis | Ever smoker | 60 | Death w/o cancer | 0.005 | (0.001, 0.014) |
| Women | Haemoptysis | Ever smoker | 70 | Breast | 0.007 | (0.002, 0.022) |
| Women | Haemoptysis | Ever smoker | 70 | Gyanecological | 0.002 | (0.001, 0.004) |
| Women | Haemoptysis | Ever smoker | 70 | Lung | 0.094 | (0.076, 0.116) |
| Women | Haemoptysis | Ever smoker | 70 | Upper GI | 0.001 | (<0.001, 0.003) |
| Women | Haemoptysis | Ever smoker | 70 | Lower GI | 0.003 | (0.001, 0.006) |
| Women | Haemoptysis | Ever smoker | 70 | Haematological | 0.005 | (0.001, 0.015) |
| Women | Haemoptysis | Ever smoker | 70 | Other cancers | 0.005 | (0.002, 0.013) |
| Women | Haemoptysis | Ever smoker | 70 | Death w/o cancer | 0.015 | (0.008, 0.028) |
| Women | Haemoptysis | Ever smoker | 80 | Breast | 0.008 | (0.003, 0.019) |
| Women | Haemoptysis | Ever smoker | 80 | Gyanecological | 0.002 | (<0.001, 0.004) |
| Women | Haemoptysis | Ever smoker | 80 | Lung | 0.108 | (0.080, 0.143) |
| Women | Haemoptysis | Ever smoker | 80 | Upper GI | 0.002 | (<0.001, 0.005) |
| Women | Haemoptysis | Ever smoker | 80 | Lower GI | 0.003 | (<0.001, 0.011) |
| Women | Haemoptysis | Ever smoker | 80 | Haematological | 0.006 | (0.002, 0.022) |
| Women | Haemoptysis | Ever smoker | 80 | Other cancers | 0.007 | (0.002, 0.019) |
| Women | Haemoptysis | Ever smoker | 80 | Death w/o cancer | 0.049 | (0.036, 0.068) |
| Women | Haemoptysis | Ever smoker | 90 | Breast | 0.009 | (0.004, 0.020) |
| Women | Haemoptysis | Ever smoker | 90 | Gyanecological | 0.001 | (<0.001, 0.003) |
| Women | Haemoptysis | Ever smoker | 90 | Lung | 0.098 | (0.074, 0.129) |
| Women | Haemoptysis | Ever smoker | 90 | Upper GI | 0.002 | (<0.001, 0.004) |
| Women | Haemoptysis | Ever smoker | 90 | Lower GI | 0.003 | (0.001, 0.009) |
| Women | Haemoptysis | Ever smoker | 90 | Haematological | 0.006 | (0.003, 0.011) |
| Women | Haemoptysis | Ever smoker | 90 | Other cancers | 0.008 | (0.003, 0.020) |
| Women | Haemoptysis | Ever smoker | 90 | Death w/o cancer | 0.151 | (0.110, 0.205) |
| Women | Jaundice | Never smoker | 30 | Gyanecological | 0.004 | (0.001, 0.009) |
| Women | Jaundice | Never smoker | 30 | Upper GI | 0.003 | (0.002, 0.005) |
| Women | Jaundice | Never smoker | 30 | Other cancers | 0.007 | (0.003, 0.014) |
| Women | Jaundice | Never smoker | 30 | Death w/o cancer | 0.002 | (<0.001, 0.005) |
| Women | Jaundice | Never smoker | 40 | Breast | 0.002 | (0.001, 0.003) |
| Women | Jaundice | Never smoker | 40 | Gyanecological | 0.001 | (<0.001, 0.004) |
| Women | Jaundice | Never smoker | 40 | Upper GI | 0.012 | (0.006, 0.027) |
| Women | Jaundice | Never smoker | 40 | Other cancers | 0.010 | (0.005, 0.019) |
| Women | Jaundice | Never smoker | 40 | Death w/o cancer | 0.004 | (0.002, 0.009) |
| Women | Jaundice | Never smoker | 50 | Breast | 0.004 | (0.002, 0.010) |
| Women | Jaundice | Never smoker | 50 | Upper GI | 0.044 | (0.028, 0.068) |
| Women | Jaundice | Never smoker | 50 | Other cancers | 0.023 | (0.016, 0.034) |
| Women | Jaundice | Never smoker | 50 | Death w/o cancer | 0.008 | (0.004, 0.015) |
| Women | Jaundice | Never smoker | 60 | Breast | 0.006 | (0.002, 0.018) |
| Women | Jaundice | Never smoker | 60 | Gyanecological | 0.002 | (<0.001, 0.006) |
| Women | Jaundice | Never smoker | 60 | Upper GI | 0.101 | (0.083, 0.123) |
| Women | Jaundice | Never smoker | 60 | Lower GI | 0.003 | (<0.001, 0.008) |
| Women | Jaundice | Never smoker | 60 | Urological | 0.002 | (<0.001, 0.004) |
| Women | Jaundice | Never smoker | 60 | Other cancers | 0.035 | (0.020, 0.063) |
| Women | Jaundice | Never smoker | 60 | Death w/o cancer | 0.015 | (0.009, 0.024) |
| Women | Jaundice | Never smoker | 70 | Breast | 0.008 | (0.004, 0.019) |
| Women | Jaundice | Never smoker | 70 | Gyanecological | 0.002 | (<0.001, 0.006) |
| Women | Jaundice | Never smoker | 70 | Lung | 0.001 | (<0.001, 0.003) |
| Women | Jaundice | Never smoker | 70 | Upper GI | 0.231 | (0.198, 0.268) |
| Women | Jaundice | Never smoker | 70 | Lower GI | 0.005 | (0.001, 0.014) |
| Women | Jaundice | Never smoker | 70 | Urological | 0.004 | (0.002, 0.009) |
| Women | Jaundice | Never smoker | 70 | Other cancers | 0.055 | (0.038, 0.080) |
| Women | Jaundice | Never smoker | 70 | Death w/o cancer | 0.045 | (0.029, 0.068) |
| Women | Jaundice | Never smoker | 80 | Breast | 0.009 | (0.004, 0.021) |
| Women | Jaundice | Never smoker | 80 | Gyanecological | 0.002 | (<0.001, 0.005) |
| Women | Jaundice | Never smoker | 80 | Lung | 0.001 | (<0.001, 0.003) |
| Women | Jaundice | Never smoker | 80 | Upper GI | 0.296 | (0.248, 0.350) |
| Women | Jaundice | Never smoker | 80 | Lower GI | 0.006 | (0.002, 0.018) |
| Women | Jaundice | Never smoker | 80 | Urological | 0.005 | (<0.001, 0.000) |
| Women | Jaundice | Never smoker | 80 | Haematological | 0.001 | (<0.001, 0.003) |
| Women | Jaundice | Never smoker | 80 | Other cancers | 0.072 | (0.050, 0.102) |
| Women | Jaundice | Never smoker | 80 | Death w/o cancer | 0.129 | (0.101, 0.164) |
| Women | Jaundice | Never smoker | 90 | Breast | 0.008 | (0.003, 0.022) |
| Women | Jaundice | Never smoker | 90 | Lung | 0.001 | (<0.001, 0.003) |
| Women | Jaundice | Never smoker | 90 | Upper GI | 0.280 | (0.237, 0.327) |
| Women | Jaundice | Never smoker | 90 | Lower GI | 0.005 | (0.001, 0.018) |
| Women | Jaundice | Never smoker | 90 | Urological | 0.005 | (0.002, 0.012) |
| Women | Jaundice | Never smoker | 90 | Haematological | 0.001 | (<0.001, 0.004) |
| Women | Jaundice | Never smoker | 90 | Other cancers | 0.080 | (0.057, 0.112) |
| Women | Jaundice | Never smoker | 90 | Death w/o cancer | 0.336 | (0.283, 0.392) |
| Women | Jaundice | Ever smoker | 30 | Gyanecological | 0.004 | (<0.001, 0.015) |
| Women | Jaundice | Ever smoker | 30 | Upper GI | 0.003 | (0.001, 0.009) |
| Women | Jaundice | Ever smoker | 30 | Other cancers | 0.007 | (0.003, 0.018) |
| Women | Jaundice | Ever smoker | 30 | Death w/o cancer | 0.002 | (<0.001, 0.005) |
| Women | Jaundice | Ever smoker | 40 | Breast | 0.002 | (<0.001, 0.004) |
| Women | Jaundice | Ever smoker | 40 | Gyanecological | 0.002 | (<0.001, 0.003) |
| Women | Jaundice | Ever smoker | 40 | Upper GI | 0.015 | (0.009, 0.026) |
| Women | Jaundice | Ever smoker | 40 | Other cancers | 0.011 | (0.005, 0.022) |
| Women | Jaundice | Ever smoker | 40 | Death w/o cancer | 0.004 | (0.001, 0.011) |
| Women | Jaundice | Ever smoker | 50 | Breast | 0.004 | (0.002, 0.008) |
| Women | Jaundice | Ever smoker | 50 | Gyanecological | 0.001 | (<0.001, 0.002) |
| Women | Jaundice | Ever smoker | 50 | Upper GI | 0.053 | (0.041, 0.068) |
| Women | Jaundice | Ever smoker | 50 | Lower GI | 0.001 | (<0.001, 0.003) |
| Women | Jaundice | Ever smoker | 50 | Urological | 0.001 | (<0.001, 0.002) |
| Women | Jaundice | Ever smoker | 50 | Other cancers | 0.024 | (0.015, 0.039) |
| Women | Jaundice | Ever smoker | 50 | Death w/o cancer | 0.009 | (0.004, 0.017) |
| Women | Jaundice | Ever smoker | 60 | Breast | 0.007 | (0.002, 0.024) |
| Women | Jaundice | Ever smoker | 60 | Gyanecological | 0.002 | (<0.001, 0.004) |
| Women | Jaundice | Ever smoker | 60 | Lung | 0.003 | (<0.001, 0.009) |
| Women | Jaundice | Ever smoker | 60 | Upper GI | 0.119 | (0.091, 0.156) |
| Women | Jaundice | Ever smoker | 60 | Lower GI | 0.002 | (<0.001, 0.006) |
| Women | Jaundice | Ever smoker | 60 | Urological | 0.003 | (0.001, 0.007) |
| Women | Jaundice | Ever smoker | 60 | Other cancers | 0.037 | (0.023, 0.059) |
| Women | Jaundice | Ever smoker | 60 | Death w/o cancer | 0.014 | (0.008, 0.024) |
| Women | Jaundice | Ever smoker | 70 | Breast | 0.008 | (0.002, 0.029) |
| Women | Jaundice | Ever smoker | 70 | Gyanecological | 0.002 | (<0.001, 0.007) |
| Women | Jaundice | Ever smoker | 70 | Lung | 0.006 | (0.001, 0.032) |
| Women | Jaundice | Ever smoker | 70 | Upper GI | 0.267 | (0.220, 0.322) |
| Women | Jaundice | Ever smoker | 70 | Lower GI | 0.005 | (0.002, 0.012) |
| Women | Jaundice | Ever smoker | 70 | Urological | 0.005 | (0.002, 0.017) |
| Women | Jaundice | Ever smoker | 70 | Haematological | 0.001 | (<0.001, 0.003) |
| Women | Jaundice | Ever smoker | 70 | Other cancers | 0.057 | (0.039, 0.084) |
| Women | Jaundice | Ever smoker | 70 | Death w/o cancer | 0.040 | (0.029, 0.056) |
| Women | Jaundice | Ever smoker | 80 | Breast | 0.009 | (0.003, 0.026) |
| Women | Jaundice | Ever smoker | 80 | Gyanecological | 0.002 | (<0.001, 0.007) |
| Women | Jaundice | Ever smoker | 80 | Lung | 0.006 | (0.001, 0.030) |
| Women | Jaundice | Ever smoker | 80 | Upper GI | 0.342 | (0.307, 0.380) |
| Women | Jaundice | Ever smoker | 80 | Lower GI | 0.005 | (0.001, 0.020) |
| Women | Jaundice | Ever smoker | 80 | Urological | 0.006 | (0.002, 0.019) |
| Women | Jaundice | Ever smoker | 80 | Haematological | 0.001 | (<0.001, 0.002) |
| Women | Jaundice | Ever smoker | 80 | Other cancers | 0.072 | (0.045, 0.114) |
| Women | Jaundice | Ever smoker | 80 | Death w/o cancer | 0.115 | (0.093, 0.143) |
| Women | Jaundice | Ever smoker | 90 | Breast | 0.008 | (0.004, 0.017) |
| Women | Jaundice | Ever smoker | 90 | Lung | 0.006 | (0.002, 0.021) |
| Women | Jaundice | Ever smoker | 90 | Upper GI | 0.325 | (0.278, 0.376) |
| Women | Jaundice | Ever smoker | 90 | Lower GI | 0.005 | (0.001, 0.016) |
| Women | Jaundice | Ever smoker | 90 | Urological | 0.006 | (0.002, 0.021) |
| Women | Jaundice | Ever smoker | 90 | Haematological | 0.001 | (<0.001, 0.004) |
| Women | Jaundice | Ever smoker | 90 | Other cancers | 0.083 | (0.060, 0.113) |
| Women | Jaundice | Ever smoker | 90 | Death w/o cancer | 0.304 | (0.249, 0.365) |
| Women | Night sweats | Never smoker | 30 | Gyanecological | 0.002 | (0.001, 0.005) |
| Women | Night sweats | Never smoker | 40 | Breast | 0.001 | (<0.001, 0.002) |
| Women | Night sweats | Never smoker | 50 | Breast | 0.002 | (0.001, 0.004) |
| Women | Night sweats | Never smoker | 50 | Haematological | 0.001 | (<0.001, 0.003) |
| Women | Night sweats | Never smoker | 60 | Breast | 0.005 | (<0.001, 0.000) |
| Women | Night sweats | Never smoker | 60 | Gyanecological | 0.001 | (<0.001, 0.002) |
| Women | Night sweats | Never smoker | 60 | Lower GI | 0.001 | (<0.001, 0.003) |
| Women | Night sweats | Never smoker | 60 | Haematological | 0.003 | (0.001, 0.008) |
| Women | Night sweats | Never smoker | 60 | Other cancers | 0.001 | (<0.001, 0.002) |
| Women | Night sweats | Never smoker | 60 | Death w/o cancer | 0.001 | (<0.001, 0.003) |
| Women | Night sweats | Never smoker | 70 | Breast | 0.007 | (0.003, 0.014) |
| Women | Night sweats | Never smoker | 70 | Gyanecological | 0.002 | (0.001, 0.005) |
| Women | Night sweats | Never smoker | 70 | Lung | 0.002 | (<0.001, 0.003) |
| Women | Night sweats | Never smoker | 70 | Lower GI | 0.003 | (0.001, 0.006) |
| Women | Night sweats | Never smoker | 70 | Urological | 0.002 | (<0.001, 0.004) |
| Women | Night sweats | Never smoker | 70 | Haematological | 0.006 | (0.002, 0.017) |
| Women | Night sweats | Never smoker | 70 | Other cancers | 0.002 | (<0.001, 0.006) |
| Women | Night sweats | Never smoker | 70 | Death w/o cancer | 0.005 | (0.003, 0.010) |
| Women | Night sweats | Never smoker | 80 | Breast | 0.009 | (0.005, 0.017) |
| Women | Night sweats | Never smoker | 80 | Gyanecological | 0.002 | (<0.001, 0.004) |
| Women | Night sweats | Never smoker | 80 | Lung | 0.002 | (<0.001, 0.004) |
| Women | Night sweats | Never smoker | 80 | Upper GI | 0.001 | (<0.001, 0.003) |
| Women | Night sweats | Never smoker | 80 | Lower GI | 0.004 | (0.002, 0.009) |
| Women | Night sweats | Never smoker | 80 | Urological | 0.003 | (0.001, 0.007) |
| Women | Night sweats | Never smoker | 80 | Haematological | 0.008 | (0.004, 0.018) |
| Women | Night sweats | Never smoker | 80 | Other cancers | 0.004 | (0.001, 0.010) |
| Women | Night sweats | Never smoker | 80 | Death w/o cancer | 0.017 | (0.008, 0.035) |
| Women | Night sweats | Never smoker | 90 | Breast | 0.010 | (0.004, 0.024) |
| Women | Night sweats | Never smoker | 90 | Gyanecological | 0.001 | (<0.001, 0.003) |
| Women | Night sweats | Never smoker | 90 | Lung | 0.002 | (<0.001, 0.004) |
| Women | Night sweats | Never smoker | 90 | Upper GI | 0.001 | (<0.001, 0.004) |
| Women | Night sweats | Never smoker | 90 | Lower GI | 0.004 | (0.002, 0.010) |
| Women | Night sweats | Never smoker | 90 | Urological | 0.003 | (0.001, 0.010) |
| Women | Night sweats | Never smoker | 90 | Haematological | 0.008 | (0.002, 0.033) |
| Women | Night sweats | Never smoker | 90 | Other cancers | 0.004 | (0.002, 0.009) |
| Women | Night sweats | Never smoker | 90 | Death w/o cancer | 0.056 | (0.030, 0.102) |
| Women | Night sweats | Ever smoker | 30 | Gyanecological | 0.003 | (0.001, 0.006) |
| Women | Night sweats | Ever smoker | 50 | Breast | 0.002 | (0.001, 0.005) |
| Women | Night sweats | Ever smoker | 50 | Lung | 0.001 | (<0.001, 0.003) |
| Women | Night sweats | Ever smoker | 50 | Haematological | 0.001 | (<0.001, 0.003) |
| Women | Night sweats | Ever smoker | 50 | Other cancers | 0.001 | (<0.001, 0.002) |
| Women | Night sweats | Ever smoker | 60 | Breast | 0.004 | (0.002, 0.012) |
| Women | Night sweats | Ever smoker | 60 | Gyanecological | 0.001 | (<0.001, 0.002) |
| Women | Night sweats | Ever smoker | 60 | Lung | 0.004 | (0.001, 0.013) |
| Women | Night sweats | Ever smoker | 60 | Lower GI | 0.001 | (<0.001, 0.002) |
| Women | Night sweats | Ever smoker | 60 | Urological | 0.001 | (<0.001, 0.002) |
| Women | Night sweats | Ever smoker | 60 | Haematological | 0.003 | (0.001, 0.008) |
| Women | Night sweats | Ever smoker | 60 | Other cancers | 0.002 | (<0.001, 0.003) |
| Women | Night sweats | Ever smoker | 60 | Death w/o cancer | 0.001 | (<0.001, 0.003) |
| Women | Night sweats | Ever smoker | 70 | Breast | 0.007 | (0.003, 0.019) |
| Women | Night sweats | Ever smoker | 70 | Gyanecological | 0.002 | (<0.001, 0.005) |
| Women | Night sweats | Ever smoker | 70 | Lung | 0.008 | (0.003, 0.024) |
| Women | Night sweats | Ever smoker | 70 | Upper GI | 0.001 | (<0.001, 0.002) |
| Women | Night sweats | Ever smoker | 70 | Lower GI | 0.003 | (0.001, 0.007) |
| Women | Night sweats | Ever smoker | 70 | Urological | 0.002 | (<0.001, 0.007) |
| Women | Night sweats | Ever smoker | 70 | Haematological | 0.007 | (0.003, 0.016) |
| Women | Night sweats | Ever smoker | 70 | Other cancers | 0.003 | (0.001, 0.006) |
| Women | Night sweats | Ever smoker | 70 | Death w/o cancer | 0.005 | (0.002, 0.015) |
| Women | Night sweats | Ever smoker | 80 | Breast | 0.009 | (0.004, 0.024) |
| Women | Night sweats | Ever smoker | 80 | Gyanecological | 0.002 | (<0.001, 0.004) |
| Women | Night sweats | Ever smoker | 80 | Lung | 0.009 | (0.003, 0.024) |
| Women | Night sweats | Ever smoker | 80 | Upper GI | 0.002 | (<0.001, 0.004) |
| Women | Night sweats | Ever smoker | 80 | Lower GI | 0.004 | (0.001, 0.012) |
| Women | Night sweats | Ever smoker | 80 | Urological | 0.004 | (0.002, 0.009) |
| Women | Night sweats | Ever smoker | 80 | Haematological | 0.008 | (0.004, 0.019) |
| Women | Night sweats | Ever smoker | 80 | Other cancers | 0.004 | (0.002, 0.007) |
| Women | Night sweats | Ever smoker | 80 | Death w/o cancer | 0.017 | (0.008, 0.039) |
| Women | Night sweats | Ever smoker | 90 | Breast | 0.011 | (0.006, 0.021) |
| Women | Night sweats | Ever smoker | 90 | Gyanecological | 0.001 | (<0.001, 0.003) |
| Women | Night sweats | Ever smoker | 90 | Lung | 0.008 | (0.002, 0.032) |
| Women | Night sweats | Ever smoker | 90 | Upper GI | 0.002 | (<0.001, 0.004) |
| Women | Night sweats | Ever smoker | 90 | Lower GI | 0.004 | (0.002, 0.008) |
| Women | Night sweats | Ever smoker | 90 | Urological | 0.004 | (0.001, 0.013) |
| Women | Night sweats | Ever smoker | 90 | Haematological | 0.009 | (0.004, 0.017) |
| Women | Night sweats | Ever smoker | 90 | Other cancers | 0.005 | (0.001, 0.016) |
| Women | Night sweats | Ever smoker | 90 | Death w/o cancer | 0.054 | (0.033, 0.087) |
| Women | Post-menopausal bleed | Never smoker | 30 | Gyanecological | 0.145 | (0.108, 0.190) |
| Women | Post-menopausal bleed | Never smoker | 40 | Gyanecological | 0.068 | (0.045, 0.101) |
| Women | Post-menopausal bleed | Never smoker | 50 | Breast | 0.002 | (<0.001, 0.004) |
| Women | Post-menopausal bleed | Never smoker | 50 | Gyanecological | 0.046 | (0.028, 0.077) |
| Women | Post-menopausal bleed | Never smoker | 50 | Other cancers | 0.001 | (<0.001, 0.002) |
| Women | Post-menopausal bleed | Never smoker | 50 | Death w/o cancer | 0.001 | (<0.001, 0.002) |
| Women | Post-menopausal bleed | Never smoker | 60 | Breast | 0.003 | (0.001, 0.007) |
| Women | Post-menopausal bleed | Never smoker | 60 | Gyanecological | 0.078 | (0.060, 0.103) |
| Women | Post-menopausal bleed | Never smoker | 60 | Lower GI | 0.001 | (<0.001, 0.002) |
| Women | Post-menopausal bleed | Never smoker | 60 | Urological | 0.002 | (<0.001, 0.003) |
| Women | Post-menopausal bleed | Never smoker | 60 | Haematological | 0.001 | (<0.001, 0.002) |
| Women | Post-menopausal bleed | Never smoker | 60 | Other cancers | 0.002 | (<0.001, 0.005) |
| Women | Post-menopausal bleed | Never smoker | 60 | Death w/o cancer | 0.002 | (<0.001, 0.005) |
| Women | Post-menopausal bleed | Never smoker | 70 | Breast | 0.005 | (0.002, 0.012) |
| Women | Post-menopausal bleed | Never smoker | 70 | Gyanecological | 0.136 | (0.111, 0.164) |
| Women | Post-menopausal bleed | Never smoker | 70 | Upper GI | 0.002 | (<0.001, 0.004) |
| Women | Post-menopausal bleed | Never smoker | 70 | Lower GI | 0.002 | (0.001, 0.004) |
| Women | Post-menopausal bleed | Never smoker | 70 | Urological | 0.003 | (0.001, 0.009) |
| Women | Post-menopausal bleed | Never smoker | 70 | Haematological | 0.002 | (<0.001, 0.004) |
| Women | Post-menopausal bleed | Never smoker | 70 | Other cancers | 0.003 | (<0.001, 0.008) |
| Women | Post-menopausal bleed | Never smoker | 70 | Death w/o cancer | 0.008 | (0.004, 0.016) |
| Women | Post-menopausal bleed | Never smoker | 80 | Breast | 0.006 | (0.004, 0.011) |
| Women | Post-menopausal bleed | Never smoker | 80 | Gyanecological | 0.130 | (0.104, 0.161) |
| Women | Post-menopausal bleed | Never smoker | 80 | Upper GI | 0.002 | (0.001, 0.004) |
| Women | Post-menopausal bleed | Never smoker | 80 | Lower GI | 0.003 | (0.001, 0.008) |
| Women | Post-menopausal bleed | Never smoker | 80 | Urological | 0.005 | (0.002, 0.013) |
| Women | Post-menopausal bleed | Never smoker | 80 | Haematological | 0.002 | (<0.001, 0.004) |
| Women | Post-menopausal bleed | Never smoker | 80 | Other cancers | 0.004 | (0.001, 0.014) |
| Women | Post-menopausal bleed | Never smoker | 80 | Death w/o cancer | 0.026 | (0.018, 0.037) |
| Women | Post-menopausal bleed | Never smoker | 90 | Breast | 0.007 | (0.004, 0.014) |
| Women | Post-menopausal bleed | Never smoker | 90 | Gyanecological | 0.096 | (0.074, 0.124) |
| Women | Post-menopausal bleed | Never smoker | 90 | Upper GI | 0.002 | (0.001, 0.004) |
| Women | Post-menopausal bleed | Never smoker | 90 | Lower GI | 0.004 | (0.002, 0.009) |
| Women | Post-menopausal bleed | Never smoker | 90 | Urological | 0.006 | (0.002, 0.018) |
| Women | Post-menopausal bleed | Never smoker | 90 | Haematological | 0.002 | (<0.001, 0.005) |
| Women | Post-menopausal bleed | Never smoker | 90 | Other cancers | 0.005 | (0.002, 0.016) |
| Women | Post-menopausal bleed | Never smoker | 90 | Death w/o cancer | 0.085 | (0.066, 0.109) |
| Women | Post-menopausal bleed | Ever smoker | 30 | Gyanecological | 0.153 | (0.122, 0.191) |
| Women | Post-menopausal bleed | Ever smoker | 40 | Gyanecological | 0.072 | (0.048, 0.105) |
| Women | Post-menopausal bleed | Ever smoker | 50 | Breast | 0.002 | (0.001, 0.003) |
| Women | Post-menopausal bleed | Ever smoker | 50 | Gyanecological | 0.049 | (0.035, 0.068) |
| Women | Post-menopausal bleed | Ever smoker | 50 | Other cancers | 0.001 | (<0.001, 0.003) |
| Women | Post-menopausal bleed | Ever smoker | 50 | Death w/o cancer | 0.001 | (<0.001, 0.003) |
| Women | Post-menopausal bleed | Ever smoker | 60 | Breast | 0.003 | (0.002, 0.006) |
| Women | Post-menopausal bleed | Ever smoker | 60 | Gyanecological | 0.083 | (0.061, 0.113) |
| Women | Post-menopausal bleed | Ever smoker | 60 | Urological | 0.002 | (<0.001, 0.005) |
| Women | Post-menopausal bleed | Ever smoker | 60 | Other cancers | 0.002 | (<0.001, 0.004) |
| Women | Post-menopausal bleed | Ever smoker | 60 | Death w/o cancer | 0.002 | (0.001, 0.006) |
| Women | Post-menopausal bleed | Ever smoker | 70 | Breast | 0.005 | (0.003, 0.010) |
| Women | Post-menopausal bleed | Ever smoker | 70 | Gyanecological | 0.144 | (0.119, 0.175) |
| Women | Post-menopausal bleed | Ever smoker | 70 | Upper GI | 0.002 | (<0.001, 0.004) |
| Women | Post-menopausal bleed | Ever smoker | 70 | Lower GI | 0.002 | (<0.001, 0.006) |
| Women | Post-menopausal bleed | Ever smoker | 70 | Urological | 0.004 | (0.002, 0.011) |
| Women | Post-menopausal bleed | Ever smoker | 70 | Haematological | 0.002 | (<0.001, 0.004) |
| Women | Post-menopausal bleed | Ever smoker | 70 | Other cancers | 0.003 | (0.002, 0.007) |
| Women | Post-menopausal bleed | Ever smoker | 70 | Death w/o cancer | 0.007 | (0.003, 0.017) |
| Women | Post-menopausal bleed | Ever smoker | 80 | Breast | 0.007 | (0.003, 0.016) |
| Women | Post-menopausal bleed | Ever smoker | 80 | Gyanecological | 0.139 | (0.115, 0.167) |
| Women | Post-menopausal bleed | Ever smoker | 80 | Lung | 0.001 | (<0.001, 0.002) |
| Women | Post-menopausal bleed | Ever smoker | 80 | Upper GI | 0.003 | (0.001, 0.006) |
| Women | Post-menopausal bleed | Ever smoker | 80 | Lower GI | 0.003 | (0.001, 0.008) |
| Women | Post-menopausal bleed | Ever smoker | 80 | Urological | 0.006 | (0.003, 0.012) |
| Women | Post-menopausal bleed | Ever smoker | 80 | Haematological | 0.002 | (0.001, 0.004) |
| Women | Post-menopausal bleed | Ever smoker | 80 | Other cancers | 0.005 | (0.002, 0.011) |
| Women | Post-menopausal bleed | Ever smoker | 80 | Death w/o cancer | 0.025 | (0.017, 0.036) |
| Women | Post-menopausal bleed | Ever smoker | 90 | Breast | 0.007 | (0.003, 0.020) |
| Women | Post-menopausal bleed | Ever smoker | 90 | Gyanecological | 0.102 | (0.081, 0.128) |
| Women | Post-menopausal bleed | Ever smoker | 90 | Upper GI | 0.003 | (0.001, 0.005) |
| Women | Post-menopausal bleed | Ever smoker | 90 | Lower GI | 0.004 | (0.001, 0.010) |
| Women | Post-menopausal bleed | Ever smoker | 90 | Urological | 0.008 | (0.003, 0.020) |
| Women | Post-menopausal bleed | Ever smoker | 90 | Haematological | 0.002 | (0.001, 0.004) |
| Women | Post-menopausal bleed | Ever smoker | 90 | Other cancers | 0.006 | (0.002, 0.015) |
| Women | Post-menopausal bleed | Ever smoker | 90 | Death w/o cancer | 0.081 | (0.059, 0.109) |
| Women | Rectal bleeding | Never smoker | 30 | Gyanecological | 0.004 | (0.002, 0.010) |
| Women | Rectal bleeding | Never smoker | 40 | Gyanecological | 0.002 | (0.001, 0.004) |
| Women | Rectal bleeding | Never smoker | 40 | Lower GI | 0.002 | (<0.001, 0.006) |
| Women | Rectal bleeding | Never smoker | 40 | Other cancers | 0.001 | (<0.001, 0.002) |
| Women | Rectal bleeding | Never smoker | 50 | Breast | 0.002 | (<0.001, 0.003) |
| Women | Rectal bleeding | Never smoker | 50 | Gyanecological | 0.001 | (<0.001, 0.002) |
| Women | Rectal bleeding | Never smoker | 50 | Lower GI | 0.009 | (0.004, 0.017) |
| Women | Rectal bleeding | Never smoker | 50 | Other cancers | 0.003 | (0.001, 0.006) |
| Women | Rectal bleeding | Never smoker | 50 | Death w/o cancer | 0.003 | (<0.001, 0.007) |
| Women | Rectal bleeding | Never smoker | 60 | Breast | 0.003 | (0.001, 0.010) |
| Women | Rectal bleeding | Never smoker | 60 | Gyanecological | 0.002 | (0.001, 0.004) |
| Women | Rectal bleeding | Never smoker | 60 | Lower GI | 0.019 | (0.013, 0.028) |
| Women | Rectal bleeding | Never smoker | 60 | Haematological | 0.001 | (<0.001, 0.003) |
| Women | Rectal bleeding | Never smoker | 60 | Other cancers | 0.004 | (0.001, 0.011) |
| Women | Rectal bleeding | Never smoker | 60 | Death w/o cancer | 0.004 | (0.001, 0.012) |
| Women | Rectal bleeding | Never smoker | 70 | Breast | 0.005 | (0.003, 0.011) |
| Women | Rectal bleeding | Never smoker | 70 | Gyanecological | 0.004 | (0.001, 0.009) |
| Women | Rectal bleeding | Never smoker | 70 | Upper GI | 0.002 | (<0.001, 0.003) |
| Women | Rectal bleeding | Never smoker | 70 | Lower GI | 0.042 | (0.031, 0.057) |
| Women | Rectal bleeding | Never smoker | 70 | Urological | 0.001 | (<0.001, 0.002) |
| Women | Rectal bleeding | Never smoker | 70 | Haematological | 0.002 | (<0.001, 0.006) |
| Women | Rectal bleeding | Never smoker | 70 | Other cancers | 0.007 | (0.004, 0.014) |
| Women | Rectal bleeding | Never smoker | 70 | Death w/o cancer | 0.014 | (0.009, 0.024) |
| Women | Rectal bleeding | Never smoker | 80 | Breast | 0.006 | (0.003, 0.015) |
| Women | Rectal bleeding | Never smoker | 80 | Gyanecological | 0.003 | (0.001, 0.009) |
| Women | Rectal bleeding | Never smoker | 80 | Upper GI | 0.003 | (0.002, 0.005) |
| Women | Rectal bleeding | Never smoker | 80 | Lower GI | 0.057 | (0.044, 0.074) |
| Women | Rectal bleeding | Never smoker | 80 | Urological | 0.002 | (<0.001, 0.005) |
| Women | Rectal bleeding | Never smoker | 80 | Haematological | 0.003 | (0.001, 0.008) |
| Women | Rectal bleeding | Never smoker | 80 | Other cancers | 0.010 | (0.005, 0.018) |
| Women | Rectal bleeding | Never smoker | 80 | Death w/o cancer | 0.049 | (0.036, 0.067) |
| Women | Rectal bleeding | Never smoker | 90 | Breast | 0.007 | (0.004, 0.012) |
| Women | Rectal bleeding | Never smoker | 90 | Gyanecological | 0.002 | (<0.001, 0.006) |
| Women | Rectal bleeding | Never smoker | 90 | Upper GI | 0.003 | (0.001, 0.007) |
| Women | Rectal bleeding | Never smoker | 90 | Lower GI | 0.059 | (0.044, 0.079) |
| Women | Rectal bleeding | Never smoker | 90 | Urological | 0.002 | (<0.001, 0.004) |
| Women | Rectal bleeding | Never smoker | 90 | Haematological | 0.003 | (0.001, 0.007) |
| Women | Rectal bleeding | Never smoker | 90 | Other cancers | 0.012 | (0.006, 0.023) |
| Women | Rectal bleeding | Never smoker | 90 | Death w/o cancer | 0.147 | (0.128, 0.168) |
| Women | Rectal bleeding | Ever smoker | 30 | Gyanecological | 0.004 | (0.002, 0.012) |
| Women | Rectal bleeding | Ever smoker | 40 | Gyanecological | 0.002 | (0.001, 0.003) |
| Women | Rectal bleeding | Ever smoker | 40 | Lower GI | 0.002 | (0.001, 0.004) |
| Women | Rectal bleeding | Ever smoker | 40 | Other cancers | 0.001 | (<0.001, 0.002) |
| Women | Rectal bleeding | Ever smoker | 50 | Breast | 0.002 | (<0.001, 0.004) |
| Women | Rectal bleeding | Ever smoker | 50 | Gyanecological | 0.001 | (<0.001, 0.002) |
| Women | Rectal bleeding | Ever smoker | 50 | Lower GI | 0.008 | (0.004, 0.013) |
| Women | Rectal bleeding | Ever smoker | 50 | Other cancers | 0.003 | (0.001, 0.006) |
| Women | Rectal bleeding | Ever smoker | 50 | Death w/o cancer | 0.002 | (<0.001, 0.005) |
| Women | Rectal bleeding | Ever smoker | 60 | Breast | 0.004 | (0.002, 0.008) |
| Women | Rectal bleeding | Ever smoker | 60 | Gyanecological | 0.002 | (0.001, 0.004) |
| Women | Rectal bleeding | Ever smoker | 60 | Lung | 0.001 | (<0.001, 0.003) |
| Women | Rectal bleeding | Ever smoker | 60 | Upper GI | 0.001 | (<0.001, 0.002) |
| Women | Rectal bleeding | Ever smoker | 60 | Lower GI | 0.018 | (0.011, 0.028) |
| Women | Rectal bleeding | Ever smoker | 60 | Haematological | 0.001 | (<0.001, 0.003) |
| Women | Rectal bleeding | Ever smoker | 60 | Other cancers | 0.004 | (0.002, 0.011) |
| Women | Rectal bleeding | Ever smoker | 60 | Death w/o cancer | 0.004 | (0.002, 0.011) |
| Women | Rectal bleeding | Ever smoker | 70 | Breast | 0.006 | (0.002, 0.015) |
| Women | Rectal bleeding | Ever smoker | 70 | Gyanecological | 0.004 | (0.001, 0.010) |
| Women | Rectal bleeding | Ever smoker | 70 | Lung | 0.002 | (<0.001, 0.007) |
| Women | Rectal bleeding | Ever smoker | 70 | Upper GI | 0.003 | (0.001, 0.006) |
| Women | Rectal bleeding | Ever smoker | 70 | Lower GI | 0.040 | (0.028, 0.058) |
| Women | Rectal bleeding | Ever smoker | 70 | Urological | 0.002 | (<0.001, 0.004) |
| Women | Rectal bleeding | Ever smoker | 70 | Haematological | 0.003 | (0.001, 0.005) |
| Women | Rectal bleeding | Ever smoker | 70 | Other cancers | 0.007 | (0.004, 0.015) |
| Women | Rectal bleeding | Ever smoker | 70 | Death w/o cancer | 0.014 | (0.009, 0.021) |
| Women | Rectal bleeding | Ever smoker | 80 | Breast | 0.006 | (0.003, 0.014) |
| Women | Rectal bleeding | Ever smoker | 80 | Gyanecological | 0.003 | (0.002, 0.006) |
| Women | Rectal bleeding | Ever smoker | 80 | Lung | 0.003 | (0.001, 0.007) |
| Women | Rectal bleeding | Ever smoker | 80 | Upper GI | 0.004 | (0.001, 0.009) |
| Women | Rectal bleeding | Ever smoker | 80 | Lower GI | 0.056 | (0.041, 0.076) |
| Women | Rectal bleeding | Ever smoker | 80 | Urological | 0.002 | (<0.001, 0.006) |
| Women | Rectal bleeding | Ever smoker | 80 | Haematological | 0.003 | (0.001, 0.007) |
| Women | Rectal bleeding | Ever smoker | 80 | Other cancers | 0.010 | (0.005, 0.021) |
| Women | Rectal bleeding | Ever smoker | 80 | Death w/o cancer | 0.047 | (0.031, 0.070) |
| Women | Rectal bleeding | Ever smoker | 90 | Breast | 0.008 | (0.005, 0.014) |
| Women | Rectal bleeding | Ever smoker | 90 | Gyanecological | 0.002 | (0.001, 0.005) |
| Women | Rectal bleeding | Ever smoker | 90 | Lung | 0.003 | (0.001, 0.006) |
| Women | Rectal bleeding | Ever smoker | 90 | Upper GI | 0.003 | (0.002, 0.007) |
| Women | Rectal bleeding | Ever smoker | 90 | Lower GI | 0.059 | (0.043, 0.080) |
| Women | Rectal bleeding | Ever smoker | 90 | Urological | 0.003 | (0.001, 0.005) |
| Women | Rectal bleeding | Ever smoker | 90 | Haematological | 0.003 | (0.001, 0.007) |
| Women | Rectal bleeding | Ever smoker | 90 | Other cancers | 0.012 | (0.007, 0.022) |
| Women | Rectal bleeding | Ever smoker | 90 | Death w/o cancer | 0.146 | (0.124, 0.170) |
| Women | Weight loss | Never smoker | 30 | Gyanecological | 0.006 | (0.003, 0.011) |
| Women | Weight loss | Never smoker | 40 | Gyanecological | 0.003 | (0.001, 0.006) |
| Women | Weight loss | Never smoker | 40 | Other cancers | 0.001 | (<0.001, 0.003) |
| Women | Weight loss | Never smoker | 40 | Death w/o cancer | 0.002 | (<0.001, 0.004) |
| Women | Weight loss | Never smoker | 50 | Breast | 0.002 | (<0.001, 0.004) |
| Women | Weight loss | Never smoker | 50 | Gyanecological | 0.002 | (<0.001, 0.003) |
| Women | Weight loss | Never smoker | 50 | Upper GI | 0.002 | (<0.001, 0.004) |
| Women | Weight loss | Never smoker | 50 | Lower GI | 0.002 | (<0.001, 0.004) |
| Women | Weight loss | Never smoker | 50 | Haematological | 0.002 | (<0.001, 0.003) |
| Women | Weight loss | Never smoker | 50 | Other cancers | 0.002 | (<0.001, 0.006) |
| Women | Weight loss | Never smoker | 50 | Death w/o cancer | 0.004 | (0.002, 0.008) |
| Women | Weight loss | Never smoker | 60 | Breast | 0.003 | (0.001, 0.009) |
| Women | Weight loss | Never smoker | 60 | Gyanecological | 0.003 | (0.001, 0.006) |
| Women | Weight loss | Never smoker | 60 | Lung | 0.002 | (<0.001, 0.006) |
| Women | Weight loss | Never smoker | 60 | Upper GI | 0.005 | (0.002, 0.013) |
| Women | Weight loss | Never smoker | 60 | Lower GI | 0.004 | (0.001, 0.010) |
| Women | Weight loss | Never smoker | 60 | Urological | 0.001 | (<0.001, 0.002) |
| Women | Weight loss | Never smoker | 60 | Haematological | 0.003 | (0.001, 0.008) |
| Women | Weight loss | Never smoker | 60 | Other cancers | 0.004 | (0.001, 0.011) |
| Women | Weight loss | Never smoker | 60 | Death w/o cancer | 0.007 | (0.004, 0.012) |
| Women | Weight loss | Never smoker | 70 | Breast | 0.005 | (0.002, 0.010) |
| Women | Weight loss | Never smoker | 70 | Gyanecological | 0.005 | (0.002, 0.012) |
| Women | Weight loss | Never smoker | 70 | Lung | 0.004 | (0.001, 0.011) |
| Women | Weight loss | Never smoker | 70 | Upper GI | 0.011 | (0.005, 0.026) |
| Women | Weight loss | Never smoker | 70 | Lower GI | 0.007 | (0.004, 0.014) |
| Women | Weight loss | Never smoker | 70 | Urological | 0.002 | (0.001, 0.005) |
| Women | Weight loss | Never smoker | 70 | Haematological | 0.006 | (0.003, 0.014) |
| Women | Weight loss | Never smoker | 70 | Other cancers | 0.007 | (0.003, 0.014) |
| Women | Weight loss | Never smoker | 70 | Death w/o cancer | 0.026 | (0.019, 0.035) |
| Women | Weight loss | Never smoker | 80 | Breast | 0.007 | (0.003, 0.018) |
| Women | Weight loss | Never smoker | 80 | Gyanecological | 0.005 | (0.001, 0.015) |
| Women | Weight loss | Never smoker | 80 | Lung | 0.005 | (0.002, 0.014) |
| Women | Weight loss | Never smoker | 80 | Upper GI | 0.015 | (0.009, 0.026) |
| Women | Weight loss | Never smoker | 80 | Lower GI | 0.011 | (0.006, 0.020) |
| Women | Weight loss | Never smoker | 80 | Urological | 0.003 | (0.001, 0.009) |
| Women | Weight loss | Never smoker | 80 | Haematological | 0.008 | (0.004, 0.014) |
| Women | Weight loss | Never smoker | 80 | Other cancers | 0.010 | (0.005, 0.017) |
| Women | Weight loss | Never smoker | 80 | Death w/o cancer | 0.084 | (0.064, 0.110) |
| Women | Weight loss | Never smoker | 90 | Breast | 0.008 | (0.004, 0.014) |
| Women | Weight loss | Never smoker | 90 | Gyanecological | 0.003 | (0.001, 0.009) |
| Women | Weight loss | Never smoker | 90 | Lung | 0.004 | (0.002, 0.010) |
| Women | Weight loss | Never smoker | 90 | Upper GI | 0.015 | (0.008, 0.028) |
| Women | Weight loss | Never smoker | 90 | Lower GI | 0.011 | (0.007, 0.018) |
| Women | Weight loss | Never smoker | 90 | Urological | 0.003 | (0.001, 0.008) |
| Women | Weight loss | Never smoker | 90 | Haematological | 0.008 | (0.004, 0.015) |
| Women | Weight loss | Never smoker | 90 | Other cancers | 0.011 | (0.006, 0.022) |
| Women | Weight loss | Never smoker | 90 | Death w/o cancer | 0.248 | (0.224, 0.274) |
| Women | Weight loss | Ever smoker | 30 | Gyanecological | 0.006 | (0.003, 0.011) |
| Women | Weight loss | Ever smoker | 30 | Death w/o cancer | 0.001 | (<0.001, 0.002) |
| Women | Weight loss | Ever smoker | 40 | Gyanecological | 0.003 | (0.001, 0.007) |
| Women | Weight loss | Ever smoker | 40 | Other cancers | 0.001 | (<0.001, 0.002) |
| Women | Weight loss | Ever smoker | 40 | Death w/o cancer | 0.002 | (<0.001, 0.003) |
| Women | Weight loss | Ever smoker | 50 | Breast | 0.002 | (<0.001, 0.004) |
| Women | Weight loss | Ever smoker | 50 | Gyanecological | 0.002 | (<0.001, 0.003) |
| Women | Weight loss | Ever smoker | 50 | Lung | 0.003 | (0.002, 0.007) |
| Women | Weight loss | Ever smoker | 50 | Upper GI | 0.002 | (<0.001, 0.005) |
| Women | Weight loss | Ever smoker | 50 | Lower GI | 0.001 | (<0.001, 0.003) |
| Women | Weight loss | Ever smoker | 50 | Haematological | 0.001 | (<0.001, 0.002) |
| Women | Weight loss | Ever smoker | 50 | Other cancers | 0.003 | (0.001, 0.005) |
| Women | Weight loss | Ever smoker | 50 | Death w/o cancer | 0.004 | (0.002, 0.009) |
| Women | Weight loss | Ever smoker | 60 | Breast | 0.004 | (0.002, 0.007) |
| Women | Weight loss | Ever smoker | 60 | Gyanecological | 0.003 | (0.001, 0.008) |
| Women | Weight loss | Ever smoker | 60 | Lung | 0.010 | (0.005, 0.022) |
| Women | Weight loss | Ever smoker | 60 | Upper GI | 0.006 | (0.002, 0.012) |
| Women | Weight loss | Ever smoker | 60 | Lower GI | 0.003 | (0.001, 0.008) |
| Women | Weight loss | Ever smoker | 60 | Urological | 0.001 | (<0.001, 0.003) |
| Women | Weight loss | Ever smoker | 60 | Haematological | 0.003 | (0.001, 0.008) |
| Women | Weight loss | Ever smoker | 60 | Other cancers | 0.005 | (0.002, 0.011) |
| Women | Weight loss | Ever smoker | 60 | Death w/o cancer | 0.007 | (0.004, 0.012) |
| Women | Weight loss | Ever smoker | 70 | Breast | 0.005 | (0.002, 0.013) |
| Women | Weight loss | Ever smoker | 70 | Gyanecological | 0.006 | (0.003, 0.010) |
| Women | Weight loss | Ever smoker | 70 | Lung | 0.020 | (0.014, 0.028) |
| Women | Weight loss | Ever smoker | 70 | Upper GI | 0.013 | (0.008, 0.022) |
| Women | Weight loss | Ever smoker | 70 | Lower GI | 0.007 | (0.004, 0.012) |
| Women | Weight loss | Ever smoker | 70 | Urological | 0.003 | (0.001, 0.008) |
| Women | Weight loss | Ever smoker | 70 | Haematological | 0.006 | (0.002, 0.018) |
| Women | Weight loss | Ever smoker | 70 | Other cancers | 0.008 | (0.003, 0.016) |
| Women | Weight loss | Ever smoker | 70 | Death w/o cancer | 0.024 | (0.016, 0.036) |
| Women | Weight loss | Ever smoker | 80 | Breast | 0.007 | (0.003, 0.017) |
| Women | Weight loss | Ever smoker | 80 | Gyanecological | 0.005 | (0.003, 0.008) |
| Women | Weight loss | Ever smoker | 80 | Lung | 0.023 | (0.013, 0.040) |
| Women | Weight loss | Ever smoker | 80 | Upper GI | 0.019 | (0.012, 0.032) |
| Women | Weight loss | Ever smoker | 80 | Lower GI | 0.010 | (0.005, 0.020) |
| Women | Weight loss | Ever smoker | 80 | Urological | 0.004 | (0.002, 0.008) |
| Women | Weight loss | Ever smoker | 80 | Haematological | 0.007 | (0.004, 0.016) |
| Women | Weight loss | Ever smoker | 80 | Other cancers | 0.009 | (0.005, 0.017) |
| Women | Weight loss | Ever smoker | 80 | Death w/o cancer | 0.080 | (0.064, 0.101) |
| Women | Weight loss | Ever smoker | 90 | Breast | 0.008 | (0.004, 0.014) |
| Women | Weight loss | Ever smoker | 90 | Gyanecological | 0.003 | (0.001, 0.008) |
| Women | Weight loss | Ever smoker | 90 | Lung | 0.021 | (0.012, 0.038) |
| Women | Weight loss | Ever smoker | 90 | Upper GI | 0.019 | (0.009, 0.039) |
| Women | Weight loss | Ever smoker | 90 | Lower GI | 0.011 | (0.005, 0.022) |
| Women | Weight loss | Ever smoker | 90 | Urological | 0.004 | (<0.001, 0.000) |
| Women | Weight loss | Ever smoker | 90 | Haematological | 0.008 | (0.004, 0.018) |
| Women | Weight loss | Ever smoker | 90 | Other cancers | 0.011 | (0.007, 0.018) |
| Women | Weight loss | Ever smoker | 90 | Death w/o cancer | 0.239 | (0.209, 0.272) |
| Men | Abdominal pain | Never smoker | 40 | Death w/o cancer | 0.002 | (0.001, 0.003) |
| Men | Abdominal pain | Never smoker | 50 | Upper GI | 0.002 | (<0.001, 0.004) |
| Men | Abdominal pain | Never smoker | 50 | Lower GI | 0.003 | (0.002, 0.007) |
| Men | Abdominal pain | Never smoker | 50 | Urological | 0.001 | (<0.001, 0.002) |
| Men | Abdominal pain | Never smoker | 50 | Haematological | 0.001 | (<0.001, 0.003) |
| Men | Abdominal pain | Never smoker | 50 | Other cancers | 0.002 | (0.001, 0.004) |
| Men | Abdominal pain | Never smoker | 50 | Death w/o cancer | 0.003 | (0.002, 0.007) |
| Men | Abdominal pain | Never smoker | 60 | Upper GI | 0.006 | (0.002, 0.015) |
| Men | Abdominal pain | Never smoker | 60 | Lower GI | 0.008 | (0.004, 0.015) |
| Men | Abdominal pain | Never smoker | 60 | Urological | 0.003 | (0.001, 0.008) |
| Men | Abdominal pain | Never smoker | 60 | Prostate | 0.007 | (0.003, 0.015) |
| Men | Abdominal pain | Never smoker | 60 | Haematological | 0.003 | (0.001, 0.006) |
| Men | Abdominal pain | Never smoker | 60 | Other cancers | 0.003 | (0.001, 0.008) |
| Men | Abdominal pain | Never smoker | 60 | Death w/o cancer | 0.007 | (0.002, 0.019) |
| Men | Abdominal pain | Never smoker | 70 | Lung | 0.002 | (<0.001, 0.004) |
| Men | Abdominal pain | Never smoker | 70 | Upper GI | 0.011 | (0.006, 0.019) |
| Men | Abdominal pain | Never smoker | 70 | Lower GI | 0.019 | (0.011, 0.032) |
| Men | Abdominal pain | Never smoker | 70 | Urological | 0.005 | (0.002, 0.013) |
| Men | Abdominal pain | Never smoker | 70 | Prostate | 0.016 | (0.008, 0.030) |
| Men | Abdominal pain | Never smoker | 70 | Haematological | 0.006 | (0.003, 0.013) |
| Men | Abdominal pain | Never smoker | 70 | Other cancers | 0.006 | (0.003, 0.013) |
| Men | Abdominal pain | Never smoker | 70 | Death w/o cancer | 0.016 | (0.009, 0.030) |
| Men | Abdominal pain | Never smoker | 80 | Lung | 0.002 | (<0.001, 0.007) |
| Men | Abdominal pain | Never smoker | 80 | Upper GI | 0.014 | (0.008, 0.022) |
| Men | Abdominal pain | Never smoker | 80 | Lower GI | 0.024 | (0.016, 0.034) |
| Men | Abdominal pain | Never smoker | 80 | Urological | 0.006 | (0.002, 0.018) |
| Men | Abdominal pain | Never smoker | 80 | Prostate | 0.019 | (0.013, 0.030) |
| Men | Abdominal pain | Never smoker | 80 | Haematological | 0.007 | (0.004, 0.014) |
| Men | Abdominal pain | Never smoker | 80 | Other cancers | 0.007 | (0.004, 0.014) |
| Men | Abdominal pain | Never smoker | 80 | Death w/o cancer | 0.044 | (0.034, 0.057) |
| Men | Abdominal pain | Never smoker | 90 | Lung | 0.002 | (0.001, 0.006) |
| Men | Abdominal pain | Never smoker | 90 | Upper GI | 0.014 | (0.009, 0.021) |
| Men | Abdominal pain | Never smoker | 90 | Lower GI | 0.029 | (0.019, 0.042) |
| Men | Abdominal pain | Never smoker | 90 | Urological | 0.006 | (0.002, 0.015) |
| Men | Abdominal pain | Never smoker | 90 | Prostate | 0.022 | (0.015, 0.032) |
| Men | Abdominal pain | Never smoker | 90 | Haematological | 0.009 | (0.005, 0.019) |
| Men | Abdominal pain | Never smoker | 90 | Other cancers | 0.010 | (0.006, 0.017) |
| Men | Abdominal pain | Never smoker | 90 | Death w/o cancer | 0.129 | (0.111, 0.149) |
| Men | Abdominal pain | Ever smoker | 40 | Death w/o cancer | 0.002 | (<0.001, 0.004) |
| Men | Abdominal pain | Ever smoker | 50 | Upper GI | 0.003 | (0.001, 0.007) |
| Men | Abdominal pain | Ever smoker | 50 | Lower GI | 0.003 | (0.001, 0.007) |
| Men | Abdominal pain | Ever smoker | 50 | Urological | 0.001 | (<0.001, 0.003) |
| Men | Abdominal pain | Ever smoker | 50 | Haematological | 0.001 | (<0.001, 0.002) |
| Men | Abdominal pain | Ever smoker | 50 | Other cancers | 0.002 | (0.001, 0.005) |
| Men | Abdominal pain | Ever smoker | 50 | Death w/o cancer | 0.003 | (0.001, 0.009) |
| Men | Abdominal pain | Ever smoker | 60 | Lung | 0.003 | (0.001, 0.007) |
| Men | Abdominal pain | Ever smoker | 60 | Upper GI | 0.008 | (0.004, 0.017) |
| Men | Abdominal pain | Ever smoker | 60 | Lower GI | 0.009 | (0.004, 0.018) |
| Men | Abdominal pain | Ever smoker | 60 | Urological | 0.004 | (0.002, 0.007) |
| Men | Abdominal pain | Ever smoker | 60 | Prostate | 0.006 | (0.003, 0.012) |
| Men | Abdominal pain | Ever smoker | 60 | Haematological | 0.003 | (0.001, 0.007) |
| Men | Abdominal pain | Ever smoker | 60 | Other cancers | 0.004 | (0.001, 0.010) |
| Men | Abdominal pain | Ever smoker | 60 | Death w/o cancer | 0.007 | (0.003, 0.017) |
| Men | Abdominal pain | Ever smoker | 70 | Lung | 0.007 | (0.003, 0.016) |
| Men | Abdominal pain | Ever smoker | 70 | Upper GI | 0.014 | (0.010, 0.021) |
| Men | Abdominal pain | Ever smoker | 70 | Lower GI | 0.019 | (0.013, 0.028) |
| Men | Abdominal pain | Ever smoker | 70 | Urological | 0.007 | (0.002, 0.022) |
| Men | Abdominal pain | Ever smoker | 70 | Prostate | 0.014 | (0.009, 0.024) |
| Men | Abdominal pain | Ever smoker | 70 | Haematological | 0.006 | (0.002, 0.012) |
| Men | Abdominal pain | Ever smoker | 70 | Other cancers | 0.007 | (0.003, 0.014) |
| Men | Abdominal pain | Ever smoker | 70 | Death w/o cancer | 0.017 | (0.010, 0.027) |
| Men | Abdominal pain | Ever smoker | 80 | Lung | 0.009 | (0.004, 0.018) |
| Men | Abdominal pain | Ever smoker | 80 | Upper GI | 0.018 | (0.010, 0.033) |
| Men | Abdominal pain | Ever smoker | 80 | Lower GI | 0.026 | (0.016, 0.040) |
| Men | Abdominal pain | Ever smoker | 80 | Urological | 0.008 | (0.003, 0.017) |
| Men | Abdominal pain | Ever smoker | 80 | Prostate | 0.018 | (0.010, 0.031) |
| Men | Abdominal pain | Ever smoker | 80 | Haematological | 0.007 | (0.003, 0.016) |
| Men | Abdominal pain | Ever smoker | 80 | Other cancers | 0.009 | (0.004, 0.019) |
| Men | Abdominal pain | Ever smoker | 80 | Death w/o cancer | 0.047 | (0.037, 0.060) |
| Men | Abdominal pain | Ever smoker | 90 | Lung | 0.009 | (0.004, 0.019) |
| Men | Abdominal pain | Ever smoker | 90 | Upper GI | 0.019 | (0.011, 0.033) |
| Men | Abdominal pain | Ever smoker | 90 | Lower GI | 0.028 | (0.018, 0.046) |
| Men | Abdominal pain | Ever smoker | 90 | Urological | 0.008 | (0.004, 0.017) |
| Men | Abdominal pain | Ever smoker | 90 | Prostate | 0.019 | (0.012, 0.029) |
| Men | Abdominal pain | Ever smoker | 90 | Haematological | 0.009 | (0.006, 0.014) |
| Men | Abdominal pain | Ever smoker | 90 | Other cancers | 0.012 | (0.007, 0.020) |
| Men | Abdominal pain | Ever smoker | 90 | Death w/o cancer | 0.135 | (0.115, 0.157) |
| Men | Abdominal bloating | Never smoker | 40 | Death w/o cancer | 0.002 | (<0.001, 0.004) |
| Men | Abdominal bloating | Never smoker | 50 | Upper GI | 0.002 | (<0.001, 0.004) |
| Men | Abdominal bloating | Never smoker | 50 | Lower GI | 0.003 | (0.001, 0.008) |
| Men | Abdominal bloating | Never smoker | 50 | Haematological | 0.001 | (<0.001, 0.003) |
| Men | Abdominal bloating | Never smoker | 50 | Other cancers | 0.001 | (<0.001, 0.003) |
| Men | Abdominal bloating | Never smoker | 50 | Death w/o cancer | 0.002 | (0.001, 0.006) |
| Men | Abdominal bloating | Never smoker | 60 | Upper GI | 0.006 | (0.003, 0.012) |
| Men | Abdominal bloating | Never smoker | 60 | Lower GI | 0.008 | (0.004, 0.017) |
| Men | Abdominal bloating | Never smoker | 60 | Urological | 0.002 | (0.001, 0.003) |
| Men | Abdominal bloating | Never smoker | 60 | Prostate | 0.006 | (0.002, 0.020) |
| Men | Abdominal bloating | Never smoker | 60 | Haematological | 0.003 | (<0.001, 0.008) |
| Men | Abdominal bloating | Never smoker | 60 | Other cancers | 0.003 | (0.001, 0.006) |
| Men | Abdominal bloating | Never smoker | 60 | Death w/o cancer | 0.006 | (0.003, 0.013) |
| Men | Abdominal bloating | Never smoker | 70 | Lung | 0.002 | (<0.001, 0.004) |
| Men | Abdominal bloating | Never smoker | 70 | Upper GI | 0.011 | (0.006, 0.020) |
| Men | Abdominal bloating | Never smoker | 70 | Lower GI | 0.017 | (0.010, 0.029) |
| Men | Abdominal bloating | Never smoker | 70 | Urological | 0.003 | (0.001, 0.009) |
| Men | Abdominal bloating | Never smoker | 70 | Prostate | 0.014 | (0.007, 0.028) |
| Men | Abdominal bloating | Never smoker | 70 | Haematological | 0.005 | (0.002, 0.011) |
| Men | Abdominal bloating | Never smoker | 70 | Other cancers | 0.004 | (0.002, 0.011) |
| Men | Abdominal bloating | Never smoker | 70 | Death w/o cancer | 0.014 | (0.008, 0.026) |
| Men | Abdominal bloating | Never smoker | 80 | Lung | 0.002 | (<0.001, 0.005) |
| Men | Abdominal bloating | Never smoker | 80 | Upper GI | 0.013 | (0.008, 0.022) |
| Men | Abdominal bloating | Never smoker | 80 | Lower GI | 0.022 | (0.014, 0.034) |
| Men | Abdominal bloating | Never smoker | 80 | Urological | 0.004 | (0.001, 0.010) |
| Men | Abdominal bloating | Never smoker | 80 | Prostate | 0.018 | (0.012, 0.027) |
| Men | Abdominal bloating | Never smoker | 80 | Haematological | 0.007 | (0.004, 0.014) |
| Men | Abdominal bloating | Never smoker | 80 | Other cancers | 0.006 | (0.003, 0.013) |
| Men | Abdominal bloating | Never smoker | 80 | Death w/o cancer | 0.040 | (0.028, 0.057) |
| Men | Abdominal bloating | Never smoker | 90 | Lung | 0.002 | (<0.001, 0.005) |
| Men | Abdominal bloating | Never smoker | 90 | Upper GI | 0.013 | (0.008, 0.022) |
| Men | Abdominal bloating | Never smoker | 90 | Lower GI | 0.026 | (0.017, 0.040) |
| Men | Abdominal bloating | Never smoker | 90 | Urological | 0.004 | (0.002, 0.008) |
| Men | Abdominal bloating | Never smoker | 90 | Prostate | 0.019 | (0.012, 0.032) |
| Men | Abdominal bloating | Never smoker | 90 | Haematological | 0.009 | (0.004, 0.020) |
| Men | Abdominal bloating | Never smoker | 90 | Other cancers | 0.008 | (0.004, 0.014) |
| Men | Abdominal bloating | Never smoker | 90 | Death w/o cancer | 0.117 | (0.094, 0.144) |
| Men | Abdominal bloating | Ever smoker | 40 | Death w/o cancer | 0.002 | (<0.001, 0.004) |
| Men | Abdominal bloating | Ever smoker | 50 | Upper GI | 0.003 | (0.001, 0.008) |
| Men | Abdominal bloating | Ever smoker | 50 | Lower GI | 0.003 | (0.001, 0.008) |
| Men | Abdominal bloating | Ever smoker | 50 | Other cancers | 0.002 | (<0.001, 0.003) |
| Men | Abdominal bloating | Ever smoker | 50 | Death w/o cancer | 0.003 | (0.001, 0.007) |
| Men | Abdominal bloating | Ever smoker | 60 | Lung | 0.002 | (<0.001, 0.005) |
| Men | Abdominal bloating | Ever smoker | 60 | Upper GI | 0.008 | (0.004, 0.015) |
| Men | Abdominal bloating | Ever smoker | 60 | Lower GI | 0.008 | (0.004, 0.015) |
| Men | Abdominal bloating | Ever smoker | 60 | Urological | 0.002 | (0.001, 0.005) |
| Men | Abdominal bloating | Ever smoker | 60 | Prostate | 0.005 | (0.002, 0.012) |
| Men | Abdominal bloating | Ever smoker | 60 | Haematological | 0.003 | (<0.001, 0.009) |
| Men | Abdominal bloating | Ever smoker | 60 | Other cancers | 0.003 | (0.001, 0.008) |
| Men | Abdominal bloating | Ever smoker | 60 | Death w/o cancer | 0.006 | (0.003, 0.013) |
| Men | Abdominal bloating | Ever smoker | 70 | Lung | 0.005 | (0.003, 0.010) |
| Men | Abdominal bloating | Ever smoker | 70 | Upper GI | 0.015 | (0.008, 0.027) |
| Men | Abdominal bloating | Ever smoker | 70 | Lower GI | 0.018 | (0.012, 0.027) |
| Men | Abdominal bloating | Ever smoker | 70 | Urological | 0.004 | (0.002, 0.011) |
| Men | Abdominal bloating | Ever smoker | 70 | Prostate | 0.013 | (0.007, 0.023) |
| Men | Abdominal bloating | Ever smoker | 70 | Haematological | 0.005 | (0.002, 0.014) |
| Men | Abdominal bloating | Ever smoker | 70 | Other cancers | 0.005 | (0.003, 0.010) |
| Men | Abdominal bloating | Ever smoker | 70 | Death w/o cancer | 0.015 | (0.009, 0.025) |
| Men | Abdominal bloating | Ever smoker | 80 | Lung | 0.006 | (0.003, 0.012) |
| Men | Abdominal bloating | Ever smoker | 80 | Upper GI | 0.017 | (0.010, 0.030) |
| Men | Abdominal bloating | Ever smoker | 80 | Lower GI | 0.022 | (0.014, 0.036) |
| Men | Abdominal bloating | Ever smoker | 80 | Urological | 0.005 | (0.002, 0.011) |
| Men | Abdominal bloating | Ever smoker | 80 | Prostate | 0.017 | (0.009, 0.033) |
| Men | Abdominal bloating | Ever smoker | 80 | Haematological | 0.007 | (0.003, 0.017) |
| Men | Abdominal bloating | Ever smoker | 80 | Other cancers | 0.008 | (0.002, 0.026) |
| Men | Abdominal bloating | Ever smoker | 80 | Death w/o cancer | 0.045 | (0.034, 0.060) |
| Men | Abdominal bloating | Ever smoker | 90 | Lung | 0.007 | (0.003, 0.019) |
| Men | Abdominal bloating | Ever smoker | 90 | Upper GI | 0.018 | (0.009, 0.034) |
| Men | Abdominal bloating | Ever smoker | 90 | Lower GI | 0.026 | (0.016, 0.042) |
| Men | Abdominal bloating | Ever smoker | 90 | Urological | 0.005 | (0.002, 0.012) |
| Men | Abdominal bloating | Ever smoker | 90 | Prostate | 0.018 | (0.011, 0.029) |
| Men | Abdominal bloating | Ever smoker | 90 | Haematological | 0.009 | (0.005, 0.015) |
| Men | Abdominal bloating | Ever smoker | 90 | Other cancers | 0.009 | (0.005, 0.017) |
| Men | Abdominal bloating | Ever smoker | 90 | Death w/o cancer | 0.128 | (0.107, 0.152) |
| Men | Breast lump | Never smoker | 30 | Other cancers | 0.003 | (0.001, 0.009) |
| Men | Breast lump | Never smoker | 40 | Other cancers | 0.005 | (0.002, 0.009) |
| Men | Breast lump | Never smoker | 50 | Haematological | 0.001 | (<0.001, 0.003) |
| Men | Breast lump | Never smoker | 50 | Other cancers | 0.010 | (0.005, 0.018) |
| Men | Breast lump | Never smoker | 50 | Death w/o cancer | 0.002 | (<0.001, 0.003) |
| Men | Breast lump | Never smoker | 60 | Upper GI | 0.001 | (<0.001, 0.003) |
| Men | Breast lump | Never smoker | 60 | Prostate | 0.004 | (0.001, 0.010) |
| Men | Breast lump | Never smoker | 60 | Haematological | 0.002 | (0.001, 0.006) |
| Men | Breast lump | Never smoker | 60 | Other cancers | 0.016 | (0.009, 0.027) |
| Men | Breast lump | Never smoker | 60 | Death w/o cancer | 0.003 | (0.001, 0.008) |
| Men | Breast lump | Never smoker | 70 | Lung | 0.002 | (<0.001, 0.004) |
| Men | Breast lump | Never smoker | 70 | Upper GI | 0.003 | (0.001, 0.006) |
| Men | Breast lump | Never smoker | 70 | Urological | 0.001 | (<0.001, 0.003) |
| Men | Breast lump | Never smoker | 70 | Prostate | 0.009 | (0.003, 0.022) |
| Men | Breast lump | Never smoker | 70 | Haematological | 0.005 | (0.002, 0.010) |
| Men | Breast lump | Never smoker | 70 | Other cancers | 0.027 | (0.019, 0.038) |
| Men | Breast lump | Never smoker | 70 | Death w/o cancer | 0.009 | (0.005, 0.017) |
| Men | Breast lump | Never smoker | 80 | Lung | 0.002 | (<0.001, 0.006) |
| Men | Breast lump | Never smoker | 80 | Upper GI | 0.004 | (0.002, 0.010) |
| Men | Breast lump | Never smoker | 80 | Urological | 0.002 | (<0.001, 0.003) |
| Men | Breast lump | Never smoker | 80 | Prostate | 0.011 | (0.005, 0.026) |
| Men | Breast lump | Never smoker | 80 | Haematological | 0.007 | (0.003, 0.016) |
| Men | Breast lump | Never smoker | 80 | Other cancers | 0.039 | (0.027, 0.056) |
| Men | Breast lump | Never smoker | 80 | Death w/o cancer | 0.026 | (0.016, 0.041) |
| Men | Breast lump | Never smoker | 90 | Lung | 0.003 | (<0.001, 0.007) |
| Men | Breast lump | Never smoker | 90 | Upper GI | 0.004 | (0.002, 0.008) |
| Men | Breast lump | Never smoker | 90 | Urological | 0.002 | (<0.001, 0.004) |
| Men | Breast lump | Never smoker | 90 | Prostate | 0.012 | (0.005, 0.027) |
| Men | Breast lump | Never smoker | 90 | Haematological | 0.008 | (0.004, 0.015) |
| Men | Breast lump | Never smoker | 90 | Other cancers | 0.051 | (0.035, 0.072) |
| Men | Breast lump | Never smoker | 90 | Death w/o cancer | 0.075 | (0.049, 0.114) |
| Men | Breast lump | Ever smoker | 30 | Other cancers | 0.004 | (0.002, 0.007) |
| Men | Breast lump | Ever smoker | 40 | Other cancers | 0.005 | (0.003, 0.009) |
| Men | Breast lump | Ever smoker | 40 | Death w/o cancer | 0.001 | (<0.001, 0.002) |
| Men | Breast lump | Ever smoker | 50 | Haematological | 0.001 | (<0.001, 0.003) |
| Men | Breast lump | Ever smoker | 50 | Other cancers | 0.011 | (0.006, 0.021) |
| Men | Breast lump | Ever smoker | 50 | Death w/o cancer | 0.002 | (<0.001, 0.004) |
| Men | Breast lump | Ever smoker | 60 | Lung | 0.003 | (0.001, 0.009) |
| Men | Breast lump | Ever smoker | 60 | Upper GI | 0.002 | (<0.001, 0.006) |
| Men | Breast lump | Ever smoker | 60 | Prostate | 0.003 | (0.001, 0.009) |
| Men | Breast lump | Ever smoker | 60 | Haematological | 0.002 | (<0.001, 0.007) |
| Men | Breast lump | Ever smoker | 60 | Other cancers | 0.020 | (0.013, 0.030) |
| Men | Breast lump | Ever smoker | 60 | Death w/o cancer | 0.004 | (0.002, 0.008) |
| Men | Breast lump | Ever smoker | 70 | Lung | 0.007 | (0.003, 0.017) |
| Men | Breast lump | Ever smoker | 70 | Upper GI | 0.004 | (0.001, 0.014) |
| Men | Breast lump | Ever smoker | 70 | Urological | 0.002 | (<0.001, 0.004) |
| Men | Breast lump | Ever smoker | 70 | Prostate | 0.008 | (0.003, 0.020) |
| Men | Breast lump | Ever smoker | 70 | Haematological | 0.004 | (0.001, 0.012) |
| Men | Breast lump | Ever smoker | 70 | Other cancers | 0.033 | (0.021, 0.051) |
| Men | Breast lump | Ever smoker | 70 | Death w/o cancer | 0.010 | (0.005, 0.020) |
| Men | Breast lump | Ever smoker | 80 | Lung | 0.008 | (0.003, 0.025) |
| Men | Breast lump | Ever smoker | 80 | Upper GI | 0.005 | (0.002, 0.012) |
| Men | Breast lump | Ever smoker | 80 | Urological | 0.002 | (<0.001, 0.006) |
| Men | Breast lump | Ever smoker | 80 | Prostate | 0.010 | (0.004, 0.023) |
| Men | Breast lump | Ever smoker | 80 | Haematological | 0.006 | (0.002, 0.016) |
| Men | Breast lump | Ever smoker | 80 | Other cancers | 0.045 | (0.031, 0.064) |
| Men | Breast lump | Ever smoker | 80 | Death w/o cancer | 0.028 | (0.015, 0.049) |
| Men | Breast lump | Ever smoker | 90 | Lung | 0.010 | (0.004, 0.026) |
| Men | Breast lump | Ever smoker | 90 | Upper GI | 0.005 | (0.001, 0.017) |
| Men | Breast lump | Ever smoker | 90 | Urological | 0.002 | (<0.001, 0.005) |
| Men | Breast lump | Ever smoker | 90 | Prostate | 0.010 | (0.003, 0.032) |
| Men | Breast lump | Ever smoker | 90 | Haematological | 0.007 | (0.002, 0.021) |
| Men | Breast lump | Ever smoker | 90 | Other cancers | 0.061 | (0.039, 0.093) |
| Men | Breast lump | Ever smoker | 90 | Death w/o cancer | 0.081 | (0.051, 0.126) |
| Men | Change in bowel habit | Never smoker | 30 | Lower GI | 0.002 | (<0.001, 0.005) |
| Men | Change in bowel habit | Never smoker | 40 | Lower GI | 0.004 | (0.002, 0.008) |
| Men | Change in bowel habit | Never smoker | 40 | Other cancers | 0.001 | (<0.001, 0.002) |
| Men | Change in bowel habit | Never smoker | 50 | Lower GI | 0.014 | (0.009, 0.023) |
| Men | Change in bowel habit | Never smoker | 50 | Other cancers | 0.002 | (<0.001, 0.005) |
| Men | Change in bowel habit | Never smoker | 50 | Death w/o cancer | 0.002 | (<0.001, 0.003) |
| Men | Change in bowel habit | Never smoker | 60 | Upper GI | 0.002 | (0.001, 0.004) |
| Men | Change in bowel habit | Never smoker | 60 | Lower GI | 0.038 | (0.025, 0.057) |
| Men | Change in bowel habit | Never smoker | 60 | Urological | 0.002 | (<0.001, 0.004) |
| Men | Change in bowel habit | Never smoker | 60 | Prostate | 0.007 | (0.004, 0.014) |
| Men | Change in bowel habit | Never smoker | 60 | Haematological | 0.002 | (0.001, 0.004) |
| Men | Change in bowel habit | Never smoker | 60 | Other cancers | 0.003 | (0.001, 0.008) |
| Men | Change in bowel habit | Never smoker | 60 | Death w/o cancer | 0.003 | (0.001, 0.009) |
| Men | Change in bowel habit | Never smoker | 70 | Lung | 0.001 | (<0.001, 0.003) |
| Men | Change in bowel habit | Never smoker | 70 | Upper GI | 0.005 | (0.002, 0.011) |
| Men | Change in bowel habit | Never smoker | 70 | Lower GI | 0.080 | (0.061, 0.104) |
| Men | Change in bowel habit | Never smoker | 70 | Urological | 0.003 | (0.001, 0.008) |
| Men | Change in bowel habit | Never smoker | 70 | Prostate | 0.017 | (0.011, 0.026) |
| Men | Change in bowel habit | Never smoker | 70 | Haematological | 0.004 | (0.002, 0.009) |
| Men | Change in bowel habit | Never smoker | 70 | Other cancers | 0.006 | (0.003, 0.012) |
| Men | Change in bowel habit | Never smoker | 70 | Death w/o cancer | 0.009 | (0.004, 0.021) |
| Men | Change in bowel habit | Never smoker | 80 | Lung | 0.002 | (<0.001, 0.003) |
| Men | Change in bowel habit | Never smoker | 80 | Upper GI | 0.005 | (0.002, 0.012) |
| Men | Change in bowel habit | Never smoker | 80 | Lower GI | 0.107 | (0.083, 0.138) |
| Men | Change in bowel habit | Never smoker | 80 | Urological | 0.004 | (0.002, 0.008) |
| Men | Change in bowel habit | Never smoker | 80 | Prostate | 0.021 | (0.012, 0.036) |
| Men | Change in bowel habit | Never smoker | 80 | Haematological | 0.005 | (0.002, 0.015) |
| Men | Change in bowel habit | Never smoker | 80 | Other cancers | 0.008 | (0.004, 0.015) |
| Men | Change in bowel habit | Never smoker | 80 | Death w/o cancer | 0.022 | (0.013, 0.037) |
| Men | Change in bowel habit | Never smoker | 90 | Lung | 0.002 | (<0.001, 0.004) |
| Men | Change in bowel habit | Never smoker | 90 | Upper GI | 0.005 | (0.002, 0.013) |
| Men | Change in bowel habit | Never smoker | 90 | Lower GI | 0.125 | (0.099, 0.156) |
| Men | Change in bowel habit | Never smoker | 90 | Urological | 0.003 | (0.001, 0.009) |
| Men | Change in bowel habit | Never smoker | 90 | Prostate | 0.023 | (0.015, 0.036) |
| Men | Change in bowel habit | Never smoker | 90 | Haematological | 0.006 | (0.002, 0.016) |
| Men | Change in bowel habit | Never smoker | 90 | Other cancers | 0.010 | (0.004, 0.024) |
| Men | Change in bowel habit | Never smoker | 90 | Death w/o cancer | 0.062 | (0.047, 0.083) |
| Men | Change in bowel habit | Ever smoker | 30 | Lower GI | 0.002 | (0.001, 0.004) |
| Men | Change in bowel habit | Ever smoker | 40 | Lower GI | 0.003 | (0.001, 0.008) |
| Men | Change in bowel habit | Ever smoker | 40 | Other cancers | 0.001 | (<0.001, 0.002) |
| Men | Change in bowel habit | Ever smoker | 50 | Upper GI | 0.001 | (<0.001, 0.002) |
| Men | Change in bowel habit | Ever smoker | 50 | Lower GI | 0.016 | (0.010, 0.024) |
| Men | Change in bowel habit | Ever smoker | 50 | Other cancers | 0.003 | (0.001, 0.005) |
| Men | Change in bowel habit | Ever smoker | 50 | Death w/o cancer | 0.002 | (<0.001, 0.004) |
| Men | Change in bowel habit | Ever smoker | 60 | Lung | 0.003 | (0.001, 0.004) |
| Men | Change in bowel habit | Ever smoker | 60 | Upper GI | 0.003 | (0.001, 0.008) |
| Men | Change in bowel habit | Ever smoker | 60 | Lower GI | 0.039 | (0.029, 0.052) |
| Men | Change in bowel habit | Ever smoker | 60 | Urological | 0.002 | (<0.001, 0.005) |
| Men | Change in bowel habit | Ever smoker | 60 | Prostate | 0.007 | (0.003, 0.013) |
| Men | Change in bowel habit | Ever smoker | 60 | Haematological | 0.002 | (<0.001, 0.004) |
| Men | Change in bowel habit | Ever smoker | 60 | Other cancers | 0.004 | (0.002, 0.010) |
| Men | Change in bowel habit | Ever smoker | 60 | Death w/o cancer | 0.003 | (0.001, 0.010) |
| Men | Change in bowel habit | Ever smoker | 70 | Lung | 0.005 | (0.003, 0.010) |
| Men | Change in bowel habit | Ever smoker | 70 | Upper GI | 0.006 | (0.003, 0.014) |
| Men | Change in bowel habit | Ever smoker | 70 | Lower GI | 0.085 | (0.071, 0.101) |
| Men | Change in bowel habit | Ever smoker | 70 | Urological | 0.004 | (0.002, 0.009) |
| Men | Change in bowel habit | Ever smoker | 70 | Prostate | 0.016 | (0.008, 0.034) |
| Men | Change in bowel habit | Ever smoker | 70 | Haematological | 0.003 | (0.001, 0.009) |
| Men | Change in bowel habit | Ever smoker | 70 | Other cancers | 0.007 | (0.002, 0.020) |
| Men | Change in bowel habit | Ever smoker | 70 | Death w/o cancer | 0.008 | (0.004, 0.017) |
| Men | Change in bowel habit | Ever smoker | 80 | Lung | 0.007 | (0.003, 0.015) |
| Men | Change in bowel habit | Ever smoker | 80 | Upper GI | 0.007 | (0.004, 0.012) |
| Men | Change in bowel habit | Ever smoker | 80 | Lower GI | 0.111 | (0.092, 0.135) |
| Men | Change in bowel habit | Ever smoker | 80 | Urological | 0.005 | (0.002, 0.010) |
| Men | Change in bowel habit | Ever smoker | 80 | Prostate | 0.019 | (0.011, 0.030) |
| Men | Change in bowel habit | Ever smoker | 80 | Haematological | 0.005 | (0.002, 0.015) |
| Men | Change in bowel habit | Ever smoker | 80 | Other cancers | 0.010 | (0.004, 0.024) |
| Men | Change in bowel habit | Ever smoker | 80 | Death w/o cancer | 0.024 | (0.017, 0.032) |
| Men | Change in bowel habit | Ever smoker | 90 | Lung | 0.007 | (0.003, 0.020) |
| Men | Change in bowel habit | Ever smoker | 90 | Upper GI | 0.007 | (0.004, 0.014) |
| Men | Change in bowel habit | Ever smoker | 90 | Lower GI | 0.128 | (0.101, 0.161) |
| Men | Change in bowel habit | Ever smoker | 90 | Urological | 0.005 | (0.002, 0.010) |
| Men | Change in bowel habit | Ever smoker | 90 | Prostate | 0.020 | (0.014, 0.028) |
| Men | Change in bowel habit | Ever smoker | 90 | Haematological | 0.006 | (0.002, 0.019) |
| Men | Change in bowel habit | Ever smoker | 90 | Other cancers | 0.012 | (0.006, 0.025) |
| Men | Change in bowel habit | Ever smoker | 90 | Death w/o cancer | 0.068 | (0.054, 0.086) |
| Men | Dyspepsia | Never smoker | 40 | Death w/o cancer | 0.001 | (<0.001, 0.002) |
| Men | Dyspepsia | Never smoker | 50 | Upper GI | 0.003 | (0.001, 0.005) |
| Men | Dyspepsia | Never smoker | 50 | Other cancers | 0.001 | (<0.001, 0.003) |
| Men | Dyspepsia | Never smoker | 50 | Death w/o cancer | 0.002 | (0.001, 0.003) |
| Men | Dyspepsia | Never smoker | 60 | Upper GI | 0.007 | (0.003, 0.016) |
| Men | Dyspepsia | Never smoker | 60 | Lower GI | 0.002 | (0.001, 0.005) |
| Men | Dyspepsia | Never smoker | 60 | Urological | 0.001 | (<0.001, 0.003) |
| Men | Dyspepsia | Never smoker | 60 | Prostate | 0.005 | (0.002, 0.011) |
| Men | Dyspepsia | Never smoker | 60 | Haematological | 0.002 | (<0.001, 0.003) |
| Men | Dyspepsia | Never smoker | 60 | Other cancers | 0.003 | (0.001, 0.006) |
| Men | Dyspepsia | Never smoker | 60 | Death w/o cancer | 0.005 | (0.003, 0.009) |
| Men | Dyspepsia | Never smoker | 70 | Lung | 0.002 | (<0.001, 0.004) |
| Men | Dyspepsia | Never smoker | 70 | Upper GI | 0.014 | (0.009, 0.023) |
| Men | Dyspepsia | Never smoker | 70 | Lower GI | 0.005 | (0.003, 0.010) |
| Men | Dyspepsia | Never smoker | 70 | Urological | 0.002 | (<0.001, 0.006) |
| Men | Dyspepsia | Never smoker | 70 | Prostate | 0.011 | (0.006, 0.020) |
| Men | Dyspepsia | Never smoker | 70 | Haematological | 0.003 | (0.002, 0.007) |
| Men | Dyspepsia | Never smoker | 70 | Other cancers | 0.004 | (0.002, 0.009) |
| Men | Dyspepsia | Never smoker | 70 | Death w/o cancer | 0.012 | (0.006, 0.024) |
| Men | Dyspepsia | Never smoker | 80 | Lung | 0.002 | (<0.001, 0.005) |
| Men | Dyspepsia | Never smoker | 80 | Upper GI | 0.017 | (0.010, 0.030) |
| Men | Dyspepsia | Never smoker | 80 | Lower GI | 0.007 | (0.003, 0.015) |
| Men | Dyspepsia | Never smoker | 80 | Urological | 0.003 | (0.001, 0.006) |
| Men | Dyspepsia | Never smoker | 80 | Prostate | 0.015 | (0.009, 0.027) |
| Men | Dyspepsia | Never smoker | 80 | Haematological | 0.005 | (0.002, 0.013) |
| Men | Dyspepsia | Never smoker | 80 | Other cancers | 0.006 | (0.002, 0.017) |
| Men | Dyspepsia | Never smoker | 80 | Death w/o cancer | 0.035 | (0.025, 0.050) |
| Men | Dyspepsia | Never smoker | 90 | Lung | 0.002 | (0.001, 0.005) |
| Men | Dyspepsia | Never smoker | 90 | Upper GI | 0.019 | (0.012, 0.029) |
| Men | Dyspepsia | Never smoker | 90 | Lower GI | 0.007 | (0.004, 0.015) |
| Men | Dyspepsia | Never smoker | 90 | Urological | 0.003 | (0.001, 0.009) |
| Men | Dyspepsia | Never smoker | 90 | Prostate | 0.016 | (0.010, 0.024) |
| Men | Dyspepsia | Never smoker | 90 | Haematological | 0.006 | (0.002, 0.013) |
| Men | Dyspepsia | Never smoker | 90 | Other cancers | 0.008 | (0.004, 0.016) |
| Men | Dyspepsia | Never smoker | 90 | Death w/o cancer | 0.104 | (0.085, 0.128) |
| Men | Dyspepsia | Ever smoker | 40 | Death w/o cancer | 0.002 | (<0.001, 0.003) |
| Men | Dyspepsia | Ever smoker | 50 | Upper GI | 0.004 | (0.001, 0.009) |
| Men | Dyspepsia | Ever smoker | 50 | Other cancers | 0.002 | (0.001, 0.004) |
| Men | Dyspepsia | Ever smoker | 50 | Death w/o cancer | 0.002 | (<0.001, 0.006) |
| Men | Dyspepsia | Ever smoker | 60 | Lung | 0.003 | (0.001, 0.007) |
| Men | Dyspepsia | Ever smoker | 60 | Upper GI | 0.010 | (0.005, 0.018) |
| Men | Dyspepsia | Ever smoker | 60 | Lower GI | 0.003 | (0.001, 0.006) |
| Men | Dyspepsia | Ever smoker | 60 | Urological | 0.002 | (<0.001, 0.005) |
| Men | Dyspepsia | Ever smoker | 60 | Prostate | 0.004 | (0.002, 0.008) |
| Men | Dyspepsia | Ever smoker | 60 | Haematological | 0.002 | (<0.001, 0.004) |
| Men | Dyspepsia | Ever smoker | 60 | Other cancers | 0.003 | (0.001, 0.007) |
| Men | Dyspepsia | Ever smoker | 60 | Death w/o cancer | 0.006 | (0.003, 0.011) |
| Men | Dyspepsia | Ever smoker | 70 | Lung | 0.007 | (0.003, 0.013) |
| Men | Dyspepsia | Ever smoker | 70 | Upper GI | 0.019 | (0.012, 0.030) |
| Men | Dyspepsia | Ever smoker | 70 | Lower GI | 0.005 | (0.002, 0.015) |
| Men | Dyspepsia | Ever smoker | 70 | Urological | 0.003 | (0.002, 0.007) |
| Men | Dyspepsia | Ever smoker | 70 | Prostate | 0.010 | (0.005, 0.022) |
| Men | Dyspepsia | Ever smoker | 70 | Haematological | 0.003 | (0.001, 0.008) |
| Men | Dyspepsia | Ever smoker | 70 | Other cancers | 0.005 | (0.002, 0.014) |
| Men | Dyspepsia | Ever smoker | 70 | Death w/o cancer | 0.014 | (0.008, 0.024) |
| Men | Dyspepsia | Ever smoker | 80 | Lung | 0.008 | (0.004, 0.017) |
| Men | Dyspepsia | Ever smoker | 80 | Upper GI | 0.023 | (0.013, 0.039) |
| Men | Dyspepsia | Ever smoker | 80 | Lower GI | 0.007 | (0.004, 0.013) |
| Men | Dyspepsia | Ever smoker | 80 | Urological | 0.004 | (0.002, 0.012) |
| Men | Dyspepsia | Ever smoker | 80 | Prostate | 0.012 | (0.007, 0.020) |
| Men | Dyspepsia | Ever smoker | 80 | Haematological | 0.005 | (0.002, 0.014) |
| Men | Dyspepsia | Ever smoker | 80 | Other cancers | 0.007 | (0.003, 0.019) |
| Men | Dyspepsia | Ever smoker | 80 | Death w/o cancer | 0.038 | (0.025, 0.056) |
| Men | Dyspepsia | Ever smoker | 90 | Lung | 0.009 | (0.004, 0.024) |
| Men | Dyspepsia | Ever smoker | 90 | Upper GI | 0.024 | (0.014, 0.039) |
| Men | Dyspepsia | Ever smoker | 90 | Lower GI | 0.008 | (0.004, 0.018) |
| Men | Dyspepsia | Ever smoker | 90 | Urological | 0.004 | (0.002, 0.012) |
| Men | Dyspepsia | Ever smoker | 90 | Prostate | 0.014 | (0.007, 0.027) |
| Men | Dyspepsia | Ever smoker | 90 | Haematological | 0.005 | (0.002, 0.012) |
| Men | Dyspepsia | Ever smoker | 90 | Other cancers | 0.010 | (0.005, 0.019) |
| Men | Dyspepsia | Ever smoker | 90 | Death w/o cancer | 0.111 | (0.091, 0.136) |
| Men | Dysphagia | Never smoker | 30 | Other cancers | 0.001 | (<0.001, 0.003) |
| Men | Dysphagia | Never smoker | 30 | Death w/o cancer | 0.002 | (0.001, 0.004) |
| Men | Dysphagia | Never smoker | 40 | Upper GI | 0.003 | (0.001, 0.008) |
| Men | Dysphagia | Never smoker | 40 | Other cancers | 0.002 | (<0.001, 0.004) |
| Men | Dysphagia | Never smoker | 40 | Death w/o cancer | 0.005 | (0.002, 0.009) |
| Men | Dysphagia | Never smoker | 50 | Upper GI | 0.014 | (0.008, 0.025) |
| Men | Dysphagia | Never smoker | 50 | Haematological | 0.001 | (<0.001, 0.002) |
| Men | Dysphagia | Never smoker | 50 | Other cancers | 0.003 | (0.001, 0.010) |
| Men | Dysphagia | Never smoker | 50 | Death w/o cancer | 0.008 | (0.005, 0.014) |
| Men | Dysphagia | Never smoker | 60 | Lung | 0.001 | (<0.001, 0.003) |
| Men | Dysphagia | Never smoker | 60 | Upper GI | 0.042 | (0.032, 0.054) |
| Men | Dysphagia | Never smoker | 60 | Lower GI | 0.002 | (<0.001, 0.006) |
| Men | Dysphagia | Never smoker | 60 | Urological | 0.001 | (<0.001, 0.002) |
| Men | Dysphagia | Never smoker | 60 | Prostate | 0.003 | (0.001, 0.010) |
| Men | Dysphagia | Never smoker | 60 | Haematological | 0.002 | (0.001, 0.004) |
| Men | Dysphagia | Never smoker | 60 | Other cancers | 0.006 | (0.003, 0.012) |
| Men | Dysphagia | Never smoker | 60 | Death w/o cancer | 0.017 | (0.011, 0.028) |
| Men | Dysphagia | Never smoker | 70 | Lung | 0.003 | (<0.001, 0.007) |
| Men | Dysphagia | Never smoker | 70 | Upper GI | 0.080 | (0.059, 0.108) |
| Men | Dysphagia | Never smoker | 70 | Lower GI | 0.005 | (0.002, 0.010) |
| Men | Dysphagia | Never smoker | 70 | Urological | 0.002 | (<0.001, 0.004) |
| Men | Dysphagia | Never smoker | 70 | Prostate | 0.008 | (0.003, 0.020) |
| Men | Dysphagia | Never smoker | 70 | Haematological | 0.005 | (0.002, 0.013) |
| Men | Dysphagia | Never smoker | 70 | Other cancers | 0.010 | (0.006, 0.017) |
| Men | Dysphagia | Never smoker | 70 | Death w/o cancer | 0.041 | (0.030, 0.055) |
| Men | Dysphagia | Never smoker | 80 | Lung | 0.003 | (0.001, 0.010) |
| Men | Dysphagia | Never smoker | 80 | Upper GI | 0.093 | (0.075, 0.114) |
| Men | Dysphagia | Never smoker | 80 | Lower GI | 0.006 | (0.003, 0.014) |
| Men | Dysphagia | Never smoker | 80 | Urological | 0.002 | (<0.001, 0.005) |
| Men | Dysphagia | Never smoker | 80 | Prostate | 0.010 | (0.005, 0.019) |
| Men | Dysphagia | Never smoker | 80 | Haematological | 0.006 | (0.002, 0.015) |
| Men | Dysphagia | Never smoker | 80 | Other cancers | 0.013 | (0.007, 0.025) |
| Men | Dysphagia | Never smoker | 80 | Death w/o cancer | 0.114 | (0.091, 0.142) |
| Men | Dysphagia | Never smoker | 90 | Lung | 0.003 | (0.001, 0.010) |
| Men | Dysphagia | Never smoker | 90 | Upper GI | 0.095 | (0.073, 0.124) |
| Men | Dysphagia | Never smoker | 90 | Lower GI | 0.006 | (0.002, 0.017) |
| Men | Dysphagia | Never smoker | 90 | Urological | 0.002 | (<0.001, 0.004) |
| Men | Dysphagia | Never smoker | 90 | Prostate | 0.010 | (0.005, 0.020) |
| Men | Dysphagia | Never smoker | 90 | Haematological | 0.006 | (0.002, 0.019) |
| Men | Dysphagia | Never smoker | 90 | Other cancers | 0.015 | (0.010, 0.022) |
| Men | Dysphagia | Never smoker | 90 | Death w/o cancer | 0.299 | (0.264, 0.337) |
| Men | Dysphagia | Ever smoker | 30 | Other cancers | 0.002 | (<0.001, 0.003) |
| Men | Dysphagia | Ever smoker | 30 | Death w/o cancer | 0.002 | (0.001, 0.005) |
| Men | Dysphagia | Ever smoker | 40 | Upper GI | 0.005 | (0.002, 0.009) |
| Men | Dysphagia | Ever smoker | 40 | Other cancers | 0.002 | (<0.001, 0.005) |
| Men | Dysphagia | Ever smoker | 40 | Death w/o cancer | 0.005 | (0.003, 0.009) |
| Men | Dysphagia | Ever smoker | 50 | Lung | 0.001 | (<0.001, 0.003) |
| Men | Dysphagia | Ever smoker | 50 | Upper GI | 0.020 | (0.011, 0.036) |
| Men | Dysphagia | Ever smoker | 50 | Haematological | 0.001 | (<0.001, 0.002) |
| Men | Dysphagia | Ever smoker | 50 | Other cancers | 0.004 | (0.002, 0.012) |
| Men | Dysphagia | Ever smoker | 50 | Death w/o cancer | 0.009 | (0.005, 0.016) |
| Men | Dysphagia | Ever smoker | 60 | Lung | 0.005 | (<0.001, 0.000) |
| Men | Dysphagia | Ever smoker | 60 | Upper GI | 0.054 | (0.038, 0.076) |
| Men | Dysphagia | Ever smoker | 60 | Lower GI | 0.002 | (0.001, 0.005) |
| Men | Dysphagia | Ever smoker | 60 | Urological | 0.001 | (<0.001, 0.003) |
| Men | Dysphagia | Ever smoker | 60 | Prostate | 0.003 | (0.001, 0.008) |
| Men | Dysphagia | Ever smoker | 60 | Haematological | 0.002 | (0.001, 0.005) |
| Men | Dysphagia | Ever smoker | 60 | Other cancers | 0.007 | (0.004, 0.014) |
| Men | Dysphagia | Ever smoker | 60 | Death w/o cancer | 0.019 | (0.013, 0.029) |
| Men | Dysphagia | Ever smoker | 70 | Lung | 0.010 | (0.005, 0.020) |
| Men | Dysphagia | Ever smoker | 70 | Upper GI | 0.103 | (0.088, 0.120) |
| Men | Dysphagia | Ever smoker | 70 | Lower GI | 0.005 | (0.002, 0.015) |
| Men | Dysphagia | Ever smoker | 70 | Urological | 0.002 | (<0.001, 0.006) |
| Men | Dysphagia | Ever smoker | 70 | Prostate | 0.007 | (0.003, 0.016) |
| Men | Dysphagia | Ever smoker | 70 | Haematological | 0.004 | (0.002, 0.012) |
| Men | Dysphagia | Ever smoker | 70 | Other cancers | 0.012 | (0.007, 0.019) |
| Men | Dysphagia | Ever smoker | 70 | Death w/o cancer | 0.043 | (0.032, 0.059) |
| Men | Dysphagia | Ever smoker | 80 | Lung | 0.013 | (0.007, 0.025) |
| Men | Dysphagia | Ever smoker | 80 | Upper GI | 0.124 | (0.095, 0.160) |
| Men | Dysphagia | Ever smoker | 80 | Lower GI | 0.006 | (0.003, 0.012) |
| Men | Dysphagia | Ever smoker | 80 | Urological | 0.002 | (<0.001, 0.006) |
| Men | Dysphagia | Ever smoker | 80 | Prostate | 0.008 | (0.003, 0.023) |
| Men | Dysphagia | Ever smoker | 80 | Haematological | 0.006 | (0.002, 0.015) |
| Men | Dysphagia | Ever smoker | 80 | Other cancers | 0.015 | (0.009, 0.025) |
| Men | Dysphagia | Ever smoker | 80 | Death w/o cancer | 0.118 | (0.095, 0.145) |
| Men | Dysphagia | Ever smoker | 90 | Lung | 0.012 | (0.006, 0.026) |
| Men | Dysphagia | Ever smoker | 90 | Upper GI | 0.123 | (0.100, 0.151) |
| Men | Dysphagia | Ever smoker | 90 | Lower GI | 0.006 | (0.003, 0.012) |
| Men | Dysphagia | Ever smoker | 90 | Urological | 0.002 | (<0.001, 0.006) |
| Men | Dysphagia | Ever smoker | 90 | Prostate | 0.008 | (0.004, 0.015) |
| Men | Dysphagia | Ever smoker | 90 | Haematological | 0.007 | (0.002, 0.020) |
| Men | Dysphagia | Ever smoker | 90 | Other cancers | 0.018 | (0.011, 0.031) |
| Men | Dysphagia | Ever smoker | 90 | Death w/o cancer | 0.311 | (0.281, 0.343) |
| Men | Dyspnoea | Never smoker | 30 | Death w/o cancer | 0.001 | (<0.001, 0.003) |
| Men | Dyspnoea | Never smoker | 40 | Death w/o cancer | 0.003 | (0.001, 0.007) |
| Men | Dyspnoea | Never smoker | 50 | Other cancers | 0.001 | (<0.001, 0.003) |
| Men | Dyspnoea | Never smoker | 50 | Death w/o cancer | 0.006 | (0.003, 0.012) |
| Men | Dyspnoea | Never smoker | 60 | Lung | 0.002 | (<0.001, 0.005) |
| Men | Dyspnoea | Never smoker | 60 | Upper GI | 0.002 | (<0.001, 0.003) |
| Men | Dyspnoea | Never smoker | 60 | Lower GI | 0.002 | (0.001, 0.004) |
| Men | Dyspnoea | Never smoker | 60 | Urological | 0.001 | (<0.001, 0.002) |
| Men | Dyspnoea | Never smoker | 60 | Prostate | 0.003 | (0.001, 0.008) |
| Men | Dyspnoea | Never smoker | 60 | Haematological | 0.002 | (0.001, 0.005) |
| Men | Dyspnoea | Never smoker | 60 | Other cancers | 0.003 | (0.001, 0.005) |
| Men | Dyspnoea | Never smoker | 60 | Death w/o cancer | 0.013 | (0.007, 0.022) |
| Men | Dyspnoea | Never smoker | 70 | Lung | 0.005 | (0.002, 0.013) |
| Men | Dyspnoea | Never smoker | 70 | Upper GI | 0.003 | (0.001, 0.008) |
| Men | Dyspnoea | Never smoker | 70 | Lower GI | 0.005 | (0.002, 0.010) |
| Men | Dyspnoea | Never smoker | 70 | Urological | 0.003 | (0.001, 0.006) |
| Men | Dyspnoea | Never smoker | 70 | Prostate | 0.008 | (0.004, 0.018) |
| Men | Dyspnoea | Never smoker | 70 | Haematological | 0.004 | (0.002, 0.010) |
| Men | Dyspnoea | Never smoker | 70 | Other cancers | 0.004 | (0.002, 0.009) |
| Men | Dyspnoea | Never smoker | 70 | Death w/o cancer | 0.030 | (0.021, 0.042) |
| Men | Dyspnoea | Never smoker | 80 | Lung | 0.006 | (0.003, 0.013) |
| Men | Dyspnoea | Never smoker | 80 | Upper GI | 0.003 | (0.001, 0.009) |
| Men | Dyspnoea | Never smoker | 80 | Lower GI | 0.006 | (0.003, 0.013) |
| Men | Dyspnoea | Never smoker | 80 | Urological | 0.003 | (0.001, 0.006) |
| Men | Dyspnoea | Never smoker | 80 | Prostate | 0.011 | (0.006, 0.018) |
| Men | Dyspnoea | Never smoker | 80 | Haematological | 0.006 | (0.003, 0.013) |
| Men | Dyspnoea | Never smoker | 80 | Other cancers | 0.006 | (0.003, 0.012) |
| Men | Dyspnoea | Never smoker | 80 | Death w/o cancer | 0.087 | (0.070, 0.107) |
| Men | Dyspnoea | Never smoker | 90 | Lung | 0.007 | (0.003, 0.015) |
| Men | Dyspnoea | Never smoker | 90 | Upper GI | 0.004 | (0.001, 0.009) |
| Men | Dyspnoea | Never smoker | 90 | Lower GI | 0.007 | (0.003, 0.013) |
| Men | Dyspnoea | Never smoker | 90 | Urological | 0.003 | (0.002, 0.007) |
| Men | Dyspnoea | Never smoker | 90 | Prostate | 0.011 | (0.006, 0.021) |
| Men | Dyspnoea | Never smoker | 90 | Haematological | 0.006 | (0.003, 0.012) |
| Men | Dyspnoea | Never smoker | 90 | Other cancers | 0.007 | (0.003, 0.017) |
| Men | Dyspnoea | Never smoker | 90 | Death w/o cancer | 0.239 | (0.212, 0.267) |
| Men | Dyspnoea | Ever smoker | 30 | Death w/o cancer | 0.002 | (<0.001, 0.004) |
| Men | Dyspnoea | Ever smoker | 40 | Death w/o cancer | 0.004 | (0.001, 0.011) |
| Men | Dyspnoea | Ever smoker | 50 | Lung | 0.003 | (<0.001, 0.000) |
| Men | Dyspnoea | Ever smoker | 50 | Other cancers | 0.002 | (0.001, 0.003) |
| Men | Dyspnoea | Ever smoker | 50 | Death w/o cancer | 0.006 | (0.003, 0.014) |
| Men | Dyspnoea | Ever smoker | 60 | Lung | 0.009 | (0.004, 0.018) |
| Men | Dyspnoea | Ever smoker | 60 | Upper GI | 0.002 | (0.001, 0.005) |
| Men | Dyspnoea | Ever smoker | 60 | Lower GI | 0.002 | (<0.001, 0.005) |
| Men | Dyspnoea | Ever smoker | 60 | Urological | 0.002 | (<0.001, 0.004) |
| Men | Dyspnoea | Ever smoker | 60 | Prostate | 0.003 | (0.001, 0.008) |
| Men | Dyspnoea | Ever smoker | 60 | Haematological | 0.002 | (<0.001, 0.005) |
| Men | Dyspnoea | Ever smoker | 60 | Other cancers | 0.003 | (0.001, 0.007) |
| Men | Dyspnoea | Ever smoker | 60 | Death w/o cancer | 0.013 | (0.007, 0.025) |
| Men | Dyspnoea | Ever smoker | 70 | Lung | 0.019 | (0.013, 0.030) |
| Men | Dyspnoea | Ever smoker | 70 | Upper GI | 0.004 | (0.001, 0.011) |
| Men | Dyspnoea | Ever smoker | 70 | Lower GI | 0.005 | (0.002, 0.011) |
| Men | Dyspnoea | Ever smoker | 70 | Urological | 0.004 | (0.002, 0.009) |
| Men | Dyspnoea | Ever smoker | 70 | Prostate | 0.007 | (0.004, 0.016) |
| Men | Dyspnoea | Ever smoker | 70 | Haematological | 0.004 | (0.002, 0.009) |
| Men | Dyspnoea | Ever smoker | 70 | Other cancers | 0.005 | (0.002, 0.014) |
| Men | Dyspnoea | Ever smoker | 70 | Death w/o cancer | 0.033 | (0.021, 0.050) |
| Men | Dyspnoea | Ever smoker | 80 | Lung | 0.024 | (0.015, 0.039) |
| Men | Dyspnoea | Ever smoker | 80 | Upper GI | 0.005 | (0.001, 0.016) |
| Men | Dyspnoea | Ever smoker | 80 | Lower GI | 0.006 | (0.003, 0.012) |
| Men | Dyspnoea | Ever smoker | 80 | Urological | 0.004 | (0.001, 0.011) |
| Men | Dyspnoea | Ever smoker | 80 | Prostate | 0.009 | (0.004, 0.022) |
| Men | Dyspnoea | Ever smoker | 80 | Haematological | 0.006 | (0.002, 0.013) |
| Men | Dyspnoea | Ever smoker | 80 | Other cancers | 0.007 | (0.003, 0.016) |
| Men | Dyspnoea | Ever smoker | 80 | Death w/o cancer | 0.093 | (0.077, 0.112) |
| Men | Dyspnoea | Ever smoker | 90 | Lung | 0.027 | (0.016, 0.043) |
| Men | Dyspnoea | Ever smoker | 90 | Upper GI | 0.005 | (0.002, 0.013) |
| Men | Dyspnoea | Ever smoker | 90 | Lower GI | 0.007 | (0.004, 0.013) |
| Men | Dyspnoea | Ever smoker | 90 | Urological | 0.004 | (0.002, 0.010) |
| Men | Dyspnoea | Ever smoker | 90 | Prostate | 0.010 | (0.005, 0.019) |
| Men | Dyspnoea | Ever smoker | 90 | Haematological | 0.007 | (0.003, 0.013) |
| Men | Dyspnoea | Ever smoker | 90 | Other cancers | 0.009 | (0.005, 0.015) |
| Men | Dyspnoea | Ever smoker | 90 | Death w/o cancer | 0.251 | (0.227, 0.275) |
| Men | Fatigue | Never smoker | 40 | Other cancers | 0.001 | (<0.001, 0.002) |
| Men | Fatigue | Never smoker | 40 | Death w/o cancer | 0.002 | (<0.001, 0.005) |
| Men | Fatigue | Never smoker | 50 | Lower GI | 0.001 | (<0.001, 0.003) |
| Men | Fatigue | Never smoker | 50 | Haematological | 0.001 | (<0.001, 0.003) |
| Men | Fatigue | Never smoker | 50 | Other cancers | 0.002 | (<0.001, 0.004) |
| Men | Fatigue | Never smoker | 50 | Death w/o cancer | 0.004 | (0.002, 0.010) |
| Men | Fatigue | Never smoker | 60 | Lung | 0.002 | (<0.001, 0.003) |
| Men | Fatigue | Never smoker | 60 | Upper GI | 0.002 | (<0.001, 0.005) |
| Men | Fatigue | Never smoker | 60 | Lower GI | 0.003 | (0.001, 0.008) |
| Men | Fatigue | Never smoker | 60 | Urological | 0.001 | (<0.001, 0.003) |
| Men | Fatigue | Never smoker | 60 | Prostate | 0.006 | (0.003, 0.012) |
| Men | Fatigue | Never smoker | 60 | Haematological | 0.003 | (0.001, 0.008) |
| Men | Fatigue | Never smoker | 60 | Other cancers | 0.003 | (0.001, 0.009) |
| Men | Fatigue | Never smoker | 60 | Death w/o cancer | 0.008 | (0.005, 0.014) |
| Men | Fatigue | Never smoker | 70 | Lung | 0.003 | (0.001, 0.008) |
| Men | Fatigue | Never smoker | 70 | Upper GI | 0.004 | (0.002, 0.010) |
| Men | Fatigue | Never smoker | 70 | Lower GI | 0.007 | (0.004, 0.012) |
| Men | Fatigue | Never smoker | 70 | Urological | 0.003 | (0.001, 0.007) |
| Men | Fatigue | Never smoker | 70 | Prostate | 0.015 | (0.010, 0.023) |
| Men | Fatigue | Never smoker | 70 | Haematological | 0.006 | (0.003, 0.012) |
| Men | Fatigue | Never smoker | 70 | Other cancers | 0.005 | (0.002, 0.012) |
| Men | Fatigue | Never smoker | 70 | Death w/o cancer | 0.021 | (0.014, 0.030) |
| Men | Fatigue | Never smoker | 80 | Lung | 0.004 | (0.002, 0.007) |
| Men | Fatigue | Never smoker | 80 | Upper GI | 0.005 | (0.002, 0.015) |
| Men | Fatigue | Never smoker | 80 | Lower GI | 0.008 | (0.004, 0.015) |
| Men | Fatigue | Never smoker | 80 | Urological | 0.003 | (0.001, 0.009) |
| Men | Fatigue | Never smoker | 80 | Prostate | 0.018 | (0.011, 0.029) |
| Men | Fatigue | Never smoker | 80 | Haematological | 0.008 | (0.005, 0.013) |
| Men | Fatigue | Never smoker | 80 | Other cancers | 0.007 | (0.003, 0.016) |
| Men | Fatigue | Never smoker | 80 | Death w/o cancer | 0.059 | (0.044, 0.079) |
| Men | Fatigue | Never smoker | 90 | Lung | 0.004 | (0.001, 0.010) |
| Men | Fatigue | Never smoker | 90 | Upper GI | 0.005 | (0.003, 0.009) |
| Men | Fatigue | Never smoker | 90 | Lower GI | 0.010 | (0.005, 0.018) |
| Men | Fatigue | Never smoker | 90 | Urological | 0.003 | (0.002, 0.007) |
| Men | Fatigue | Never smoker | 90 | Prostate | 0.019 | (0.013, 0.029) |
| Men | Fatigue | Never smoker | 90 | Haematological | 0.009 | (0.005, 0.016) |
| Men | Fatigue | Never smoker | 90 | Other cancers | 0.010 | (0.005, 0.017) |
| Men | Fatigue | Never smoker | 90 | Death w/o cancer | 0.167 | (0.140, 0.198) |
| Men | Fatigue | Ever smoker | 30 | Death w/o cancer | 0.001 | (<0.001, 0.003) |
| Men | Fatigue | Ever smoker | 40 | Other cancers | 0.001 | (<0.001, 0.002) |
| Men | Fatigue | Ever smoker | 40 | Death w/o cancer | 0.002 | (<0.001, 0.006) |
| Men | Fatigue | Ever smoker | 50 | Lung | 0.002 | (<0.001, 0.003) |
| Men | Fatigue | Ever smoker | 50 | Upper GI | 0.001 | (<0.001, 0.002) |
| Men | Fatigue | Ever smoker | 50 | Lower GI | 0.001 | (<0.001, 0.002) |
| Men | Fatigue | Ever smoker | 50 | Haematological | 0.001 | (<0.001, 0.003) |
| Men | Fatigue | Ever smoker | 50 | Other cancers | 0.002 | (0.001, 0.004) |
| Men | Fatigue | Ever smoker | 50 | Death w/o cancer | 0.004 | (0.002, 0.010) |
| Men | Fatigue | Ever smoker | 60 | Lung | 0.005 | (0.002, 0.012) |
| Men | Fatigue | Ever smoker | 60 | Upper GI | 0.003 | (0.001, 0.005) |
| Men | Fatigue | Ever smoker | 60 | Lower GI | 0.003 | (0.001, 0.007) |
| Men | Fatigue | Ever smoker | 60 | Urological | 0.002 | (0.001, 0.003) |
| Men | Fatigue | Ever smoker | 60 | Prostate | 0.006 | (0.003, 0.013) |
| Men | Fatigue | Ever smoker | 60 | Haematological | 0.003 | (0.001, 0.008) |
| Men | Fatigue | Ever smoker | 60 | Other cancers | 0.004 | (0.002, 0.006) |
| Men | Fatigue | Ever smoker | 60 | Death w/o cancer | 0.010 | (0.005, 0.018) |
| Men | Fatigue | Ever smoker | 70 | Lung | 0.012 | (0.006, 0.020) |
| Men | Fatigue | Ever smoker | 70 | Upper GI | 0.005 | (0.002, 0.011) |
| Men | Fatigue | Ever smoker | 70 | Lower GI | 0.007 | (0.003, 0.014) |
| Men | Fatigue | Ever smoker | 70 | Urological | 0.003 | (0.001, 0.009) |
| Men | Fatigue | Ever smoker | 70 | Prostate | 0.013 | (0.007, 0.022) |
| Men | Fatigue | Ever smoker | 70 | Haematological | 0.006 | (0.003, 0.012) |
| Men | Fatigue | Ever smoker | 70 | Other cancers | 0.007 | (0.003, 0.015) |
| Men | Fatigue | Ever smoker | 70 | Death w/o cancer | 0.022 | (0.015, 0.032) |
| Men | Fatigue | Ever smoker | 80 | Lung | 0.015 | (0.007, 0.029) |
| Men | Fatigue | Ever smoker | 80 | Upper GI | 0.007 | (0.004, 0.012) |
| Men | Fatigue | Ever smoker | 80 | Lower GI | 0.010 | (0.004, 0.023) |
| Men | Fatigue | Ever smoker | 80 | Urological | 0.004 | (0.002, 0.010) |
| Men | Fatigue | Ever smoker | 80 | Prostate | 0.016 | (0.009, 0.029) |
| Men | Fatigue | Ever smoker | 80 | Haematological | 0.008 | (0.004, 0.016) |
| Men | Fatigue | Ever smoker | 80 | Other cancers | 0.009 | (0.005, 0.014) |
| Men | Fatigue | Ever smoker | 80 | Death w/o cancer | 0.064 | (0.047, 0.088) |
| Men | Fatigue | Ever smoker | 90 | Lung | 0.016 | (0.008, 0.030) |
| Men | Fatigue | Ever smoker | 90 | Upper GI | 0.007 | (0.003, 0.015) |
| Men | Fatigue | Ever smoker | 90 | Lower GI | 0.009 | (0.005, 0.019) |
| Men | Fatigue | Ever smoker | 90 | Urological | 0.005 | (0.002, 0.013) |
| Men | Fatigue | Ever smoker | 90 | Prostate | 0.017 | (0.010, 0.028) |
| Men | Fatigue | Ever smoker | 90 | Haematological | 0.010 | (0.005, 0.020) |
| Men | Fatigue | Ever smoker | 90 | Other cancers | 0.011 | (0.005, 0.023) |
| Men | Fatigue | Ever smoker | 90 | Death w/o cancer | 0.178 | (0.157, 0.201) |
| Men | Haematuria | Never smoker | 30 | Urological | 0.003 | (<0.001, 0.007) |
| Men | Haematuria | Never smoker | 40 | Urological | 0.009 | (0.005, 0.015) |
| Men | Haematuria | Never smoker | 40 | Death w/o cancer | 0.002 | (<0.001, 0.003) |
| Men | Haematuria | Never smoker | 50 | Urological | 0.022 | (0.014, 0.035) |
| Men | Haematuria | Never smoker | 50 | Prostate | 0.003 | (<0.001, 0.007) |
| Men | Haematuria | Never smoker | 50 | Other cancers | 0.001 | (<0.001, 0.003) |
| Men | Haematuria | Never smoker | 50 | Death w/o cancer | 0.003 | (0.001, 0.007) |
| Men | Haematuria | Never smoker | 60 | Lower GI | 0.001 | (<0.001, 0.003) |
| Men | Haematuria | Never smoker | 60 | Urological | 0.055 | (0.040, 0.076) |
| Men | Haematuria | Never smoker | 60 | Prostate | 0.018 | (0.012, 0.029) |
| Men | Haematuria | Never smoker | 60 | Haematological | 0.002 | (<0.001, 0.003) |
| Men | Haematuria | Never smoker | 60 | Other cancers | 0.002 | (<0.001, 0.005) |
| Men | Haematuria | Never smoker | 60 | Death w/o cancer | 0.006 | (0.003, 0.010) |
| Men | Haematuria | Never smoker | 70 | Lung | 0.001 | (<0.001, 0.003) |
| Men | Haematuria | Never smoker | 70 | Upper GI | 0.002 | (<0.001, 0.003) |
| Men | Haematuria | Never smoker | 70 | Lower GI | 0.003 | (0.001, 0.009) |
| Men | Haematuria | Never smoker | 70 | Urological | 0.097 | (0.078, 0.119) |
| Men | Haematuria | Never smoker | 70 | Prostate | 0.044 | (0.034, 0.057) |
| Men | Haematuria | Never smoker | 70 | Haematological | 0.003 | (0.001, 0.007) |
| Men | Haematuria | Never smoker | 70 | Other cancers | 0.004 | (0.002, 0.007) |
| Men | Haematuria | Never smoker | 70 | Death w/o cancer | 0.016 | (0.010, 0.025) |
| Men | Haematuria | Never smoker | 80 | Lung | 0.001 | (<0.001, 0.003) |
| Men | Haematuria | Never smoker | 80 | Upper GI | 0.002 | (<0.001, 0.003) |
| Men | Haematuria | Never smoker | 80 | Lower GI | 0.004 | (0.002, 0.008) |
| Men | Haematuria | Never smoker | 80 | Urological | 0.117 | (0.099, 0.137) |
| Men | Haematuria | Never smoker | 80 | Prostate | 0.057 | (0.043, 0.075) |
| Men | Haematuria | Never smoker | 80 | Haematological | 0.003 | (0.002, 0.006) |
| Men | Haematuria | Never smoker | 80 | Other cancers | 0.005 | (0.002, 0.009) |
| Men | Haematuria | Never smoker | 80 | Death w/o cancer | 0.043 | (0.032, 0.057) |
| Men | Haematuria | Never smoker | 90 | Lung | 0.002 | (<0.001, 0.004) |
| Men | Haematuria | Never smoker | 90 | Upper GI | 0.002 | (<0.001, 0.004) |
| Men | Haematuria | Never smoker | 90 | Lower GI | 0.005 | (0.002, 0.013) |
| Men | Haematuria | Never smoker | 90 | Urological | 0.127 | (0.102, 0.157) |
| Men | Haematuria | Never smoker | 90 | Prostate | 0.059 | (0.045, 0.078) |
| Men | Haematuria | Never smoker | 90 | Haematological | 0.004 | (0.001, 0.011) |
| Men | Haematuria | Never smoker | 90 | Other cancers | 0.006 | (0.003, 0.013) |
| Men | Haematuria | Never smoker | 90 | Death w/o cancer | 0.121 | (0.101, 0.144) |
| Men | Haematuria | Ever smoker | 30 | Urological | 0.004 | (0.002, 0.008) |
| Men | Haematuria | Ever smoker | 40 | Urological | 0.012 | (0.007, 0.021) |
| Men | Haematuria | Ever smoker | 40 | Death w/o cancer | 0.002 | (<0.001, 0.004) |
| Men | Haematuria | Ever smoker | 50 | Urological | 0.029 | (0.021, 0.039) |
| Men | Haematuria | Ever smoker | 50 | Prostate | 0.002 | (0.001, 0.004) |
| Men | Haematuria | Ever smoker | 50 | Other cancers | 0.001 | (<0.001, 0.003) |
| Men | Haematuria | Ever smoker | 50 | Death w/o cancer | 0.003 | (0.002, 0.007) |
| Men | Haematuria | Ever smoker | 60 | Lung | 0.002 | (<0.001, 0.006) |
| Men | Haematuria | Ever smoker | 60 | Upper GI | 0.001 | (<0.001, 0.002) |
| Men | Haematuria | Ever smoker | 60 | Lower GI | 0.002 | (<0.001, 0.003) |
| Men | Haematuria | Ever smoker | 60 | Urological | 0.076 | (0.059, 0.097) |
| Men | Haematuria | Ever smoker | 60 | Prostate | 0.017 | (0.010, 0.027) |
| Men | Haematuria | Ever smoker | 60 | Haematological | 0.001 | (<0.001, 0.003) |
| Men | Haematuria | Ever smoker | 60 | Other cancers | 0.003 | (0.001, 0.006) |
| Men | Haematuria | Ever smoker | 60 | Death w/o cancer | 0.007 | (0.004, 0.012) |
| Men | Haematuria | Ever smoker | 70 | Lung | 0.004 | (0.002, 0.011) |
| Men | Haematuria | Ever smoker | 70 | Upper GI | 0.002 | (<0.001, 0.005) |
| Men | Haematuria | Ever smoker | 70 | Lower GI | 0.003 | (0.001, 0.008) |
| Men | Haematuria | Ever smoker | 70 | Urological | 0.129 | (0.108, 0.154) |
| Men | Haematuria | Ever smoker | 70 | Prostate | 0.039 | (0.026, 0.057) |
| Men | Haematuria | Ever smoker | 70 | Haematological | 0.002 | (<0.001, 0.006) |
| Men | Haematuria | Ever smoker | 70 | Other cancers | 0.004 | (0.002, 0.009) |
| Men | Haematuria | Ever smoker | 70 | Death w/o cancer | 0.016 | (0.010, 0.025) |
| Men | Haematuria | Ever smoker | 80 | Lung | 0.006 | (0.003, 0.012) |
| Men | Haematuria | Ever smoker | 80 | Upper GI | 0.003 | (<0.001, 0.007) |
| Men | Haematuria | Ever smoker | 80 | Lower GI | 0.004 | (0.001, 0.011) |
| Men | Haematuria | Ever smoker | 80 | Urological | 0.153 | (0.131, 0.178) |
| Men | Haematuria | Ever smoker | 80 | Prostate | 0.048 | (0.037, 0.063) |
| Men | Haematuria | Ever smoker | 80 | Haematological | 0.003 | (0.001, 0.006) |
| Men | Haematuria | Ever smoker | 80 | Other cancers | 0.005 | (0.002, 0.017) |
| Men | Haematuria | Ever smoker | 80 | Death w/o cancer | 0.046 | (0.032, 0.065) |
| Men | Haematuria | Ever smoker | 90 | Lung | 0.006 | (0.003, 0.012) |
| Men | Haematuria | Ever smoker | 90 | Upper GI | 0.003 | (0.001, 0.005) |
| Men | Haematuria | Ever smoker | 90 | Lower GI | 0.005 | (0.002, 0.013) |
| Men | Haematuria | Ever smoker | 90 | Urological | 0.164 | (0.133, 0.201) |
| Men | Haematuria | Ever smoker | 90 | Prostate | 0.052 | (0.037, 0.073) |
| Men | Haematuria | Ever smoker | 90 | Haematological | 0.004 | (0.002, 0.010) |
| Men | Haematuria | Ever smoker | 90 | Other cancers | 0.007 | (0.002, 0.021) |
| Men | Haematuria | Ever smoker | 90 | Death w/o cancer | 0.126 | (0.108, 0.147) |
| Men | Haemoptysis | Never smoker | 30 | Other cancers | 0.001 | (<0.001, 0.002) |
| Men | Haemoptysis | Never smoker | 30 | Death w/o cancer | 0.001 | (<0.001, 0.003) |
| Men | Haemoptysis | Never smoker | 40 | Other cancers | 0.001 | (<0.001, 0.002) |
| Men | Haemoptysis | Never smoker | 40 | Death w/o cancer | 0.003 | (0.001, 0.007) |
| Men | Haemoptysis | Never smoker | 50 | Lung | 0.005 | (0.002, 0.010) |
| Men | Haemoptysis | Never smoker | 50 | Haematological | 0.001 | (<0.001, 0.003) |
| Men | Haemoptysis | Never smoker | 50 | Other cancers | 0.002 | (0.001, 0.006) |
| Men | Haemoptysis | Never smoker | 50 | Death w/o cancer | 0.005 | (0.003, 0.010) |
| Men | Haemoptysis | Never smoker | 60 | Lung | 0.015 | (0.009, 0.024) |
| Men | Haemoptysis | Never smoker | 60 | Upper GI | 0.002 | (0.001, 0.005) |
| Men | Haemoptysis | Never smoker | 60 | Lower GI | 0.002 | (<0.001, 0.004) |
| Men | Haemoptysis | Never smoker | 60 | Prostate | 0.003 | (0.001, 0.007) |
| Men | Haemoptysis | Never smoker | 60 | Haematological | 0.003 | (0.002, 0.006) |
| Men | Haemoptysis | Never smoker | 60 | Other cancers | 0.004 | (0.002, 0.010) |
| Men | Haemoptysis | Never smoker | 60 | Death w/o cancer | 0.011 | (0.007, 0.019) |
| Men | Haemoptysis | Never smoker | 70 | Lung | 0.034 | (0.025, 0.047) |
| Men | Haemoptysis | Never smoker | 70 | Upper GI | 0.005 | (0.002, 0.013) |
| Men | Haemoptysis | Never smoker | 70 | Lower GI | 0.004 | (0.001, 0.009) |
| Men | Haemoptysis | Never smoker | 70 | Urological | 0.002 | (<0.001, 0.004) |
| Men | Haemoptysis | Never smoker | 70 | Prostate | 0.008 | (0.004, 0.016) |
| Men | Haemoptysis | Never smoker | 70 | Haematological | 0.007 | (0.003, 0.014) |
| Men | Haemoptysis | Never smoker | 70 | Other cancers | 0.007 | (0.003, 0.019) |
| Men | Haemoptysis | Never smoker | 70 | Death w/o cancer | 0.028 | (0.019, 0.042) |
| Men | Haemoptysis | Never smoker | 80 | Lung | 0.044 | (0.030, 0.065) |
| Men | Haemoptysis | Never smoker | 80 | Upper GI | 0.005 | (0.002, 0.010) |
| Men | Haemoptysis | Never smoker | 80 | Lower GI | 0.004 | (0.001, 0.014) |
| Men | Haemoptysis | Never smoker | 80 | Urological | 0.002 | (<0.001, 0.004) |
| Men | Haemoptysis | Never smoker | 80 | Prostate | 0.009 | (0.004, 0.020) |
| Men | Haemoptysis | Never smoker | 80 | Haematological | 0.010 | (0.005, 0.019) |
| Men | Haemoptysis | Never smoker | 80 | Other cancers | 0.009 | (0.004, 0.020) |
| Men | Haemoptysis | Never smoker | 80 | Death w/o cancer | 0.078 | (0.060, 0.100) |
| Men | Haemoptysis | Never smoker | 90 | Lung | 0.048 | (0.034, 0.067) |
| Men | Haemoptysis | Never smoker | 90 | Upper GI | 0.005 | (0.002, 0.011) |
| Men | Haemoptysis | Never smoker | 90 | Lower GI | 0.004 | (0.002, 0.013) |
| Men | Haemoptysis | Never smoker | 90 | Urological | 0.002 | (<0.001, 0.004) |
| Men | Haemoptysis | Never smoker | 90 | Prostate | 0.009 | (0.005, 0.015) |
| Men | Haemoptysis | Never smoker | 90 | Haematological | 0.011 | (0.005, 0.026) |
| Men | Haemoptysis | Never smoker | 90 | Other cancers | 0.012 | (0.005, 0.028) |
| Men | Haemoptysis | Never smoker | 90 | Death w/o cancer | 0.217 | (0.189, 0.248) |
| Men | Haemoptysis | Ever smoker | 30 | Other cancers | 0.001 | (<0.001, 0.002) |
| Men | Haemoptysis | Ever smoker | 30 | Death w/o cancer | 0.001 | (<0.001, 0.003) |
| Men | Haemoptysis | Ever smoker | 40 | Lung | 0.002 | (<0.001, 0.004) |
| Men | Haemoptysis | Ever smoker | 40 | Other cancers | 0.002 | (<0.001, 0.004) |
| Men | Haemoptysis | Ever smoker | 40 | Death w/o cancer | 0.003 | (0.002, 0.007) |
| Men | Haemoptysis | Ever smoker | 50 | Lung | 0.017 | (0.011, 0.026) |
| Men | Haemoptysis | Ever smoker | 50 | Upper GI | 0.001 | (<0.001, 0.002) |
| Men | Haemoptysis | Ever smoker | 50 | Haematological | 0.002 | (<0.001, 0.004) |
| Men | Haemoptysis | Ever smoker | 50 | Other cancers | 0.003 | (0.001, 0.006) |
| Men | Haemoptysis | Ever smoker | 50 | Death w/o cancer | 0.006 | (0.003, 0.012) |
| Men | Haemoptysis | Ever smoker | 60 | Lung | 0.059 | (0.043, 0.079) |
| Men | Haemoptysis | Ever smoker | 60 | Upper GI | 0.003 | (0.001, 0.007) |
| Men | Haemoptysis | Ever smoker | 60 | Lower GI | 0.002 | (<0.001, 0.003) |
| Men | Haemoptysis | Ever smoker | 60 | Urological | 0.001 | (<0.001, 0.003) |
| Men | Haemoptysis | Ever smoker | 60 | Prostate | 0.003 | (0.001, 0.007) |
| Men | Haemoptysis | Ever smoker | 60 | Haematological | 0.003 | (0.001, 0.010) |
| Men | Haemoptysis | Ever smoker | 60 | Other cancers | 0.005 | (0.003, 0.010) |
| Men | Haemoptysis | Ever smoker | 60 | Death w/o cancer | 0.012 | (0.008, 0.019) |
| Men | Haemoptysis | Ever smoker | 70 | Lung | 0.129 | (0.105, 0.157) |
| Men | Haemoptysis | Ever smoker | 70 | Upper GI | 0.005 | (0.002, 0.010) |
| Men | Haemoptysis | Ever smoker | 70 | Lower GI | 0.003 | (0.001, 0.009) |
| Men | Haemoptysis | Ever smoker | 70 | Urological | 0.002 | (<0.001, 0.004) |
| Men | Haemoptysis | Ever smoker | 70 | Prostate | 0.006 | (0.002, 0.015) |
| Men | Haemoptysis | Ever smoker | 70 | Haematological | 0.006 | (0.003, 0.013) |
| Men | Haemoptysis | Ever smoker | 70 | Other cancers | 0.008 | (0.004, 0.016) |
| Men | Haemoptysis | Ever smoker | 70 | Death w/o cancer | 0.028 | (0.019, 0.040) |
| Men | Haemoptysis | Ever smoker | 80 | Lung | 0.164 | (0.126, 0.210) |
| Men | Haemoptysis | Ever smoker | 80 | Upper GI | 0.006 | (0.002, 0.015) |
| Men | Haemoptysis | Ever smoker | 80 | Lower GI | 0.004 | (0.002, 0.013) |
| Men | Haemoptysis | Ever smoker | 80 | Urological | 0.002 | (<0.001, 0.004) |
| Men | Haemoptysis | Ever smoker | 80 | Prostate | 0.007 | (0.003, 0.016) |
| Men | Haemoptysis | Ever smoker | 80 | Haematological | 0.008 | (0.003, 0.022) |
| Men | Haemoptysis | Ever smoker | 80 | Other cancers | 0.011 | (0.006, 0.019) |
| Men | Haemoptysis | Ever smoker | 80 | Death w/o cancer | 0.076 | (0.058, 0.099) |
| Men | Haemoptysis | Ever smoker | 90 | Lung | 0.176 | (0.147, 0.209) |
| Men | Haemoptysis | Ever smoker | 90 | Upper GI | 0.006 | (0.002, 0.021) |
| Men | Haemoptysis | Ever smoker | 90 | Lower GI | 0.004 | (0.001, 0.014) |
| Men | Haemoptysis | Ever smoker | 90 | Urological | 0.002 | (<0.001, 0.004) |
| Men | Haemoptysis | Ever smoker | 90 | Prostate | 0.007 | (0.004, 0.013) |
| Men | Haemoptysis | Ever smoker | 90 | Haematological | 0.009 | (0.004, 0.022) |
| Men | Haemoptysis | Ever smoker | 90 | Other cancers | 0.013 | (0.006, 0.026) |
| Men | Haemoptysis | Ever smoker | 90 | Death w/o cancer | 0.208 | (0.166, 0.256) |
| Men | Jaundice | Never smoker | 30 | Upper GI | 0.002 | (<0.001, 0.005) |
| Men | Jaundice | Never smoker | 30 | Other cancers | 0.009 | (0.002, 0.034) |
| Men | Jaundice | Never smoker | 30 | Death w/o cancer | 0.005 | (0.003, 0.010) |
| Men | Jaundice | Never smoker | 40 | Upper GI | 0.010 | (0.006, 0.019) |
| Men | Jaundice | Never smoker | 40 | Haematological | 0.002 | (<0.001, 0.004) |
| Men | Jaundice | Never smoker | 40 | Other cancers | 0.012 | (0.006, 0.022) |
| Men | Jaundice | Never smoker | 40 | Death w/o cancer | 0.011 | (0.007, 0.019) |
| Men | Jaundice | Never smoker | 50 | Upper GI | 0.043 | (0.030, 0.061) |
| Men | Jaundice | Never smoker | 50 | Lower GI | 0.002 | (<0.001, 0.004) |
| Men | Jaundice | Never smoker | 50 | Urological | 0.002 | (<0.001, 0.003) |
| Men | Jaundice | Never smoker | 50 | Haematological | 0.003 | (0.002, 0.008) |
| Men | Jaundice | Never smoker | 50 | Other cancers | 0.025 | (0.015, 0.039) |
| Men | Jaundice | Never smoker | 50 | Death w/o cancer | 0.019 | (0.012, 0.032) |
| Men | Jaundice | Never smoker | 60 | Lung | 0.002 | (<0.001, 0.005) |
| Men | Jaundice | Never smoker | 60 | Upper GI | 0.115 | (0.091, 0.144) |
| Men | Jaundice | Never smoker | 60 | Lower GI | 0.004 | (0.002, 0.007) |
| Men | Jaundice | Never smoker | 60 | Urological | 0.004 | (0.001, 0.014) |
| Men | Jaundice | Never smoker | 60 | Prostate | 0.004 | (0.001, 0.014) |
| Men | Jaundice | Never smoker | 60 | Haematological | 0.007 | (0.004, 0.014) |
| Men | Jaundice | Never smoker | 60 | Other cancers | 0.041 | (0.028, 0.060) |
| Men | Jaundice | Never smoker | 60 | Death w/o cancer | 0.038 | (0.028, 0.051) |
| Men | Jaundice | Never smoker | 70 | Lung | 0.003 | (0.001, 0.006) |
| Men | Jaundice | Never smoker | 70 | Upper GI | 0.204 | (0.158, 0.259) |
| Men | Jaundice | Never smoker | 70 | Lower GI | 0.008 | (0.003, 0.021) |
| Men | Jaundice | Never smoker | 70 | Urological | 0.006 | (0.002, 0.022) |
| Men | Jaundice | Never smoker | 70 | Prostate | 0.008 | (0.003, 0.028) |
| Men | Jaundice | Never smoker | 70 | Haematological | 0.012 | (0.006, 0.022) |
| Men | Jaundice | Never smoker | 70 | Other cancers | 0.061 | (0.039, 0.092) |
| Men | Jaundice | Never smoker | 70 | Death w/o cancer | 0.085 | (0.065, 0.111) |
| Men | Jaundice | Never smoker | 80 | Lung | 0.003 | (<0.001, 0.010) |
| Men | Jaundice | Never smoker | 80 | Upper GI | 0.226 | (0.187, 0.271) |
| Men | Jaundice | Never smoker | 80 | Lower GI | 0.009 | (0.004, 0.018) |
| Men | Jaundice | Never smoker | 80 | Urological | 0.006 | (0.002, 0.017) |
| Men | Jaundice | Never smoker | 80 | Prostate | 0.009 | (0.004, 0.020) |
| Men | Jaundice | Never smoker | 80 | Haematological | 0.014 | (0.007, 0.028) |
| Men | Jaundice | Never smoker | 80 | Other cancers | 0.079 | (0.055, 0.111) |
| Men | Jaundice | Never smoker | 80 | Death w/o cancer | 0.210 | (0.182, 0.242) |
| Men | Jaundice | Never smoker | 90 | Lung | 0.003 | (<0.001, 0.010) |
| Men | Jaundice | Never smoker | 90 | Upper GI | 0.199 | (0.161, 0.242) |
| Men | Jaundice | Never smoker | 90 | Lower GI | 0.008 | (0.003, 0.024) |
| Men | Jaundice | Never smoker | 90 | Urological | 0.004 | (0.002, 0.010) |
| Men | Jaundice | Never smoker | 90 | Prostate | 0.007 | (0.003, 0.017) |
| Men | Jaundice | Never smoker | 90 | Haematological | 0.014 | (0.007, 0.026) |
| Men | Jaundice | Never smoker | 90 | Other cancers | 0.086 | (0.062, 0.118) |
| Men | Jaundice | Never smoker | 90 | Death w/o cancer | 0.472 | (0.407, 0.537) |
| Men | Jaundice | Ever smoker | 30 | Upper GI | 0.002 | (<0.001, 0.006) |
| Men | Jaundice | Ever smoker | 30 | Other cancers | 0.011 | (0.006, 0.023) |
| Men | Jaundice | Ever smoker | 30 | Death w/o cancer | 0.006 | (0.003, 0.010) |
| Men | Jaundice | Ever smoker | 40 | Upper GI | 0.012 | (0.007, 0.021) |
| Men | Jaundice | Ever smoker | 40 | Haematological | 0.002 | (<0.001, 0.005) |
| Men | Jaundice | Ever smoker | 40 | Other cancers | 0.014 | (0.009, 0.022) |
| Men | Jaundice | Ever smoker | 40 | Death w/o cancer | 0.013 | (0.008, 0.022) |
| Men | Jaundice | Ever smoker | 50 | Lung | 0.002 | (<0.001, 0.005) |
| Men | Jaundice | Ever smoker | 50 | Upper GI | 0.057 | (0.042, 0.077) |
| Men | Jaundice | Ever smoker | 50 | Lower GI | 0.002 | (<0.001, 0.003) |
| Men | Jaundice | Ever smoker | 50 | Urological | 0.002 | (0.001, 0.005) |
| Men | Jaundice | Ever smoker | 50 | Haematological | 0.004 | (0.001, 0.009) |
| Men | Jaundice | Ever smoker | 50 | Other cancers | 0.028 | (0.020, 0.041) |
| Men | Jaundice | Ever smoker | 50 | Death w/o cancer | 0.021 | (0.014, 0.033) |
| Men | Jaundice | Ever smoker | 60 | Lung | 0.005 | (0.002, 0.012) |
| Men | Jaundice | Ever smoker | 60 | Upper GI | 0.148 | (0.130, 0.169) |
| Men | Jaundice | Ever smoker | 60 | Lower GI | 0.004 | (0.001, 0.011) |
| Men | Jaundice | Ever smoker | 60 | Urological | 0.005 | (0.002, 0.016) |
| Men | Jaundice | Ever smoker | 60 | Prostate | 0.003 | (0.001, 0.008) |
| Men | Jaundice | Ever smoker | 60 | Haematological | 0.007 | (0.002, 0.022) |
| Men | Jaundice | Ever smoker | 60 | Other cancers | 0.049 | (0.031, 0.076) |
| Men | Jaundice | Ever smoker | 60 | Death w/o cancer | 0.041 | (0.028, 0.059) |
| Men | Jaundice | Ever smoker | 70 | Lung | 0.010 | (0.006, 0.018) |
| Men | Jaundice | Ever smoker | 70 | Upper GI | 0.264 | (0.215, 0.319) |
| Men | Jaundice | Ever smoker | 70 | Lower GI | 0.007 | (0.002, 0.023) |
| Men | Jaundice | Ever smoker | 70 | Urological | 0.008 | (0.002, 0.026) |
| Men | Jaundice | Ever smoker | 70 | Prostate | 0.007 | (0.003, 0.017) |
| Men | Jaundice | Ever smoker | 70 | Haematological | 0.011 | (0.005, 0.028) |
| Men | Jaundice | Ever smoker | 70 | Other cancers | 0.071 | (0.047, 0.105) |
| Men | Jaundice | Ever smoker | 70 | Death w/o cancer | 0.084 | (0.068, 0.105) |
| Men | Jaundice | Ever smoker | 80 | Lung | 0.012 | (0.006, 0.022) |
| Men | Jaundice | Ever smoker | 80 | Upper GI | 0.285 | (0.242, 0.331) |
| Men | Jaundice | Ever smoker | 80 | Lower GI | 0.009 | (0.003, 0.026) |
| Men | Jaundice | Ever smoker | 80 | Urological | 0.007 | (0.004, 0.015) |
| Men | Jaundice | Ever smoker | 80 | Prostate | 0.008 | (0.003, 0.019) |
| Men | Jaundice | Ever smoker | 80 | Haematological | 0.013 | (0.008, 0.023) |
| Men | Jaundice | Ever smoker | 80 | Other cancers | 0.087 | (0.067, 0.114) |
| Men | Jaundice | Ever smoker | 80 | Death w/o cancer | 0.207 | (0.175, 0.242) |
| Men | Jaundice | Ever smoker | 90 | Lung | 0.010 | (0.005, 0.018) |
| Men | Jaundice | Ever smoker | 90 | Upper GI | 0.246 | (0.202, 0.296) |
| Men | Jaundice | Ever smoker | 90 | Lower GI | 0.008 | (0.002, 0.027) |
| Men | Jaundice | Ever smoker | 90 | Urological | 0.006 | (0.002, 0.014) |
| Men | Jaundice | Ever smoker | 90 | Prostate | 0.006 | (0.001, 0.021) |
| Men | Jaundice | Ever smoker | 90 | Haematological | 0.012 | (0.007, 0.023) |
| Men | Jaundice | Ever smoker | 90 | Other cancers | 0.095 | (0.071, 0.125) |
| Men | Jaundice | Ever smoker | 90 | Death w/o cancer | 0.456 | (0.386, 0.528) |
| Men | Night sweats | Never smoker | 40 | Haematological | 0.002 | (<0.001, 0.003) |
| Men | Night sweats | Never smoker | 50 | Lower GI | 0.001 | (<0.001, 0.002) |
| Men | Night sweats | Never smoker | 50 | Urological | 0.001 | (<0.001, 0.002) |
| Men | Night sweats | Never smoker | 50 | Haematological | 0.003 | (0.001, 0.005) |
| Men | Night sweats | Never smoker | 50 | Other cancers | 0.002 | (<0.001, 0.004) |
| Men | Night sweats | Never smoker | 50 | Death w/o cancer | 0.002 | (<0.001, 0.004) |
| Men | Night sweats | Never smoker | 60 | Lung | 0.002 | (<0.001, 0.006) |
| Men | Night sweats | Never smoker | 60 | Lower GI | 0.003 | (0.001, 0.007) |
| Men | Night sweats | Never smoker | 60 | Urological | 0.003 | (0.001, 0.009) |
| Men | Night sweats | Never smoker | 60 | Prostate | 0.007 | (0.003, 0.015) |
| Men | Night sweats | Never smoker | 60 | Haematological | 0.006 | (0.002, 0.019) |
| Men | Night sweats | Never smoker | 60 | Other cancers | 0.003 | (0.001, 0.007) |
| Men | Night sweats | Never smoker | 60 | Death w/o cancer | 0.004 | (0.001, 0.011) |
| Men | Night sweats | Never smoker | 70 | Lung | 0.005 | (0.002, 0.013) |
| Men | Night sweats | Never smoker | 70 | Upper GI | 0.001 | (<0.001, 0.004) |
| Men | Night sweats | Never smoker | 70 | Lower GI | 0.007 | (0.002, 0.021) |
| Men | Night sweats | Never smoker | 70 | Urological | 0.006 | (0.003, 0.014) |
| Men | Night sweats | Never smoker | 70 | Prostate | 0.018 | (0.011, 0.029) |
| Men | Night sweats | Never smoker | 70 | Haematological | 0.013 | (0.006, 0.027) |
| Men | Night sweats | Never smoker | 70 | Other cancers | 0.006 | (0.002, 0.016) |
| Men | Night sweats | Never smoker | 70 | Death w/o cancer | 0.009 | (0.005, 0.016) |
| Men | Night sweats | Never smoker | 80 | Lung | 0.007 | (0.002, 0.020) |
| Men | Night sweats | Never smoker | 80 | Upper GI | 0.001 | (<0.001, 0.005) |
| Men | Night sweats | Never smoker | 80 | Lower GI | 0.009 | (0.004, 0.022) |
| Men | Night sweats | Never smoker | 80 | Urological | 0.007 | (0.002, 0.029) |
| Men | Night sweats | Never smoker | 80 | Prostate | 0.022 | (0.014, 0.036) |
| Men | Night sweats | Never smoker | 80 | Haematological | 0.016 | (0.008, 0.034) |
| Men | Night sweats | Never smoker | 80 | Other cancers | 0.007 | (0.003, 0.015) |
| Men | Night sweats | Never smoker | 80 | Death w/o cancer | 0.025 | (0.015, 0.040) |
| Men | Night sweats | Never smoker | 90 | Lung | 0.007 | (0.003, 0.017) |
| Men | Night sweats | Never smoker | 90 | Upper GI | 0.002 | (<0.001, 0.005) |
| Men | Night sweats | Never smoker | 90 | Lower GI | 0.011 | (0.004, 0.026) |
| Men | Night sweats | Never smoker | 90 | Urological | 0.008 | (0.003, 0.018) |
| Men | Night sweats | Never smoker | 90 | Prostate | 0.024 | (0.015, 0.039) |
| Men | Night sweats | Never smoker | 90 | Haematological | 0.021 | (0.010, 0.044) |
| Men | Night sweats | Never smoker | 90 | Other cancers | 0.010 | (0.005, 0.021) |
| Men | Night sweats | Never smoker | 90 | Death w/o cancer | 0.072 | (0.045, 0.114) |
| Men | Night sweats | Ever smoker | 40 | Haematological | 0.001 | (<0.001, 0.003) |
| Men | Night sweats | Ever smoker | 40 | Other cancers | 0.001 | (<0.001, 0.002) |
| Men | Night sweats | Ever smoker | 40 | Death w/o cancer | 0.001 | (<0.001, 0.002) |
| Men | Night sweats | Ever smoker | 50 | Lung | 0.002 | (0.001, 0.004) |
| Men | Night sweats | Ever smoker | 50 | Lower GI | 0.001 | (<0.001, 0.003) |
| Men | Night sweats | Ever smoker | 50 | Urological | 0.002 | (<0.001, 0.005) |
| Men | Night sweats | Ever smoker | 50 | Haematological | 0.002 | (0.001, 0.006) |
| Men | Night sweats | Ever smoker | 50 | Other cancers | 0.002 | (<0.001, 0.006) |
| Men | Night sweats | Ever smoker | 50 | Death w/o cancer | 0.002 | (<0.001, 0.004) |
| Men | Night sweats | Ever smoker | 60 | Lung | 0.009 | (0.004, 0.023) |
| Men | Night sweats | Ever smoker | 60 | Lower GI | 0.003 | (0.001, 0.008) |
| Men | Night sweats | Ever smoker | 60 | Urological | 0.005 | (0.002, 0.016) |
| Men | Night sweats | Ever smoker | 60 | Prostate | 0.006 | (0.003, 0.013) |
| Men | Night sweats | Ever smoker | 60 | Haematological | 0.007 | (0.002, 0.018) |
| Men | Night sweats | Ever smoker | 60 | Other cancers | 0.004 | (0.002, 0.011) |
| Men | Night sweats | Ever smoker | 60 | Death w/o cancer | 0.004 | (0.002, 0.008) |
| Men | Night sweats | Ever smoker | 70 | Lung | 0.020 | (0.009, 0.042) |
| Men | Night sweats | Ever smoker | 70 | Upper GI | 0.002 | (<0.001, 0.004) |
| Men | Night sweats | Ever smoker | 70 | Lower GI | 0.007 | (0.002, 0.021) |
| Men | Night sweats | Ever smoker | 70 | Urological | 0.008 | (0.004, 0.018) |
| Men | Night sweats | Ever smoker | 70 | Prostate | 0.015 | (0.007, 0.031) |
| Men | Night sweats | Ever smoker | 70 | Haematological | 0.012 | (0.005, 0.025) |
| Men | Night sweats | Ever smoker | 70 | Other cancers | 0.006 | (0.002, 0.020) |
| Men | Night sweats | Ever smoker | 70 | Death w/o cancer | 0.009 | (0.005, 0.016) |
| Men | Night sweats | Ever smoker | 80 | Lung | 0.026 | (0.016, 0.041) |
| Men | Night sweats | Ever smoker | 80 | Upper GI | 0.002 | (<0.001, 0.006) |
| Men | Night sweats | Ever smoker | 80 | Lower GI | 0.010 | (0.004, 0.023) |
| Men | Night sweats | Ever smoker | 80 | Urological | 0.010 | (0.003, 0.031) |
| Men | Night sweats | Ever smoker | 80 | Prostate | 0.019 | (0.011, 0.032) |
| Men | Night sweats | Ever smoker | 80 | Haematological | 0.017 | (0.007, 0.037) |
| Men | Night sweats | Ever smoker | 80 | Other cancers | 0.009 | (0.005, 0.019) |
| Men | Night sweats | Ever smoker | 80 | Death w/o cancer | 0.026 | (0.016, 0.042) |
| Men | Night sweats | Ever smoker | 90 | Lung | 0.028 | (0.017, 0.046) |
| Men | Night sweats | Ever smoker | 90 | Upper GI | 0.002 | (<0.001, 0.005) |
| Men | Night sweats | Ever smoker | 90 | Lower GI | 0.012 | (0.005, 0.028) |
| Men | Night sweats | Ever smoker | 90 | Urological | 0.011 | (0.005, 0.026) |
| Men | Night sweats | Ever smoker | 90 | Prostate | 0.021 | (0.013, 0.032) |
| Men | Night sweats | Ever smoker | 90 | Haematological | 0.021 | (0.009, 0.044) |
| Men | Night sweats | Ever smoker | 90 | Other cancers | 0.012 | (0.006, 0.023) |
| Men | Night sweats | Ever smoker | 90 | Death w/o cancer | 0.079 | (0.048, 0.128) |
| Men | Rectal bleeding | Never smoker | 30 | Lower GI | 0.002 | (0.001, 0.003) |
| Men | Rectal bleeding | Never smoker | 40 | Lower GI | 0.003 | (0.001, 0.006) |
| Men | Rectal bleeding | Never smoker | 40 | Death w/o cancer | 0.001 | (<0.001, 0.003) |
| Men | Rectal bleeding | Never smoker | 50 | Lower GI | 0.012 | (0.007, 0.020) |
| Men | Rectal bleeding | Never smoker | 50 | Other cancers | 0.002 | (<0.001, 0.003) |
| Men | Rectal bleeding | Never smoker | 50 | Death w/o cancer | 0.003 | (0.001, 0.007) |
| Men | Rectal bleeding | Never smoker | 60 | Upper GI | 0.002 | (<0.001, 0.005) |
| Men | Rectal bleeding | Never smoker | 60 | Lower GI | 0.030 | (0.020, 0.044) |
| Men | Rectal bleeding | Never smoker | 60 | Urological | 0.001 | (<0.001, 0.003) |
| Men | Rectal bleeding | Never smoker | 60 | Prostate | 0.006 | (0.003, 0.015) |
| Men | Rectal bleeding | Never smoker | 60 | Haematological | 0.002 | (<0.001, 0.003) |
| Men | Rectal bleeding | Never smoker | 60 | Other cancers | 0.003 | (0.001, 0.006) |
| Men | Rectal bleeding | Never smoker | 60 | Death w/o cancer | 0.007 | (0.003, 0.015) |
| Men | Rectal bleeding | Never smoker | 70 | Lung | 0.001 | (<0.001, 0.002) |
| Men | Rectal bleeding | Never smoker | 70 | Upper GI | 0.004 | (0.001, 0.009) |
| Men | Rectal bleeding | Never smoker | 70 | Lower GI | 0.065 | (0.041, 0.103) |
| Men | Rectal bleeding | Never smoker | 70 | Urological | 0.003 | (0.001, 0.006) |
| Men | Rectal bleeding | Never smoker | 70 | Prostate | 0.014 | (0.008, 0.025) |
| Men | Rectal bleeding | Never smoker | 70 | Haematological | 0.003 | (0.001, 0.007) |
| Men | Rectal bleeding | Never smoker | 70 | Other cancers | 0.005 | (0.002, 0.012) |
| Men | Rectal bleeding | Never smoker | 70 | Death w/o cancer | 0.016 | (0.010, 0.025) |
| Men | Rectal bleeding | Never smoker | 80 | Lung | 0.001 | (<0.001, 0.003) |
| Men | Rectal bleeding | Never smoker | 80 | Upper GI | 0.005 | (0.002, 0.009) |
| Men | Rectal bleeding | Never smoker | 80 | Lower GI | 0.085 | (0.070, 0.104) |
| Men | Rectal bleeding | Never smoker | 80 | Urological | 0.003 | (<0.001, 0.007) |
| Men | Rectal bleeding | Never smoker | 80 | Prostate | 0.019 | (0.013, 0.028) |
| Men | Rectal bleeding | Never smoker | 80 | Haematological | 0.004 | (0.002, 0.010) |
| Men | Rectal bleeding | Never smoker | 80 | Other cancers | 0.006 | (0.003, 0.015) |
| Men | Rectal bleeding | Never smoker | 80 | Death w/o cancer | 0.046 | (0.033, 0.065) |
| Men | Rectal bleeding | Never smoker | 90 | Lung | 0.001 | (<0.001, 0.003) |
| Men | Rectal bleeding | Never smoker | 90 | Upper GI | 0.004 | (0.002, 0.010) |
| Men | Rectal bleeding | Never smoker | 90 | Lower GI | 0.096 | (0.077, 0.119) |
| Men | Rectal bleeding | Never smoker | 90 | Urological | 0.003 | (0.001, 0.007) |
| Men | Rectal bleeding | Never smoker | 90 | Prostate | 0.019 | (0.012, 0.033) |
| Men | Rectal bleeding | Never smoker | 90 | Haematological | 0.004 | (0.002, 0.010) |
| Men | Rectal bleeding | Never smoker | 90 | Other cancers | 0.008 | (0.004, 0.016) |
| Men | Rectal bleeding | Never smoker | 90 | Death w/o cancer | 0.129 | (0.109, 0.152) |
| Men | Rectal bleeding | Ever smoker | 30 | Lower GI | 0.002 | (0.001, 0.003) |
| Men | Rectal bleeding | Ever smoker | 30 | Death w/o cancer | 0.001 | (<0.001, 0.002) |
| Men | Rectal bleeding | Ever smoker | 40 | Lower GI | 0.003 | (0.001, 0.008) |
| Men | Rectal bleeding | Ever smoker | 40 | Death w/o cancer | 0.002 | (<0.001, 0.004) |
| Men | Rectal bleeding | Ever smoker | 50 | Lower GI | 0.012 | (0.007, 0.020) |
| Men | Rectal bleeding | Ever smoker | 50 | Other cancers | 0.002 | (<0.001, 0.004) |
| Men | Rectal bleeding | Ever smoker | 50 | Death w/o cancer | 0.003 | (0.001, 0.010) |
| Men | Rectal bleeding | Ever smoker | 60 | Lung | 0.002 | (<0.001, 0.004) |
| Men | Rectal bleeding | Ever smoker | 60 | Upper GI | 0.003 | (0.001, 0.005) |
| Men | Rectal bleeding | Ever smoker | 60 | Lower GI | 0.031 | (0.021, 0.046) |
| Men | Rectal bleeding | Ever smoker | 60 | Urological | 0.002 | (0.001, 0.003) |
| Men | Rectal bleeding | Ever smoker | 60 | Prostate | 0.006 | (0.003, 0.011) |
| Men | Rectal bleeding | Ever smoker | 60 | Haematological | 0.002 | (<0.001, 0.004) |
| Men | Rectal bleeding | Ever smoker | 60 | Other cancers | 0.003 | (0.001, 0.009) |
| Men | Rectal bleeding | Ever smoker | 60 | Death w/o cancer | 0.007 | (0.003, 0.016) |
| Men | Rectal bleeding | Ever smoker | 70 | Lung | 0.004 | (0.001, 0.012) |
| Men | Rectal bleeding | Ever smoker | 70 | Upper GI | 0.005 | (0.002, 0.015) |
| Men | Rectal bleeding | Ever smoker | 70 | Lower GI | 0.068 | (0.050, 0.092) |
| Men | Rectal bleeding | Ever smoker | 70 | Urological | 0.003 | (0.001, 0.008) |
| Men | Rectal bleeding | Ever smoker | 70 | Prostate | 0.013 | (0.007, 0.023) |
| Men | Rectal bleeding | Ever smoker | 70 | Haematological | 0.003 | (0.001, 0.007) |
| Men | Rectal bleeding | Ever smoker | 70 | Other cancers | 0.006 | (0.002, 0.016) |
| Men | Rectal bleeding | Ever smoker | 70 | Death w/o cancer | 0.018 | (0.010, 0.032) |
| Men | Rectal bleeding | Ever smoker | 80 | Lung | 0.005 | (0.002, 0.010) |
| Men | Rectal bleeding | Ever smoker | 80 | Upper GI | 0.006 | (0.002, 0.016) |
| Men | Rectal bleeding | Ever smoker | 80 | Lower GI | 0.091 | (0.070, 0.118) |
| Men | Rectal bleeding | Ever smoker | 80 | Urological | 0.004 | (0.001, 0.011) |
| Men | Rectal bleeding | Ever smoker | 80 | Prostate | 0.016 | (0.008, 0.031) |
| Men | Rectal bleeding | Ever smoker | 80 | Haematological | 0.004 | (0.001, 0.010) |
| Men | Rectal bleeding | Ever smoker | 80 | Other cancers | 0.008 | (0.004, 0.013) |
| Men | Rectal bleeding | Ever smoker | 80 | Death w/o cancer | 0.048 | (0.036, 0.064) |
| Men | Rectal bleeding | Ever smoker | 90 | Lung | 0.006 | (0.002, 0.013) |
| Men | Rectal bleeding | Ever smoker | 90 | Upper GI | 0.006 | (0.003, 0.012) |
| Men | Rectal bleeding | Ever smoker | 90 | Lower GI | 0.100 | (0.078, 0.127) |
| Men | Rectal bleeding | Ever smoker | 90 | Urological | 0.004 | (0.002, 0.011) |
| Men | Rectal bleeding | Ever smoker | 90 | Prostate | 0.017 | (0.011, 0.027) |
| Men | Rectal bleeding | Ever smoker | 90 | Haematological | 0.004 | (0.002, 0.011) |
| Men | Rectal bleeding | Ever smoker | 90 | Other cancers | 0.010 | (0.004, 0.021) |
| Men | Rectal bleeding | Ever smoker | 90 | Death w/o cancer | 0.140 | (0.117, 0.167) |
| Men | Weight loss | Never smoker | 30 | Other cancers | 0.001 | (<0.001, 0.002) |
| Men | Weight loss | Never smoker | 30 | Death w/o cancer | 0.002 | (<0.001, 0.004) |
| Men | Weight loss | Never smoker | 40 | Haematological | 0.001 | (<0.001, 0.004) |
| Men | Weight loss | Never smoker | 40 | Other cancers | 0.002 | (<0.001, 0.003) |
| Men | Weight loss | Never smoker | 40 | Death w/o cancer | 0.004 | (0.002, 0.008) |
| Men | Weight loss | Never smoker | 50 | Lung | 0.001 | (<0.001, 0.002) |
| Men | Weight loss | Never smoker | 50 | Upper GI | 0.004 | (0.002, 0.007) |
| Men | Weight loss | Never smoker | 50 | Lower GI | 0.003 | (0.001, 0.006) |
| Men | Weight loss | Never smoker | 50 | Urological | 0.002 | (<0.001, 0.004) |
| Men | Weight loss | Never smoker | 50 | Prostate | 0.001 | (<0.001, 0.003) |
| Men | Weight loss | Never smoker | 50 | Haematological | 0.002 | (<0.001, 0.006) |
| Men | Weight loss | Never smoker | 50 | Other cancers | 0.004 | (0.001, 0.010) |
| Men | Weight loss | Never smoker | 50 | Death w/o cancer | 0.007 | (0.004, 0.013) |
| Men | Weight loss | Never smoker | 60 | Lung | 0.004 | (0.001, 0.013) |
| Men | Weight loss | Never smoker | 60 | Upper GI | 0.010 | (0.006, 0.018) |
| Men | Weight loss | Never smoker | 60 | Lower GI | 0.007 | (0.003, 0.018) |
| Men | Weight loss | Never smoker | 60 | Urological | 0.004 | (0.001, 0.010) |
| Men | Weight loss | Never smoker | 60 | Prostate | 0.011 | (0.006, 0.018) |
| Men | Weight loss | Never smoker | 60 | Haematological | 0.006 | (0.003, 0.013) |
| Men | Weight loss | Never smoker | 60 | Other cancers | 0.006 | (0.003, 0.012) |
| Men | Weight loss | Never smoker | 60 | Death w/o cancer | 0.015 | (0.009, 0.026) |
| Men | Weight loss | Never smoker | 70 | Lung | 0.009 | (0.004, 0.019) |
| Men | Weight loss | Never smoker | 70 | Upper GI | 0.020 | (0.012, 0.035) |
| Men | Weight loss | Never smoker | 70 | Lower GI | 0.016 | (0.010, 0.027) |
| Men | Weight loss | Never smoker | 70 | Urological | 0.006 | (0.003, 0.013) |
| Men | Weight loss | Never smoker | 70 | Prostate | 0.025 | (0.016, 0.039) |
| Men | Weight loss | Never smoker | 70 | Haematological | 0.011 | (0.005, 0.023) |
| Men | Weight loss | Never smoker | 70 | Other cancers | 0.011 | (0.006, 0.020) |
| Men | Weight loss | Never smoker | 70 | Death w/o cancer | 0.035 | (0.024, 0.050) |
| Men | Weight loss | Never smoker | 80 | Lung | 0.012 | (0.006, 0.021) |
| Men | Weight loss | Never smoker | 80 | Upper GI | 0.024 | (0.016, 0.036) |
| Men | Weight loss | Never smoker | 80 | Lower GI | 0.021 | (0.014, 0.031) |
| Men | Weight loss | Never smoker | 80 | Urological | 0.008 | (0.002, 0.023) |
| Men | Weight loss | Never smoker | 80 | Prostate | 0.031 | (0.021, 0.046) |
| Men | Weight loss | Never smoker | 80 | Haematological | 0.015 | (0.008, 0.026) |
| Men | Weight loss | Never smoker | 80 | Other cancers | 0.015 | (0.008, 0.027) |
| Men | Weight loss | Never smoker | 80 | Death w/o cancer | 0.099 | (0.078, 0.124) |
| Men | Weight loss | Never smoker | 90 | Lung | 0.012 | (0.007, 0.020) |
| Men | Weight loss | Never smoker | 90 | Upper GI | 0.023 | (0.014, 0.037) |
| Men | Weight loss | Never smoker | 90 | Lower GI | 0.023 | (0.013, 0.040) |
| Men | Weight loss | Never smoker | 90 | Urological | 0.007 | (0.004, 0.015) |
| Men | Weight loss | Never smoker | 90 | Prostate | 0.032 | (0.022, 0.046) |
| Men | Weight loss | Never smoker | 90 | Haematological | 0.017 | (0.010, 0.029) |
| Men | Weight loss | Never smoker | 90 | Other cancers | 0.018 | (0.011, 0.030) |
| Men | Weight loss | Never smoker | 90 | Death w/o cancer | 0.262 | (0.229, 0.298) |
| Men | Weight loss | Ever smoker | 30 | Other cancers | 0.002 | (<0.001, 0.003) |
| Men | Weight loss | Ever smoker | 30 | Death w/o cancer | 0.002 | (0.001, 0.004) |
| Men | Weight loss | Ever smoker | 40 | Upper GI | 0.001 | (<0.001, 0.002) |
| Men | Weight loss | Ever smoker | 40 | Haematological | 0.002 | (<0.001, 0.003) |
| Men | Weight loss | Ever smoker | 40 | Other cancers | 0.002 | (0.001, 0.004) |
| Men | Weight loss | Ever smoker | 40 | Death w/o cancer | 0.004 | (0.002, 0.011) |
| Men | Weight loss | Ever smoker | 50 | Lung | 0.004 | (0.003, 0.007) |
| Men | Weight loss | Ever smoker | 50 | Upper GI | 0.004 | (0.002, 0.011) |
| Men | Weight loss | Ever smoker | 50 | Lower GI | 0.003 | (0.001, 0.007) |
| Men | Weight loss | Ever smoker | 50 | Urological | 0.002 | (<0.001, 0.005) |
| Men | Weight loss | Ever smoker | 50 | Prostate | 0.001 | (<0.001, 0.002) |
| Men | Weight loss | Ever smoker | 50 | Haematological | 0.002 | (0.001, 0.006) |
| Men | Weight loss | Ever smoker | 50 | Other cancers | 0.004 | (0.002, 0.009) |
| Men | Weight loss | Ever smoker | 50 | Death w/o cancer | 0.008 | (0.004, 0.016) |
| Men | Weight loss | Ever smoker | 60 | Lung | 0.016 | (0.010, 0.025) |
| Men | Weight loss | Ever smoker | 60 | Upper GI | 0.014 | (0.008, 0.024) |
| Men | Weight loss | Ever smoker | 60 | Lower GI | 0.007 | (0.003, 0.018) |
| Men | Weight loss | Ever smoker | 60 | Urological | 0.005 | (0.002, 0.011) |
| Men | Weight loss | Ever smoker | 60 | Prostate | 0.009 | (0.004, 0.020) |
| Men | Weight loss | Ever smoker | 60 | Haematological | 0.006 | (0.002, 0.014) |
| Men | Weight loss | Ever smoker | 60 | Other cancers | 0.008 | (0.004, 0.017) |
| Men | Weight loss | Ever smoker | 60 | Death w/o cancer | 0.016 | (0.009, 0.027) |
| Men | Weight loss | Ever smoker | 70 | Lung | 0.034 | (0.026, 0.045) |
| Men | Weight loss | Ever smoker | 70 | Upper GI | 0.027 | (0.017, 0.044) |
| Men | Weight loss | Ever smoker | 70 | Lower GI | 0.015 | (0.007, 0.031) |
| Men | Weight loss | Ever smoker | 70 | Urological | 0.009 | (0.004, 0.019) |
| Men | Weight loss | Ever smoker | 70 | Prostate | 0.021 | (0.013, 0.033) |
| Men | Weight loss | Ever smoker | 70 | Haematological | 0.011 | (0.008, 0.017) |
| Men | Weight loss | Ever smoker | 70 | Other cancers | 0.012 | (0.007, 0.021) |
| Men | Weight loss | Ever smoker | 70 | Death w/o cancer | 0.038 | (0.026, 0.054) |
| Men | Weight loss | Ever smoker | 80 | Lung | 0.044 | (0.032, 0.059) |
| Men | Weight loss | Ever smoker | 80 | Upper GI | 0.031 | (0.020, 0.047) |
| Men | Weight loss | Ever smoker | 80 | Lower GI | 0.021 | (0.013, 0.035) |
| Men | Weight loss | Ever smoker | 80 | Urological | 0.010 | (0.005, 0.019) |
| Men | Weight loss | Ever smoker | 80 | Prostate | 0.027 | (0.016, 0.045) |
| Men | Weight loss | Ever smoker | 80 | Haematological | 0.015 | (0.009, 0.025) |
| Men | Weight loss | Ever smoker | 80 | Other cancers | 0.016 | (0.010, 0.028) |
| Men | Weight loss | Ever smoker | 80 | Death w/o cancer | 0.102 | (0.084, 0.122) |
| Men | Weight loss | Ever smoker | 90 | Lung | 0.047 | (0.031, 0.069) |
| Men | Weight loss | Ever smoker | 90 | Upper GI | 0.031 | (0.022, 0.044) |
| Men | Weight loss | Ever smoker | 90 | Lower GI | 0.023 | (0.015, 0.036) |
| Men | Weight loss | Ever smoker | 90 | Urological | 0.010 | (0.005, 0.019) |
| Men | Weight loss | Ever smoker | 90 | Prostate | 0.026 | (0.019, 0.036) |
| Men | Weight loss | Ever smoker | 90 | Haematological | 0.017 | (0.009, 0.030) |
| Men | Weight loss | Ever smoker | 90 | Other cancers | 0.021 | (0.014, 0.031) |
| Men | Weight loss | Ever smoker | 90 | Death w/o cancer | 0.271 | (0.238, 0.306) |

### Appendix 6. Additional detail on the risk of specific cancer sites.

*Risk of breast cancer*

Risk of breast cancer in women presenting with any of the studied symptoms other than breast lump was always under the 3% urgent referral threshold (Figures 4 and 5), while for those presenting with breast lump it reached this threshold from age 35 (smokers) / 40 (non-smokers). In women with breast lump risk appeared to increase rapidly with age, and exceeded 10% from age 50 and 20% from age 60. Breast cancer in men was grouped into the 'other cancers’ category (see below).

*Risk of gynaecological cancers*

Risk of gynaecological cancer exceeded the 3% urgent referral threshold in women presenting with post-menopausal bleeding (Figures 4 and 5); post-menopausal bleeding at any age appeared associated with greater than 3% risk (although estimates for younger ages were based on very small sample sizes). While gynaecological cancer did not reach the 3% threshold for other 14 studied symptoms, there was notable risk of gynaecological cancer in women presenting with haematuria and abdominal bloating. For example, 40-year-old female non-smokers presenting with haematuria had a risk of gynaecological cancer of 0.6% (95% CI 0.2% to 1.5%, Figure 4, Appendix 5 Table 1), and with abdominal bloating a risk of 0.7% (95% CI 0.3% to 1.8%).

*Risk of lung cancer*

Lung cancer risk in non-smokers was generally minimal across the studied symptoms, except for haemoptysis. Male non-smokers presenting with haemoptysis had 3% risk of lung cancer from age 70 (Table 2, Figure 2). In smokers, there was notable lung cancer risk in patients presenting with haemoptysis (3% or higher risk from age 55 for male smokers, 60 for female smokers, Figures 3 and 5) and for male smokers presenting with weight loss (reaching 3% risk from age 70). For dyspnoea and all three non-specific symptoms studied (fatigue, night sweats, weight loss), there was elevated risk of lung cancer for male and female smokers presenting, though without reaching 3%.

*Risk of upper GI cancers*

Risk of upper GI cancers exceeded the 3% referral threshold for both men and women presenting with dysphagia (from age 60 in men, 70 in women) and jaundice (from age 50). Patients presenting with dyspepsia, abdominal pain, abdominal bloating and weight loss also had notable risk of upper GI cancer, but not reaching the 3% threshold except for male smokers presenting with weight loss, where the risk of upper GI cancer reached 3% from age 75. The specific types of upper GI cancer varied by index symptom. Pancreatic cancer was the most commonly diagnosed cancer following a presentation with jaundice (237 pancreatic cancers of 456 total cancers in patients with jaundice as index symptom, Appendix 2 Table 1), while dysphagia was more frequently associated with oesophageal cancer (558 oesophageal cancers of 1,036 total cancers).

*Risk of lower GI cancers*

Risk of lower GI cancers exceeded the 3% referral threshold for patients presenting with rectal bleeding or change in bowel habit from around age 60 in men and around age 70 in women. Patients presenting with abdominal pain and abdominal bloating, and also non-specific symptoms such as fatigue, night sweats, and weight loss, appeared to have a 1-2% risk of lower GI cancer from around age 60.

*Risk of urological cancers*

Risk of urological cancer was higher in smokers than non-smokers, and reached the 3% urgent referral threshold for patients presenting with haematuria from age 55 in men and age 60/65 in female smokers/non-smokers.

*Risk of prostate cancer*

Risk of prostate cancer reached 3% from around age 70 in men presenting with haematuria, and from age 80 in male non-smokers presenting with weight loss. Older men presenting with many of the studied symptoms had 1-2% risk of prostate cancer (including following presentation with abdominal pain and bloating, rectal bleeding, change in bowel habit, dyspepsia, fatigue, night sweats and weight loss).

*Risk of haematological cancers*

While many symptoms were associated with relatively large hazard ratios for haematological cancer, the risk of haematological cancers did not reach the 3% referral threshold for any of the studied symptoms. Night sweats in older male patients were associated with the highest risk of haematological cancers.

*Risk of other cancers*

Risk of other cancers, including breast cancer in men and all other cancers excluding non-melanoma skin cancer, reached the 3% urgent referral threshold from around age 55 for patients presenting with jaundice, and from around age 75 for men presenting with breast lump.

### Appendix 7. Additional detail on the limitations of this study

This section expands on the brief summary of limitations given in the main text of the article.

First, the consideration of deaths in patients with cancer. In this analysis, only the first of the two outcomes (either diagnosis of cancer or death) was considered, and so deaths in patients with cancer were not examined in detail, beyond briefly in Table 1. Research to inform diagnostic strategies may additionally consider whether certain symptomatic presentations were associated with worse outcomes in patients diagnosed with cancer, but the focus of this study was cancer incidence alone.

Second, the assessment of smoking-status used in this study was crude, classifying patients as ‘ever-smokers’ if they ever had a positive smoking-related Read code, and ‘never-smokers’ otherwise. This will have led some patients with minimal or no history of smoking being classed as smokers, and did not allow any examination of possible dose-response relationships with measures such as pack-years of smoking [1,2]. A more refined stratification of risk related to exposure to smoking (e.g., including smoking intensity) may be attempted in future as this could further support referral decisions.

Third, each patient was included at most once in the analysis. Landmark methods with multiple observations per patient would have made better use of the longitudinal data from the electronic health record [3], and would have avoided the need to remove patients from the random sample from consideration for their symptom status. Using landmark approaches would have allowed consideration of symptom history (i.e., patients who have previously presented with a symptom and have returned again with that symptom may be different risk of cancer than those presenting for the first time). We considered adopting landmark methods in this analysis but decided against it due to challenges in accurately estimating variances, and the relatively limited precision benefits with an analysis cohort that already included 1.7 million patients.

Fourth, examination of risk of cancer in patients with multiple symptoms was limited. Analysis included all symptoms patients had at index when estimating cause-specific hazard ratios, but we did not consider possible interactions between index symptoms and we simulated cumulative incidence curves for single index symptoms only. Further simulations could be carried out to produce cumulative incidence curves for combinations of index symptoms, although given only a small fraction of patients have multiple index symptoms the value of such analyses may be limited in practice.

Fifth, analysis relies entirely on symptoms that were recorded in coded form in the electronic health record. We know that coded data does not capture all symptoms, or even all symptoms that are entered in free-text in the patient’s record [4,5], despite the proportion of cancer patients in CPRD who are symptomatic being similar to the proportion of patients in the National Cancer Diagnosis Audit for whom a symptom was judged to be relevant to their diagnosis [6,7]. This motivated the use of a random sample as a reference group rather than an apparently-symptom-free comparison group; we could be confident that patients with a recorded symptom had that symptom, but it was harder to be certain that a patient without a recorded symptom did not have a symptom. It would be preferable to have access to free-text data to get better ascertainment of patients’ symptom status, bearing in mind that weather the symptom was recorded using structured code or entered in free-text may in itself be informative of risk level. Yet data protection concerns mean that access to free-text data for research is limited at present.

Sixth, while we examine 15 different symptoms possibly related to an underlying cancer diagnosis, there are over 70 other symptoms that could have additionally been examined [8]. Further, certain comorbidities may be linked to both the presence of a symptom and underlying risk of cancer, for example COPD is linked to both dyspnoea and lung cancer risk [9]. The symptomatic presentation and diagnostic management of patients with cancer and prior comorbidities may vary to that observed in patients without pre-existing chronic conditions [10]. There are practical limits on how much can be examined in a multi-exposure multi-outcome descriptive study, and examining the complicated associations between comorbidities, symptoms, and individual cancer sites is likely best served by carefully designed single-outcome studies.

*Citations in above text (different numbering from article main-text)*

1	Hippisley-Cox J, Coupland C. Symptoms and risk factors to identify women with suspected cancer in primary care: derivation and validation of an algorithm. *The British Journal of General Practice*. 2013;63:e11.

2	Hippisley-Cox J, Coupland C. Symptoms and risk factors to identify men with suspected cancer in primary care: derivation and validation of an algorithm. *British Journal of General Practice*. 2013;63:e1–10.

3	Keogh RH, Seaman SR, Barrett JK, *et al.* Dynamic Prediction of Survival in Cystic Fibrosis: A Landmarking Analysis Using UK Patient Registry Data. *Epidemiology*. 2019;30:29–37.

4	Kostopoulou O, Tracey C, Delaney BC. Can decision support combat incompleteness and bias in routine primary care data? *Journal of the American Medical Informatics Association*. 2021;28:1461–7.

5	Price SJ, Stapley SA, Shephard E, *et al.* Is omission of free text records a possible source of data loss and bias in Clinical Practice Research Datalink studies? A case-control study. *BMJ open*. 2016;6. doi: 10.1136/BMJOPEN-2016-011664

6	Barclay M, Renzi C, Antoniou A, *et al.* Phenotypes and rates of cancer-relevant symptoms and tests in the year before cancer diagnosis in UK Biobank and CPRD Gold. *PLOS Digital Health*. 2023;2:e0000383.

7	Zakkak N, Barclay ME, Swann R, *et al.* The presenting symptom signatures of incident cancer: evidence from the English 2018 National Cancer Diagnosis Audit. *Br J Cancer*. 2023;1–11.

8	Moore SF, Price SJ, Chowienczyk S, *et al.* The impact of changing risk thresholds on the number of people in England eligible for urgent investigation for possible cancer: an observational cross-sectional study. *British Journal of Cancer*. 2021;125:1593–7.

9	Skillrud D, Offord K, Miller R. Higher Risk of Lung Cancer in Chronic Obstructive Pulmonary Disease. *Annals of Internal Medicine*. 1986;105:503.

10	Majano SB, Lyratzopoulos G, Rachet B, *et al.* Do presenting symptoms, use of pre-diagnostic endoscopy and risk of emergency cancer diagnosis vary by comorbidity burden and type in patients with colorectal cancer? *British Journal of Cancer 2021 126:4*. 2021;126:652–63.
